# Thinner cortex is associated with psychosis onset in individuals at Clinical High Risk for Developing Psychosis: An ENIGMA Working Group mega-analysis

**DOI:** 10.1101/2021.01.05.20248768

**Authors:** Maria Jalbrzikowski, Rebecca A. Hayes, Stephen J. Wood, Dorte Nordholm, Juan H. Zhou, Paolo Fusar-Poli, Peter J. Uhlhaas, Tsutomu Takahashi, Gisela Sugranyes, Yoo Bin Kwak, Daniel H. Mathalon, Naoyuki Katagiri, Christine I. Hooker, Lukasz Smigielski, Tiziano Colibazzi, Esther Via, Jinsong Tang, Shinsuke Koike, Paul E. Rasser, Chantal Michel, Irina Lebedeva, Wenche ten Velden Hegelstad, Camilo de la Fuente-Sandoval, James A. Waltz, Romina R.M. Mizrahi, Cheryl Corcoran, Franz Resch, Christian K. Tamnes, Shalaila S. Haas, Imke L.J. Lemmers-Jansen, Ingrid Agartz, Paul Allen, Ole A. Andreassen, Kimberley Atkinson, Peter Bachman, Inmaculada Baeza, Helen Baldwin, Cali F. Bartholomeusz, Kolbjørn S. Brønnick, Sabrina Catalano, Michael W.L. Chee, Xiaogang Chen, Kang Ik K. Cho, Rebecca E. Cooper, Vanessa L. Cropley, Montserrat Dolz, Bjørn H. Ebdrup, Adriana Fortea, Louise B. Glenthøj, Birte Y. Glenthøj, Lieuwe de Haan, Holly K. Hamilton, Mathew A. Harris, Kristen M. Haut, Ying He, Karsten Heekeren, Andreas Heinz, Daniela Hubl, Wu Jeong Hwang, Michael Kaess, Kiyoto Kasai, Minah Kim, Jochen Kindler, Mallory J. Klaunig, Tina D. Kristensen, Jun Soo Kwon, Stephen M. Lawrie, Jimmy Lee, Pablo León-Ortiz, Ashleigh Lin, Rachel L. Loewy, Xiaoqian Ma, Daniel H. Mathalon, Patrick McGorry, Philip McGuire, Masafumi Mizuno, Paul Møller, Toma Moncada-Habib, Daniel Munoz-Samons, Barnaby Nelson, Takahiro Nemoto, Merete Nordentoft, Maria A. Omelchenko, Ketil Oppedal, Lijun Ouyang, Christos Pantelis, Jose C. Pariente, Jayachandra Raghava, Francisco Reyes-Madrigal, Brian J. Roach, Jan I. Røssberg, Wulf Rössler, Dean F. Salisbury, Daiki Sasabayashi, Ulrich Schall, Jason Schiffman, Florian Schlagenhauf, Andre Schmidt, Mikkel E. Sørensen, Michio Suzuki, Anastasia Theodoridou, Alexander S. Tomyshev, Jordina Tor, Tor G Vaernes, Dennis Velakoulis, Gloria D. Venegoni, Sophia Vinogradov, Christina Wenneberg, Lars T. Westlye, Hidenori Yamasue, Liu Yuan, Alison R. Yung, Thérèse A.M.J. van Amelsvoort, Jessica A. Turner, Theo G.M. van Erp, Paul M. Thompson, Dennis Hernaus, Koppel, Borgwardt

**Affiliations:** Amsterdam UMC, Amsterdam, The Netherlands; Arkin, Amsterdam, The Netherlands; Center for Clinical Intervention and Neuropsychiatric Schizophrenia Research, CINS, Mental Health Centre Glostrup, University of Copenhagen, Glostrup, Denmark; Center for Evolutionary Cognitive Sciences, Graduate School of Art and Sciences, The University of Tokyo; Center for Sleep and Cognition, Yong Loo Lin School of Medicine, National University of Singapore, Singapore; Center for the Neurobiology of Learning and Memory, 309 Qureshey Research Lab, Irvine, CA, USA; Center for Translational Magnetic Resonance Research, Yong Loo Lin School of Medicine, National University of Singapore, Singapore; Centre for Clinical Intervention and Neuropsychiatric Schizophrenia Research (CINS), Glostrup, Denmark; Centre for Mental Health, Faculty of Health, Arts and Design, School of Health Sciences, Swinburne University, Melbourne, Australia; Centre for Neuropsychiatric Schizophrenia Research, CNSR, Glostrup, Denmark; Centre for Sleep and Cognition, Yong Loo Lin School of Medicine, National University of Singapore, Singapore; Centre for Youth Mental Health, University of Melbourne, Australia; Child and Adolescent Mental Health Research Group, Institut de Recerca Sant Joan de Déu, Barcelona, Spain; Child and Adolescent Psychiatry and Psychology Department, Hospital Sant Joan de Déu, Barcelona, Spain; Clinic for Child and Adolescent Psychiatry, University Hospital of Heidelberg, Heidelberg, Germany; Clinical Translational Neuroscience Laboratory, Department of Psychiatry and Human Behavior, University of California Irvine, Irvine, CA, USA; Columbia University Department of Psychiatry, New York, USA; Copenhagen Research Center for Mental Health - CORE, Mental Health Center Copenhagen, Copenhagen University Hospital, Denmark; Department for Mental Health Research and Development, Division of Mental Health and Addiction, Vestre Viken Hospital Trust, Norway; Department of Brain and Behavioral Sciences, University of Pavia, Italy; Department of Brain and Cognitive Sciences, Seoul National University College of Natural Sciences, Republic of Korea; Department of Child and Adolescent Psychiatry and Psychology, Institute of Neuroscience, Hospital Clinic Barcelona, Fundació Clínic Recerca Biomèdica, Universitat de Barcelona, Spain; Department of Child and Adolescent Psychiatry and Psychology, Institute of Neuroscience, Hospital Clinic Barcelona, Institut d’Investigacions Biomèdiques August Pi i Sunyer (IDIBAPS), CIBERSAM, Universitat de Barcelona, Spain; Department of Child and Adolescent Psychiatry, Psychiatric University Hospital Zurich, University of Zurich, Switzerland; Department of Child and Adolescent Psychiatry, Center of Psychosocial Medicine, University of Heidelberg, Heidelberg, Germany; Department of Child and Adolescent Psychiatry, Charité Universitätsmedizin, Berlin, Germany; Department of Clinical Medicine, Faculty of Health and Medical Sciences, University of Copenhagen, Denmark; Department of Clinical Physiology, Nuclear Medicine and PET, Functional Imaging Unit, Glostrup, Denmark; Department of Neuropsychiatry, Graduate School of Medicine, The University of Tokyo, Japan; Department of Neuropsychiatry, Seoul National University Hospital, Republic of Korea; Department of Neuropsychiatry, Toho University School of Medicine, Japan; Department of Neuropsychiatry, University of Toyama Graduate School of Medicine and Pharmaceutical Sciences, Toyama, Japan; Department of Pharmacology and Toxicology, University of Toronto, Canada; Department of Psychiatric Research, Diakonhjemmet Hospital, Oslo, Norway; Department of Psychiatry & Behavioral Sciences, University of California, San Francisco, CA, USA; Department of Psychiatry & Neuropsychology, School for Mental Health and Neuroscience, Maastricht University, The Netherlands; Department of Psychiatry (UPK), University of Basel, Switzerland; Department of Psychiatry and Behavioral Sciences, Rush University Medical Center, Chicago IL; Department of Psychiatry and Psychotherapy I, LVR-Hospital Cologne, Cologne, Germany; Department of Psychiatry and Psychotherapy, Charité – Universitätsmedizin Berlin, corporate member of Freie Universität Berlin, Humboldt-Universität zu Berlin, and Berlin Institute of Health, Germany; Department of Psychiatry and Psychotherapy, Charité Universitätsmedizin, Berlin, Germany; Department of Psychiatry and Psychotherapy, University of Lübeck, Germany; Department of Psychiatry, Hamamatsu University School of Medicine, Hamamatsu City, Japan; Department of Psychiatry, Icahn School of Medicine at Mount Sinai, New York, NY, USA; Department of Psychiatry, Psychiatry Neuroimaging Laboratory, Brigham and Women’s Hospital, Harvard Medical School, Boston, MA, USA; Department of Psychiatry, Psychotherapy and Psychosomatics, Psychiatric University Hospital Zurich, University of Zurich, Switzerland; Department of Psychiatry, Rush University Medical Center, Chicago, IL, USA; Department of Psychiatry, Seoul National University College of Medicine, Republic of Korea; Department of Psychiatry, Sir Run Run Shaw Hospital, School of Medicine, Zhejiang University, Hangzhou, Zhejiang, China; Department of Psychiatry, the Second Xiangya Hospital, Central South University, Changsha, Hunan, China; Department of Psychiatry, University of California, San Francisco, CA, USA; Department of Psychiatry, University of Edinburgh, UK; Department of Psychiatry, University of Minnesota, MN, USA; Department of Psychiatry, University of Pittsburgh, PA, USA; Department of Psychological Science, University of California, Irvine, CA, USA; Department of Psychology, University of Maryland, Baltimore County, MD, USA; Department of Psychology, University of Oslo, Norway; Department of Psychology, University of Roehampton, London, UK; Department of Psychosis Studies, Institute of Psychiatry, Psychology and Neuroscience, King’s College London, UK; Department of Psychosis, Institute of Mental Health, Singapore; Division of Psychiatry, University of Edinburgh, Edinburgh, UK; Douglas Research Center, Montreal, Canada; Early Intervention in Psychosis Advisory Unit for South-East Norway, TIPS Sør-Øst, Division of Mental Health and Addiction, Oslo University Hospital, Oslo, Norway; EPIC Lab, Department of Psychosis Studies, King’s College London, UK; Faculty of Behavioural and Movement sciences, Department of Clinical, Neuro and Developmental Psychology, Vrije Universiteit Amsterdam, The Netherlands; Faculty of Social Sciences,University of Stavanger, Norway; Florey Institute of Neuroscience and Mental Health, Australia; Georgia State University, Atlanta, GA, USA; Hunan Key Laboratory of Psychiatry and Mental Health, Changsha, Hunan, China; Imaging Genetics Center, Mark & Mary Stevens Institute for Neuroimaging & Informatics, Keck USC School of Medicine, Los Angeles, CA, USA; Institute of Neuroscience and Psychology, University of Glasgow, Glasgow, UK; James J Peters VA, Bronx, NY, USA (MIRECC); Key Laboratory of Medical Neurobiology of Zhejiang Province, Zhejiang, China; King’s College London, UK; Laboratory of Experimental Psychiatry, Instituto Nacional de Neurología y Neurocirugía, Mexico City, Mexico; Lee Kong Chian School of Medicine, Nanyang Technological University, Singapore; Maastricht University, The Netherlands; Magnetic Resonance Imaging core facility, Institut d’Investigacions Biomèdiques August Pi i Sunyer (IDIBAPS), Barcelona, Spain; Maryland Psychiatric Research Center, University of Maryland School of Medicine, MD, USA; McGill University, Montreal, Canada; Melbourne Neuropsychiatry Centre, Department of Psychiatry, University of Melbourne & Melbourne Health, Melbourne, Australia; Mental Health Research Center, Moscow, Russia; National Clinical Research Center for Geriatric Disorders, Xiangya Hospital, Central South University, Changsha, China; Neuropsychiatry, The Royal Melbourne Hospital, Australia; New York State Psychiatric Institute, NY, USA; NIHR Maudsley Biomedical Research Centre (South London and Maudsley NHS Foundation Trust & King’s College London), UK; NORMENT, Division of Mental Health and Addiction, Oslo University Hospital & Institute of Clinical Medicine, University of Oslo, Oslo, Norway; Orygen, The University of Melbourne, Melbourne, Australia; Priority Centre for Brain & Mental Health Research, The University of Newcastle, Australia; Priority Research Centre for Stroke and Brain Injury, The University of Newcastle, Australia; Priority Research Centre Grow Up Well, The University of Newcastle, Australia; PROMENTA Research Center, Department of Psychology, University of Oslo, Norway; Psychosis Studies, Institute of Psychiatry, Psychology and Neuroscience, King’s College London, UK; Research Center for Idling Brain Science, University of Toyama, Toyama, Japan; San Francisco Veterans Affairs Health Care System, San Francisco, CA; School of Psychology, University of Birmingham, UK; Stavanger Medical Imaging Laboratory (SMIL), Stavanger University Hospital, Norway; Telethon Kids Institute, The University of Western Australia, Perth, Australia; The University of Tokyo Institute for Diversity and Adaptation of Human Mind (UTIDAHM), Japan; TIPS Network for Clinical Research in Psychosis, Stavanger University Hospital, Norway; Translational Research Center, University Hospital of Psychiatry and Psychotherapy, University of Bern, Bern, Switzerland; University Hospital of Child and Adolescent Psychiatry and Psychotherapy, University of Bern, Bern, Switzerland

**Keywords:** magnetic resonance imaging, cortical thickness, ultra high-risk for psychosis, prodromal, schizophrenia, at-risk mental states, gray matter, brain volume, CHR, clinical high risk

## Abstract

**Importance:** The ENIGMA clinical high risk for psychosis (CHR) initiative, the largest pooled CHR-neuroimaging sample to date, aims to discover robust neurobiological markers of psychosis risk in a sample with known heterogeneous outcomes.

**Objective:** We investigated baseline structural neuroimaging differences between CHR subjects and healthy controls (HC), and between CHR participants who later developed a psychotic disorder (CHR-PS+) and those who did *not* (CHR-PS-). We assessed associations with age by group and conversion status, and similarities between the patterns of effect size maps for psychosis conversion and those found in other large-scale psychosis studies.

**Design, Setting, and Participants:** Baseline T1-weighted MRI data were pooled from 31 international sites participating in the ENIGMA CHR Working Group. MRI scans were processed using harmonized protocols and analyzed within a mega- and meta-analysis framework from January-October 2020.

**Main Outcome(s) and Measure(s):** Measures of regional cortical thickness (CT), surface area (SA), and subcortical volumes were extracted from T1-weighted MRI scans. Independent variables were group (CHR, HC) and conversion status (CHR-PS+, CHR-PS-, HC).

**Results:** The final dataset consisted of 3,169 participants (CHR=1,792, HC=1,377, age range: 9.5 to 39.8 years, 45% female). Using longitudinal clinical information, we identified CHR-PS+ (N=253) and CHR-PS- (N=1,234). CHR exhibited widespread thinner cortex compared to HC (average *d*=-0.125, range: -0.09 to -0.17), but not SA or subcortical volume. Thinner cortex in the fusiform, superior temporal, and paracentral regions was associated with psychosis conversion (average *d*=-0.22). Age showed a stronger negative association with left fusiform and left paracentral CT in HC, compared to CHR-PS+. Regional CT psychosis conversion effect sizes resembled patterns of CT alterations observed in other ENIGMA studies of psychosis.

**Conclusions and Relevance:** We provide evidence for widespread subtle CT reductions in CHR. The pattern of regions displaying greater CT alterations in CHR-PS+ were similar to those reported in other large-scale investigations of psychosis. Additionally, a subset of these regions displayed abnormal age associations. Widespread CT disruptions coupled with abnormal age associations in CHR may point to disruptions in postnatal brain developmental processes.

**Key Points:** *Question:* How do baseline brain morphometric features relate to later psychosis conversion in individuals at clinical high risk (CHR)?

*Findings:* In the largest coordinated international analysis to date, reduced baseline cortical thickness, but not cortical surface area or subcortical volume, was more pronounced in CHR, in a manner highly consistent with thinner cortex in established psychosis. Regions that displayed greater cortical thinning in future psychosis converters additionally displayed abnormal associations with age.

*Meaning:* CHR status and later transition to psychosis is robustly associated with reduced cortical thickness. Abnormal age associations and specificity to cortical thickness may point to aberrant postnatal brain development in CHR, including pruning and myelination.

## 1. INTRODUCTION

The clinical high-risk paradigm is a widely used framework to investigate mechanisms underlying psychosis vulnerability. Help-seeking individuals who do not meet diagnostic criteria for a psychotic disorder, but typically present with subthreshold psychotic symptoms and accumulating risk factors, are considered at clinical high-risk (CHR) for developing psychosis^1–3^. An estimated 18-20% of CHR individuals develop a psychotic disorder within 2 years of identification^4^, although conversion rates vary^5–11^, likely due to heterogeneous recruitment and sampling strategies, and interventions applied^2,13^. However, despite decades of research, the nature of morphometrical alterations associated with psychosis conversion remains largely unknown. Here, we aim to address this question by combining all available structural neuroimaging data in CHR to date, in an attempt to better understand brain mechanisms associated with psychosis risk and conversion in this population.

A large body of work has used structural magnetic resonance imaging (sMRI) to investigate morphometric brain alterations in CHR individuals^14–37^. However, the extent to which characteristic baseline structural neuroimaging differences exist between those at CHR who later develop a psychotic disorder (CHR-PS+) compared to those who do *not* (CHR-PS-) is debated. Many studies failed to find baseline differences between CHR-PS+ and CHR-PS- ^15,35,38,39^, though a meta-analysis and multi-center study found lower prefrontal and temporal volumes or cortical thickness (CT) in CHR-PS+^40,41^. High attrition rates in CHR samples^42–44^, coupled with low psychosis conversion rates^4,45^, often yield insufficient power to detect between-group structural brain differences. Moreover, small sample sizes can be associated with inflated effect sizes^46,47^, so effect sizes of prior studies that found structural brain alterations in CHR may be overestimated. Although multi-site consortia aim to address these challenges^34,48–50^, the largest published sMRI studies to date included fewer than 50 CHR-PS+^38,41^. Furthermore, it is currently unknown whether group differences are robust enough to predict outcomes.

Importantly, many CHR participants are adolescents or young adults, a timeframe associated with psychosis onset^51,52^. Several brain regions implicated in psychosis show protracted developmental courses continuing through adolescence^53–63^, suggesting that morphometric alterations associated with psychosis risk vary with age. Indeed, there are developmental influences on psychotic symptom presentation^64–68^, perhaps driven by differences in regional brain changes. It is not fully understood how age-related patterns in brain morphometry in CHR differ from normal development. Thus, using a developmental framework to examine whether morphometric alterations in CHR are influenced by age may provide important insights into mechanisms associated with psychosis risk, and the stability of neuroimaging measures associated with psychosis risk across development.

Finally, it is unknown whether baseline brain alterations associated with future conversion to psychosis resemble those observed in other large-scale psychosis studies. Understanding whether morphometric alterations in CHR overlap with those observed in individuals who have schizophrenia^69–71^ and individuals with a genetic subtype of psychosis^72,73^ will provide insights into convergent or distinct alterations across the psychosis spectrum.

To address these questions, we founded the Enhancing Neuro Imaging Genetics through Meta-Analysis (ENIGMA) Clinical High Risk Working Group in 2018. Using baseline sMRI data and longitudinal clinical information from 31 sites, this study addressed the following questions:

1. Do CHR and healthy control (HC) participants differ in CT, surface area (SA), and/or subcortical volumes?
2. Is there a neuroanatomic signature associated with future transition to a psychotic disorder (CHR-PS+ versus CHR-PS-versus HC)?
3. Do structural neuroimaging measures identified in Aims 1 and 2 display group differences in age associations, suggestive of abnormal developmental trajectories?
4. Is the pattern of morphometric alteration associated with psychosis conversion similar to that observed in ENIGMA studies of psychosis?

## 2. METHODS

### Participants

We included 1,792 CHR and 1,377 HC from 31 sites participating in the ENIGMA CHR Working Group (Table 1). CHR data consisted of CHR-PS+ (N=253), CHR-PS- (N=1,234), and CHR individuals without follow-up data (CHR-UNK, N=305). CHR participants met CAARMS (N=821) or SIPS (N=971) CHR criteria (see details in Supplement). Site-specific inclusion/exclusion criteria are detailed in **eTable 1**. All sites obtained Institutional Review Board approval prior to data collection. Informed written consent was obtained from every participant or the participant’s guardian (for participants <18 years). All studies were conducted in accordance with the Declaration of Helsinki^74^.

**Table 1.**
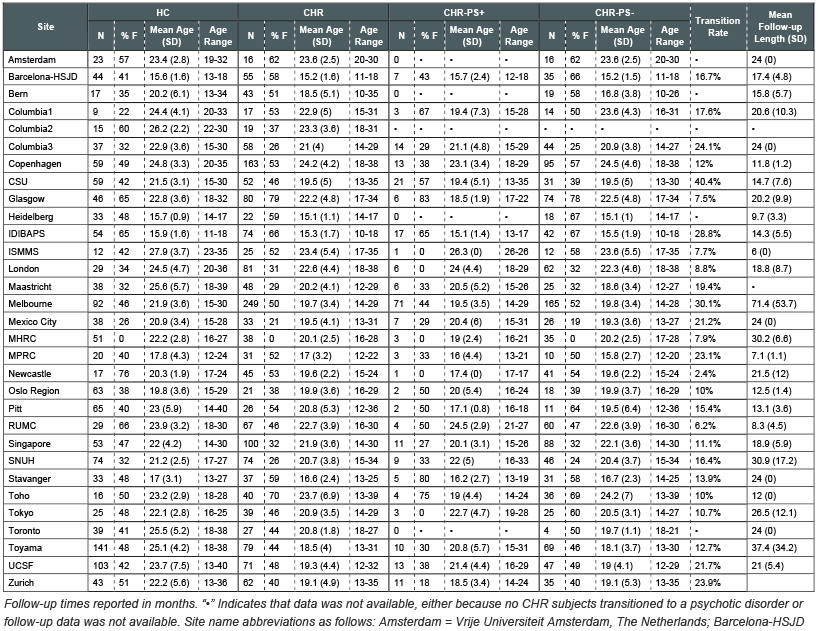
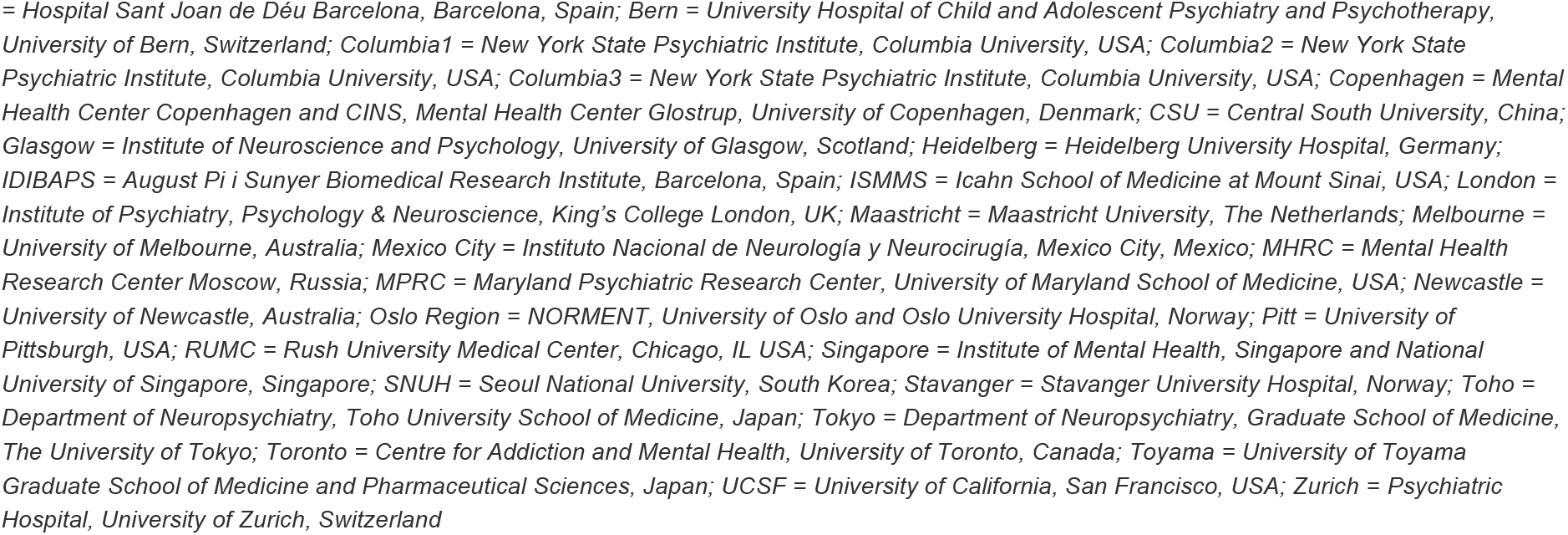
Age and sex information for healthy controls (HC) and clinical high risk for psychosis (CHR) participants at each site

| Site | HC |  |  |  | CHR |  |  |  | CHR-PS+ |  |  |  | CHR-PS- |  |  |  | Transition Rate | Mean Follow-up Length (SD) |
| --- | --- | --- | --- | --- | --- | --- | --- | --- | --- | --- | --- | --- | --- | --- | --- | --- | --- | --- |
|  | N | % F | Mean Age (SD) | Age Range | N | % F | Mean Age (SD) | Age Range | N | % F | Mean Age (SD) | Age Range | N | % F | Mean Age (SD) | Age Range |  |  |
| Amsterdam | 23 | 57 | 23.4 (2.8) | 19-32 | 16 | 62 | 23.6 (2.5) | 20-30 | 0 | • | • | • | 16 | 62 | 23.6 (2.5) | 20-30 | • | 24 (0) |
| Barcelona-HSJD | 44 | 41 | 15.6 (1.6) | 13-18 | 55 | 58 | 15.2 (1.6) | 11-18 | 7 | 43 | 15.7 (2.4) | 12-18 | 35 | 66 | 15.2 (1.5) | 11-18 | 16.7% | 17.4 (4.8) |
| Bern | 17 | 35 | 20.2 (6.1) | 13-34 | 43 | 51 | 18.5 (5.1) | 10-35 | 0 | • | • | • | 19 | 58 | 16.8 (3.8) | 10-26 | • | 15.8 (5.7) |
| Columbia1 | 9 | 22 | 24.4 (4.1) | 20-33 | 17 | 53 | 22.9 (5) | 15-31 | 3 | 67 | 19.4 (7.3) | 15-28 | 14 | 50 | 23.6 (4.3) | 16-31 | 17.6% | 20.6 (10.3) |
| Columbia2 | 15 | 60 | 26.2 (2.2) | 22-30 | 19 | 37 | 23.3 (3.6) | 18-31 | • | • | • | • | • | • | • | • | • | • |
| Columbia3 | 37 | 32 | 22.9 (3.6) | 15-30 | 58 | 26 | 21 (4) | 14-29 | 14 | 29 | 21.1 (4.8) | 15-29 | 44 | 25 | 20.9 (3.8) | 14-27 | 24.1% | 24 (0) |
| Copenhagen | 59 | 49 | 24.8 (3.3) | 20-35 | 163 | 53 | 24.2 (4.2) | 18-38 | 13 | 38 | 23.1 (3.4) | 18-29 | 95 | 57 | 24.5 (4.6) | 18-38 | 12% | 11.8 (1.2) |
| CSU | 59 | 42 | 21.5 (3.1) | 15-30 | 52 | 46 | 19.5 (5) | 13-35 | 21 | 57 | 19.4 (5.1) | 13-35 | 31 | 39 | 19.5 (5) | 13-30 | 40.4% | 14.7 (7.6) |
| Glasgow | 46 | 65 | 22.8 (3.6) | 18-32 | 80 | 79 | 22.2 (4.8) | 17-34 | 6 | 83 | 18.5 (1.9) | 17-22 | 74 | 78 | 22.5 (4.8) | 17-34 | 7.5% | 20.2 (9.9) |
| Heidelberg | 33 | 48 | 15.7 (0.9) | 14-17 | 22 | 59 | 15.1 (1.1) | 14-17 | 0 | • | • | • | 18 | 67 | 15.1 (1) | 14-17 | • | 9.7 (3.3) |
| IDIBAPS | 54 | 65 | 15.9 (1.6) | 11-18 | 74 | 66 | 15.3 (1.7) | 10-18 | 17 | 65 | 15.1 (1.4) | 13-17 | 42 | 67 | 15.5 (1.9) | 10-18 | 28.8% | 14.3 (5.5) |
| ISMMS | 12 | 42 | 27.9 (3.7) | 23-35 | 25 | 52 | 23.4 (5.4) | 17-35 | 1 | 0 | 26.3 (0) | 26-26 | 12 | 58 | 23.6 (5.5) | 17-35 | 7.7% | 6 (0) |
| London | 29 | 34 | 24.5 (4.7) | 20-36 | 81 | 31 | 22.6 (4.4) | 18-38 | 6 | 0 | 24 (4.4) | 18-29 | 62 | 32 | 22.3 (4.6) | 18-38 | 8.8% | 18.8 (8.7) |
| Maastricht | 38 | 32 | 25.6 (5.7) | 18-39 | 48 | 29 | 20.2 (4.1) | 12-29 | 6 | 33 | 20.5 (5.2) | 15-26 | 25 | 32 | 18.6 (3.4) | 12-27 | 19.4% | • |
| Melbourne | 92 | 46 | 21.9 (3.6) | 15-30 | 249 | 50 | 19.7 (3.4) | 14-29 | 71 | 44 | 19.5 (3.5) | 14-29 | 165 | 52 | 19.8 (3.4) | 14-28 | 30.1% | 71.4 (53.7) |
| Mexico City | 38 | 26 | 20.9 (3.4) | 15-28 | 33 | 21 | 19.5 (4.1) | 13-31 | 7 | 29 | 20.4 (6) | 15-31 | 26 | 19 | 19.3 (3.6) | 13-27 | 21.2% | 24 (0) |
| MHRC | 51 | 0 | 22.2 (2.8) | 16-27 | 38 | 0 | 20.1 (2.5) | 16-28 | 3 | 0 | 19 (2.4) | 16-21 | 35 | 0 | 20.2 (2.5) | 17-28 | 7.9% | 30.2 (6.6) |
| MPRC | 20 | 40 | 17.8 (4.3) | 12-24 | 31 | 52 | 17 (3.2) | 12-22 | 3 | 33 | 16 (4.4) | 13-21 | 10 | 50 | 15.8 (2.7) | 12-20 | 23.1% | 7.1 (1.1) |
| Newcastle | 17 | 76 | 20.3 (1.9) | 17-24 | 45 | 53 | 19.6 (2.2) | 15-24 | 1 | 0 | 17.4 (0) | 17-17 | 41 | 54 | 19.6 (2.2) | 15-24 | 2.4% | 21.5 (12) |
| Oslo Region | 63 | 38 | 19.8 (3.6) | 15-29 | 21 | 38 | 19.9 (3.6) | 16-29 | 2 | 50 | 20 (5.4) | 16-24 | 18 | 39 | 19.9 (3.7) | 16-29 | 10% | 12.5 (1.4) |
| Pitt | 65 | 40 | 23 (5.9) | 14-40 | 26 | 54 | 20.8 (5.3) | 12-36 | 2 | 50 | 17.1 (0.8) | 16-18 | 11 | 64 | 19.5 (6.4) | 12-36 | 15.4% | 13.1 (3.6) |
| RUMC | 29 | 66 | 23.9 (3.2) | 18-30 | 67 | 46 | 22.7 (3.9) | 16-30 | 4 | 50 | 24.5 (2.9) | 21-27 | 60 | 47 | 22.6 (3.9) | 16-30 | 6.2% | 8.3 (4.5) |
| Singapore | 53 | 47 | 22 (4.2) | 14-30 | 100 | 32 | 21.9 (3.6) | 14-30 | 11 | 27 | 20.1 (3.1) | 15-26 | 88 | 32 | 22.1 (3.6) | 14-30 | 11.1% | 18.9 (5.9) |
| SNUH | 74 | 32 | 21.2 (2.5) | 17-27 | 74 | 26 | 20.7 (3.8) | 15-34 | 9 | 33 | 22 (5) | 16-33 | 46 | 24 | 20.4 (3.7) | 15-34 | 16.4% | 30.9 (17.2) |
| Stavanger | 33 | 48 | 17 (3.1) | 13-27 | 37 | 59 | 16.6 (2.4) | 13-25 | 5 | 80 | 16.2 (2.7) | 13-19 | 31 | 58 | 16.7 (2.3) | 14-25 | 13.9% | 24 (0) |
| Toho | 16 | 50 | 23.2 (2.9) | 18-28 | 40 | 70 | 23.7 (6.9) | 13-39 | 4 | 75 | 19 (4.4) | 14-24 | 36 | 69 | 24.2 (7) | 13-39 | 10% | 12 (0) |
| Tokyo | 25 | 48 | 22.1 (2.8) | 16-25 | 39 | 46 | 20.9 (3.5) | 14-29 | 3 | 0 | 22.7 (4.7) | 19-28 | 25 | 60 | 20.5 (3.1) | 14-27 | 10.7% | 26.5 (12.1) |
| Toronto | 39 | 41 | 25.5 (5.2) | 18-38 | 27 | 44 | 20.8 (1.8) | 18-27 | 0 | • | • | • | 4 | 50 | 19.7 (1.1) | 18-21 | • | 24 (0) |
| Toyama | 141 | 48 | 25.1 (4.2) | 18-38 | 79 | 44 | 18.5 (4) | 13-31 | 10 | 30 | 20.8 (5.7) | 15-31 | 69 | 46 | 18.1 (3.7) | 13-30 | 12.7% | 37.4 (34.2) |
| UCSF | 103 | 42 | 23.7 (7.5) | 13-40 | 71 | 48 | 19.3 (4.4) | 12-32 | 13 | 38 | 21.4 (4.4) | 16-29 | 47 | 49 | 19 (4.1) | 12-29 | 21.7% | 21 (5.4) |
| Zurich | 43 | 51 | 22.2 (5.6) | 13-36 | 62 | 40 | 19.1 (4.9) | 13-35 | 11 | 18 | 18.5 (3.4) | 14-24 | 35 | 40 | 19.1 (5.3) | 13-35 | 23.9% |  |

Follow-up times reported in months. “•” Indicates that data was not available, either because no CHR subjects transitioned to a psychotic disorder or follow-up data was not available. Site name abbreviations as follows: Amsterdam = Vrije Universiteit Amsterdam, The Netherlands; Barcelona-HSJD

### Image Acquisition and Processing

Thirty-one sites contributed T1-weighted MRI brain scans from 50 MRI scanners (**eTable 2** for MRI acquisition details). We conducted power calculations to estimate the sample size necessary to detect statistically significant effects (see **Supplement** for details).

We extract structural gray matter measures of 68 CT, 68 SA measures, and 16 subcortical volume measures using the FreeSurfer analysis software^75–78^. We implemented the ENIGMA consortium quality assessment pipeline^70,73,79–82^ (**Supplement** for further details).

Prior to conducting the mega-analysis, all neuroimaging data were adjusted for scanner protocol effects using neuroComBat^83^, a modified version of ComBat^84^ that increases the statistical power and precision of neuroimaging effect size estimates^83^.

### Statistical Analyses

#### Group and conversion-related differences in structural neuroimaging metrics

We assessed group differences using general linear models (GLMs) within a mega-analysis framework, with each sMRI measure (i.e., regional CT, SA or subcortical volume) as the dependent variable and group (HC/CHR) or conversion status (CHR-PS+/CHR-PS-/HC) as the independent variable. We included age, age^2^, sex, and estimated total intracranial volume (ICV) as covariates in all models, and corrected for multiple comparisons (N=155) using the False Discovery Rate (FDR^85^) method. We considered *q*-values<0.05 statistically significant.

For all structural neuroimaging measures, we conducted pairwise comparisons and calculated Cohen’s *d* effect sizes between two (CHR vs. HC) or three groups of interest (CHR-PS+ vs. HC/CHR-PS+ vs. CHR-PS-/CHR-PS-vs. HC). We also conducted analyses within a meta-analysis framework and fitted linear mixed models of sMRI data (not adjusted with neuroCombat), with scanner as a random effect. We investigated sMRI differences associated with the specific psychosis-risk syndromes (e.g., Attenuated Positive Symptom Syndrome; details in **Supplement**).

To evaluate the stability of group and conversion status differences, we performed analyses statistically controlling for psychotropic medication exposure at baseline. To assess effects of site, we conducted jackknife resampling analyses, i.e., iteratively removing one site’s data and re-running respective analyses^86,87^. sMRI measures that failed to show a group or conversion status effect at *q*<0.05 in >10% of jackknife iterations (i.e., 4/31 sites) were considered “unstable”.

To assess the meaningfulness of obtained effect sizes we used two analytic approaches: equivalence testing (to assess whether observed differences fell within the upper and lower bounds of a predefined smallest effect size of interest, providing support for the *absence* of a meaningful effect) and minimal-effects testing (to assess whether observed effects *were greater* than the same pre-defined effect size^88^). Upper/lower bounds (representing the positive/negative predefined smallest effect size of interest) were set to *d* = +/- 0.15 (further details and effect size rationale, see **Supplement**).

#### Group and conversion-related differences in sMRI age-associations

We used general additive models^89,90^ to model group and conversion status differences in the relationship between age and sMRI measures. General additive models extend GLMs by allowing for a non-linear relationship between independent and dependent variables. Here, age is modeled as a smooth function (continuous derivatives), which is akin to the “age-related” slope in a GLM. Analyses were confined to sMRI measures that displayed statistically significant effects of I) HC vs. CHR (N=56 measures) and II) CHR-PS+ vs. HC/CHR-PS+ vs. CHR-PS-/CHR-PS- vs. HC (N=4). We examined the effect of baseline age, group/conversion status, and the interaction between the two variables. Sex and estimated ICV were included as covariates. Similar to prior work examining age effects during adolescent development^91,92^, we restricted our sample’s age range to 12-25 years (**eTable 3**). Details on post-hoc analyses for significant interaction effects are provided in the **Supplement**.

#### Comparison of psychosis conversion-related effects to other ENIGMA findings

We computed Spearman’s rank correlations to assess the extent to which the pattern of observed effect sizes (Cohen’s *d’s* for CHR-PS+ vs. HC and CHR-PS-) correlated with the pattern found in prior psychosis studies, specifically the ENIGMA Schizophrenia(SZ vs. HC^69,70^) and ENIGMA 22q11.2 Deletion Syndrome (22q11DS with psychosis vs. 22q11DS without psychosis^72,73^) Working Groups. As a control, we compared CHR-PS+/CHR-PS- vs. HC effect sizes to effect sizes published by the Major Depressive Disorder Working Group (MDD vs. HC^93^). Additional details of correlation analyses are reported in the **Supplement**.

## 3. RESULTS

### Sample Characteristics

Site demographics are reported in **Table 1**. See **eTables 4-6** for IQ and symptom comparisons. Few CHR participants reported typical (<1%) and/or atypical antipsychotic (12.4%) medication use (**eTable 7**).

### Widespread lower CT in CHR versus HC

Compared to HC, the CHR group had smaller global neuroimaging measures: estimated ICV (*d*=-0.13, CI=-0.2 to -0.06), mean CT (*d*=-0.18, CI=-0.25 to -0.11) and total SA (*d*=-0.15, CI=-0.22 to -0.08). Fifty-three additional regions exhibited a significant effect of group (*q*<0.05, **eTable 8**). The largest group effects were observed for widespread lower CT in CHR vs. HC (42/68 comparisons, *d* range=-0.09 to -0.17; **Figure 1A** for overview; **eTable 8** and **eFigure 1** for details). Few subcortical (3/16) and SA (8/68) group differences were observed (*d* range=- 0.09 to -0.16). No group-by-sex interactions were detected (all *q*>0.05).

**Figure 1.**
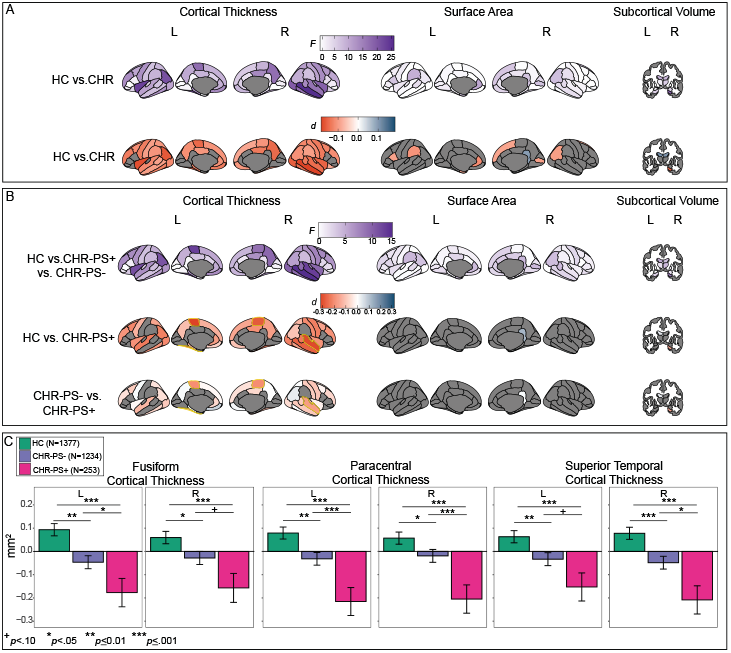
Effect sizes for mega-analysis of group and conversion status. **A**. Overview of effect sizes for HC vs. CHR. The top row reflects the results of the overall generalized linear model. A deeper purple color indicates a greater effect of group (HC vs. CHR) in this region. We observed the greatest effects of group in cortical thickness measures. The second row indicates the pairwise effect sizes for HC vs. CHR, in regions that were statistically significant (*q*<0.05) in the overall comparison (top row of **A**.). Regions that were not statistically significant in the overall comparison are in gray. In comparison to HC, CHR exhibited lower cortical thickness across the cortex. Red color indicates that HC has a larger value in comparison to CHR for this region. **B**. Overview of effect sizes for HC vs. CHR-PS+ vs. CHR-PS-. The top row reflects the results of the overall generalized linear model. A deeper purple color indicates a greater effect of conversion status (HC vs. CHR-PS+ vs. CHR-PS-) in this region. The second and third rows indicate the pairwise effect sizes for HC vs. CHR-PS+ and CHR-PS- vs. CHR-PS+, respectively. Pairwise comparisons are presented in regions that were statistically significant (*q*<0.05) in the overall comparison (top row of **B**.). Regions that were not statistically significant in the overall comparison are in gray. Regions that CHR-PS+ had lower cortical thickness in comparison to HC and CHR-PS-, CHR-PS+ are highlighted in yellow. **C**. Bar graphs for regions in which CHR-PS+ (pink) had lower CT in comparison to CHR-PS- (purple) and HC (green).

Group difference magnitudes remained similar when controlling for antipsychotic and psychotropic medication use (**eTable 9**). 91% of group differences remained statistically significant (*q*<0.05) in >90% of jackknife resampling iterations (eFigure 2), providing evidence that site effects were not driving results. Equivalence testing revealed that effect sizes for group differences were likely not meaningfully greater than our *a priori*-defined smallest effect size of interest in 3/56 measures (**eFigure 1, eTable 10**).

**Figure 2.**
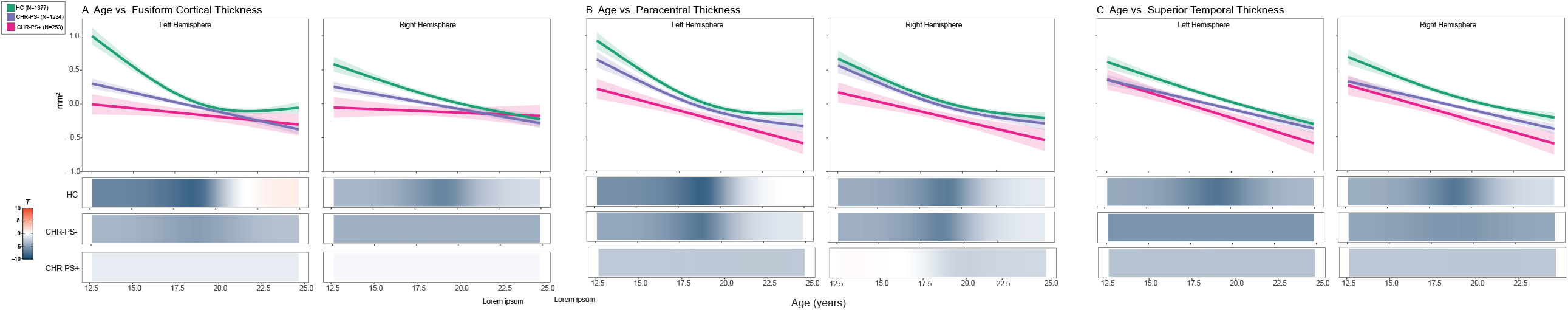
Age effects of regions that exhibited an effect of conversion status. HC are in green, CHR-PS+ are pink, and CHR-PS-are purple. Each graph has a line of best fit for the effect of age on the respective neuroimaging measures. Shading around the line indicates the standard error. The bars underneath the age plots reflect the derivative of the slope, i.e., the rate of change taking place at a particular age, scaled as a pseudo *t*-statistic, based on the posterior simulation. Age effects are plotted for A. fusiform cortical thickness, B. paracentral cortical thickness, and C. superior temporal cortical thickness.

Mega- and meta-analyses of neuroimaging data prior to neuroCombat harmonization provided similar results (Spearman’s ρ>0.93; **eTable 11**). No sMRI measures were uniquely sensitive to psychosis-risk syndrome (see **Supplement** and **eTables 12-15** forresults).

### Thinner paracentral, fusiform, and superior temporal CT are associated with psychosis conversion

Forty-eight structural neuroimaging measures (N_CT_=37) exhibited a significant overall effect of psychosis conversion status (*q*<0.05, **Figure 1B**; **eFigure 3** and **eTable 16**). The three groups differed from each other on four structural neuroimaging measures: in comparison to HC and CHR-PS-, CHR-PS+ exhibited thinner cortex in bilateral paracentral, right superior temporal, and left fusiform regions (**Figure 1C**, average *d* of four sMRI measures=-0.22). CHR-PS+ and CHR-PS- (vs. HC) exhibited thinner cortex in the left superior temporal and right fusiform regions;similar trends were observed for CHR-PS+ vs. CHR-PS-differences (*p*<0.08; Figure 1C). Results remained stable when length of follow-up period was included as a covariate.

**Figure 3.**
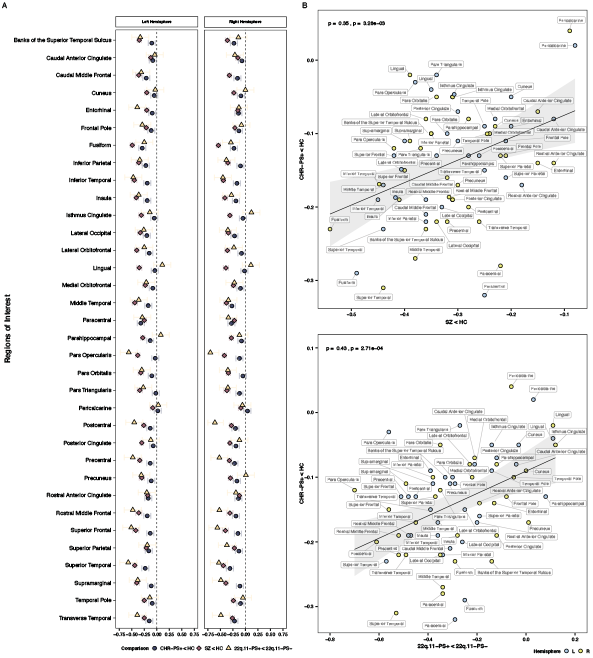
Cortical thickness alterations in CHR-PS+, schizophrenia, and 22q.11DS **A**. Magnitude of cortical thickness alterations for the three groups. Whole-brain cortical thickness effect sizes were not reported for SZ and 22q.11DS in original publications; values shown are the hemispheres with the smallest effect size. Colored bars represent standard error. **B**. Spearman’s rank correlations between cortical thickness alterations in CHR-PS+ and individuals with schizophrenia (top) and 22q.11 DS with psychosis (bottom).

Using minimal-effects testing, we observed that effect sizes for bilateral paracentral (L Z=-2.43, *p*=0.02; R Z=-1.86 *p*=0.06), right superior temporal (Z=-2.29, *p*=0.02) and left fusiform (Z=-2.00, *p*=0.05) in CHR-PS+ vs. HC were all significantly *greater* than 0.15, at least at trend level, underscoring the presence of notable group differences.

In the remaining follow-up comparisons, CHR-PS+ and CHR-PS-differed from HC at *p*<0.05, but there were no significant differences between CHR-PS+ and CHR-PS- (29/48 comparisons; **eTable 16**). We observed no conversion status-by-sex interactions (all *q*>0.05).

When we controlled for antipsychotic or psychotropic medication exposure, most overall psychosis conversion effects remained statistically significant (**eTable 17**). sMRI measures that differentiated the three groups showed an overall conversion effect at *q*<0.05 in all jackknife iterations, ruling out site effects as a potential confound (**eFigure 2**). No pairwise comparisons for structural neuroimaging measures that showed an overall conversion effect at *q*<0.05 were statistically equivalent to our predefined smallest effect size of interest (**eTable 18**). There were no statistically significant psychosis-risk syndrome-by-conversion status interactions (**eTable 19**).

### CHR-PS+ fail to show expected age-associations with fusiform and paracentral CT

There were no significant group-by-age interactions in the 56 neuroimaging measures that differed between CHR and HC (*q*>0.05, **eTable 20**). In contrast, 2/4 sMRI measures in which all groups differed (HC vs. CHR-PS+ vs. CHR-PS-) displayed a statistically significant conversion status-by-age interaction.

In left fusiform, age-CT associations differed between CHR-PS+ vs. HC (F=9.8, *p*=4.9e-05, *q*=5.9e-04) and CHR-PS- vs. HC (F=8.7, *p*=1.5e-04, *q*=9.1e-04, Figure 2A, left). Between ages 12-16, HC showed a stronger negative association between age and CT, compared to CHR-PS+ and CHR-PS-, suggesting a reduced rate of CT change in CHR-PS+ and CHR-PS-. Though the age-by-conversion status interaction was not statistically significant, a similar pattern emerged for the right fusiform CT (Figure 2A, right).

Age-effects in the left paracentral CT differed between CHR-PS+ vs. HC (F=5.9, *p*=4.9e-03, *q*=0.02, Figure 2B, left). Between 12-15.8 years of age, HC showed a stronger negative association between age and CT in comparison to CHR-PS+. Age-CT associations did not differ between CHR-PS- vs. HC (F=0.2, *p*=0.69). This pattern of results was not observed for the right paracentral CT (Figure 2B, right).

We found no significant age-by-conversion status interactions for the superior temporal CT (Figure 2C); all three groups showed negative CT-age associations.

### CT aberrations in CHR-PS+ resemble the pattern observed in SZ and 22q11DS with a psychotic disorder diagnosis, but not MDD

**Figure 3A** provides a visual overview of CT alterations in CHR-PS+ relative to SZ and individuals with 22q11DS and a psychotic disorder. The pattern of baseline CT alterations in CHR-PS+ (CHR-PS+ vs. HC effect sizes) correlated significantly with that observed in SZ (ρ=0.35, *p*_*permute*_=0.004, **Figure 3B** top) and individuals with 22q11DS and psychosis (ρ=0.43, *p*_*permute*_=0.001, **Figure 3B** bottom). CT alterations in CHR did *not* correlate with those observed in MDD (CHR-PS+ ρ=-0.03) and slopes for CHR-PS+/SZ and CHR-PS+/MDD correlations significantly differed (Steiger’s Z=2.06, *p*_*permute*_=0.008). No significant correlations were observed for SA (SZ ρ=-0.03; 22q.11DS ρ=-0.06, eFigure 5). Subcortical volume alterations in CHR-PS+ correlated with those observed in SZ (ρ=0.54, *p*=0.03) and a similar non-significant trend was observed for 22q11DS and psychosis (ρ=0.46, *p*=0.07). Associations for CHR-PS- (vs. HC) effect sizes were similar to those reported here (see **Supplement**).

## 4. DISCUSSION

We conducted the largest multisite neuroimaging investigation to date in CHR participants, examining baseline structural neuroimaging measures associated with later transition to psychosis. We found widespread thinner cortex in CHR, consistent with previously-reported CT alterations in individuals with an established psychotic disorder. Compared to CHR-PS- and HC, at baseline, CHR-PS+ exhibited thinner cortex in bilateral paracentral, right fusiform and left superior temporal regions, with effect sizes significantly greater than what we considered to be meaningful *a priori*. Our results were robust to effects of medication exposure, sex, site, and length of follow-up period. Findings from this international effort suggest that conversion to psychosis amongst those at high risk is associated with thinner cortex at baseline.

We identified widespread regional thinner cortex in CHR compared to HC. Thinner cortex has been observed in SZ, as well as other psychiatric disorders^69,79,93–95^. Importantly, the overall pattern of thinner cortex in CHR resembled that observed in SZ and individuals with 22q11DS and a psychotic disorder, but not in MDD. For CHR-PS+, correlations with SZ CT alterations were significantly greater than the relationship observed with MDD CT alterations. Taken together, our results suggest that the overall constellation of reported CT alterations resembles the regional pattern of alterations observed in SZ and genetic disorders associated with psychosis, and thus argues in favour of psychosis-specific thinner cortex in CHR

We also observed thinner cortex specific to psychosis conversion in paracentral, superior temporal and fusiform CT; CHR-PS+ exhibit significantly lower CT than CHR-PS- and HC in these regions. Lower baseline CT and/or volume in these regions has previously been reported in CHR-PS+^96,97^ (data not used here). Furthermore, longitudinal CT decreases in these regions have been associated with transition to psychosis in CHR^17,98,99^. The magnitude of altered CT in CHR-PS+ in paracentral, superior temporal and fusiform regions was highly consistent with findings in SZ^70,100–102^ and lower fusiform and paracentral CT has been observed in non-clinical voice hearers^103^. Thus, the observed pattern of CT alterations in CHR-PS+ is consilient with morphometric disruptions across the wider psychosis spectrum.

Consistent with previous CHR studies examining baseline neuroimaging associations with later conversion to psychosis^96,104^, we did not observe widespread subcortical volume or SA alterations associated with later psychosis transition. Taken together, these results suggest that CT reductions may be among the most widespread, robust, and specific morphometric changes associated with psychosis risk and conversion, compared to SA or subcortical volume.

The neuroanatomic pattern of group differences and age-associated disruptions observed in CHR may provide important insights into mechanisms underlying increased risk for psychosis. Pre-clinical models ^105,106^ and recent genome-wide association studies^107^ suggest that genetic variants associated with SZ are linked to the regulation of neural progenitor cells during fetal development, while genetic markers associated with CT were associated with regulatory processes in adulthood. Thus, CT alterations may be the end result of maladaptive maturation-related mechanisms that occur during post-fetal development, including proliferation, synaptic pruning and/or myelination^108–111^. Thinner CT, particularly in early adolescence^112,113^ (**Figure 2**), could reflect abnormal synaptic plasticity or pruning, which have both been implicated in *in vitro* SZ models^114^. Although excessive synaptic pruning is one plausible explanation for thinner cortex associated with psychosis transition, recent evidence suggests that intracortical myelination and/or expression of myelin-related genes may be mechanisms driving cortical thinning^115,116^. To better understand neurobiological mechanisms underlying psychosis transition in CHR, investigations of concomitant measures of cortical thickness, macroscale white matter tracts, and intracortical myelination are necessary.

Even if CT reductions in CHR were robust, effect sizes for between-group differences were nevertheless small-to-moderate and accounted for ∼1% of the variance in CHR-PS+ vs. CHR-PS-comparisons. The subtle nature of these morphometric alterations underscores the importance of adequate statistical power, achievable only through large-scale multi-site collaborations. Consistent with recent work showing that SZ polygenic risk scores only improved differentiation of CHR-PS+ and HC (not CHR-PS+ from CHR-PS-)^117^, we anticipate that baseline, univariate sMRI metrics will to have a similar impact on psychosis risk prediction algorithms. Given the logistic and financial challenges that MRI brings, the use of MRI metrics in isolation may not be feasible or useful for psychosis risk prediction. A viable solution may be to adopt sequential assessment frameworks, as recently implemented^34^. Alternatively, sMRI alterations may be a better predictor of general psychopathology, and would be better suited for transdiagnostic risk prediction models^118,119^.

### Limitations

One limitation common to multi-site studies is that data were collected from multiple scanners, although leave-one-out analyses suggest that site effects were not prominent. Secondly, this initial study focused on baseline cross-sectional data, and did not investigate progressive sMRI changes (trajectories) associated with psychosis conversion, as identified in prior work^17,19,38,97–99,120–123^. This is a future goal of the ENIGMA CHR Working Group, now that feasibility of this collaboration has been established.

## Conclusions and Future Directions

In the largest study of brain abnormalities in CHR to date, we found robust evidence for a subtle, widespread pattern of CT alterations, consistent with observations in psychosis. The specificity of these alterations to CT - as well as age-associated deviations in regions sensitive to psychosis conversion - may point to abnormal development processes. These findings also point to age ranges (i.e., early adolescence) when morphometric abnormalities in CHR might be greatest.

## Data Availability

All code used in this project is publicly available here: https://osf.io/p9ntr/ Due to individual site IRB restrictions, data is not publicly available. Please contact the ENIGMA CHR Working Group co-chairs if you are interested in joining and participating in the working group. Then you will be able to submit secondary data proposals.

## Acknowledgements

BHE (Bjørn H. Ebdrup) is part of the Advisory Board of Eli Lilly Denmark A/S, Janssen-Cilag, Lundbeck Pharma A/S, and Takeda Pharmaceutical Company Ltd; and has received lecture fees from Bristol-Myers Squibb, Otsuka Pharma Scandinavia AB, Eli Lilly Company, Boehringer Ingelheim Danmark A/S, and Lundbeck Pharma A/S. DH (Dennis Hernaus) has received financial compensation as a consultant for P1vital Produicts Ltd. F-RM (Francisco Reyes-Madrigal) has received compensation as a speaker for Janssen (Johnson & Johnson). All reported activities were unrelated to the work presented in this manuscript.

## Table and Figure Legends

**Table 1.** Demographic descriptives for every site in HC and CHR, and CHR-PS+ and CHR-PS- The Ns for CHR-PS+ and CHR-PS- do not always sum to the sample CHR N because most sites lose individuals to follow-up (CHR-UNK).

