## supplemental text & tables for "Thinner cortex is associated with psychosis onset in individuals at Clinical High Risk for Developing Psychosis: An ENIGMA Working Group mega-analysis"

### Table of Contents

**Supplementary Text**

Supplemental Methods
Supplemental Results

**Supplementary Tables**

**eTable 1.** Inclusion and exclusion participant criteria per site
**eTable 2.** Scanner and acquisition parameters by site
**eTable 3.** Age and sex of HC vs. CHR vs. CHR-PS+ vs. CHR-PS- in 12-25 year old age range (Age-associated analyses)
**eTable 4.** Intelligence quotient comparisons of HC vs. CHR by site
**eTable 5.** Intelligence quotient comparisons of CHR-PS+ vs. CHR-PS- vs. CHR-UNK by site
**eTable 6.** Symptom differences between CHR groups (CHR-PS+, CHR-PS-, CHR-UNK) as measured by the SIPS or CAARMS
**eTable 7.** CHR current medication information reported by site
**eTable 8.** CHR vs. HC effect size overview (post-ComBat mega analysis)
**eTable 9.** CHR vs. HC antipsychotic medication effects
**eTable 10.** CHR vs. HC equivalence testing results
**eTable 11.** CHR vs. HC effect size overview (pre-ComBat mega and meta analysis)
**eTable 12A.** CHR subgroup (APS vs. BIPS vs. GRD vs. HC) effect size overview
**eTable 12B.** CHR subgroup (APS vs. BIPS vs. GRD vs. HC) effect size overview (continued)
**eTable 13.** APS subgroup (APS vs. no APS assignment vs. HC) effect size overview
**eTable 14.** BIPS subgroup (BIPS vs. no BIPS assignment vs. HC) effect size overview
**eTable 15.** GRD subgroup (GRD vs. no GRD assignment vs. HC) effect size overview
**eTable 16**. CHR-PS+ vs. CHR-PS- vs. HC effect size overview (post-ComBat mega analysis)
**eTable 17.** CHR-PS+ vs. CHR-PS- vs. HC antipsychotic medication effects
**eTable 18.** CHR-PS+ vs. CHR-PS- vs. HC equivalence testing results
**eTable 19.** CHR subgroup-by-psychosis conversion interaction analyses effect size overview
**eTable 20.** Age effects of CHR vs HC
**eTable 21.** Age effects on conversion status

**Supplementary Figures**

**eFigure 1.** CHR vs. HC effect size overview (post-ComBat mega analysis)
**eFigure 2.** Jackknife resampling analysis for neuroimaging regions meeting statistical significance (q<.05) in Aim 1
**eFigure 3.** CHR-PS+ vs. CHR-PS- vs. HC effect size overview (post-ComBat mega analysis)
**eFigure 4.** CHR-PS+ vs. SZ vs. 22q11DS effect size overview
**eFigure 5.** Surface Area, Subcortical Volume and global measure correlations between CHR-PS+, SZ, and 22q11DS

**Supplementary Citations**

[1,2]

***Supplemental Methods***

***CHR Inclusion/Exclusion Criteria: CAARMS***

Eight hundred twenty one CHR participants were included in this study based on their responses to the Comprehensive Assessment of At-Risk Mental States (CAARMS[1,2]) The CAARMS is a semi-structured interview administered by trained clinician. The CAARMS consists of seven subscales: positive symptoms, cognitive change, emotional disturbances, negative symptoms, behavioral change, motor change, and general psychopathology. The positive symptom subscale and an assessment of social and occupational functioning are used to determine CHR eligibility and syndrome subgroup (see below).

The positive symptom subscale consists of four positive symptoms (P1=unusual thought content, P2=non-bizarre ideas, P3=perceptual abnormalities, P4=disorganized speech). Each symptom is rated on a scale from 0-6, where a score of "0" means that the symptom is not present and "6" means that the symptom is endorsed at a fully psychotic level. CHR participants met criteria for at least one of three syndromes: 1) Attenuated Psychotic Symptoms, 2) Brief Limited Intermittent Psychotic Symptoms, and/or 3) a trait/vulnerability condition.

A CHR participant meets criteria for Attenuated Psychotic Symptoms when a) any one of the positive symptoms (P1-P4) is rated within a specific subthreshold range (i.e., P1 & P2=3-5, P3=3-4, P4=4-5) and occurs at least once a month or b) the severity of any one of the positive symptoms is rated a "6" on P1, P2, or P4 or at least a "5" on P3, but occurs no more than twice a week for longer than an hour, or occurs no more than six times a week, for less than an hour. The symptoms should have been present in the previous twelve months, and not for longer than five years. A CHR participant meets criteria for Brief Limited Intermittent Psychotic Symptoms when the severity of any one of the positive symptoms is rated a "6" (or at least a "5" on P3), but occurring at a frequency of at least three times a week for greater than an hour or occurring daily for less than an hour. Similar to Attenuated Psychotic Symptoms, the symptoms should have been present in the previous twelve months, and not for longer than five years. A CHR participant meets the trait/vulnerability condition if they have 1) a first degree relative with a psychotic disorder or 2) meets criteria for schizotypal personality disorder. For all risk syndromes, CHR participants must have experienced either 1) at least a 30% drop in functioning (as measured by the SOFAS[3]) within the last year and they have to maintain this level for a minimum of a month or they 2) receive a SOFAS score <50 for the past 12 months or longer.

***CHR Inclusion/Exclusion Criteria: SIPS***

Nine-hundred seventy one CHR participants were included in this study based on their responses to the Structured Interview for Prodromal Syndromes/Structured Interview for Psychosis-Risk Syndromes (SIPS[4,5]). The SIPS is a semi-structured interview administered by a trained clinician. The SIPS is made up of four subscales: positive, negative, disorganization, and general symptoms. The position symptom subscale is used to determine CHR eligibility. The positive symptom subscale consists of five positive symptoms (P1=unusual thought content/delusional ideas, P2=suspiciousness/persecutory ideas, P3=grandiose ideas, P4=perceptual abnormalities/hallucinations, P5=disorganized communication). Each symptom is rated on a scale from 0-6, where a score of "0" means that the symptom is not present and "6" means that the symptom is endorsed at a fully psychotic level. CHR participants met criteria for at least one of three syndromes : 1) Attenuated Positive Symptom Syndrome, 2) Brief Intermittent Psychotic Symptom Syndrome, and/or 3) Genetic Risk and Deterioration Syndrome. An individual meets Attenuated Positive Symptom Syndrome criteria when any one of the positive symptoms (P1-P5) is rated as "subthreshold": the participant receives a score of 3 (moderate), 4 (moderately severe), or 5 (severe but not psychotic). Positive subthreshold symptoms must have started or gotten worse in the last year. Positive subthreshold symptoms must occur at a frequency of at least one time per week in the last month. An individual meets Brief Intermittent Psychotic Symptom Syndrome criteria when one of the positive symptoms is rated a 6 (a severe and psychotic level) for a brief period of time (~several minutes at a time). Positive symptoms must have started in the past 3 months and must have occurred at least once in the last month. An individual meets Genetic Risk and Deterioration Syndrome criteria when they have a first-degree relative who has a psychotic disorder and they have experienced a 30% decrease in functioning over the past year (as measured by the Global Assessment of Functioning).

***Combining CHR Risk Syndromes from CAARMS and SIPS*** We combined the CAARMs and SIPS psychosis-risk syndromes when we analyzed the syndrome's effects. Attenuated Psychotic Symptoms (CAARMS) and Attenuated Positive Symptom Syndrome (SIPS) were combined into the "APS" psychosis-risk syndrome group. We combined Brief Limited Intermittent Psychotic Symptoms (CAARMS) and Brief Intermittent Psychotic Symptom Syndrome (SIPS) into the "BIPS" psychosis-risk syndrome group. Finally, the trait/vulnerability condition (CAARMS) and Genetic Risk and Deterioration Syndrome (SIPS) were combined into the "GRD" group.

***CHR Exclusion Criteria: All Sites*** Participants were not included in the study if they currently or previously endorsed or presented with positive symptoms at a psychotic level of intensity.

***Power calculations***

To determine statistical power for Aims 1 and 2, we conducted 1,000 Monte Carlo simulations of the proposed models, based on preliminary data distributions. Sample sizes reported are based on 80% power to achieve the desired effect. In our preliminary data, we observed 0.15-0.19 standard deviation change in group status for every one standard deviation change in structural neuroimaging metrics (N~150 CHR). For **aim 1** (HC vs. CHR), we observed in simulation that we would need a minimum sample size N=750 (40% HC, 60% CHR) to detect a group difference of beta=0.17. For **aim 2** (CHR-PS+ vs. CHR-PS-), we determined that we would require a CHR sample size of N=1200 with an estimated 20% conversion status rate to detect a group difference of beta=0.17.

***Pre-ComBat mega analysis and random-effects meta analysis***

All 155 GLMs (i.e. for every structural neuroimaging measure; CT in 68 regions, SA in 68 regions, subcortical volume in 16 regions and 6 whole-brain measures) were re-run using data prior to correction for site effects using neuroComBat using a 1) mega analysis framework ("pre-ComBat mega-analysis"), and, 2) meta analysis framework ("pre-ComBat meta-analysis"), to allow comparisons to previous work[6–8] and to ensure robustness of results. For the pre-ComBat mega analysis, scanner was treated as a random effect. To obtain the site-specific effect size of interest for the pre-ComBat meta analysis, each GLM was run separately for every study site, with scanner as an additional covariate. Study site-specific effect sizes were then entered into a random-effects meta analysis to obtain the effect size for group and conversion status differences in structural neuroimaging measures. All meta analyses were conducted using the metafor package (version 2.4.0[9]) in R.

***Aim 1: CHR syndrome analyses***

Out of the 1792 CHR participants, 142 CHR participants did not have CHR subgroup assignments and were removed from all subsequent analyses. For all structural neuroimaging metrics, we first tested the main effect of CHR subgroup (APS (N=1320) vs. BIPS (N=50) vs. GRD (N=98) vs. HC) in participants with only one subgroup assignment (CHR N=1468; HC N=1377). To include CHR participants with multiple subgroup assignments (CHR N=1650; HC N= 1377), we also ran a series of linear models assessing main effects of the individual subgroup assignment (APS (N=1500) vs. no APS assignment (N=150) vs. HC (N=1377); BIPs (N=91) vs. no BIPS assignment (N=1559) vs. HC (N=1377); GRD (N=249) vs. no GRD assignment (N=1401) vs. HC(N=1377)).

***Aim 2: CHR syndrome by conversion status analyses***

We also assessed the interaction between psychosis conversion status (CHR-PS+ vs. CHR-PS- vs. HC) and CHR subgroup (APS vs. BIPS vs. GRD vs. HC) in participants with one subgroup assignment and conversion status data. Additional linear models assessing interactions between conversion status (CHR-PS+ vs. CHR-PS- vs. HC) and individual subgroup assignment (APS vs. no APS vs. HC; BIPs vs. no BIPS vs. HC; GRD vs. no GRD vs. HC) were also run.

Similar to the analyses reported in the main text of the manuscript, we included age, $age^2$, sex, and estimated total ICV as covariates in all models. ICV was not used as a covariate for three whole-brain measures: mean CT , total SA, and estimated ICV.

***Equivalence testing bounds***

We used equivalence testing[10] to assess whether observed sMRI effect sizes were not meaningfully greater than zero. To these aims, two one-sided t-tests were conducted against the upper and lower equivalence bounds of a predefined smallest effect size of interest. If the observed effect size's 90% confidence interval falls within the upper and lower equivalence bounds, the presence of a meaningful or "worthwhile" effect can be rejected[11]. We conducted these tests on all regions displaying an overall effect of group or conversion.

For regions displaying a conversion effect, and in which CHR-PS+ differed from CHR-PS- and HC, we conducted minimal effects testing. The aim of this analyses was to investigate whether any of the observed effects in CHR-PS+ were meaningfully greater than the smallest effect size of interest, which would emphasize their potential for use as e.g. a predictor in risk prediction models in future studies.

All upper and lower equivalence bounds (representing the positive and negative smallest effect size of interest) were set to Cohen's *d* = +/- 0.15. Thus, for all equivalence tests, we investigated whether any of the observed effect sizes fell within the 0.15 - -0.15 range. We based these bounds on our *a priori* power calculations (see above) indicating 80% power to detect an effect of conversion status of Cohen's *d*=~0.17 in a sample of 1200 CHR, assuming a transition rate of 20%. Moreover, this effect size is highly similar to the average effect size for SZ-HC group differences in cortical thickness reported in a recent large-scale meta-analysis[7] (Cohen's *d*=.154). Equivalence tests[12] were conducted using the TOSTER package (version 0.3.4) in R.

***GAM Post-Hoc Analyses***

When statistically significant group-by-smoothed age interactions were observed ($q<0.05$), we assessed for pairwise differences in the smoothed effect of age, by taking the difference between the two smooths and considered an age range where 95% confidence intervals did not overlap as a statistically significant effect. This analysis would be akin to comparing the strength of the slopes between two groups in a linear model; however, the GAM extends this approach by identifying more restricted age ranges where smoothed effects of age (age-related slope in GLM) differ. Similar to Aim 1, we re-ran all GAM analyses covarying for antipsychotic and psychotropic medication status. We also employed jackknife analyses to assess the stability of observed age-associated group differences.

We also performed a posterior simulation on the first derivative of the GAM fits in each group separately to identify periods of age-associated changes. Following previous work[13] and established guidelines[14], we used a multivariate normal distribution whose vector of means and covariance were defined by the fitted GAM parameters to simulate 10,000 GAM fits and their first derivatives (generated at 0.1 year age intervals). Significant intervals of age-related change in a structural neuroimaging measure were defined as ages when the confidence intervals (95%) of simulated GAM fits did not include zero ($p < 0.05$).

***Correlation Analyses***

For all correlations we used Spearman's rank correlations. To investigate whether the difference in correlation coefficients between CHR-PS+-SZ CT effect sizes and CHR-PS+-MDD CT effect sizes was significant (see main text, aim 3), we used Steiger's Test[15] for dependent correlations. Next, we used Monte Carlo permutation testing to generate a null distribution of correlation coefficients, and effect sizes representing the difference in correlations, by randomly shuffling correlation coefficients ($n_{permutations}=2000$, $\alpha_{permutate}=0.05$). All correlation analyses were carried out using the cocor package[16] (v1.1.3) in R.

##### *Supplemental Results*

##### *Aim 1 Psychosis-Risk Syndrome Results*

When examining differences in structural neuroimaging measures between CHR subgroups in participants with only one subgroup assignment, 13 brain regions passed correction for multiple comparisons (eTable 11 A/B). In all these regions, except for the lateral ventricles, group differences were driven by reductions in the APS group in comparison to HC, and often also by reductions in the APS group compared to BIPS and GRD.

When the main of effect of APS subgroup (vs. no APS assignment vs. HC) was assessed, there were 33 structural neuroimaging measures that passed correction for multiple comparisons (eTable 12). In 32 of these regions, APS-CHR exhibited significantly reduced CT, SA, or subcortical volume in comparison to HC. In a subset of seven structural neuroimaging measures, APS-CHR additionally exhibited reductions compared to CHR participants that did not have an APS assignment: bilateral paracentral (left: *d*=-0.18, CI=-0.35 - -0.01, *p*=0.04, right: *d*=-0.19, CI=-0.36 - -0.02, *p*=0.03), right caudal middle frontal (d=-0.19, CI=-0.36 - -0.02, p=0.03), right superior parietal (*d*=-0.20, CI=-0.36 - -0.03, *p*=0.02), right inferior parietal (*d*=-0.18, CI=-0.35 - -0.01, *p*=0.04) and left precuneus (*d*=-0.22, CI=-0.39 - -0.05, *p*=0.01) CT, as well as right temporal pole SA (*d*=-0.25, CI=-0.42 - -0.08, *p*=0.004). CT and SA reductions in CHR without APS assignment compared to HC were observed in right superior temporal CT (*d*=-0.20, CI=-0.37 - -0.03, *p*=0.02), left supramarginal SA (*d*=-0.18, CI=-0.35 - -0.01, *p*=0.04) and increased SA was observed in right temporal pole (*d*=0.27, CI=0.10 - 0.44, *p*=0.002).

Importantly, all regions identified in APS analyses were also identified in the HC vs. CHR analyses described in the main text, with highly similar effect sizes (for these results, see eTable 7).

Analyses of the main effect of BIPS subgroup assignment revealed 23 structural neuroimaging measures that passed correction for multiple comparisons (eTable 13). These results were primarily driven by CT, SA, or subcortical volume reductions in CHR individuals without BIPS assignment (the majority being CHR-APS) compared to HC, which were significant in all 23 regions. Group differences between all three groups were not observed in any structural neuroimaging measure. In comparison to BIPS-CHR, CHR individuals without BIPS assignment did not exhibit statistically significant differences in any of these 23 regions (all *p*>0.09). CT and SA reductions in CHR-BIPS compared to HC were observed in a subset of structural neuroimaging measures; right insula CT (d=-0.25, CI=-0.47 - -0.04, p=0.02), left inferior parietal CT (*d*=-0.25, CI=-0.46 - -0.04, *p*=0.02) and right pericalcarine SA (*d*=-0.26, CI=-0.48 - -0.04, *p*=0.02).

For the main effect of GRD subgroup assignment, 22 brain regions passed correction for multiple comparisons (eTable 14). Similar to the BIPS main effect analyses, CHR individuals without GRD assignment (again, the majority being CHR-APS) showed significant CT, SA, or subcortical volume reductions compared to HC in 21 of these regions. Group differences between all three subgroups were not observed in any structural neuroimaging measure. In comparison to CHR individuals without GRD assignment, CHR-GRD only exhibited significantly lower right inferior parietal SA (*d*=-0.18, CI=-0.32 - -0.05, *p*=0.009). In 12/22 of these regions, GRD-CHR exhibited significant reductions in CT, SA, or subcortical volume in comparison to HC (*d* range=-0.25 - -0.14).

In summary, the CHR subgroup main effect analyses reported above reveal a pattern of results in CHR-APS that is similar to the widespread pattern of structural neuroimaging changes in CHR vs. HC. Given that the great majority of CHR (91%) met criteria for APS syndrome, we speculate that the CHR subgroup results reconfirm the HC vs. CHR analyses, rather than supporting the existence of structural neuroimaging changes that are unique to APS, compared to BIPS or GRD. This interpretation is moreover confirmed by the observation that CHR subgroup main effect analyses only revealed changes in structural neuroimaging measures in APS that were also observed in the psychosis risk (CHR vs. HC) analyses; no regions uniquely associated with APS (and not psychosis risk) were observed.

##### *Aim 2 Psychosis-Risk Syndrome by Conversion Status Results*

After correcting for multiple comparisons, there were no significant interactions between CHR subgroup and psychosis conversion status; not when running the interaction analysis in CHR with only one subgroup assignment (eTable 18 “all-by-conversion) or when each subgroup status was examined individually (eTable 18 “APS/BIPS/GRD-by-conversion”).

##### *Aim 3 CHR-PS- correlations*

CHR-PS- (vs. HC) effect sizes for CT were significantly correlated with those observed in SZ ($\rho=0.35$, $p=0.003$) and individuals with 22q.11DS and psychosis ($\rho=0.31$, $p=0.009$). No CHR-PS- - SZ correlations were observed for SA ($\rho=0.17$), although a correlation was observed with 22q11DS with psychosis ($\rho=0.35$, $p<0.001$). Subcortical volume reductions in CHR-PS- correlated with those observed in SZ ($\rho=0.71$, $p<0.001$) and individuals with 22q11Ds and psychosis ($\rho=0.64$, $p=0.007$).

**eTable 1.** Inclusion and exclusion participant criteria per site

| Full Site Name | Abbreviated Site Name | All Subjects Major Inclusion Criteria | CHR Inclusion Criteria | HC Inclusion Criteria | Previous publications |
|---|---|---|---|---|---|
| Vrije Universiteit Amsterdam, The Netherlands | Amsterdam | • 16-31 years old<br>• No spasticity<br>• Sufficient command of the Dutch language<br>• IQ ≥80 | • Met CHR inclusion criteria via CAARMS<br>• Within 1 year of CHR assessment | • CAARMS and SOFAS score <55<br>• No family history of psychiatric disorders | 17, 18, 19 |
| Hospital Sant Joan de Déu Barcelona, Barcelona, Spain | Barcelona-HSJD | • 10-17 years old<br>• No lifetime neurological disorder or traumatic brain injury diagnosis<br>• Fluent in Spanish or Catalan<br>• IQ >70 | • Met CHR inclusion criteria via SIPS<br>• Some participants were also considered "Genetic Risk and Deterioration Syndrome" if they had an affected second-degree relative (82)<br>• Some participants also met criteria for Attentuated Negative Symptom Psychosis-Risk Syndrome (83)<br>• No autism spectrum disorder | • No current psychiatric disorder diagnosis<br>• No current psychotropic medication use<br>• No lifetime psychotic disorder diagnosis<br>• No first- or second-degree relative with psychotic disorder diagnosis | None |
| University Hospital of Child and Adolescent Psychiatry and Psychotherapy, University of Bern, Switzerland | Bern | • 8-40 years old<br>• No current neurological disorder diagnosis<br>• Fluent in German, French or English<br>• IQ >70 (via clinical impression) | • Met CHR inclusion criteria via SIPS<br>• Some participants also met criteria for basic symptom CHR via the SPI-A (81) | • No lifetime psychotic disorder diagnosis | 20 |
| New York State Psychiatric Institute, Columbia University, USA | Columbia1 | • 15-35 years old<br>• No major medical or neurological disorder diagnosis<br>• IQ >70<br>• No lifetime substance use disorder<br>• No lifetime history of traumatic head injury<br>• No imminent risk of harm to self or others | • Met CHR inclusion criteria via SIPS | • No SCID Axis I and/or Axis II Cluster C disorder diagnosis<br>• Not adopted | 21 |
| New York State Psychiatric Institute, Columbia University, USA | Columbia2 | • 18-30 years old<br>• No lifetime major medical or neurological disorders<br>• IQ >70 | • Met CHR inclusion criteria via SIPS | • No lifetime DSM-IV psychiatric disorder diagnosis | None |
| New York State Psychiatric Institute, Columbia University, USA | Columbia3 | • 14-30 years old<br>• No lifetime major medical or neurological disorders<br>• IQ >70 | • Met CHR inclusion criteria via SIPS<br>• CHR symptoms do NOT occur solely in the context of substance use | • No lifetime psychiatric disorder diagnosis<br>• No lifetime history of meeting criteria for psychosis-risk syndrome via the SIPS<br>• No current substance abuse | 22 |
| Mental Health Center Copenhagen and CINS, Mental Health Center Glostrup, University of Copenhagen, Denmark | Copenhagen_1 | • 18-40 years old<br>• No organic brain disorder diagnosis (e.g., epilepsy, inflammatory brain disease)<br>• Fluent in Danish<br>• IQ >70<br>• Not currently pregnant | • Met CHR inclusion criteria via CAARMS<br>• No psychiatric symptoms that could be explained by a physical illness with psychotropic effect (e.g. delirium) or acute intoxication (e.g. cannabis use)<br>• No diagnosis of a serious developmental disorder (e.g. Asperger's syndrome)<br>• Lifetime neuroleptic exposure equivalent is less than total lifetime haloperidol dose of >50 mg<br>• No current mood stabiliser use or recreational use of ketamine | • No lifetime DSM-IV psychiatric disorder diagnosis | 23, 24 |
| Mental Health Center Copenhagen | | • 18-40 years old<br>• No organic brain disorder | • Met CHR inclusion criteria via CAARMS<br>• No psychiatric symptoms that could be explained by a physical illness with psychotropic effect | | |

| | | | | | |
|---|---|---|---|---|---|
| and CINS, Mental Health Center Glostrup, University of Copenhagen, Denmark | Copenhagen_2 | diagnosis (e.g., epilepsy, inflammatory brain disease)<br>• Fluent in Danish<br>• IQ >70<br>• Not currently pregnant | (e.g. delirium) or acute intoxication (e.g. cannabis use)<br>• No diagnosis of a serious developmental disorder (e.g. Asperger's syndrome)<br>• Not currently receiving methylphenidate | • No lifetime DSM-IV psychiatric disorder diagnosis | None |
| Central South University, China | CSU | • 13-30 years old<br>• No lifetime neurological disease or organic brain abnormalities (examined by neuroradiologist blinded to group allocation)<br>• Fluent in Chinese<br>• IQ >70<br>• Not currently pregnant<br>• No lifetime drug or alcohol dependence | • Met CHR inclusion criteria via SIPS<br>• No lifetime psychotropic medication use | | None |
| Institute of Neuroscience and Psychology, University of Glasgow, Scotland | Glasgow | • 16-35 years old<br>• No organic cause for presentation<br>• Fluent in English<br>• IQ >70 | • Met CHR inclusion criteria via CAARMS<br>• Some participants also met criteria for basic symptom CHR via the SPI-A (81) | • No current psychiatric disorder diagnosis<br>• No current substance abuse<br>• No first-degree relative with a psychotic disorder diagnosis | 25 |
| Heidelberg University Hospital, Germany | Heidelberg | • 14-18 years old<br>• No lifetime neurological disorder diagnosis<br>• Fluent in German<br>• IQ >85<br>• No illegal drug use in past 12 months | • Met APS CHR inclusion criteria via SIPS<br>• No lifetime DSM-IV Axis I diagnosis<br>• No psychotropic medication exposure | • No lifetime DSM-IV Axis-I disorder diagnosis<br>• No psychotropic medication prescription<br>• Does not meet APS psychosis risk syndrome via SIPS | None |
| August Pi i Sunyer Biomedical Research Institute, Barcelona, Spain | IDIBAPS | • 10-17 years old<br>• No lifetime neurological disorder or traumatic brain injury diagnosis<br>• Fluent in Spanish or Catalan<br>• IQ >70 | • Met CHR inclusion criteria via SIPS<br>• Some participants were also considered "Genetic Risk and Deterioration Syndrome" if they had an affected second-degree relative (82)<br>• Some participants also met criteria for Attentuated Negative Symptom Psychosis-Risk Syndrome (83)<br>• No autism spectrum disorder | • No current psychiatric disorder diagnosis<br>• No current psychotropic medication use<br>• No lifetime psychotic disorder diagnosis<br>• No first- or second-degree relative with psychotic disorder diagnosis | 26 |
| Icahn School of Medicine at Mount Sinai, USA | ISMMS | 15-35 years old<br>• No major medical or neurological disorder diagnosis<br>• No lifetime traumatic head injury<br>• Fluent in English<br>• IQ >70<br>• No MRI contraindications<br>• No lifetime substance use disorder<br>• No imminent risk of harm to self or others | • Met CHR inclusion criteria via SIPS | • No SCID Axis I diagnosis, Axis II Cluster C disorder<br>• Not adopted | 21, 27 |
| Institute of Psychiatry, Psychology & Neuroscience, King's College London, UK | London_1 | • 18-35 years old<br>• Male<br>• No lifetime neurological disorder diagnosis<br>• IQ >70<br>• No contraindications to intranasal oxytocin or placebo<br>• No acute intoxication on day of scanning<br>• No exposure to antipsychotics | • Met CHR inclusion criteria via CAARMS<br>• No current substance use disorder | • Not Applicable: No Healthy Controls recruited as part of this study | 28, 29 |
| | | • 18-35 years old | | | |

| | | | | | |
|---|---|---|---|---|---|
| Institute of Psychiatry, Psychology & Neuroscience, King's College London, UK | London_2 | • No lifetime neurological disorder diagnosis<br>• Fluent in English<br>• IQ within normal range<br>• No drug or alcohol dependence, as specified in DSM-IV<br>• No recreational drug use in past two weeks | • Met CHR inclusion criteria via CAARMS | • No lifetime psychiatric disorder diagnosis<br>• No current psychotropic medication use | 30, 31 |
| Maastricht University, The Netherlands | Maastricht | • 12-55 years old<br>• No lifetime neurological disorder diagnosis<br>• Fluent in Dutch<br>• IQ >70 | • Met CHR inclusion criteria via SIPS<br>• No lifetime psychotic disorder for more than one week, based on DSM-IV criteria<br>• Clear evidence that the psychosis-risk syndrome is NOT due to a (non-schizophrenia-spectrum) psychiatric disorder | • No DSM-IV psychiatric disorder diagnosis (lifetime)<br>• No psychotropic medication prescription (lifetime)<br>• No first- or second-degree relative with a psychotic disorder diagnosis | 32-34 |
| University of Melbourne, Australia | Melbourne | • 14-30 years old<br>• No history of substantial head injury, seizures, neurologic diseases, impaired thyroid function, corticosteroid use<br>• Fluent in English<br>• No alcohol or substance abuse or dependence | • Met CHR inclusion criteria via CAARMS<br>• Absence of known organic cause for presentation<br>• Lifetime neuroleptic exposure is less than total lifetime haloperidol use of >15mg | • No lifetime psychiatric disorder diagnosis<br>• No family history of psychotic illness | 35-38 |
| Instituto Nacional de Neurología y Neurocirugía, Mexico City, Mexico | Mexico City | • 13-34 years old<br>• No lifetime neurological disorder or traumatic brain injury diagnosis | • Met CHR inclusion criteria via SIPS<br>• No other major psychiatric disorder diagnosis | • No lifetime psychotic disorder diagnosis<br>• No lifetime psychiatric disorder diagnosis<br>• No first- or second-degree relative with psychotic disorder diagnosis | 39-42 |
| Mental Health Research Center Moscow, Russia | MHRC | • 16-28 years old<br>• Male<br>• Right-handed<br>• No lifetime neurological disorder diagnosis<br>• No lifetime drug or alcohol dependence | • Met APS or BIPS CHR inclusion criteria via SIPS<br>• No lifetime mental or behavioural disorders due to psychoactive substance use (ICD-10 F10-F19) | • No lifetime psychiatric disorder diagnosis<br>• No lifetime psychotropic medication prescription<br>• No family history of psychiatric or neurological disorders | 43,44 |
| Maryland Psychiatric Research Center, University of Maryland School of Medicine, USA | MPRC | • 15-35 years old<br>• No major medical or neurological disorder diagnosis<br>• No lifetime traumatic head injury<br>• Fluent in English<br>• IQ >70<br>• No MRI contraindications<br>• No lifetime substance use disorder<br>• No imminent risk of harm to self or others | • Met CHR inclusion criteria via SIPS | • No lifetime psychiatric disorder diagnosis | 45,46 |
| University of Newcastle, Australia | Newcastle | • 13-25 years old<br>• No antipsychotic pharmacotherapy<br>• IQ ≥70<br>• No lifetime drug abuse or dependence diagnosis<br>• No head injury with loss of consciousness (>15 mins)<br>• No organic brain impairment<br>• No hearing impairment (>20dB SPL)<br>• No history of nasal trauma. | • Met CHR inclusion criteria via CAARMS<br>• Drop of 30% in Global Assessment of Functioning (GAF; past 12 months) | • No DSM-IV psychiatric disorder diagnosis<br>• No lifetime treatment for depression or anxiety<br>• Does not meet criteria for the CAARMS psychosis risk syndrome<br>• No first-degree family member with schizophrenia | None |
| | | | • Met CHR inclusion criteria via SIPS | • No lifetime psychiatric | |

| | | | | | |
|---|---|---|---|---|---|
| NORMENT, University of Oslo and Oslo University Hospital, Norway | Oslo Region | • 15-30 years old<br>• No organic cause for presentation<br>• Fluent in Norwegian<br>• IQ >70 | • Some participants also met basic symptoms cognitive high-risk criteria via SPI-A (81)<br>• No antipsychotic medication equivalent to a dose of ≥5 mg Olanzapin per day (current or ≥4 weeks lifetime)<br>• Clear evidence that CHR symptoms are not substance-induced | disorder diagnosis<br>• No lifetime psychotropic medication use<br>• No lifetime psychotic disorder diagnosis<br>• No first-degree relative with psychotic disorder diagnosis<br>• No lifetime substance abuse disorder diagnosis | None |
| University of Pittsburgh, USA | Pitt | • 12-40 years old<br>• No lifetime neurological disorder diagnosis<br>• Fluent in English<br>• IQ >80 | • Met CHR inclusion criteria via SIPS<br>• Clear evidence that the psychosis-risk syndrome is NOT due to a (non-schizophrenia-spectrum) psychiatric disorder | • No lifetime DSM-IV disorder diagnosis<br>• No lifetime psychotropic medication exposure<br>• No lifetime psychosis risk syndrome (SIPS criteria)<br>• No first-degree relative with a psychotic disorder diagnosis | None |
| Rush University Medical Center, Chicago, IL USA | RUMC | • 12-35 years old<br>• No lifetime major medical illness or neurological disorder diagnosis that could confound behavioral or neural measurements, incl. diabetes, epilepsy, multiple sclerosis, or traumatic brain injury with severity 7+ on TBI scale<br>• IQ >70 | • Met CHR inclusion criteria via SIPS<br>• Clear evidence that the psychosis-risk syndrome is NOT due to a (non-schizophrenia-spectrum) psychiatric disorder | • No current Axis I/II disorders<br>• Positive symptoms rated 1 or lower on the SIPS | 47 |
| Institute of Mental Health, Singapore and National University of Singapore, Singapore | Singapore | • 14-29 years old<br>• No lifetime neurological disorder diagnosis<br>• Fluent in English<br>• No history of intellectual disability<br>• No lifetime history of illicit substance use | • Met CHR inclusion Criteria via CAARMS<br>• No current antipsychotic or mood stabilizer use<br>• No medical cause associated with psychotic symptoms | • No lifetime psychiatric disorder diagnosis<br>• No first-degree relative with a psychotic disorder diagnosis | 48-50 |
| Seoul National University, South Korea | SNUH | • 15-34 years old<br>• Female or male<br>• No lifetime neurological disorder diagnosis or traumatic brain injury<br>• Fluent in Korean<br>• IQ >70<br>• No MRI contraindications | • Met CHR inclusion criteria via SIPS | • No lifetime psychiatric disorder diagnosis, inclusing psychotic disorder diagnosis<br>• No current psychotropic medication prescription<br>• No first- or second-degree relative with a psychotic disorder diagnosis<br>• No lifetime neurological disorder diagnosis | 51 |
| Stavanger University Hospital, Norway | Stavanger | • 13-65 years old<br>• Female, male<br>• No known neurological or endocrine disorders related to CHR symptoms<br>• Able to speak Scandinavian language<br>• IQ >70<br>• No MRI contraindications | • Met CHR inclusion criteria via CAARMS<br>• Symptoms not better accounted for by major Axis I, Axis II ,or substance use disorder<br>• No current antipsychotic medication use<br>• Not more than 4 weeks lifetime antipsychotic medication use | • No lifetime psychiatric disorder diagnosis<br>• No lifetime psychotropic medication exposure<br>• No lifetime psychosis risk syndrome (CAARMS criteria)<br>• No first-degree relatives with a psychotic disorder diagnosis<br>• No current substance use disorder | None |
| Department of Neuropsychiatry, Toho University School of Medicine, Japan | Toho | • 16-35 years old<br>• No lifetime neurological disorder diagnosis<br>• Fluent in Japanese<br>• IQ >70<br>• No MRI contraindications | • Met CHR inclusion criteria via SIPS | • No lifetime psychiatric disorder diagnosis<br>• No psychotropic medication exposure | 52-54 |
| Department of Neuropsychiatry, | | • 15-30 years old<br>• No neurological disorder diagnosis or traumatic brain injury with known cognitive consequences and/or loss of consciousness >5 mins | • Met CHR inclusion criteria via | • No psychiatric condition (SCID-NP criteria)<br>• No psychotropic medication | |

| | | | | | |
|---|---|---|---|---|---|
| Graduate School of Medicine, The University of Tokyo | Tokyo | • Fluent in Japanese<br>• IQ ≥70<br>• No substance use disorder or substance abuse<br>• No history of electroconvulsive therapy | SIPS | use<br>• No first-degree relatives with a psychotic disorder diagnosis | 56-64 |
| Centre for Addiction and Mental Health, University of Toronto, Canada | Toronto | • 18-40 years old<br>• No current neurological disorder diagnosis<br>• Fluent in English or French<br>• IQ >70 (via clinical impression)<br>• Not currently pregnant<br>• No current drug or alcohol abuse<br>• No lifetime history of severe head trauma | • Met CHR inclusion criteria via SIPS | • No lifetime psychiatric disorder diagnosis<br>• No psychotropic medication prescription<br>• No relatives with a psychiatric disorder diagnosis | 65, 66 |
| University of Toyama Graduate School of Medicine and Pharmaceutical Sciences, Japan | Toyama | • 12-42 years old<br>• No lifetime neurological disorder diagnosis<br>• Fluent in Japanese<br>• No MRI contraindications | • Met CHR inclusion criteria via CAARMS | • No lifetime psychiatric disorder diagnosis<br>• No lifetime psychotropic medication exposure<br>• No first-degree relative with a psychiatric disorder diagnosis<br>• Physically healthy at time of MRI scanning<br>• No history of severe obstetric complications, serious head trauma, serious medical disease (e.g. neurological illness, thyroid dysfunction, diabetes, and hypertension), steroid use, or substance abuse. | 55, 57, 58, 67-75 |
| University of California, San Francisco, USA | UCSF | • 11-30 years old<br>• No lifetime neurological disorder diagnosis<br>• Fluent in English<br>• No MRI contraindications<br>• No substance dependence (past 12 monhts, excluding nicotine)<br>• No head injury<br>• Good physical health | • Met CHR inclusion criteria via SIPS | • No lifetime DSM-IV Axis I disorder diagnosis<br>• No first-degree relatives with a psychotic disorder diagnosis | 76 |
| Psychiatric Hospital, University of Zurich, Switzerland | Zurich | • 13-36 years old<br>• No neurological disorder diagnosis, no organic brain abnormalities (confirmed by neuroradiologist blinded to group allocation)<br>• Fluent in German<br>• IQ ≥80<br>• No pregnancy<br>• No drug or alcohol dependence | • Met CHR inclusion criteria via SIPS<br>• No other psychiatric disorder diagnosis | • No lifetime psychiatric diagnosis via DSM-IV<br>• No psychotropic medication use<br>• No first-degree relatives with a psychotic disorder diagnosis | 77-80 |

*See bibliography for citations*

**eTable 2.** Scanner and acquisition parameters by site

| Site | Scanner Manufacturer | Scanner Model | Tesla Strength | Repetition Time (ms) | Echo Time (ms) | Flip Angle | Voxel Size (mm) | Freesurfer version | N HC | N CHR |
|---|---|---|---|---|---|---|---|---|---|---|
| Amsterdam | Phillips | Image MR Series | 3 | 8280 | 3.8 | • | • | 6.0.0 | 23 | 16 |
| Barcelona-HSJD | GE | Signa HD | 1.5 | 9.5 | 3.9 | 8° | 0.94x0.94x1.2 | 6.0.0 | 6 | 22 |
| | GE | SIGNA HDxt OPTIMA EDITION | 1.5 | 9.5 | 3.9 | 8° | 0.94x0.94x1.2 | 6.0.0 | 38 | 33 |
| Bern | Siemens | Magnetom Verio | 3 | 7920 | 2.48 | 16° | 1.0x1.0x1.0 | 6.0.0 | 17 | 43 |
| Columbia1 | GE | Discovery MR750 | 3 | 7840 | 3.1 | 12° | 0.8x0.8x0.8 | 6.0.0 | 9 | 17 |
| Columbia2 | GE | Discovery MR750 | 3 | 7900 | 3.1 | 12° | 0.8x0.8x0.8 | 6.0.0 | 15 | 19 |
| Columbia3 | GE | Signa | 3 | • | • | 11° | 0.9766 x 0.9766 x 1.0000 | 5.3 | 37 | 58 |
| Copenhagen | Philips | Achieva | 3 | 10.028 | 4.6 | 8° | 0.75x0.75x0.80 | 6.0.0 | • | • |
| | Philips | Achieva | 3 | 10.01 | 4.6 | 8° | 0.75x0.75x0.80 | 6.0.0 | • | 52 |
| CSU | Siemens | Skyra | 3 | 2530 | 2.33 | 7° | 1.0x1.0x1.0 | 6.0.0 | 59 | 52 |
| Glasgow | Siemens | Trio | 3 | 2250 | 2.6 | 9° | 1x1x1 | 6.0.0 | 46 | 80 |
| Heidelberg | Siemens | PET/MR | 3 | 2300 | 2.98 | 9° | 1x1x1 | 6.0.0 | 24 | 8 |
| | Siemens | Tim Trio | 3 | 2300 | 2.98 | 9° | 1x1x1 | 6.0.0 | 9 | 14 |
| IDIBAPS | Siemens | Tim Trio | 3 | 2300 | 3.01 | 9° | 0.94x0.94x1 | 6.0.0 | 39 | 51 |
| | Siemens | Prisma fit | 3 | 2300 | 3.01 | 9° | 0.94x0.94x1 | 6.0.0 | 15 | 15 |
| | Siemens | Prisma fit | 3 | 2300 | 2.98 | 9° | 1x1x1.2 | 6.0.0 | • | 8 |
| ISMMS | Siemens | Skyra | 3 | 2400 | 2.07 | 8° | 0.8x0.8x0.8 | 6.0.0 | 12 | 25 |
| London | GE | Electric Discovery MR750 | 3 | 7.31 | 3.016 | 11° | 1.1 x 1.1. x 1.2 | 6.0.0 | 29 | 81 |
| | GE | Signa HDx | 3 | 7.144 | 2.848 ms | 20° | 1.1 x 1.1. x 1.1 | 6.0.0 | 29 | 81 |
| Maastricht | Philips | Intera | 3 | 2250 | 4.6 | 8° | 1.17×1.17×1.20 | 6.0.0 | 33 | 48 |
| | Philips | Achieva | 3 | 2250 | 4.6 | 8° | 1.2x.8x.8 | 6.0.0 | 5 | • |
| Melbourne | Siemens | Trio | 3 | 1900 | 2.15 | 90° | 0.5x0.5x1.0 | 6.0.0 | 11 | 18 |
| | Siemens | Trio | 3 | 2300 | 2.98 | 9° | 1.0x1.0x1.2 | 6.0.0 | 20 | • |
| | GE | Signa | 1.5 | 14.3 | 3.3 | 30° | 0.938x0.938x1.5 | 6.0.0 | 61 | 54 |
| | GE | LX Horizon | 3 | 36 | 9 | 30° | 0.4883x0.4883x2 | 6.0.0 | • | 90 |
| | GE | LX Horizon | 3 | 36 | 9 | 30° | 0.4883x0.4883x2 | 6.0.0 | • | 83 |
| | GE | LX Horizon | 3 | 36 | 9 | 30° | 0.4883x0.4883x2 | 6.0.0 | • | 4 |
| Mexico City | GE | Signa_HDx | 3 | 1340 | 5.7 | 20° | 1.2x1.2x1.2 | 6.0.0 | 38 | 33 |
| MHRC | Philips | Achieva | 3 | 8.2 | 3.7 | 8° | 0.83×0.83x1.0 | 6.0.0 | 51 | 38 |
| MPRC | Siemens | Trio | 3 | 2400 | 2.2 | 8° | 0.8x0.8x0.8 | 6.0.0 | 19 | 11 |
| | Siemens | Prisma | 3 | 2400 | 2.2 | 8° | 0.8x0.8x0.8 | 6.0.0 | 1 | 20 |
| Newcastle | Siemens | Avanto | 1.5 | 1980 | 4.3 | 15° | .9765625 x 0.9765625 x 1 | 6.0.0 | 13 | 45 |
| | Siemens | Avanto | 1.5 | 1980 | 4.3 | 15° | .9765625 x 0.9765625 x 1 | 6.0.0 | 4 | • |
| Oslo Region | GE | Signa HDxt | 3 | 7800 | 2956 | 12° | 1.0×1.0×1.2 | 5.3 | 39 | 13 |
| | GE | Discovery MR750 | 3 | 8,16 | 3,18 | 12° | 1.0×1.0×1.0 | 5.3 | 24 | 8 |
| Pitt | Siemens | Prisma | 3 | 2400 | 2.2 | 8° | 0.8x0.8x0.8 | 6.0.0 | 65 | 26 |
| RUMC | Siemens | Verio | 3 | 2530 | 2.27 | 7° | 1.0x1.0x1.0 | 6.0.0 | • | 7 |
| | Siemens | Verio | 3 | 2530 | 2.27 | 7° | 1.0x1.0x1.0 | 6.0.0 | • | 18 |
| | Siemens | Verio | 3 | 2530 | 2.27 | 7° | 1.0x1.0x1.0 | 6.0.0 | 19 | 26 |
| | Siemens | Verio | 3 | 2530 | 2.27 | 7° | 1.0x1.0x1.0 | 6.0.0 | 10 | 16 |
| Singapore | Siemens | Tim Trio | 3 | 2300 | 3 | 9° | 1.0x1.0x1.0 | 6.0.0 | 53 | 100 |
| SNUH | Siemens | Magnetom TrioTim | 3 | 1670 | 1.89 | 9° | 1 x 0.98 x 0.98 | 6.0.0 | 74 | 74 |
| Stavanger | GE Medical | 450 DISCOVERY | 1,5 | 7,9 | 3,1 | 12° | • | 5.3 | 33 | 37 |

| | | | | | | | | | | |
|---|---|---|---|---|---|---|---|---|---|---|
| | systems | | | | | | | | | |
| Toho | Toshiba Medical Systems | EXCELART Vantage | 1.5 | • | • | 35° | 0.98 × 0.98 × 1.0 mm | 5.2 | 16 | 40 |
| Tokyo | GE | SIGNA HDx | 3 | 6.8 | 1.94 | 20° | 1.0x1.0x1.0 | 6.0.0 | 21 | 29 |
| | GE | DISCOVERY MR750W | 3 | 8.46 | 3.25 | 20° | 1.0x1.0x1.0 | 6.0.0 | 4 | 10 |
| Toronto | GE | Discovery MR750 | 3 | 6736 | 2.99 | 8° | 0.9x0.9x0.9 | 6.0.0 | 39 | 27 |
| Toyama | Siemens | Magnetom Vision | 1.5 | 2400 | 5 | 40° | 1.0x1.0x1.0 | 6.0.0 | 52 | 22 |
| | Siemens | Magnetom Verio | 3 | 2300 | 2.9 | 9° | 1.0x1.0x1.2 | 6.0.0 | 89 | 57 |
| UCSF | Siemens | Magnetom TrioTim | 3 | 2300 | 2.95 | 9° | 1.0x1.0x1.2 | 5.1 | 103 | 71 |
| Zurich | Phillips | Philips Achieva TX | 3 | 8.3 | 3.8 | 8° | 1.0x1.0x1.0 | 6.0.0 | 5 | 26 |
| | Phillips | Philips Achieva TX | 3 | 8.3 | 3.8 | 8° | 1.0x1.0x1.0 | 6.0.0 | 38 | 36 |

*Unavailable information left blank*

**eTable 3.** Age and sex of HC vs. CHR vs. CHR-PS+ vs. CHR-PS- in 12-25 year old age range (Age-associated analyses)

| Metric | HC | CHR | CHR-PS+ | CHR-PS- |
|---|---|---|---|---|
| N | 1009 | 1472 | 217 | 1014 |
| Age Mean (SD) | 19.8 (3.1) | 18.9 (2.9) | 18.5 (3) | 18.9 (2.9) |
| Age Range | 12.6-25 | 12.5-24.9 | 12.6-24.6 | 12.5-24.9 |
| % Female | 45 | 47 | 41 | 49 |

*Because no follow-up data is available for some participants, CHR-PS+ N and CHR-PS- N will not sum to the total CHR N*

**eTable 4.** Intelligence quotient comparisons of HC vs. CHR by site

| Site | IQ Method | HC | | CHR | | F | p |
|---|---|---|---|---|---|---|---|
| | | N | Mean IQ (SD) | N | Mean IQ (SD) | | |
| Barcelona-HSJD | WAIS-III | 11 | 111.8 (7) | 9 | 92.3 (13.3) | 17.77 | 5.20e-04 |
| | WASI-II | 31 | 110.7 (13.8) | 37 | 96.3 (12.9) | 19.59 | 3.69e-05 |
| Copenhagen | WAIS-III | 59 | 113 (13.6) | 163 | 103 (13.1) | 24.31 | 1.63e-06 |
| CSU | WISC-I+PC | 59 | 145.8 (19.7) | 52 | 125.6 (26) | 21.58 | 9.55e-06 |
| Glasgow | NART | 46 | 115.1 (6.1) | 80 | 114.2 (5.9) | 0.55 | 0.46 |
| Heidelberg | WISC-IV | 33 | 117.8 (9.7) | 22 | 103.8 (11.9) | 22.87 | 1.42e-05 |
| IDIBAPS | WAIS-III | 16 | 104.2 (9.8) | 13 | 106.4 (12.4) | 0.27 | 0.61 |
| | WAIS-IV | 1 | 119 (NA) | 8 | 100.2 (18.6) | 0.9 | 0.37 |
| | WISC-IV | 24 | 106.7 (11.9) | 42 | 96.7 (14.5) | 8.17 | 5.75e-03 |
| | WISC-V | 1 | 116 (NA) | 3 | 86.7 (6.8) | 13.93 | 0.06 |
| Maastricht | DART | 38 | 107.1 (15.8) | 48 | 102.6 (9.9) | 2.31 | 0.13 |
| Melbourne | NART | 61 | 119.7 (32.3) | 0 | • | • | • |
| | WASI | 0 | • | 56 | 100.3 (14) | • | • |
| | WASI-II | 31 | 117 (10.5) | 18 | 105.9 (11.3) | 11.9 | 1.20e-03 |
| MPRC | WASI | 1 | 96 (NA) | 3 | 108.7 (20.7) | 0.28 | 0.65 |
| | WASI-II | 0 | • | 10 | 110.9 (9.9) | • | • |
| Newcastle | WASI-II | 17 | 120.2 (10.1) | 44 | 107.9 (15.3) | 9.3 | 3.43e-03 |
| Oslo Region | WASI | 48 | 111 (10.4) | 20 | 105.2 (12.6) | 3.84 | 0.05 |
| Pitt | WASI | 65 | 109.9 (8.4) | 26 | 101.6 (10.6) | 15.35 | 1.78e-04 |
| RUMC | WASI | 10 | 113.8 (8.7) | 16 | 108.5 (13.7) | 0.97 | 0.34 |
| | WRAT | 19 | 105.9 (13.6) | 51 | 110.3 (17.9) | 0.94 | 0.34 |
| SNUH | K-WAIS | 74 | 110.2 (12) | 74 | 108.9 (11.6) | 0.47 | 0.49 |
| Stavanger | WAIS-IV | 33 | 99.5 (7.6) | 37 | 96.7 (11.8) | 1.34 | 0.25 |
| Toho | WAIS-III | 0 | • | 10 | 93.8 (17.8) | • | • |
| Tokyo | JART25 | 25 | 106.4 (10.3) | 39 | 105.8 (9.7) | 0.06 | 0.81 |
| Toronto | RBANS | 39 | 88.8 (14.2) | 27 | 90.1 (12.2) | 0.15 | 0.70 |
| Toyama | JART50 | 85 | 110.1 (6.8) | 72 | 97.1 (9.1) | 104.28 | 4.82e-19 |
| UCSF | WASI-II | 43 | 114.5 (11.8) | 71 | 110.6 (14.7) | 1.72 | 0.19 |
| Zurich | MWT-B | 23 | 108.2 (13.3) | 25 | 100.2 (13.1) | 4.31 | 0.04 |
| | WISC | 13 | 114.8 (18.7) | 33 | 106 (14.3) | 2.95 | 0.09 |

*These sites did not have intelligence quotient data available and were omitted from the table: Amsterdam, Columbia, Bern, ISMMS, London, Mexico City, MHRC, and Singapore. Table reports results from two-way ANOVA group comparisons. F and p marked as "•" if intelligence quotient data were not available for at least one group.*

**eTable 5.** Intelligence quotient comparisons of CHR-PS+ vs. CHR-PS- vs. CHR-UNK by site

| Site | IQ Method | CHR-PS+ | | CHR-PS- | | CHR-UNK | | F | p |
|---|---|---|---|---|---|---|---|---|---|
| | | N | Mean IQ (SD) | N | Mean IQ (SD) | N | Mean IQ (SD) | | |
| Barcelona-HSJD | WAIS-III | 2 | 77.5 (9.2) | 7 | 96.6 (11.3) | 0 | • | 4.68 | 0.07 |
| | WASI-II | 4 | 102.5 (14.7) | 24 | 92.5 (12.3) | 9 | 103.9 (10.5) | 3.49 | 0.04 |
| Copenhagen | WAIS-III | 13 | 101 (17.2) | 95 | 103.7 (12.7) | 55 | 102.3 (12.8) | 0.37 | 0.69 |
| CSU | WISC-I+PC | 21 | 120.3 (26.7) | 31 | 129.2 (25.3) | 0 | • | 1.48 | 0.23 |
| Glasgow | NART | 6 | 114 (6.1) | 74 | 114.3 (5.9) | 0 | • | 0.01 | 0.92 |
| Heidelberg | WISC-IV | 0 | • | 18 | 99.2 (11.2) | 4 | 99.2 (11.2] | 0.71 | 0.41 |
| IDIBAPS | WAIS-III | 3 | 109.3 (4.2) | 10 | 105.5 (14) | 0 | • | 0.21 | 0.66 |
| | WAIS-IV | 1 | 116 (NA) | 5 | 96.8 (19.1) | 2 | 101 (25.5) | 0.37 | 0.71 |
| | WISC-IV | 10 | 103.4 (13.4) | 23 | 93.7 (12.3) | 9 | 97.2 (19.4) | 1.63 | 0.21 |
| | WISC-V | 1 | 79 (NA) | 1 | 89 (NA) | 1 | 92 (NA) | • | • |
| Maastricht | DART | 6 | 105.8 (6.8) | 25 | 100 (9.2) | 17 | 105.3 (11.1) | 1.88 | 0.16 |
| Melbourne | WASI | 8 | 95.1 (16.4) | 48 | 101.1 (13.5) | 0 | • | 1.28 | 0.26 |
| | WASI-II | 1 | 125 (NA) | 6 | 101.5 (6.8) | 11 | 106.5 (12.1) | 2.14 | 0.15 |
| MPRC | WASI | 1 | 131 (NA) | 1 | 90 (NA) | 1 | 105 (NA) | • | • |
| | WASI-II | 0 | • | 3 | 111.2 (12.2) | 7 | 111.2 (12.2] | 0.01 | 0.91 |
| Newcastle | WASI-II | 1 | 75 (NA) | 40 | 108.8 (14.9) | 3 | 107.3 (10.8) | 2.57 | 0.09 |
| Oslo Region | WASI | 2 | 118.5 (10.6) | 18 | 103.7 (12.1) | 0 | • | 2.7 | 0.12 |
| Pitt | WASI | 2 | 104 (5.7) | 11 | 103.1 (7.8) | 13 | 99.9 (13.3) | 0.3 | 0.74 |
| RUMC | WASI | 1 | 90 (NA) | 12 | 114.2 (10.3) | 3 | 91.7 (4.5) | 8.47 | 4.41e-03 |
| | WRAT | 3 | 108 (10.4) | 48 | 110.5 (18.3) | 0 | • | 0.05 | 0.82 |
| SNUH | K-WAIS | 9 | 108.3 (9.1) | 46 | 109.6 (12) | 19 | 107.4 (12) | 0.26 | 0.77 |
| Stavanger | WAIS-IV | 5 | 98 (13) | 31 | 96.4 (11.9) | 1 | • | 0.07 | 0.79 |
| Toho | WAIS-III | 1 | 81 (NA) | 9 | 95.2 (18.2) | 0 | • | 0.55 | 0.48 |
| Tokyo | JART25 | 3 | 108.3 (10) | 25 | 106.1 (10) | 11 | 104.4 (9.9) | 0.22 | 0.80 |
| Toronto | RBANS | 0 | • | 4 | 89.2 (12.6) | 23 | 89.2 (12.6] | 0.83 | 0.37 |
| Toyama | JART50 | 9 | 96.8 (9.9) | 63 | 97.2 (9.1) | 0 | • | 0.01 | 0.90 |
| UCSF | WASI-II | 13 | 113.2 (14.4) | 47 | 110.2 (15.1) | 11 | 108.5 (13.5) | 0.27 | 0.76 |
| Zurich | MWT-B | 5 | 86.6 (7.7) | 11 | 108.2 (13.3) | 9 | 98.1 (7.8) | 7.39 | 3.52e-03 |
| | WISC | 5 | 90.8 (7.5) | 23 | 108.7 (13.7) | 5 | 108.8 (13.8) | 3.97 | 0.03 |

*These sites did not have intelligence quotient data available and were omitted from the table: Amsterdam, Columbia, Bern, ISMMS, London, Mexico City, MHRC, and Singapore. Table reports results from three-way ANOVA group comparisons. F and p marked as "•" if intelligence quotient data were not available for at least one group.*

**eTable 6.** Symptom differences between CHR groups (CHR-PS+, CHR-PS-, CHR-UNK) as measured by the SIPS or CAARMS

| Test | Subtest | CHR-PS+ | | CHR-PS- | | CHR-UNK | | F | p | Pairwise Comparisons |
|---|---|---|---|---|---|---|---|---|---|---|
| | | N | Mean Score (SD) | N | Mean Score (SD) | N | Mean Score (SD) | | | |
| A. SIPS | Positive Symptoms | 133 | 12.1 (5.3) | 599 | 10.6 (4.7) | 207 | 11 (4.2) | 7.67 | 4.97e-04 | CHR-PS+ > CHR-PS-; CHR-PS+ > CHR-UNK |
| | Negative Symptoms | 130 | 14.1 (6.8) | 561 | 12.5 (6.6) | 193 | 12.1 (6.1) | 3.26 | 0.04 | CHR-PS+ > CHR-PS-; CHR-PS+ > CHR-UNK |
| | Disorganized Symptoms | 129 | 6.3 (4.1) | 561 | 5.7 (3.6) | 193 | 4.8 (3.2) | 7.81 | 4.35e-04 | CHR-PS+ > CHR-UNK; CHR-PS- > CHR-UNK |
| | General Symptoms | 129 | 8.5 (4.4) | 564 | 8.1 (4.4) | 193 | 8.2 (4.4) | 1.40 | 0.25 | • |
| B. CAARMS | Positive Symptoms | 48 | 10.7 (4) | 457 | 10 (4.1) | 84 | 11.4 (3.5) | 0.49 | 0.61 | • |
| | Negative Symptoms | 12 | 7.2 (4.6) | 135 | 6.5 (3.6) | 24 | 6.6 (3.8) | 0.04 | 0.96 | • |
| | Cognitive Changes | 10 | 4 (1.8) | 92 | 3.5 (1.9) | 21 | 2.2 (1.8) | 3.46 | 0.04 | CHR-PS+ > CHR-UNK; CHR-PS- > CHR-UNK |
| | Emotional Disturbances | 10 | 4.1 (2) | 92 | 4 (2.5) | 21 | 3 (2.5) | 0.63 | 0.53 | • |
| | Behavioral Changes | 11 | 9.4 (5.5) | 136 | 8.8 (4.2) | 24 | 7.2 (4.3) | 0.92 | 0.40 | • |
| | Motor/Physical Changes | 9 | 2.9 (2.7) | 86 | 2.9 (2.3) | 21 | 2 (1.9) | 4.47 | 0.01 | No pairwise comparisons were significant |
| | General Psychopathology | 8 | 13.1 (7.6) | 131 | 13.2 (6.4) | 24 | 11.7 (6.2) | 1.15 | 0.32 | • |

*Table reports results of linear mixed model analyses examining effect of group (HC vs. CHR-PS+ vs. CHR-PS-) on each respective symptom measure. Site was included in the model as a random effect. Any statistically significant effects of group (p<.05) in the overall model were followed-up with pairwise comparisons. "•" indicates that the pairwise comparisons were not conducted, due to lack of statistical significance in overall model. Significant [p<0.05] pairwise results reported.*

**eTable 7.** CHR current medication information reported by site

| Site | N CHR | Typical Antipsychotics | | Atypical Antipsychotics | | Lithium | | Anticonvulsants | | Stimulants | | Other Psychotropic Medications | |
|---|---|---|---|---|---|---|---|---|---|---|---|---|---|
| | | N | % | N | % | N | % | N | % | N | % | N | % |
| Amsterdam | 16 | 0 | 0% | 0 | 0% | 0 | 0% | 1 | 6% | 1 | 6% | 9 | 56% |
| Barcelona-HSJD | 54 | 0 | 0% | 20 | 37% | 0 | 0% | 0 | 0% | 5 | 9% | 27 | 50% |
| Bern | 29 | 1 | 3% | 3 | 10% | 1 | 3% | 1 | 3% | 1 | 3% | 4 | 14% |
| Columbia1 | 17 | 0 | 0% | 7 | 41% | 1 | 6% | 3 | 18% | 0 | 0% | 0 | 0% |
| Columbia3 | 57 | 0 | 0% | 12 | 21% | 0 | 0% | 3 | 5% | 6 | 11% | 20 | 35% |
| Copenhagen | 158 | 0 | 0% | 42 | 27% | 0 | 0% | 7 | 4% | 0 | 0% | 53 | 34% |
| CSU | 52 | 0 | 0% | 0 | 0% | 0 | 0% | 0 | 0% | 0 | 0% | 0 | 0% |
| Glasgow | 80 | 1 | 1% | 2 | 2% | 1 | 1% | 1 | 1% | 0 | 0% | 0 | 0% |
| Heidelberg | 22 | 0 | 0% | 0 | 0% | 0 | 0% | 0 | 0% | 0 | 0% | 0 | 0% |
| IDIBAPS | 74 | 1 | 1% | 33 | 45% | 1 | 1% | 5 | 7% | 7 | 9% | 32 | 43% |
| ISMMS | 25 | 0 | 0% | 6 | 24% | 0 | 0% | 2 | 8% | 1 | 4% | 0 | 0% |
| London | 81 | 0 | 0% | 8 | 10% | 0 | 0% | 0 | 0% | 0 | 0% | 0 | 0% |
| Maastricht | 31 | 9 | 29% | 0 | 0% | 1 | 3% | 0 | 0% | 4 | 13% | 11 | 35% |
| Melbourne | 18 | 0 | 0% | 0 | 0% | 0 | 0% | 0 | 0% | 0 | 0% | 12 | 67% |
| Mexico City | 33 | 0 | 0% | 0 | 0% | 0 | 0% | 0 | 0% | 0 | 0% | 0 | 0% |
| MHRC | 36 | 18 | 50% | 29 | 81% | 0 | 0% | 0 | 0% | 0 | 0% | 0 | 0% |
| MPRC | 31 | 0 | 0% | 6 | 19% | 1 | 3% | 1 | 3% | 9 | 29% | 9 | 29% |
| Newcastle | 41 | 0 | 0% | 1 | 2% | 0 | 0% | 0 | 0% | 1 | 2% | 0 | 0% |
| Oslo Region | 21 | 0 | 0% | 4 | 19% | 0 | 0% | 0 | 0% | 0 | 0% | 5 | 24% |
| Pitt | 25 | 0 | 0% | 9 | 36% | 1 | 4% | 0 | 0% | 3 | 12% | 16 | 64% |
| RUMC | 64 | 0 | 0% | 1 | 2% | 0 | 0% | 1 | 2% | 1 | 2% | 7 | 11% |
| Singapore | 100 | 0 | 0% | 1 | 1% | 0 | 0% | 0 | 0% | 0 | 0% | 16 | 16% |
| SNUH | 74 | 0 | 0% | 14 | 19% | 0 | 0% | 0 | 0% | 0 | 0% | 24 | 32% |
| Stavanger | 37 | 0 | 0% | 2 | 5% | 0 | 0% | 0 | 0% | 0 | 0% | 0 | 0% |
| Tokyo | 39 | 3 | 8% | 18 | 46% | 2 | 5% | 0 | 0% | 0 | 0% | 20 | 51% |
| Toronto | 26 | 0 | 0% | 6 | 23% | 0 | 0% | 0 | 0% | 4 | 15% | 10 | 38% |
| Toyama | 79 | 1 | 1% | 14 | 18% | 0 | 0% | 2 | 3% | 0 | 0% | 26 | 33% |
| UCSF | 68 | 0 | 0% | 13 | 19% | 3 | 4% | 1 | 1% | 3 | 4% | 22 | 32% |
| Zurich | 62 | 0 | 0% | 0 | 0% | 1 | 2% | 1 | 2% | 4 | 6% | 2 | 3% |

*These sites did not have medication data available and were omitted from the table: Columbia2 and Toho.*

**eTable 8.** CHR vs. HC effect size overview (post-ComBat mega analysis)

| | | Mega Analysis (post-ComBat) | | | |
|---|---|---|---|---|---|
| **Region of Interest** | **Hemisphere** | **Cohen's d** | **95% Confidence Interval** | **p** | **q** |
| **Cortical Thickness** | | | | | |
| Banks of the Superior Temporal Sulcus | L | **-0.09** | **(-0.16 - -0.02)** | **0.01** | **0.03** |
| | R | **-0.13** | **(-0.2 - -0.06)** | **2.33e-04** | **2.17e-03** |
| Caudal Anterior Cingulate | L | -0.08 | (-0.15 - -0.01) | 0.02 | 0.05 |
| | R | -0.07 | (-0.14 - 0) | 0.06 | 0.13 |
| Caudal Middle Frontal | L | **-0.12** | **(-0.19 - -0.04)** | **1.45e-03** | **7.50e-03** |
| | R | **-0.12** | **(-0.2 - -0.05)** | **6.13e-04** | **4.13e-03** |
| Cuneus | L | 0.01 | (-0.06 - 0.08) | 0.83 | 0.89 |
| | R | -0.05 | (-0.12 - 0.02) | 0.18 | 0.30 |
| Entorhinal | L | -0.02 | (-0.09 - 0.05) | 0.51 | 0.65 |
| | R | -0.06 | (-0.13 - 0.01) | 0.10 | 0.19 |
| Frontal Pole | L | -0.05 | (-0.12 - 0.02) | 0.20 | 0.32 |
| | R | -0.08 | (-0.15 - -0.01) | 0.03 | 0.07 |
| Fusiform | L | **-0.17** | **(-0.24 - -0.1)** | **1.55e-06** | **7.07e-05** |
| | R | **-0.11** | **(-0.18 - -0.04)** | **2.33e-03** | **0.01** |
| Inferior Parietal | L | **-0.16** | **(-0.24 - -0.09)** | **5.31e-06** | **1.65e-04** |
| | R | **-0.11** | **(-0.18 - -0.04)** | **2.10e-03** | **0.01** |
| Inferior Temporal | L | **-0.13** | **(-0.2 - -0.06)** | **4.00e-04** | **2.95e-03** |
| | R | **-0.15** | **(-0.22 - -0.08)** | **4.36e-05** | **5.63e-04** |
| Insula | L | **-0.17** | **(-0.24 - -0.1)** | **1.83e-06** | **7.07e-05** |
| | R | **-0.16** | **(-0.23 - -0.08)** | **1.66e-05** | **3.45e-04** |
| Isthmus Cingulate | L | -0.03 | (-0.1 - 0.04) | 0.41 | 0.56 |
| | R | -0.06 | (-0.13 - 0.01) | 0.08 | 0.16 |
| Lateral Occipital | L | **-0.12** | **(-0.19 - -0.05)** | **1.22e-03** | **6.99e-03** |
| | R | **-0.16** | **(-0.23 - -0.09)** | **1.04e-05** | **2.70e-04** |
| Lateral Orbitofrontal | L | -0.08 | (-0.15 - -0.01) | 0.03 | 0.07 |
| | R | **-0.09** | **(-0.16 - -0.02)** | **0.02** | **0.05** |
| Lingual | L | -0.04 | (-0.11 - 0.04) | 0.33 | 0.46 |
| | R | -0.01 | (-0.08 - 0.07) | 0.88 | 0.91 |
| Medial Orbitofrontal | L | **-0.13** | **(-0.2 - -0.06)** | **2.33e-04** | **2.17e-03** |
| | R | **-0.11** | **(-0.18 - -0.03)** | **3.56e-03** | **0.01** |
| Middle Temporal | L | **-0.10** | **(-0.17 - -0.03)** | **7.30e-03** | **0.02** |
| | R | **-0.18** | **(-0.26 - -0.11)** | **3.81e-07** | **5.46e-05** |
| Paracentral | L | **-0.15** | **(-0.22 - -0.08)** | **3.24e-05** | **5.58e-04** |
| | R | **-0.11** | **(-0.18 - -0.04)** | **2.97e-03** | **0.01** |
| Parahippocampal | L | **-0.13** | **(-0.2 - -0.06)** | **4.63e-04** | **3.26e-03** |
| | R | -0.07 | (-0.15 - 0) | 0.04 | 0.09 |
| Pars Opercularis | L | -0.05 | (-0.12 - 0.02) | 0.19 | 0.30 |
| | R | -0.07 | (-0.14 - 0) | 0.04 | 0.10 |
| Pars Orbitalis | L | -0.06 | (-0.13 - 0.02) | 0.12 | 0.23 |
| | R | 0.00 | (-0.07 - 0.07) | 0.94 | 0.97 |
| Pars Triangularis | L | 0.01 | (-0.06 - 0.08) | 0.70 | 0.81 |
| | R | -0.05 | (-0.12 - 0.02) | 0.15 | 0.27 |
| Pericalcarine | L | 0.02 | (-0.05 - 0.09) | 0.54 | 0.68 |
| | R | 0.05 | (-0.02 - 0.12) | 0.18 | 0.30 |
| Postcentral | L | **-0.10** | **(-0.17 - -0.02)** | **8.30e-03** | **0.03** |
| | R | **-0.12** | **(-0.19 - -0.05)** | **1.14e-03** | **6.77e-03** |

| | | | | | |
|---|---|---|---|---|---|
| Posterior Cingulate | L | **-0.11** | **(-0.18 - -0.03)** | **3.48e-03** | **0.01** |
| | R | **-0.13** | **(-0.2 - -0.06)** | **2.38e-04** | **2.17e-03** |
| Precentral | L | **-0.13** | **(-0.2 - -0.06)** | **3.51e-04** | **2.72e-03** |
| | R | **-0.13** | **(-0.2 - -0.06)** | **2.60e-04** | **2.24e-03** |
| Precuneus | L | **-0.11** | **(-0.18 - -0.04)** | **1.68e-03** | **8.39e-03** |
| | R | **-0.15** | **(-0.22 - -0.08)** | **4.02e-05** | **5.63e-04** |
| Rostral Anterior Cingulate | L | **-0.11** | **(-0.18 - -0.04)** | **2.38e-03** | **0.01** |
| | R | -0.08 | (-0.15 - -0.01) | 0.02 | 0.06 |
| Rostral Middle Frontal | L | **-0.13** | **(-0.2 - -0.06)** | **2.91e-04** | **2.37e-03** |
| | R | **-0.12** | **(-0.19 - -0.04)** | **1.43e-03** | **7.50e-03** |
| Superior Frontal | L | **-0.11** | **(-0.18 - -0.04)** | **2.79e-03** | **0.01** |
| | R | **-0.09** | **(-0.16 - -0.02)** | **0.01** | **0.04** |
| Superior Parietal | L | **-0.10** | **(-0.18 - -0.03)** | **3.88e-03** | **0.01** |
| | R | **-0.12** | **(-0.19 - -0.05)** | **8.12e-04** | **5.24e-03** |
| Superior Temporal | L | **-0.12** | **(-0.19 - -0.05)** | **1.05e-03** | **6.48e-03** |
| | R | **-0.15** | **(-0.22 - -0.08)** | **5.31e-05** | **6.33e-04** |
| Supramarginal | L | **-0.11** | **(-0.18 - -0.04)** | **3.27e-03** | **0.01** |
| | R | **-0.10** | **(-0.17 - -0.03)** | **7.48e-03** | **0.02** |
| Temporal Pole | L | -0.06 | (-0.13 - 0.02) | 0.13 | 0.23 |
| | R | -0.08 | (-0.15 - -0.01) | 0.02 | 0.06 |
| Transverse Temporal | L | -0.07 | (-0.14 - 0) | 0.05 | 0.10 |
| | R | **-0.11** | **(-0.18 - -0.04)** | **2.43e-03** | **0.01** |
| **Surface Area** | | | | | |
| Banks of the Superior Temporal Sulcus | L | -0.07 | (-0.14 - 0) | 0.05 | 0.11 |
| | R | -0.07 | (-0.15 - 0) | 0.04 | 0.09 |
| Caudal Anterior Cingulate | L | -0.01 | (-0.08 - 0.06) | 0.73 | 0.81 |
| | R | -0.08 | (-0.15 - -0.01) | 0.04 | 0.08 |
| Caudal Middle Frontal | L | -0.05 | (-0.12 - 0.02) | 0.17 | 0.29 |
| | R | -0.02 | (-0.09 - 0.05) | 0.57 | 0.70 |
| Cuneus | L | -0.05 | (-0.12 - 0.02) | 0.20 | 0.32 |
| | R | -0.08 | (-0.15 - -0.01) | 0.03 | 0.08 |
| Entorhinal | L | **-0.09** | **(-0.16 - -0.02)** | **0.01** | **0.04** |
| | R | -0.02 | (-0.1 - 0.05) | 0.50 | 0.64 |
| Frontal Pole | L | 0.04 | (-0.03 - 0.11) | 0.25 | 0.37 |
| | R | 0.05 | (-0.03 - 0.12) | 0.21 | 0.32 |
| Fusiform | L | -0.05 | (-0.12 - 0.03) | 0.21 | 0.32 |
| | R | -0.07 | (-0.14 - 0) | 0.04 | 0.09 |
| Inferior Parietal | L | -0.06 | (-0.13 - 0.01) | 0.10 | 0.19 |
| | R | **-0.10** | **(-0.17 - -0.03)** | **6.85e-03** | **0.02** |
| Inferior Temporal | L | -0.03 | (-0.1 - 0.04) | 0.48 | 0.62 |
| | R | -0.03 | (-0.1 - 0.04) | 0.45 | 0.59 |
| Insula | L | -0.06 | (-0.13 - 0.01) | 0.11 | 0.21 |
| | R | -0.04 | (-0.11 - 0.03) | 0.30 | 0.43 |
| Isthmus Cingulate | L | 0.00 | (-0.07 - 0.07) | 0.98 | 0.99 |
| | R | **0.09** | **(0.02 - 0.16)** | **9.49e-03** | **0.03** |
| Lateral Occipital | L | 0.00 | (-0.07 - 0.07) | 0.99 | 0.99 |
| | R | -0.01 | (-0.08 - 0.06) | 0.81 | 0.87 |
| Lateral Orbitofrontal | L | -0.02 | (-0.09 - 0.05) | 0.64 | 0.76 |
| | R | -0.02 | (-0.09 - 0.05) | 0.57 | 0.70 |
| Lingual | L | -0.05 | (-0.12 - 0.02) | 0.16 | 0.27 |

| | | | | | |
|---|---|---|---|---|---|
| | R | -0.02 | (-0.09 - 0.05) | 0.64 | 0.76 |
| Medial Orbitofrontal | L | -0.01 | (-0.08 - 0.06) | 0.73 | 0.81 |
| | R | -0.08 | (-0.15 - -0.01) | 0.03 | 0.07 |
| Middle Temporal | L | 0.02 | (-0.06 - 0.09) | 0.67 | 0.78 |
| | R | -0.07 | (-0.14 - 0) | 0.05 | 0.11 |
| Paracentral | L | 0.00 | (-0.07 - 0.07) | 0.95 | 0.97 |
| | R | 0.02 | (-0.05 - 0.09) | 0.59 | 0.72 |
| Parahippocampal | L | -0.04 | (-0.12 - 0.03) | 0.22 | 0.33 |
| | R | 0.06 | (-0.02 - 0.13) | 0.13 | 0.23 |
| Pars Opercularis | L | **-0.09** | **(-0.16 - -0.02)** | **9.85e-03** | **0.03** |
| | R | -0.01 | (-0.08 - 0.06) | 0.81 | 0.87 |
| Pars Orbitalis | L | 0.03 | (-0.04 - 0.1) | 0.47 | 0.62 |
| | R | 0.03 | (-0.04 - 0.1) | 0.44 | 0.59 |
| Pars Triangularis | L | -0.05 | (-0.12 - 0.02) | 0.15 | 0.27 |
| | R | 0.01 | (-0.06 - 0.08) | 0.71 | 0.81 |
| Pericalcarine | L | -0.05 | (-0.12 - 0.02) | 0.20 | 0.32 |
| | R | **-0.09** | **(-0.17 - -0.02)** | **9.29e-03** | **0.03** |
| Postcentral | L | 0.00 | (-0.07 - 0.07) | 0.99 | 0.99 |
| | R | -0.04 | (-0.11 - 0.03) | 0.26 | 0.38 |
| Posterior Cingulate | L | -0.04 | (-0.11 - 0.03) | 0.31 | 0.44 |
| | R | -0.06 | (-0.13 - 0.01) | 0.08 | 0.17 |
| Precentral | L | 0.05 | (-0.02 - 0.12) | 0.14 | 0.26 |
| | R | 0.01 | (-0.06 - 0.08) | 0.72 | 0.81 |
| Precuneus | L | -0.03 | (-0.1 - 0.04) | 0.38 | 0.53 |
| | R | -0.04 | (-0.11 - 0.03) | 0.31 | 0.44 |
| Rostral Anterior Cingulate | L | **-0.12** | **(-0.19 - -0.04)** | **1.43e-03** | **7.50e-03** |
| | R | -0.06 | (-0.13 - 0.01) | 0.09 | 0.17 |
| Rostral Middle Frontal | L | -0.01 | (-0.08 - 0.06) | 0.75 | 0.82 |
| | R | -0.02 | (-0.09 - 0.05) | 0.55 | 0.69 |
| Superior Frontal | L | -0.03 | (-0.1 - 0.05) | 0.48 | 0.62 |
| | R | **-0.09** | **(-0.16 - -0.02)** | **0.01** | **0.03** |
| Superior Parietal | L | 0.01 | (-0.06 - 0.08) | 0.81 | 0.87 |
| | R | 0.01 | (-0.06 - 0.08) | 0.73 | 0.81 |
| Superior Temporal | L | -0.03 | (-0.1 - 0.04) | 0.42 | 0.56 |
| | R | -0.05 | (-0.12 - 0.02) | 0.18 | 0.30 |
| Supramarginal | L | **-0.11** | **(-0.18 - -0.04)** | **2.69e-03** | **0.01** |
| | R | -0.07 | (-0.14 - 0) | 0.06 | 0.12 |
| Temporal Pole | L | -0.03 | (-0.1 - 0.04) | 0.40 | 0.55 |
| | R | 0.05 | (-0.02 - 0.12) | 0.18 | 0.30 |
| Transverse Temporal | L | -0.04 | (-0.11 - 0.03) | 0.23 | 0.35 |
| | R | -0.02 | (-0.09 - 0.06) | 0.67 | 0.78 |
| **Subcortical Volume** | | | | | |
| Amygdala | L | -0.04 | (-0.11 - 0.03) | 0.30 | 0.43 |
| | R | -0.08 | (-0.15 - -0.01) | 0.03 | 0.07 |
| Caudate | L | 0.02 | (-0.05 - 0.09) | 0.60 | 0.72 |
| | R | -0.01 | (-0.08 - 0.06) | 0.72 | 0.81 |
| Hippocampus | L | -0.08 | (-0.15 - -0.01) | 0.02 | 0.06 |
| | R | **-0.16** | **(-0.23 - -0.08)** | **1.78e-05** | **3.45e-04** |
| Lateral Ventricles | L | **0.11** | **(0.04 - 0.18)** | **2.18e-03** | **0.01** |
| | R | **0.10** | **(0.03 - 0.17)** | **5.08e-03** | **0.02** |

| | | | | | |
|---|---|---|---|---|---|
| Nucleus Accumbens | L | -0.04 | (-0.11 - 0.03) | 0.29 | 0.42 |
| | R | -0.01 | (-0.08 - 0.06) | 0.84 | 0.89 |
| Pallidum | L | -0.02 | (-0.09 - 0.05) | 0.63 | 0.75 |
| | R | -0.02 | (-0.09 - 0.05) | 0.66 | 0.77 |
| Putamen | L | 0.00 | (-0.08 - 0.07) | 0.89 | 0.92 |
| | R | -0.01 | (-0.08 - 0.06) | 0.85 | 0.89 |
| Thalamus | L | -0.05 | (-0.12 - 0.02) | 0.19 | 0.30 |
| | R | -0.09 | (-0.16 - -0.01) | 0.02 | 0.05 |
| **Global** | | | | | |
| Estimated Intracranial Volume | N/A | **-0.13** | **(-0.2 - -0.06)** | **2.08e-04** | **2.17e-03** |
| Mean Cortical Thickness | | **-0.18** | **(-0.25 - -0.11)** | **7.05e-07** | **5.46e-05** |
| Total Surface Area | | **-0.15** | **(-0.22 - -0.08)** | **3.76e-05** | **5.63e-04** |

*Results marked in bold are significant at q<0.05*

**eTable 9.** CHR vs. HC antipsychotic medication effects

| | | Original Results | | | AP-corrected | | |
|---|---|---|---|---|---|---|---|
| Region of Interest | Hemisphere | Cohen's d | 95% Confidence Interval | q | Cohen's d | 95% Confidence Interval | q |
| **Cortical Thickness** | | | | | | | |
| Banks of the Superior Temporal Sulcus | L | -0.09 | (-0.16 - -0.02) | 0.03 | -0.09 | (-0.16 - -0.02) | 0.02 |
| | R | -0.13 | (-0.2 - -0.06) | 2.17e-03 | -0.12 | (-0.19 - -0.05) | 2.15e-03 |
| Caudal Middle Frontal | L | -0.12 | (-0.19 - -0.04) | 7.50e-03 | -0.10 | (-0.17 - -0.03) | 7.72e-03 |
| | R | -0.12 | (-0.2 - -0.05) | 4.13e-03 | -0.11 | (-0.19 - -0.04) | 3.42e-03 |
| Fusiform | L | -0.17 | (-0.24 - -0.1) | 7.07e-05 | -0.16 | (-0.23 - -0.09) | 1.84e-04 |
| | R | -0.11 | (-0.18 - -0.04) | 0.01 | -0.09 | (-0.16 - -0.02) | 0.01 |
| Inferior Parietal | L | -0.16 | (-0.24 - -0.09) | 1.65e-04 | -0.15 | (-0.22 - -0.08) | 2.63e-04 |
| | R | -0.11 | (-0.18 - -0.04) | 0.01 | -0.09 | (-0.17 - -0.02) | 0.01 |
| Inferior Temporal | L | -0.13 | (-0.2 - -0.06) | 2.95e-03 | -0.12 | (-0.19 - -0.05) | 2.92e-03 |
| | R | -0.15 | (-0.22 - -0.08) | 5.63e-04 | -0.12 | (-0.19 - -0.05) | 2.99e-03 |
| Insula | L | -0.17 | (-0.24 - -0.1) | 7.07e-05 | -0.17 | (-0.24 - -0.1) | 9.99e-05 |
| | R | -0.16 | (-0.23 - -0.08) | 3.45e-04 | -0.14 | (-0.21 - -0.07) | 4.67e-04 |
| Lateral Occipital | L | -0.12 | (-0.19 - -0.05) | 6.99e-03 | -0.12 | (-0.19 - -0.05) | 2.15e-03 |
| | R | -0.16 | (-0.23 - -0.09) | 2.70e-04 | -0.16 | (-0.23 - -0.09) | 1.03e-04 |
| Lateral Orbitofrontal | R | -0.09 | (-0.16 - -0.02) | 0.05 | -0.08 | (-0.15 - -0.01) | 0.03 |
| Medial Orbitofrontal | L | -0.13 | (-0.2 - -0.06) | 2.17e-03 | -0.13 | (-0.2 - -0.06) | 1.75e-03 |
| | R | -0.11 | (-0.18 - -0.03) | 0.01 | -0.11 | (-0.18 - -0.04) | 6.16e-03 |
| Middle Temporal | L | -0.10 | (-0.17 - -0.03) | 0.02 | -0.08 | (-0.15 - -0.01) | 0.02 |
| | R | -0.18 | (-0.26 - -0.11) | 5.46e-05 | -0.16 | (-0.24 - -0.09) | 1.03e-04 |
| Paracentral | L | -0.15 | (-0.22 - -0.08) | 5.58e-04 | -0.14 | (-0.21 - -0.07) | 4.67e-04 |
| | R | -0.11 | (-0.18 - -0.04) | 0.01 | -0.11 | (-0.18 - -0.03) | 6.28e-03 |
| Parahippocampal | L | -0.13 | (-0.2 - -0.06) | 3.26e-03 | -0.13 | (-0.2 - -0.06) | 1.37e-03 |
| Postcentral | L | -0.10 | (-0.17 - -0.02) | 0.03 | -0.09 | (-0.16 - -0.02) | 0.02 |
| | R | -0.12 | (-0.19 - -0.05) | 6.77e-03 | -0.12 | (-0.19 - -0.04) | 3.31e-03 |
| Posterior Cingulate | L | -0.11 | (-0.18 - -0.03) | 0.01 | -0.11 | (-0.18 - -0.04) | 5.01e-03 |
| | R | -0.13 | (-0.2 - -0.06) | 2.17e-03 | -0.12 | (-0.19 - -0.05) | 2.18e-03 |
| Precentral | L | -0.13 | (-0.2 - -0.06) | 2.72e-03 | -0.12 | (-0.19 - -0.05) | 2.49e-03 |
| | R | -0.13 | (-0.2 - -0.06) | 2.24e-03 | -0.12 | (-0.19 - -0.05) | 2.36e-03 |
| Precuneus | L | -0.11 | (-0.18 - -0.04) | 8.39e-03 | -0.11 | (-0.18 - -0.03) | 6.28e-03 |
| | R | -0.15 | (-0.22 - -0.08) | 5.63e-04 | -0.14 | (-0.21 - -0.06) | 9.16e-04 |
| Rostral Anterior Cingulate | L | -0.11 | (-0.18 - -0.04) | 0.01 | -0.11 | (-0.18 - -0.04) | 5.91e-03 |
| Rostral Middle Frontal | L | -0.13 | (-0.2 - -0.06) | 2.37e-03 | -0.12 | (-0.19 - -0.05) | 2.15e-03 |
| | R | -0.12 | (-0.19 - -0.04) | 7.50e-03 | -0.11 | (-0.18 - -0.04) | 4.40e-03 |
| Superior Frontal | L | -0.11 | (-0.18 - -0.04) | 0.01 | -0.10 | (-0.17 - -0.02) | 0.01 |
| | R | -0.09 | (-0.16 - -0.02) | 0.04 | -0.09 | (-0.16 - -0.02) | 0.01 |
| Superior Parietal | L | -0.10 | (-0.18 - -0.03) | 0.01 | -0.10 | (-0.17 - -0.03) | 6.92e-03 |
| | R | -0.12 | (-0.19 - -0.05) | 5.24e-03 | -0.12 | (-0.19 - -0.05) | 2.18e-03 |
| Superior Temporal | L | -0.12 | (-0.19 - -0.05) | 6.48e-03 | -0.10 | (-0.17 - -0.03) | 0.01 |
| | R | -0.15 | (-0.22 - -0.08) | 6.33e-04 | -0.13 | (-0.2 - -0.06) | 1.31e-03 |
| Supramarginal | L | -0.11 | (-0.18 - -0.04) | 0.01 | -0.09 | (-0.16 - -0.02) | 0.01 |
| | R | -0.10 | (-0.17 - -0.03) | 0.02 | -0.09 | (-0.16 - -0.02) | 0.02 |
| Transverse Temporal | R | -0.11 | (-0.18 - -0.04) | 0.01 | -0.11 | (-0.18 - -0.04) | 5.91e-03 |
| **Surface Area** | | | | | | | |
| Entorhinal | L | -0.09 | (-0.16 - -0.02) | 0.04 | -0.07 | (-0.15 - 0) | 0.04 |
| Inferior Parietal | R | -0.10 | (-0.17 - -0.03) | 0.02 | -0.09 | (-0.16 - -0.01) | 0.02 |

| | | | | | | | |
|---|---|---|---|---|---|---|---|
| Isthmus Cingulate | R | 0.09 | (0.02 - 0.16) | 0.03 | 0.08 | (0.01 - 0.15) | 0.02 |
| Pars Opercularis | L | -0.09 | (-0.16 - -0.02) | 0.03 | -0.07 | (-0.14 - 0) | 0.05 |
| Pericalcarine | R | -0.09 | (-0.17 - -0.02) | 0.03 | -0.09 | (-0.16 - -0.02) | 0.02 |
| Rostral Anterior Cingulate | L | -0.12 | (-0.19 - -0.04) | 7.50e-03 | -0.10 | (-0.17 - -0.03) | 7.77e-03 |
| Superior Frontal | R | -0.09 | (-0.16 - -0.02) | 0.03 | -0.08 | (-0.15 - 0) | 0.04 |
| Supramarginal | L | -0.11 | (-0.18 - -0.04) | 0.01 | -0.10 | (-0.17 - -0.03) | 0.01 |
| **Subcortical Volume** | | | | | | | |
| Hippocampus | R | -0.16 | (-0.23 - -0.08) | 3.45e-04 | -0.13 | (-0.2 - -0.06) | 1.21e-03 |
| Lateral Ventricles | L | 0.11 | (0.04 - 0.18) | 0.01 | 0.09 | (0.02 - 0.17) | 0.01 |
| | R | 0.10 | (0.03 - 0.17) | 0.02 | 0.09 | (0.02 - 0.16) | 0.02 |
| Thalamus | R | -0.09 | (-0.16 - -0.01) | 0.05 | -0.06 | (-0.13 - 0.01) | 0.08 |
| **Global** | | | | | | | |
| Estimated Intracranial Volume | N/A | -0.13 | (-0.2 - -0.06) | 2.17e-03 | -0.15 | (-0.22 - -0.07) | 4.50e-04 |
| Mean Cortical Thickness | N/A | -0.18 | (-0.25 - -0.11) | 5.46e-05 | -0.17 | (-0.24 - -0.1) | 9.99e-05 |
| Total Surface Area | N/A | -0.15 | (-0.22 - -0.08) | 5.63e-04 | -0.14 | (-0.21 - -0.07) | 6.65e-04 |

*Results reported under "Original Results" are from the CHR vs. HC post-ComBat mega analysis (see also eTable 7). All reported regions were significant at q<0.05. "AP-corrected" refers to post-ComBat mega analysis including antipsychotic medication as an additional covariate.*

**eTable 10.** CHR vs. HC equivalence testing results

| | | | Equivalence Testing Results | | | |
|---|---|---|---|---|---|---|
| **Region of Interest** | **Hemisphere** | **Cohen's d** | **90% CI** | **p ΔL** | **p ΔU** | **equivalent to SESOI** |
| **Cortical Thickness** | | | | | | |
| Banks of the Superior Temporal Sulcus | L | -0.09 | (-0.15 - -0.03) | 0.06 | 1.07e-11 | no |
| | R | -0.13 | (-0.19 - -0.07) | 0.32 | 2.48e-15 | no |
| Caudal Middle Frontal | L | -0.12 | (-0.17 - -0.06) | 0.17 | 1.08e-13 | no |
| | R | -0.12 | (-0.18 - -0.06) | 0.24 | 1.83e-14 | no |
| Fusiform | L | -0.17 | (-0.23 - -0.11) | 0.75 | 1.65e-19 | no |
| | R | -0.11 | (-0.17 - -0.05) | 0.13 | 2.97e-13 | no |
| Inferior Parietal | L | -0.16 | (-0.22 - -0.11) | 0.66 | 1.57e-18 | no |
| | R | -0.11 | (-0.17 - -0.05) | 0.14 | 2.55e-13 | no |
| Inferior Temporal | L | -0.13 | (-0.19 - -0.07) | 0.27 | 7.33e-15 | no |
| | R | -0.15 | (-0.21 - -0.09) | 0.48 | 8.84e-17 | no |
| Insula | L | -0.17 | (-0.23 - -0.11) | 0.74 | 2.24e-19 | no |
| | R | -0.16 | (-0.22 - -0.1) | 0.56 | 1.34e-17 | no |
| Lateral Occipital | L | -0.12 | (-0.18 - -0.06) | 0.18 | 7.41e-14 | no |
| | R | -0.16 | (-0.22 - -0.1) | 0.60 | 5.72e-18 | no |
| Lateral Orbitofrontal | R | -0.09 | (-0.15 - -0.03) | 0.04 | 2.86e-11 | yes |
| Medial Orbitofrontal | L | -0.13 | (-0.19 - -0.07) | 0.32 | 2.43e-15 | no |
| | R | -0.11 | (-0.16 - -0.05) | 0.11 | 7.90e-13 | no |
| Middle Temporal | L | -0.10 | (-0.16 - -0.04) | 0.07 | 4.59e-12 | no |
| | R | -0.18 | (-0.24 - -0.12) | 0.83 | 1.43e-20 | no |
| Paracentral | L | -0.15 | (-0.21 - -0.09) | 0.51 | 4.88e-17 | no |
| | R | -0.11 | (-0.17 - -0.05) | 0.12 | 5.15e-13 | no |
| Parahippocampal | L | -0.13 | (-0.19 - -0.07) | 0.26 | 9.15e-15 | no |
| Postcentral | L | -0.10 | (-0.16 - -0.04) | 0.07 | 7.36e-12 | no |
| | R | -0.12 | (-0.18 - -0.06) | 0.19 | 8.09e-14 | no |
| Posterior Cingulate | L | -0.11 | (-0.17 - -0.05) | 0.11 | 7.33e-13 | no |
| | R | -0.13 | (-0.19 - -0.07) | 0.32 | 2.56e-15 | no |
| Precentral | L | -0.13 | (-0.19 - -0.07) | 0.29 | 6.70e-15 | no |
| | R | -0.13 | (-0.19 - -0.07) | 0.32 | 3.62e-15 | no |
| Precuneus | L | -0.11 | (-0.17 - -0.05) | 0.16 | 1.54e-13 | no |
| | R | -0.15 | (-0.21 - -0.09) | 0.49 | 7.52e-17 | no |
| Rostral Anterior Cingulate | L | -0.11 | (-0.17 - -0.05) | 0.13 | 3.29e-13 | no |
| Rostral Middle Frontal | L | -0.13 | (-0.19 - -0.07) | 0.30 | 3.63e-15 | no |
| | R | -0.12 | (-0.17 - -0.06) | 0.17 | 1.03e-13 | no |
| Superior Frontal | L | -0.11 | (-0.17 - -0.05) | 0.12 | 4.57e-13 | no |
| | R | -0.09 | (-0.15 - -0.03) | 0.05 | 1.75e-11 | yes |
| Superior Parietal | L | -0.10 | (-0.16 - -0.05) | 0.10 | 9.70e-13 | no |
| | R | -0.12 | (-0.18 - -0.06) | 0.21 | 3.22e-14 | no |
| Superior Temporal | L | -0.12 | (-0.18 - -0.06) | 0.20 | 5.98e-14 | no |
| | R | -0.15 | (-0.21 - -0.09) | 0.46 | 1.34e-16 | no |
| Supramarginal | L | -0.11 | (-0.17 - -0.05) | 0.12 | 7.52e-13 | no |
| | R | -0.10 | (-0.16 - -0.04) | 0.07 | 5.01e-12 | no |
| Transverse Temporal | R | -0.11 | (-0.17 - -0.05) | 0.13 | 3.33e-13 | no |
| **Surface Area** | | | | | | |
| Entorhinal | L | -0.09 | (-0.15 - -0.03) | 0.05 | 1.39e-11 | no |
| Inferior Parietal | R | -0.10 | (-0.16 - -0.04) | 0.07 | 3.54e-12 | no |
| Isthmus Cingulate | R | 0.09 | (0.03 - 0.15) | 7.58e-12 | 0.06 | no |

| | | | | | | |
|---|---|---|---|---|---|---|
| Pars Opercularis | L | -0.09 | (-0.15 - -0.03) | 0.06 | 8.57e-12 | no |
| Pericalcarine | R | -0.09 | (-0.15 - -0.03) | 0.07 | 9.30e-12 | no |
| Rostral Anterior Cingulate | L | -0.12 | (-0.17 - -0.06) | 0.17 | 9.81e-14 | no |
| Superior Frontal | R | -0.09 | (-0.15 - -0.03) | 0.05 | 1.11e-11 | no |
| Supramarginal | L | -0.11 | (-0.17 - -0.05) | 0.13 | 5.03e-13 | no |
| **Subcortical Volume** | | | | | | |
| Hippocampus | R | -0.16 | (-0.21 - -0.1) | 0.56 | 1.46e-17 | no |
| Lateral Ventricles | L | 0.11 | (0.05 - 0.17) | 3.20e-13 | 0.15 | no |
| | R | 0.10 | (0.04 - 0.16) | 2.17e-12 | 0.09 | no |
| Thalamus | R | -0.09 | (-0.14 - -0.03) | 0.04 | 3.80e-11 | yes |
| **Global** | | | | | | |
| Estimated Intracranial Volume | N/A | -0.13 | (-0.19 - -0.07) | 0.33 | 1.67e-15 | no |
| Mean Cortical Thickness | N/A | -0.18 | (-0.24 - -0.12) | 0.79 | 3.94e-20 | no |
| Total Surface Area | N/A | -0.15 | (-0.21 - -0.09) | 0.49 | 5.73e-17 | no |

*ΔU/L refer to upper (+0.15) and lower (-0.15) equivalence bounds. SESOI = smallest effect size of interest. All regions of interest were significant at q<0.05 in the CHR vs. HC post-ComBat mega analysis (see also eTable 7).*

**eTable 11.** CHR vs. HC effect size overview (pre-ComBat mega and meta analysis)

| | | Mega Analysis (pre-ComBat) | | | | Meta Analysis (pre-ComBat) | | | |
|---|---|---|---|---|---|---|---|---|---|
| Region of Interest | Hemisphere | Cohen's d | 95% Confidence Interval | p | q | Cohen's d | 95% Confidence Interval | p | q |
| **Cortical Thickness** | | | | | | | | | |
| Banks of the Superior Temporal Sulcus | L | **-0.09** | **(-0.16 - -0.02)** | **0.02** | **0.05** | -0.09 | (-0.26 - 0.07) | 0.27 | 0.84 |
| | R | **-0.12** | **(-0.2 - -0.05)** | **6.03e-04** | **3.60e-03** | -0.15 | (-0.32 - 0.01) | 0.07 | 0.63 |
| Caudal Anterior Cingulate | L | **-0.09** | **(-0.16 - -0.02)** | **0.02** | **0.04** | -0.11 | (-0.28 - 0.05) | 0.19 | 0.65 |
| | R | -0.07 | (-0.14 - 0) | 0.05 | 0.10 | -0.09 | (-0.25 - 0.08) | 0.31 | 0.86 |
| Caudal Middle Frontal | L | **-0.12** | **(-0.19 - -0.05)** | **1.11e-03** | **5.42e-03** | -0.12 | (-0.28 - 0.05) | 0.16 | 0.65 |
| | R | **-0.12** | **(-0.19 - -0.05)** | **1.12e-03** | **5.42e-03** | -0.15 | (-0.31 - 0.02) | 0.08 | 0.63 |
| Cuneus | L | -0.01 | (-0.08 - 0.06) | 0.81 | 0.87 | -0.02 | (-0.18 - 0.15) | 0.82 | 0.97 |
| | R | -0.07 | (-0.14 - 0) | 0.06 | 0.11 | -0.06 | (-0.22 - 0.11) | 0.50 | 0.97 |
| Entorhinal | L | -0.01 | (-0.09 - 0.06) | 0.69 | 0.80 | -0.02 | (-0.18 - 0.15) | 0.83 | 0.97 |
| | R | -0.05 | (-0.12 - 0.02) | 0.16 | 0.27 | -0.05 | (-0.22 - 0.11) | 0.52 | 0.97 |
| Frontal Pole | L | -0.06 | (-0.13 - 0.01) | 0.11 | 0.20 | -0.06 | (-0.23 - 0.1) | 0.47 | 0.97 |
| | R | -0.07 | (-0.14 - 0) | 0.04 | 0.10 | -0.09 | (-0.25 - 0.08) | 0.29 | 0.86 |
| Fusiform | L | **-0.18** | **(-0.25 - -0.11)** | **5.98e-07** | **3.09e-05** | -0.17 | (-0.33 - 0) | 0.05 | 0.63 |
| | R | **-0.12** | **(-0.19 - -0.05)** | **1.01e-03** | **5.23e-03** | -0.09 | (-0.26 - 0.07) | 0.28 | 0.85 |
| Inferior Parietal | L | **-0.17** | **(-0.24 - -0.1)** | **1.72e-06** | **5.35e-05** | -0.18 | (-0.35 - -0.02) | 0.03 | 0.63 |
| | R | **-0.12** | **(-0.19 - -0.05)** | **9.84e-04** | **5.23e-03** | -0.14 | (-0.3 - 0.03) | 0.10 | 0.63 |
| Inferior Temporal | L | **-0.13** | **(-0.2 - -0.06)** | **2.67e-04** | **1.97e-03** | -0.14 | (-0.3 - 0.03) | 0.11 | 0.63 |
| | R | **-0.15** | **(-0.23 - -0.08)** | **2.02e-05** | **3.06e-04** | -0.15 | (-0.31 - 0.02) | 0.08 | 0.63 |
| Insula | L | **-0.17** | **(-0.24 - -0.1)** | **2.26e-06** | **5.83e-05** | -0.18 | (-0.34 - -0.01) | 0.04 | 0.63 |
| | R | **-0.16** | **(-0.23 - -0.08)** | **1.76e-05** | **3.03e-04** | -0.15 | (-0.31 - 0.02) | 0.08 | 0.63 |
| Isthmus Cingulate | L | -0.05 | (-0.12 - 0.02) | 0.20 | 0.33 | -0.06 | (-0.23 - 0.1) | 0.45 | 0.97 |
| | R | -0.08 | (-0.15 - -0.01) | 0.03 | 0.08 | -0.07 | (-0.24 - 0.09) | 0.40 | 0.93 |
| Lateral Occipital | L | **-0.13** | **(-0.2 - -0.05)** | **5.43e-04** | **3.51e-03** | -0.13 | (-0.3 - 0.03) | 0.12 | 0.63 |
| | R | **-0.16** | **(-0.23 - -0.09)** | **9.83e-06** | **1.91e-04** | -0.16 | (-0.33 - 0) | 0.05 | 0.63 |
| Lateral Orbitofrontal | L | -0.08 | (-0.16 - -0.01) | 0.02 | 0.05 | -0.13 | (-0.29 - 0.04) | 0.13 | 0.63 |
| | R | **-0.10** | **(-0.17 - -0.03)** | **5.67e-03** | **0.02** | -0.12 | (-0.29 - 0.04) | 0.14 | 0.64 |
| Lingual | L | -0.03 | (-0.1 - 0.04) | 0.36 | 0.51 | -0.04 | (-0.21 - 0.13) | 0.64 | 0.97 |
| | R | -0.02 | (-0.1 - 0.05) | 0.50 | 0.65 | -0.02 | (-0.19 - 0.14) | 0.77 | 0.97 |
| Medial Orbitofrontal | L | **-0.15** | **(-0.22 - -0.08)** | **2.52e-05** | **3.06e-04** | -0.19 | (-0.36 - -0.03) | 0.02 | 0.63 |
| | R | **-0.13** | **(-0.2 - -0.06)** | **3.41e-04** | **2.40e-03** | -0.14 | (-0.3 - 0.03) | 0.10 | 0.63 |
| Middle Temporal | L | **-0.10** | **(-0.17 - -0.03)** | **8.03e-03** | **0.03** | -0.11 | (-0.28 - 0.05) | 0.17 | 0.65 |
| | R | **-0.18** | **(-0.26 - -0.11)** | **4.18e-07** | **3.09e-05** | -0.19 | (-0.36 - -0.03) | 0.02 | 0.63 |
| Paracentral | L | **-0.15** | **(-0.22 - -0.08)** | **2.56e-05** | **3.06e-04** | -0.17 | (-0.34 - -0.01) | 0.04 | 0.63 |
| | R | **-0.12** | **(-0.19 - -0.05)** | **1.16e-03** | **5.47e-03** | -0.10 | (-0.27 - 0.06) | 0.22 | 0.73 |
| Parahippocampal | L | **-0.11** | **(-0.18 - -0.04)** | **2.12e-03** | **9.13e-03** | -0.13 | (-0.29 - 0.04) | 0.13 | 0.63 |
| | R | -0.07 | (-0.14 - 0) | 0.05 | 0.11 | -0.08 | (-0.25 - 0.08) | 0.34 | 0.87 |
| Pars Opercularis | L | -0.07 | (-0.14 - 0) | 0.07 | 0.13 | -0.04 | (-0.2 - 0.13) | 0.68 | 0.97 |
| | R | -0.08 | (-0.15 - -0.01) | 0.02 | 0.05 | -0.07 | (-0.24 - 0.09) | 0.38 | 0.92 |
| Pars Orbitalis | L | -0.06 | (-0.13 - 0.01) | 0.09 | 0.16 | -0.09 | (-0.26 - 0.07) | 0.28 | 0.85 |
| | R | 0.00 | (-0.07 - 0.07) | 0.99 | 1.00 | -0.02 | (-0.19 - 0.14) | 0.78 | 0.97 |
| Pars Triangularis | L | 0.00 | (-0.07 - 0.07) | 0.95 | 0.97 | -0.03 | (-0.19 - 0.14) | 0.76 | 0.97 |
| | R | -0.07 | (-0.14 - 0) | 0.04 | 0.10 | -0.09 | (-0.25 - 0.08) | 0.30 | 0.86 |
| Pericalcarine | L | 0.02 | (-0.05 - 0.1) | 0.50 | 0.65 | 0.01 | (-0.16 - 0.18) | 0.90 | 0.99 |
| | R | 0.05 | (-0.03 - 0.12) | 0.22 | 0.35 | 0.03 | (-0.14 - 0.19) | 0.76 | 0.97 |
| | L | **-0.10** | **(-0.17 - -0.03)** | **7.87e-03** | **0.03** | -0.08 | (-0.25 - 0.08) | 0.33 | 0.87 |

| | | | | | | | | | |
|---|---|---|---|---|---|---|---|---|---|
| Postcentral | R | **-0.11** | **(-0.19 - -0.04)** | **1.79e-03** | **7.92e-03** | -0.13 | (-0.29 - 0.04) | 0.14 | 0.63 |
| Posterior Cingulate | L | **-0.12** | **(-0.19 - -0.05)** | **6.52e-04** | **3.74e-03** | -0.13 | (-0.29 - 0.04) | 0.13 | 0.63 |
| | R | **-0.16** | **(-0.23 - -0.09)** | **9.78e-06** | **1.91e-04** | -0.16 | (-0.33 - 0.01) | 0.06 | 0.63 |
| Precentral | L | **-0.13** | **(-0.21 - -0.06)** | **2.42e-04** | **1.87e-03** | -0.12 | (-0.29 - 0.04) | 0.15 | 0.65 |
| | R | **-0.13** | **(-0.21 - -0.06)** | **2.27e-04** | **1.86e-03** | -0.14 | (-0.31 - 0.03) | 0.10 | 0.63 |
| Precuneus | L | **-0.15** | **(-0.22 - -0.08)** | **2.38e-05** | **3.06e-04** | -0.12 | (-0.29 - 0.04) | 0.14 | 0.63 |
| | R | **-0.18** | **(-0.25 - -0.11)** | **9.86e-07** | **3.82e-05** | -0.17 | (-0.33 - 0) | 0.05 | 0.63 |
| Rostral Anterior Cingulate | L | **-0.10** | **(-0.17 - -0.03)** | **4.09e-03** | **0.02** | -0.11 | (-0.28 - 0.05) | 0.17 | 0.65 |
| | R | -0.08 | (-0.15 - -0.01) | 0.03 | 0.07 | -0.11 | (-0.28 - 0.05) | 0.18 | 0.65 |
| Rostral Middle Frontal | L | **-0.15** | **(-0.22 - -0.08)** | **5.36e-05** | **5.94e-04** | -0.17 | (-0.34 - -0.01) | 0.04 | 0.63 |
| | R | **-0.14** | **(-0.21 - -0.07)** | **1.24e-04** | **1.13e-03** | -0.17 | (-0.34 - 0) | 0.04 | 0.63 |
| Superior Frontal | L | **-0.12** | **(-0.2 - -0.05)** | **5.70e-04** | **3.53e-03** | -0.14 | (-0.3 - 0.03) | 0.11 | 0.63 |
| | R | **-0.11** | **(-0.18 - -0.04)** | **2.53e-03** | **0.01** | -0.13 | (-0.29 - 0.04) | 0.13 | 0.63 |
| Superior Parietal | L | **-0.11** | **(-0.18 - -0.04)** | **3.42e-03** | **0.01** | -0.13 | (-0.3 - 0.04) | 0.12 | 0.63 |
| | R | **-0.13** | **(-0.2 - -0.06)** | **5.05e-04** | **3.40e-03** | -0.15 | (-0.32 - 0.01) | 0.07 | 0.63 |
| Superior Temporal | L | **-0.11** | **(-0.18 - -0.04)** | **1.78e-03** | **7.92e-03** | -0.11 | (-0.28 - 0.05) | 0.19 | 0.65 |
| | R | **-0.14** | **(-0.22 - -0.07)** | **6.63e-05** | **6.85e-04** | -0.15 | (-0.31 - 0.02) | 0.08 | 0.63 |
| Supramarginal | L | **-0.11** | **(-0.18 - -0.04)** | **2.24e-03** | **9.37e-03** | -0.13 | (-0.29 - 0.04) | 0.13 | 0.63 |
| | R | **-0.10** | **(-0.17 - -0.03)** | **5.20e-03** | **0.02** | -0.13 | (-0.29 - 0.04) | 0.13 | 0.63 |
| Temporal Pole | L | -0.03 | (-0.11 - 0.04) | 0.34 | 0.49 | -0.04 | (-0.21 - 0.12) | 0.61 | 0.97 |
| | R | -0.06 | (-0.13 - 0.01) | 0.10 | 0.19 | -0.07 | (-0.24 - 0.09) | 0.39 | 0.92 |
| Transverse Temporal | L | -0.07 | (-0.14 - 0) | 0.04 | 0.10 | -0.05 | (-0.22 - 0.11) | 0.52 | 0.97 |
| | R | **-0.10** | **(-0.18 - -0.03)** | **3.97e-03** | **0.02** | -0.09 | (-0.25 - 0.08) | 0.30 | 0.86 |
| **Surface Area** | | | | | | | | | |
| Banks of the Superior Temporal Sulcus | L | -0.06 | (-0.13 - 0.01) | 0.08 | 0.16 | -0.03 | (-0.2 - 0.13) | 0.68 | 0.97 |
| | R | -0.08 | (-0.15 - -0.01) | 0.03 | 0.08 | -0.04 | (-0.2 - 0.13) | 0.66 | 0.97 |
| Caudal Anterior Cingulate | L | -0.01 | (-0.08 - 0.06) | 0.84 | 0.88 | -0.01 | (-0.17 - 0.16) | 0.92 | 0.99 |
| | R | -0.07 | (-0.14 - 0) | 0.05 | 0.10 | -0.05 | (-0.22 - 0.11) | 0.51 | 0.97 |
| Caudal Middle Frontal | L | -0.05 | (-0.12 - 0.02) | 0.16 | 0.28 | -0.03 | (-0.19 - 0.14) | 0.73 | 0.97 |
| | R | -0.02 | (-0.09 - 0.05) | 0.64 | 0.79 | -0.01 | (-0.17 - 0.16) | 0.93 | 0.99 |
| Cuneus | L | -0.04 | (-0.11 - 0.03) | 0.25 | 0.40 | -0.03 | (-0.19 - 0.14) | 0.73 | 0.97 |
| | R | -0.07 | (-0.14 - 0) | 0.05 | 0.10 | -0.04 | (-0.2 - 0.13) | 0.66 | 0.97 |
| Entorhinal | L | **-0.09** | **(-0.16 - -0.02)** | **0.02** | **0.05** | -0.08 | (-0.25 - 0.08) | 0.33 | 0.87 |
| | R | -0.02 | (-0.09 - 0.05) | 0.62 | 0.76 | -0.02 | (-0.19 - 0.14) | 0.79 | 0.97 |
| Frontal Pole | L | 0.04 | (-0.03 - 0.11) | 0.31 | 0.47 | 0.05 | (-0.12 - 0.21) | 0.56 | 0.97 |
| | R | 0.04 | (-0.03 - 0.11) | 0.32 | 0.48 | 0.03 | (-0.13 - 0.2) | 0.69 | 0.97 |
| Fusiform | L | -0.03 | (-0.1 - 0.04) | 0.36 | 0.51 | 0.00 | (-0.16 - 0.17) | 0.98 | 1.00 |
| | R | -0.07 | (-0.14 - 0) | 0.06 | 0.12 | -0.05 | (-0.21 - 0.12) | 0.59 | 0.97 |
| Inferior Parietal | L | -0.06 | (-0.13 - 0.01) | 0.11 | 0.20 | -0.03 | (-0.19 - 0.14) | 0.75 | 0.97 |
| | R | **-0.09** | **(-0.16 - -0.02)** | **0.01** | **0.04** | -0.04 | (-0.2 - 0.13) | 0.67 | 0.97 |
| Inferior Temporal | L | -0.02 | (-0.09 - 0.05) | 0.57 | 0.72 | 0.02 | (-0.15 - 0.18) | 0.82 | 0.97 |
| | R | -0.02 | (-0.1 - 0.05) | 0.50 | 0.65 | 0.01 | (-0.16 - 0.17) | 0.92 | 0.99 |
| Insula | L | -0.06 | (-0.13 - 0.01) | 0.10 | 0.19 | -0.02 | (-0.18 - 0.15) | 0.82 | 0.97 |
| | R | -0.03 | (-0.11 - 0.04) | 0.34 | 0.49 | -0.02 | (-0.18 - 0.14) | 0.81 | 0.97 |
| Isthmus Cingulate | L | 0.01 | (-0.06 - 0.08) | 0.84 | 0.88 | 0.01 | (-0.15 - 0.18) | 0.86 | 0.97 |
| | R | **0.09** | **(0.02 - 0.16)** | **0.02** | **0.05** | 0.11 | (-0.05 - 0.28) | 0.18 | 0.65 |
| Lateral Occipital | L | 0.01 | (-0.06 - 0.08) | 0.82 | 0.88 | 0.03 | (-0.14 - 0.19) | 0.75 | 0.97 |
| | R | -0.01 | (-0.08 - 0.06) | 0.86 | 0.90 | 0.02 | (-0.14 - 0.19) | 0.78 | 0.97 |
| Lateral Orbitofrontal | L | -0.02 | (-0.09 - 0.05) | 0.55 | 0.69 | 0.00 | (-0.16 - 0.17) | 0.95 | 0.99 |
| | R | -0.01 | (-0.09 - 0.06) | 0.68 | 0.80 | -0.01 | (-0.17 - 0.16) | 0.94 | 0.99 |

| | | | | | | | | | |
|---|---|---|---|---|---|---|---|---|---|
| Lingual | L | -0.05 | (-0.12 - 0.02) | 0.18 | 0.30 | -0.03 | (-0.19 - 0.14) | 0.74 | 0.97 |
| | R | -0.01 | (-0.08 - 0.06) | 0.79 | 0.86 | 0.02 | (-0.15 - 0.18) | 0.86 | 0.97 |
| Medial Orbitofrontal | L | -0.01 | (-0.08 - 0.06) | 0.74 | 0.82 | 0.02 | (-0.14 - 0.19) | 0.79 | 0.97 |
| | R | -0.07 | (-0.14 - 0) | 0.05 | 0.10 | -0.04 | (-0.21 - 0.12) | 0.60 | 0.97 |
| Middle Temporal | L | 0.02 | (-0.06 - 0.09) | 0.66 | 0.80 | 0.05 | (-0.11 - 0.22) | 0.53 | 0.97 |
| | R | -0.07 | (-0.14 - 0) | 0.05 | 0.10 | 0.00 | (-0.17 - 0.16) | 1.00 | 1.00 |
| Paracentral | L | 0.00 | (-0.07 - 0.07) | 1.00 | 1.00 | 0.00 | (-0.17 - 0.16) | 0.99 | 1.00 |
| | R | 0.01 | (-0.06 - 0.08) | 0.73 | 0.82 | 0.00 | (-0.16 - 0.17) | 0.96 | 0.99 |
| Parahippocampal | L | -0.04 | (-0.11 - 0.03) | 0.27 | 0.42 | -0.02 | (-0.18 - 0.15) | 0.84 | 0.97 |
| | R | 0.06 | (-0.01 - 0.13) | 0.11 | 0.20 | 0.10 | (-0.06 - 0.27) | 0.23 | 0.73 |
| Pars Opercularis | L | **-0.09** | **(-0.16 - -0.02)** | **0.01** | **0.03** | -0.07 | (-0.24 - 0.09) | 0.38 | 0.92 |
| | R | -0.01 | (-0.08 - 0.06) | 0.72 | 0.82 | 0.01 | (-0.16 - 0.17) | 0.94 | 0.99 |
| Pars Orbitalis | L | 0.02 | (-0.05 - 0.09) | 0.51 | 0.65 | 0.04 | (-0.13 - 0.2) | 0.66 | 0.97 |
| | R | 0.03 | (-0.04 - 0.1) | 0.35 | 0.51 | 0.03 | (-0.14 - 0.19) | 0.76 | 0.97 |
| Pars Triangularis | L | -0.06 | (-0.13 - 0.01) | 0.12 | 0.21 | -0.04 | (-0.21 - 0.12) | 0.62 | 0.97 |
| | R | 0.01 | (-0.06 - 0.08) | 0.71 | 0.82 | 0.00 | (-0.16 - 0.17) | 0.99 | 1.00 |
| Pericalcarine | L | -0.05 | (-0.12 - 0.02) | 0.18 | 0.31 | -0.04 | (-0.21 - 0.12) | 0.61 | 0.97 |
| | R | **-0.09** | **(-0.16 - -0.02)** | **0.02** | **0.05** | -0.08 | (-0.25 - 0.08) | 0.32 | 0.86 |
| Postcentral | L | -0.01 | (-0.08 - 0.06) | 0.84 | 0.88 | 0.03 | (-0.14 - 0.2) | 0.72 | 0.97 |
| | R | -0.04 | (-0.11 - 0.03) | 0.32 | 0.48 | -0.05 | (-0.22 - 0.11) | 0.52 | 0.97 |
| Posterior Cingulate | L | -0.03 | (-0.1 - 0.04) | 0.42 | 0.57 | -0.02 | (-0.19 - 0.14) | 0.80 | 0.97 |
| | R | -0.06 | (-0.13 - 0.01) | 0.10 | 0.18 | -0.04 | (-0.21 - 0.12) | 0.60 | 0.97 |
| Precentral | L | 0.05 | (-0.02 - 0.12) | 0.18 | 0.31 | 0.06 | (-0.11 - 0.22) | 0.49 | 0.97 |
| | R | 0.01 | (-0.06 - 0.08) | 0.72 | 0.82 | 0.01 | (-0.16 - 0.17) | 0.94 | 0.99 |
| Precuneus | L | -0.03 | (-0.1 - 0.04) | 0.41 | 0.57 | -0.02 | (-0.19 - 0.14) | 0.80 | 0.97 |
| | R | -0.03 | (-0.1 - 0.04) | 0.39 | 0.54 | -0.03 | (-0.19 - 0.14) | 0.73 | 0.97 |
| Rostral Anterior Cingulate | L | **-0.10** | **(-0.17 - -0.03)** | **5.11e-03** | **0.02** | -0.07 | (-0.24 - 0.09) | 0.39 | 0.92 |
| | R | -0.05 | (-0.12 - 0.02) | 0.14 | 0.24 | -0.02 | (-0.19 - 0.14) | 0.80 | 0.97 |
| Rostral Middle Frontal | L | -0.01 | (-0.09 - 0.06) | 0.69 | 0.80 | -0.01 | (-0.17 - 0.16) | 0.94 | 0.99 |
| | R | -0.03 | (-0.1 - 0.04) | 0.40 | 0.55 | -0.02 | (-0.18 - 0.15) | 0.83 | 0.97 |
| Superior Frontal | L | -0.03 | (-0.1 - 0.04) | 0.43 | 0.57 | -0.04 | (-0.2 - 0.13) | 0.68 | 0.97 |
| | R | **-0.09** | **(-0.16 - -0.02)** | **0.01** | **0.04** | -0.07 | (-0.23 - 0.1) | 0.42 | 0.94 |
| Superior Parietal | L | 0.00 | (-0.07 - 0.07) | 0.93 | 0.96 | 0.02 | (-0.15 - 0.18) | 0.84 | 0.97 |
| | R | 0.01 | (-0.06 - 0.08) | 0.87 | 0.90 | 0.03 | (-0.13 - 0.2) | 0.71 | 0.97 |
| Superior Temporal | L | -0.02 | (-0.09 - 0.05) | 0.52 | 0.66 | 0.02 | (-0.15 - 0.18) | 0.86 | 0.97 |
| | R | -0.05 | (-0.12 - 0.02) | 0.19 | 0.31 | -0.01 | (-0.17 - 0.16) | 0.94 | 0.99 |
| Supramarginal | L | **-0.11** | **(-0.18 - -0.04)** | **3.24e-03** | **0.01** | -0.07 | (-0.24 - 0.09) | 0.38 | 0.92 |
| | R | -0.07 | (-0.14 - 0) | 0.06 | 0.12 | -0.07 | (-0.24 - 0.09) | 0.38 | 0.92 |
| Temporal Pole | L | -0.02 | (-0.1 - 0.05) | 0.50 | 0.65 | -0.03 | (-0.19 - 0.14) | 0.75 | 0.97 |
| | R | 0.04 | (-0.03 - 0.11) | 0.23 | 0.36 | 0.06 | (-0.1 - 0.23) | 0.45 | 0.97 |
| Transverse Temporal | L | -0.04 | (-0.11 - 0.03) | 0.31 | 0.47 | -0.01 | (-0.17 - 0.16) | 0.93 | 0.99 |
| | R | -0.01 | (-0.08 - 0.06) | 0.73 | 0.82 | -0.02 | (-0.18 - 0.15) | 0.84 | 0.97 |
| **Subcortical Volume** | | | | | | | | | |
| Amygdala | L | -0.03 | (-0.1 - 0.04) | 0.45 | 0.60 | 0.02 | (-0.15 - 0.18) | 0.84 | 0.97 |
| | R | -0.07 | (-0.14 - 0) | 0.04 | 0.10 | -0.04 | (-0.2 - 0.13) | 0.67 | 0.97 |
| Caudate | L | 0.01 | (-0.06 - 0.09) | 0.69 | 0.80 | 0.03 | (-0.13 - 0.2) | 0.68 | 0.97 |
| | R | -0.01 | (-0.08 - 0.06) | 0.81 | 0.87 | 0.00 | (-0.17 - 0.16) | 0.98 | 1.00 |
| Hippocampus | L | -0.08 | (-0.15 - 0) | 0.04 | 0.09 | -0.07 | (-0.23 - 0.1) | 0.42 | 0.94 |
| | R | **-0.14** | **(-0.21 - -0.07)** | **8.64e-05** | **8.37e-04** | -0.14 | (-0.3 - 0.03) | 0.10 | 0.63 |
| | L | **0.10** | **(0.03 - 0.17)** | **6.90e-03** | **0.02** | 0.10 | (-0.06 - 0.27) | 0.23 | 0.73 |

| | | | | | | | | | |
|---|---|---|---|---|---|---|---|---|---|
| Lateral Ventricles | R | **0.09** | **(0.02 - 0.16)** | **0.01** | **0.04** | 0.09 | (-0.08 - 0.25) | 0.31 | 0.86 |
| Nucleus Accumbens | L | -0.04 | (-0.11 - 0.03) | 0.32 | 0.48 | -0.02 | (-0.19 - 0.14) | 0.79 | 0.97 |
| | R | 0.00 | (-0.07 - 0.07) | 0.97 | 0.98 | 0.02 | (-0.15 - 0.18) | 0.85 | 0.97 |
| Pallidum | L | -0.02 | (-0.09 - 0.05) | 0.58 | 0.72 | 0.05 | (-0.11 - 0.22) | 0.54 | 0.97 |
| | R | -0.01 | (-0.08 - 0.06) | 0.74 | 0.82 | 0.04 | (-0.12 - 0.21) | 0.63 | 0.97 |
| Putamen | L | -0.02 | (-0.09 - 0.05) | 0.65 | 0.79 | 0.04 | (-0.13 - 0.2) | 0.67 | 0.97 |
| | R | -0.01 | (-0.08 - 0.06) | 0.74 | 0.82 | 0.03 | (-0.14 - 0.19) | 0.74 | 0.97 |
| Thalamus | L | -0.03 | (-0.1 - 0.04) | 0.36 | 0.51 | -0.01 | (-0.18 - 0.15) | 0.88 | 0.98 |
| | R | -0.07 | (-0.14 - 0) | 0.04 | 0.10 | -0.03 | (-0.19 - 0.14) | 0.73 | 0.97 |
| **Global** | | | | | | | | | |
| Estimated Intracranial Volume | N/A | **-0.12** | **(-0.19 - -0.05)** | **9.11e-04** | **5.04e-03** | -0.12 | (-0.28 - 0.05) | 0.16 | 0.65 |
| Mean Cortical Thickness | | **-0.20** | **(-0.27 - -0.13)** | **2.59e-08** | **4.02e-06** | -0.20 | (-0.36 - -0.03) | 0.02 | 0.63 |
| Total Surface Area | | **-0.14** | **(-0.21 - -0.07)** | **1.49e-04** | **1.28e-03** | -0.12 | (-0.28 - 0.05) | 0.17 | 0.65 |

*Results marked in bold are significant at q<0.05*

**eTable 12A.** CHR subgroup (APS vs. BIPS vs. GRD vs. HC) effect size overview

| | | Overall Test Statistics | | | APS < HC | | | BIPS < HC | | | GRD < HC | | |
|---|---|---|---|---|---|---|---|---|---|---|---|---|---|
| Region of Interest | Hemisphere | F | p | q | Cohen's d | 95% Confidence Interval | p | Cohen's d | 95% Confidence Interval | p | Cohen's d | 95% Confidence Interval | p |
| **Cortical Thickness** | | | | | | | | | | | | | |
| Banks of the Superior Temporal Sulcus | L | 2.41 | 0.07 | 0.19 | **-0.09** | **(-0.17 - -0.02)** | **0.02** | 0.05 | (-0.24 - 0.33) | 0.75 | 0.06 | (-0.15 - 0.27) | 0.56 |
| | R | 4.18 | 5.84e-03 | 0.06 | **-0.13** | **(-0.21 - -0.06)** | **6.47e-04** | 0.06 | (-0.22 - 0.34) | 0.68 | -0.10 | (-0.31 - 0.11) | 0.36 |
| Caudal Anterior Cingulate | L | 1.81 | 0.14 | 0.30 | -0.07 | (-0.14 - 0.01) | 0.09 | -0.26 | (-0.55 - 0.02) | 0.07 | -0.01 | (-0.22 - 0.2) | 0.94 |
| | R | 3.94 | 8.08e-03 | 0.06 | **-0.08** | **(-0.16 - -0.01)** | **0.04** | -0.22 | (-0.5 - 0.06) | 0.13 | 0.21 | (0 - 0.42) | 0.05 |
| Caudal Middle Frontal | L | 2.36 | 0.07 | 0.20 | **-0.09** | **(-0.16 - -0.01)** | **0.02** | -0.22 | (-0.51 - 0.07) | 0.14 | 0.02 | (-0.18 - 0.23) | 0.82 |
| | R | 3.98 | 7.73e-03 | 0.06 | **-0.12** | **(-0.19 - -0.04)** | **2.33e-03** | 0.06 | (-0.23 - 0.34) | 0.69 | 0.08 | (-0.12 - 0.29) | 0.42 |
| Cuneus | L | 1.44 | 0.23 | 0.41 | 0.00 | (-0.08 - 0.08) | 0.97 | 0.28 | (-0.01 - 0.57) | 0.06 | 0.09 | (-0.12 - 0.29) | 0.41 |
| | R | 2.55 | 0.05 | 0.17 | -0.04 | (-0.12 - 0.03) | 0.26 | **0.34** | **(0.06 - 0.62)** | **0.02** | -0.02 | (-0.23 - 0.18) | 0.83 |
| Entorhinal | L | 0.29 | 0.83 | 0.91 | 0.01 | (-0.07 - 0.08) | 0.83 | -0.09 | (-0.38 - 0.19) | 0.52 | -0.06 | (-0.26 - 0.15) | 0.58 |
| | R | 0.75 | 0.52 | 0.70 | -0.03 | (-0.1 - 0.05) | 0.51 | -0.15 | (-0.43 - 0.13) | 0.30 | -0.12 | (-0.32 - 0.09) | 0.27 |
| Frontal Pole | L | 1.21 | 0.30 | 0.49 | -0.05 | (-0.13 - 0.02) | 0.19 | 0.17 | (-0.12 - 0.45) | 0.25 | 0.01 | (-0.2 - 0.21) | 0.95 |
| | R | 1.47 | 0.22 | 0.40 | -0.07 | (-0.14 - 0.01) | 0.09 | -0.19 | (-0.48 - 0.1) | 0.20 | 0.03 | (-0.18 - 0.24) | 0.77 |
| Fusiform | L | **5.81** | **5.92e-04** | **0.02** | **-0.16** | **(-0.24 - -0.08)** | **4.41e-05** | -0.08 | (-0.36 - 0.2) | 0.57 | 0.00 | (-0.2 - 0.21) | 0.98 |
| | R | 1.86 | 0.13 | 0.29 | **-0.09** | **(-0.17 - -0.01)** | **0.02** | 0.00 | (-0.28 - 0.29) | 0.98 | -0.07 | (-0.28 - 0.13) | 0.48 |
| Inferior Parietal | L | **6.35** | **2.75e-04** | **0.02** | **-0.16** | **(-0.23 - -0.08)** | **5.57e-05** | -0.22 | (-0.51 - 0.06) | 0.13 | 0.05 | (-0.15 - 0.26) | 0.61 |
| | R | 3.93 | 8.24e-03 | 0.06 | **-0.12** | **(-0.19 - -0.04)** | **2.87e-03** | -0.07 | (-0.35 - 0.22) | 0.64 | 0.11 | (-0.09 - 0.32) | 0.28 |
| Inferior Temporal | L | 2.94 | 0.03 | 0.12 | **-0.10** | **(-0.18 - -0.03)** | **7.65e-03** | -0.10 | (-0.39 - 0.18) | 0.49 | -0.19 | (-0.39 - 0.02) | 0.08 |
| | R | 3.83 | 9.51e-03 | 0.06 | **-0.13** | **(-0.2 - -0.05)** | **1.10e-03** | -0.16 | (-0.45 - 0.12) | 0.27 | -0.13 | (-0.34 - 0.08) | 0.22 |
| Insula | L | **5.56** | **8.41e-04** | **0.02** | **-0.15** | **(-0.23 - -0.07)** | **1.14e-04** | -0.06 | (-0.34 - 0.22) | 0.68 | **-0.21** | **(-0.42 - -0.01)** | **0.04** |
| | R | **4.80** | **2.46e-03** | **0.04** | **-0.14** | **(-0.21 - -0.06)** | **3.83e-04** | -0.25 | (-0.54 - 0.03) | 0.08 | -0.11 | (-0.31 - 0.1) | 0.31 |
| Isthmus Cingulate | L | 0.58 | 0.63 | 0.77 | 0.00 | (-0.08 - 0.07) | 0.91 | -0.18 | (-0.47 - 0.1) | 0.22 | 0.04 | (-0.16 - 0.25) | 0.69 |
| | R | 0.46 | 0.71 | 0.84 | -0.05 | (-0.12 - 0.03) | 0.24 | -0.04 | (-0.32 - 0.25) | 0.81 | -0.02 | (-0.22 - 0.19) | 0.88 |
| Lateral Occipital | L | 3.72 | 0.01 | 0.06 | **-0.12** | **(-0.19 - -0.04)** | **2.47e-03** | -0.13 | (-0.41 - 0.16) | 0.39 | 0.08 | (-0.13 - 0.28) | 0.48 |
| | R | **4.58** | **3.31e-03** | **0.05** | **-0.14** | **(-0.22 - -0.06)** | **3.37e-04** | -0.21 | (-0.49 - 0.08) | 0.16 | -0.07 | (-0.28 - 0.14) | 0.49 |
| | | | | | | **(-0.16 -** | | | (-0.34 - | | | (-0.14 - | |

| | | | | | | | | | | | | | |
|---|---|---|---|---|---|---|---|---|---|---|---|---|---|
| Lateral Orbitofrontal | L | 2.03 | 0.11 | 0.27 | **-0.09** | **-0.01)** | **0.03** | -0.06 | 0.23) | 0.69 | 0.07 | 0.27) | 0.52 |
| | R | 1.71 | 0.16 | 0.32 | **-0.08** | **(-0.16 - -0.01)** | **0.04** | 0.00 | (-0.29 - 0.29) | 0.98 | 0.04 | (-0.17 - 0.25) | 0.70 |
| Lingual | L | 1.87 | 0.13 | 0.29 | -0.04 | (-0.11 - 0.04) | 0.35 | 0.27 | (-0.01 - 0.56) | 0.06 | 0.07 | (-0.14 - 0.28) | 0.51 |
| | R | 0.23 | 0.88 | 0.94 | 0.00 | (-0.07 - 0.08) | 0.95 | 0.10 | (-0.19 - 0.38) | 0.51 | 0.06 | (-0.15 - 0.26) | 0.61 |
| Medial Orbitofrontal | L | **4.57** | **3.38e-03** | **0.05** | **-0.14** | **(-0.22 - -0.06)** | **3.28e-04** | -0.03 | (-0.31 - 0.26) | 0.85 | -0.16 | (-0.36 - 0.05) | 0.13 |
| | R | 2.61 | 0.05 | 0.16 | **-0.11** | **(-0.18 - -0.03)** | **6.82e-03** | -0.03 | (-0.31 - 0.26) | 0.84 | 0.02 | (-0.19 - 0.22) | 0.88 |
| Middle Temporal | L | 1.95 | 0.12 | 0.28 | **-0.09** | **(-0.17 - -0.02)** | **0.02** | -0.10 | (-0.38 - 0.19) | 0.52 | -0.04 | (-0.25 - 0.16) | 0.69 |
| | R | **7.38** | **6.35e-05** | **9.84e-03** | **-0.18** | **(-0.26 - -0.1)** | **4.32e-06** | -0.21 | (-0.49 - 0.07) | 0.15 | -0.04 | (-0.25 - 0.16) | 0.68 |
| Paracentral | L | **5.17** | **1.45e-03** | **0.03** | **-0.14** | **(-0.22 - -0.07)** | **2.07e-04** | 0.08 | (-0.21 - 0.36) | 0.58 | 0.01 | (-0.2 - 0.21) | 0.95 |
| | R | 3.07 | 0.03 | 0.12 | **-0.09** | **(-0.17 - -0.02)** | **0.02** | 0.21 | (-0.07 - 0.49) | 0.14 | 0.02 | (-0.18 - 0.23) | 0.83 |
| Parahippocampal | L | 3.82 | 9.55e-03 | 0.06 | **-0.12** | **(-0.19 - -0.04)** | **2.87e-03** | 0.16 | (-0.12 - 0.44) | 0.27 | -0.11 | (-0.31 - 0.1) | 0.31 |
| | R | 1.31 | 0.27 | 0.46 | -0.06 | (-0.14 - 0.02) | 0.12 | 0.13 | (-0.15 - 0.41) | 0.38 | -0.08 | (-0.29 - 0.12) | 0.43 |
| Pars Opercularis | L | 0.70 | 0.55 | 0.71 | -0.04 | (-0.11 - 0.04) | 0.36 | 0.14 | (-0.15 - 0.43) | 0.33 | 0.01 | (-0.2 - 0.22) | 0.92 |
| | R | 0.41 | 0.74 | 0.86 | -0.04 | (-0.12 - 0.04) | 0.29 | 0.01 | (-0.27 - 0.3) | 0.92 | 0.01 | (-0.2 - 0.21) | 0.95 |
| Pars Orbitalis | L | 0.40 | 0.76 | 0.87 | -0.04 | (-0.11 - 0.04) | 0.33 | 0.04 | (-0.25 - 0.32) | 0.80 | 0.01 | (-0.2 - 0.22) | 0.92 |
| | R | 0.49 | 0.69 | 0.84 | 0.02 | (-0.05 - 0.1) | 0.58 | -0.05 | (-0.34 - 0.23) | 0.71 | -0.09 | (-0.3 - 0.12) | 0.39 |
| Pars Triangularis | L | 0.60 | 0.61 | 0.77 | 0.04 | (-0.04 - 0.12) | 0.30 | 0.10 | (-0.18 - 0.38) | 0.49 | -0.04 | (-0.25 - 0.16) | 0.68 |
| | R | 0.70 | 0.55 | 0.71 | -0.04 | (-0.12 - 0.03) | 0.27 | 0.11 | (-0.18 - 0.4) | 0.46 | -0.04 | (-0.25 - 0.16) | 0.68 |
| Pericalcarine | L | 2.93 | 0.03 | 0.12 | 0.00 | (-0.07 - 0.08) | 0.96 | **0.41** | **(0.12 - 0.7)** | **5.19e-03** | 0.11 | (-0.1 - 0.31) | 0.32 |
| | R | 2.77 | 0.04 | 0.14 | 0.06 | (-0.02 - 0.13) | 0.14 | **0.33** | **(0.04 - 0.62)** | **0.03** | 0.18 | (-0.03 - 0.39) | 0.09 |
| Postcentral | L | 1.57 | 0.19 | 0.36 | -0.07 | (-0.15 - 0.01) | 0.07 | 0.00 | (-0.29 - 0.28) | 0.98 | -0.16 | (-0.37 - 0.05) | 0.13 |
| | R | 3.20 | 0.02 | 0.10 | **-0.11** | **(-0.19 - -0.04)** | **4.13e-03** | -0.20 | (-0.49 - 0.09) | 0.17 | 0.00 | (-0.21 - 0.21) | 1.00 |
| Posterior Cingulate | L | 2.46 | 0.06 | 0.19 | **-0.09** | **(-0.17 - -0.02)** | **0.02** | -0.19 | (-0.47 - 0.1) | 0.20 | 0.03 | (-0.17 - 0.24) | 0.75 |
| | R | 3.42 | 0.02 | 0.08 | **-0.12** | **(-0.2 - -0.05)** | **1.42e-03** | -0.09 | (-0.37 - 0.19) | 0.54 | -0.05 | (-0.25 - 0.16) | 0.65 |
| Precentral | L | 2.06 | 0.10 | 0.26 | **-0.10** | **(-0.17 - -0.02)** | **0.01** | -0.07 | (-0.35 - 0.21) | 0.64 | -0.05 | (-0.26 - 0.15) | 0.62 |
| | R | 3.47 | 0.02 | 0.08 | **-0.12** | **(-0.19 - -0.04)** | **2.68e-03** | -0.01 | (-0.29 - 0.27) | 0.95 | 0.06 | (-0.15 - 0.26) | 0.60 |
| Precuneus | L | 3.77 | 0.01 | 0.06 | **-0.11** | **(-0.18 - -0.03)** | **5.80e-03** | 0.14 | (-0.14 - 0.43) | 0.33 | 0.09 | (-0.12 - 0.3) | 0.41 |
| | R | 2.96 | 0.03 | 0.12 | **-0.12** | **(-0.19 - -0.04)** | **3.06e-03** | -0.05 | (-0.33 - 0.24) | 0.74 | -0.09 | (-0.29 - 0.12) | 0.41 |
| Rostral Anterior | L | 1.77 | 0.15 | 0.31 | **-0.08** | **(-0.16 - -0.01)** | **0.03** | -0.10 | (-0.39 - 0.19) | 0.50 | -0.11 | (-0.32 - 0.09) | 0.28 |

| | | | | | | | | | | | | | |
|---|---|---|---|---|---|---|---|---|---|---|---|---|---|
| Cingulate | R | 1.97 | 0.12 | 0.28 | -0.08 | (-0.15 - 0) | 0.05 | 0.15 | (-0.13 - 0.43) | 0.30 | -0.10 | (-0.31 - 0.1) | 0.33 |
| Rostral Middle Frontal | L | 4.11 | 6.45e-03 | 0.06 | **-0.12** | **(-0.2 - -0.05)** | **1.61e-03** | -0.25 | (-0.54 - 0.04) | 0.09 | 0.02 | (-0.19 - 0.23) | 0.86 |
| | R | 2.69 | 0.04 | 0.15 | **-0.11** | **(-0.18 - -0.03)** | **5.32e-03** | -0.13 | (-0.42 - 0.15) | 0.37 | -0.07 | (-0.28 - 0.14) | 0.51 |
| Superior Frontal | L | 1.59 | 0.19 | 0.36 | **-0.08** | **(-0.16 - -0.01)** | **0.03** | -0.07 | (-0.36 - 0.21) | 0.62 | -0.01 | (-0.21 - 0.2) | 0.96 |
| | R | 1.62 | 0.18 | 0.35 | **-0.08** | **(-0.15 - 0)** | **0.05** | -0.04 | (-0.33 - 0.24) | 0.78 | 0.06 | (-0.15 - 0.26) | 0.59 |
| Superior Parietal | L | 2.12 | 0.10 | 0.25 | **-0.09** | **(-0.17 - -0.01)** | **0.02** | -0.02 | (-0.3 - 0.26) | 0.90 | 0.05 | (-0.15 - 0.26) | 0.61 |
| | R | 4.16 | 5.98e-03 | 0.06 | **-0.12** | **(-0.19 - -0.04)** | **2.78e-03** | -0.05 | (-0.33 - 0.24) | 0.75 | 0.13 | (-0.07 - 0.34) | 0.21 |
| Superior Temporal | L | 2.77 | 0.04 | 0.14 | **-0.11** | **(-0.19 - -0.03)** | **4.85e-03** | -0.10 | (-0.39 - 0.18) | 0.48 | -0.01 | (-0.21 - 0.2) | 0.96 |
| | R | **4.52** | **3.64e-03** | **0.05** | **-0.13** | **(-0.21 - -0.06)** | **7.14e-04** | -0.20 | (-0.49 - 0.09) | 0.17 | -0.19 | (-0.39 - 0.02) | 0.08 |
| Supramarginal | L | 1.53 | 0.21 | 0.38 | **-0.08** | **(-0.15 - 0)** | **0.05** | -0.05 | (-0.33 - 0.23) | 0.73 | -0.13 | (-0.33 - 0.08) | 0.22 |
| | R | 1.84 | 0.14 | 0.30 | **-0.09** | **(-0.16 - -0.01)** | **0.03** | -0.11 | (-0.39 - 0.18) | 0.46 | 0.01 | (-0.19 - 0.22) | 0.89 |
| Temporal Pole | L | 1.05 | 0.37 | 0.54 | -0.07 | (-0.14 - 0.01) | 0.09 | -0.02 | (-0.31 - 0.26) | 0.87 | -0.09 | (-0.3 - 0.11) | 0.37 |
| | R | 1.66 | 0.17 | 0.34 | **-0.09** | **(-0.16 - -0.01)** | **0.03** | -0.05 | (-0.33 - 0.24) | 0.74 | -0.03 | (-0.24 - 0.17) | 0.74 |
| Transverse Temporal | L | 0.45 | 0.72 | 0.85 | -0.04 | (-0.12 - 0.03) | 0.28 | -0.01 | (-0.3 - 0.27) | 0.93 | -0.07 | (-0.27 - 0.14) | 0.52 |
| | R | 2.80 | 0.04 | 0.14 | **-0.08** | **(-0.16 - 0)** | **0.04** | **-0.33** | **(-0.62 - -0.05)** | **0.02** | -0.03 | (-0.24 - 0.18) | 0.80 |
| **Surface Area** | | | | | | | | | | | | | |
| Banks of the Superior Temporal Sulcus | L | 1.27 | 0.28 | 0.47 | -0.07 | (-0.15 - 0.01) | 0.08 | 0.09 | (-0.2 - 0.38) | 0.54 | -0.04 | (-0.25 - 0.17) | 0.68 |
| | R | 2.25 | 0.08 | 0.22 | -0.07 | (-0.14 - 0.01) | 0.09 | 0.19 | (-0.09 - 0.47) | 0.19 | -0.16 | (-0.36 - 0.05) | 0.14 |
| Caudal Anterior Cingulate | L | 1.17 | 0.32 | 0.49 | -0.02 | (-0.1 - 0.06) | 0.60 | 0.24 | (-0.05 - 0.52) | 0.10 | -0.06 | (-0.27 - 0.14) | 0.55 |
| | R | 1.91 | 0.13 | 0.29 | **-0.09** | **(-0.17 - -0.01)** | **0.02** | 0.04 | (-0.25 - 0.32) | 0.80 | -0.02 | (-0.23 - 0.19) | 0.85 |
| Caudal Middle Frontal | L | 0.99 | 0.40 | 0.57 | -0.06 | (-0.13 - 0.02) | 0.16 | 0.10 | (-0.18 - 0.39) | 0.47 | 0.01 | (-0.2 - 0.21) | 0.93 |
| | R | 1.86 | 0.13 | 0.29 | -0.05 | (-0.12 - 0.03) | 0.24 | 0.21 | (-0.08 - 0.49) | 0.15 | 0.11 | (-0.1 - 0.32) | 0.29 |
| Cuneus | L | 2.21 | 0.09 | 0.23 | -0.07 | (-0.14 - 0.01) | 0.09 | 0.24 | (-0.04 - 0.53) | 0.10 | -0.03 | (-0.24 - 0.18) | 0.75 |
| | R | 1.73 | 0.16 | 0.32 | **-0.08** | **(-0.16 - -0.01)** | **0.03** | 0.04 | (-0.24 - 0.33) | 0.78 | -0.10 | (-0.31 - 0.11) | 0.36 |
| Entorhinal | L | 2.64 | 0.05 | 0.16 | **-0.11** | **(-0.18 - -0.03)** | **5.67e-03** | -0.12 | (-0.4 - 0.17) | 0.42 | -0.04 | (-0.24 - 0.17) | 0.74 |
| | R | 1.40 | 0.24 | 0.42 | -0.05 | (-0.12 - 0.03) | 0.25 | -0.05 | (-0.33 - 0.23) | 0.74 | 0.15 | (-0.06 - 0.36) | 0.16 |
| Frontal Pole | L | 0.48 | 0.70 | 0.84 | 0.04 | (-0.04 - 0.11) | 0.34 | 0.04 | (-0.24 - 0.32) | 0.79 | 0.09 | (-0.11 - 0.3) | 0.37 |
| | R | 1.94 | 0.12 | 0.28 | 0.04 | (-0.03 - 0.12) | 0.27 | **0.32** | **(0.03 - 0.6)** | **0.03** | -0.03 | (-0.24 - 0.17) | 0.76 |
| Fusiform | L | 1.82 | 0.14 | 0.30 | -0.04 | (-0.12 - 0.03) | 0.27 | 0.23 | (-0.05 - 0.52) | 0.11 | -0.13 | (-0.33 - 0.08) | 0.23 |

| | | | | | | | | | | | | | |
|---|---|---|---|---|---|---|---|---|---|---|---|---|---|
| | R | 2.68 | 0.05 | 0.15 | **-0.08** | **(-0.16 - -0.01)** | **0.03** | 0.23 | (-0.06 - 0.51) | 0.12 | -0.05 | (-0.25 - 0.16) | 0.65 |
| Inferior Parietal | L | 0.95 | 0.41 | 0.58 | -0.06 | (-0.14 - 0.02) | 0.13 | 0.01 | (-0.27 - 0.29) | 0.96 | -0.10 | (-0.3 - 0.11) | 0.35 |
| | R | 1.96 | 0.12 | 0.28 | **-0.08** | **(-0.16 - -0.01)** | **0.03** | 0.03 | (-0.25 - 0.31) | 0.84 | -0.15 | (-0.35 - 0.06) | 0.16 |
| Inferior Temporal | L | 0.81 | 0.49 | 0.66 | -0.04 | (-0.12 - 0.04) | 0.29 | 0.14 | (-0.14 - 0.43) | 0.32 | -0.03 | (-0.24 - 0.17) | 0.75 |
| | R | 2.42 | 0.06 | 0.19 | -0.02 | (-0.1 - 0.06) | 0.62 | **0.34** | **(0.05 - 0.62)** | **0.02** | -0.11 | (-0.32 - 0.09) | 0.28 |
| Insula | L | 0.98 | 0.40 | 0.57 | -0.06 | (-0.14 - 0.01) | 0.11 | -0.10 | (-0.39 - 0.18) | 0.47 | 0.01 | (-0.2 - 0.21) | 0.95 |
| | R | 0.37 | 0.78 | 0.88 | -0.04 | (-0.12 - 0.04) | 0.31 | -0.03 | (-0.32 - 0.25) | 0.84 | -0.05 | (-0.25 - 0.16) | 0.65 |
| Isthmus Cingulate | L | 0.16 | 0.93 | 0.96 | -0.01 | (-0.09 - 0.06) | 0.72 | 0.06 | (-0.23 - 0.34) | 0.69 | -0.05 | (-0.25 - 0.16) | 0.67 |
| | R | 3.48 | 0.02 | 0.08 | **0.08** | **(0 - 0.15)** | **0.04** | **0.41** | **(0.12 - 0.69)** | **5.47e-03** | 0.05 | (-0.15 - 0.26) | 0.61 |
| Lateral Occipital | L | 3.73 | 0.01 | 0.06 | 0.00 | (-0.08 - 0.07) | 0.93 | **0.40** | **(0.12 - 0.69)** | **5.73e-03** | -0.19 | (-0.39 - 0.02) | 0.08 |
| | R | 0.62 | 0.60 | 0.75 | -0.03 | (-0.1 - 0.05) | 0.47 | 0.15 | (-0.13 - 0.43) | 0.30 | -0.03 | (-0.24 - 0.18) | 0.81 |
| Lateral Orbitofrontal | L | 0.32 | 0.81 | 0.90 | -0.02 | (-0.1 - 0.05) | 0.59 | 0.01 | (-0.27 - 0.3) | 0.93 | 0.07 | (-0.13 - 0.28) | 0.49 |
| | R | 0.34 | 0.80 | 0.89 | -0.03 | (-0.11 - 0.04) | 0.39 | 0.04 | (-0.25 - 0.32) | 0.79 | 0.02 | (-0.18 - 0.23) | 0.83 |
| Lingual | L | 1.12 | 0.34 | 0.51 | -0.05 | (-0.13 - 0.02) | 0.16 | -0.13 | (-0.41 - 0.16) | 0.38 | -0.13 | (-0.34 - 0.08) | 0.22 |
| | R | 0.53 | 0.66 | 0.81 | -0.02 | (-0.09 - 0.06) | 0.66 | 0.16 | (-0.13 - 0.45) | 0.27 | -0.02 | (-0.23 - 0.18) | 0.82 |
| Medial Orbitofrontal | L | 0.13 | 0.94 | 0.97 | -0.02 | (-0.09 - 0.06) | 0.66 | 0.05 | (-0.24 - 0.33) | 0.75 | -0.03 | (-0.24 - 0.18) | 0.78 |
| | R | 1.57 | 0.19 | 0.36 | **-0.08** | **(-0.15 - 0)** | **0.04** | -0.02 | (-0.3 - 0.27) | 0.91 | -0.12 | (-0.33 - 0.09) | 0.26 |
| Middle Temporal | L | 0.73 | 0.54 | 0.70 | 0.00 | (-0.07 - 0.08) | 0.95 | 0.21 | (-0.08 - 0.49) | 0.15 | 0.04 | (-0.17 - 0.25) | 0.71 |
| | R | 1.17 | 0.32 | 0.49 | -0.07 | (-0.14 - 0.01) | 0.09 | 0.02 | (-0.26 - 0.3) | 0.90 | -0.11 | (-0.31 - 0.1) | 0.30 |
| Paracentral | L | 0.17 | 0.92 | 0.96 | -0.03 | (-0.1 - 0.05) | 0.50 | 0.01 | (-0.27 - 0.29) | 0.94 | 0.00 | (-0.21 - 0.2) | 0.97 |
| | R | 0.84 | 0.47 | 0.65 | -0.01 | (-0.08 - 0.07) | 0.88 | 0.00 | (-0.29 - 0.28) | 0.99 | 0.16 | (-0.05 - 0.37) | 0.13 |
| Parahippocampal | L | 1.52 | 0.21 | 0.38 | -0.04 | (-0.11 - 0.04) | 0.37 | -0.27 | (-0.56 - 0.01) | 0.06 | -0.10 | (-0.3 - 0.11) | 0.35 |
| | R | 0.75 | 0.52 | 0.70 | 0.06 | (-0.02 - 0.13) | 0.15 | 0.05 | (-0.24 - 0.33) | 0.75 | 0.08 | (-0.13 - 0.28) | 0.47 |
| Pars Opercularis | L | 2.11 | 0.10 | 0.25 | **-0.09** | **(-0.17 - -0.01)** | **0.02** | -0.14 | (-0.42 - 0.15) | 0.35 | 0.03 | (-0.18 - 0.23) | 0.79 |
| | R | 0.59 | 0.62 | 0.77 | -0.03 | (-0.1 - 0.05) | 0.49 | 0.07 | (-0.22 - 0.35) | 0.64 | 0.09 | (-0.12 - 0.3) | 0.40 |
| Pars Orbitalis | L | 1.12 | 0.34 | 0.51 | 0.01 | (-0.07 - 0.09) | 0.78 | 0.19 | (-0.09 - 0.47) | 0.19 | 0.14 | (-0.06 - 0.35) | 0.18 |
| | R | 0.69 | 0.56 | 0.71 | 0.03 | (-0.05 - 0.11) | 0.45 | 0.17 | (-0.11 - 0.46) | 0.23 | 0.07 | (-0.14 - 0.28) | 0.51 |
| Pars Triangularis | L | 1.25 | 0.29 | 0.47 | -0.07 | (-0.14 - 0.01) | 0.08 | 0.01 | (-0.27 - 0.29) | 0.95 | 0.05 | (-0.16 - 0.25) | 0.65 |
| | R | 0.18 | 0.91 | 0.96 | 0.01 | (-0.06 - | 0.76 | 0.10 | (-0.18 - | 0.49 | 0.00 | (-0.2 - 0.21) | 0.99 |

| | | | | | | | | | | | | | |
|---|---|---|---|---|---|---|---|---|---|---|---|---|---|
| | | | | | | 0.09) | | | 0.38) | | | | |
| Pericalcarine | L | 1.05 | 0.37 | 0.54 | -0.05 | (-0.13 - 0.02) | 0.16 | 0.13 | (-0.16 - 0.42) | 0.38 | -0.01 | (-0.21 - 0.2) | 0.95 |
| | R | 2.27 | 0.08 | 0.22 | **-0.10** | **(-0.18 - -0.02)** | **9.71e-03** | -0.07 | (-0.36 - 0.22) | 0.65 | -0.08 | (-0.29 - 0.13) | 0.43 |
| Postcentral | L | 0.10 | 0.96 | 0.97 | 0.01 | (-0.07 - 0.08) | 0.85 | -0.02 | (-0.3 - 0.26) | 0.88 | -0.05 | (-0.25 - 0.16) | 0.67 |
| | R | 1.24 | 0.30 | 0.48 | -0.04 | (-0.12 - 0.04) | 0.29 | -0.24 | (-0.53 - 0.04) | 0.10 | 0.02 | (-0.18 - 0.23) | 0.83 |
| Posterior Cingulate | L | 1.39 | 0.24 | 0.42 | -0.05 | (-0.12 - 0.03) | 0.22 | 0.14 | (-0.14 - 0.43) | 0.32 | -0.14 | (-0.35 - 0.07) | 0.19 |
| | R | 1.76 | 0.15 | 0.31 | -0.06 | (-0.14 - 0.01) | 0.10 | -0.01 | (-0.3 - 0.27) | 0.93 | -0.20 | (-0.4 - 0.01) | 0.06 |
| Precentral | L | 1.20 | 0.31 | 0.49 | 0.04 | (-0.03 - 0.12) | 0.29 | 0.25 | (-0.04 - 0.53) | 0.09 | 0.03 | (-0.18 - 0.24) | 0.77 |
| | R | 0.23 | 0.87 | 0.94 | 0.01 | (-0.07 - 0.09) | 0.77 | 0.12 | (-0.17 - 0.4) | 0.43 | 0.03 | (-0.18 - 0.23) | 0.80 |
| Precuneus | L | 1.05 | 0.37 | 0.54 | -0.04 | (-0.11 - 0.04) | 0.34 | -0.12 | (-0.4 - 0.16) | 0.41 | -0.16 | (-0.36 - 0.05) | 0.13 |
| | R | 0.44 | 0.73 | 0.85 | -0.04 | (-0.11 - 0.04) | 0.34 | -0.11 | (-0.39 - 0.18) | 0.46 | -0.04 | (-0.25 - 0.17) | 0.71 |
| Rostral Anterior Cingulate | L | 3.78 | 0.01 | 0.06 | **-0.12** | **(-0.2 - -0.05)** | **1.75e-03** | 0.08 | (-0.2 - 0.36) | 0.59 | 0.02 | (-0.19 - 0.22) | 0.87 |
| | R | 2.16 | 0.09 | 0.24 | **-0.09** | **(-0.16 - -0.01)** | **0.02** | 0.08 | (-0.21 - 0.36) | 0.59 | 0.04 | (-0.17 - 0.25) | 0.71 |
| Rostral Middle Frontal | L | 2.43 | 0.06 | 0.19 | -0.02 | (-0.1 - 0.06) | 0.62 | **0.34** | **(0.05 - 0.62)** | **0.02** | -0.12 | (-0.32 - 0.09) | 0.27 |
| | R | **4.48** | **3.81e-03** | **0.05** | -0.02 | (-0.1 - 0.05) | 0.58 | **0.41** | **(0.13 - 0.69)** | **4.73e-03** | **-0.22** | **(-0.43 - -0.02)** | **0.03** |
| Superior Frontal | L | 3.24 | 0.02 | 0.10 | -0.06 | (-0.14 - 0.02) | 0.13 | 0.28 | (0 - 0.56) | 0.05 | 0.14 | (-0.06 - 0.35) | 0.17 |
| | R | 3.65 | 0.01 | 0.06 | **-0.12** | **(-0.19 - -0.04)** | **2.39e-03** | 0.10 | (-0.18 - 0.39) | 0.47 | 0.01 | (-0.2 - 0.21) | 0.95 |
| Superior Parietal | L | 0.12 | 0.95 | 0.97 | 0.01 | (-0.07 - 0.08) | 0.83 | -0.06 | (-0.34 - 0.22) | 0.69 | 0.04 | (-0.17 - 0.25) | 0.71 |
| | R | 0.63 | 0.59 | 0.75 | 0.02 | (-0.06 - 0.09) | 0.66 | 0.17 | (-0.11 - 0.45) | 0.24 | -0.06 | (-0.27 - 0.15) | 0.58 |
| Superior Temporal | L | 0.15 | 0.93 | 0.96 | -0.03 | (-0.1 - 0.05) | 0.51 | -0.02 | (-0.31 - 0.27) | 0.90 | -0.01 | (-0.22 - 0.19) | 0.89 |
| | R | 0.91 | 0.44 | 0.60 | -0.06 | (-0.13 - 0.02) | 0.14 | 0.02 | (-0.26 - 0.3) | 0.89 | -0.10 | (-0.31 - 0.11) | 0.35 |
| Supramarginal | L | 2.82 | 0.04 | 0.14 | **-0.10** | **(-0.18 - -0.02)** | **0.01** | -0.21 | (-0.5 - 0.08) | 0.16 | -0.14 | (-0.35 - 0.06) | 0.18 |
| | R | 1.47 | 0.22 | 0.40 | -0.07 | (-0.14 - 0.01) | 0.09 | -0.11 | (-0.39 - 0.17) | 0.45 | -0.15 | (-0.36 - 0.06) | 0.15 |
| Temporal Pole | L | 0.06 | 0.98 | 0.98 | -0.01 | (-0.08 - 0.07) | 0.88 | -0.06 | (-0.34 - 0.23) | 0.70 | 0.01 | (-0.2 - 0.21) | 0.96 |
| | R | 3.25 | 0.02 | 0.10 | 0.03 | (-0.05 - 0.11) | 0.42 | 0.24 | (-0.04 - 0.52) | 0.10 | **0.29** | **(0.08 - 0.5)** | **6.33e-03** |
| Transverse Temporal | L | 1.16 | 0.32 | 0.49 | -0.05 | (-0.13 - 0.03) | 0.20 | -0.18 | (-0.46 - 0.11) | 0.23 | 0.06 | (-0.14 - 0.27) | 0.56 |
| | R | 0.96 | 0.41 | 0.58 | -0.01 | (-0.09 - 0.06) | 0.71 | 0.02 | (-0.27 - 0.3) | 0.91 | -0.18 | (-0.38 - 0.03) | 0.09 |
| **Subcortical Volume** | | | | | | | | | | | | | |
| Amygdala | L | 0.73 | 0.54 | 0.70 | -0.04 | (-0.12 - 0.03) | 0.26 | 0.08 | (-0.2 - 0.37) | 0.56 | 0.04 | (-0.17 - 0.25) | 0.71 |

| | | | | | | | | | | | | | |
|---|---|---|---|---|---|---|---|---|---|---|---|---|---|
| | R | 3.05 | 0.03 | 0.12 | **-0.10** | **(-0.17 - -0.02)** | **0.01** | 0.19 | (-0.09 - 0.47) | 0.19 | 0.00 | (-0.21 - 0.2) | 0.98 |
| Caudate | L | 0.47 | 0.71 | 0.84 | 0.02 | (-0.06 - 0.1) | 0.60 | -0.14 | (-0.43 - 0.15) | 0.35 | -0.02 | (-0.23 - 0.19) | 0.84 |
| | R | 0.25 | 0.86 | 0.93 | -0.02 | (-0.1 - 0.06) | 0.62 | -0.11 | (-0.39 - 0.18) | 0.45 | -0.03 | (-0.24 - 0.18) | 0.78 |
| Hippocampus | L | 1.25 | 0.29 | 0.47 | -0.07 | (-0.15 - 0) | 0.07 | -0.07 | (-0.35 - 0.22) | 0.65 | 0.02 | (-0.19 - 0.23) | 0.85 |
| | R | **5.59** | **8.01e-04** | **0.02** | **-0.16** | **(-0.23 - -0.08)** | **6.45e-05** | -0.20 | (-0.48 - 0.08) | 0.17 | -0.06 | (-0.26 - 0.15) | 0.59 |
| Lateral Ventricles | L | 4.09 | 6.54e-03 | 0.06 | **0.13** | **(0.05 - 0.2)** | **1.12e-03** | -0.02 | (-0.31 - 0.27) | 0.88 | -0.05 | (-0.26 - 0.15) | 0.62 |
| | R | 3.79 | 9.99e-03 | 0.06 | **0.11** | **(0.04 - 0.19)** | **3.65e-03** | -0.18 | (-0.47 - 0.11) | 0.22 | 0.11 | (-0.1 - 0.32) | 0.30 |
| Nucleus Accumbens | L | 0.28 | 0.84 | 0.91 | -0.01 | (-0.09 - 0.06) | 0.76 | -0.10 | (-0.38 - 0.19) | 0.51 | -0.07 | (-0.28 - 0.14) | 0.50 |
| | R | 0.05 | 0.98 | 0.98 | 0.01 | (-0.07 - 0.09) | 0.81 | -0.03 | (-0.32 - 0.25) | 0.82 | -0.01 | (-0.22 - 0.2) | 0.92 |
| Pallidum | L | 0.35 | 0.79 | 0.89 | -0.02 | (-0.1 - 0.05) | 0.55 | 0.02 | (-0.26 - 0.3) | 0.88 | 0.07 | (-0.13 - 0.28) | 0.49 |
| | R | 1.06 | 0.36 | 0.54 | -0.03 | (-0.1 - 0.05) | 0.52 | -0.09 | (-0.37 - 0.19) | 0.53 | 0.15 | (-0.06 - 0.36) | 0.16 |
| Putamen | L | 0.16 | 0.93 | 0.96 | -0.01 | (-0.08 - 0.07) | 0.86 | 0.04 | (-0.24 - 0.33) | 0.78 | -0.06 | (-0.27 - 0.14) | 0.55 |
| | R | 0.31 | 0.82 | 0.91 | -0.02 | (-0.09 - 0.06) | 0.67 | 0.10 | (-0.19 - 0.4) | 0.48 | 0.03 | (-0.17 - 0.24) | 0.74 |
| Thalamus | L | 1.30 | 0.27 | 0.46 | -0.05 | (-0.13 - 0.03) | 0.20 | 0.04 | (-0.25 - 0.32) | 0.80 | 0.12 | (-0.08 - 0.33) | 0.24 |
| | R | 2.45 | 0.06 | 0.19 | **-0.09** | **(-0.17 - -0.02)** | **0.02** | -0.09 | (-0.37 - 0.19) | 0.53 | 0.08 | (-0.13 - 0.28) | 0.47 |
| **Global** | | | | | | | | | | | | | |
| Estimated Intracranial Volume | N/A | 4.28 | 5.03e-03 | 0.06 | **-0.13** | **(-0.21 - -0.06)** | **5.20e-04** | -0.15 | (-0.43 - 0.13) | 0.29 | -0.14 | (-0.35 - 0.07) | 0.18 |
| Mean Cortical Thickness | | **5.87** | **5.42e-04** | **0.02** | **-0.16** | **(-0.24 - -0.09)** | **3.06e-05** | -0.12 | (-0.4 - 0.17) | 0.43 | -0.05 | (-0.26 - 0.16) | 0.64 |
| Total Surface Area | | **6.29** | **3.01e-04** | **0.02** | **-0.16** | **(-0.24 - -0.09)** | **3.37e-05** | 0.03 | (-0.25 - 0.31) | 0.84 | -0.19 | (-0.39 - 0.02) | 0.08 |

*Results marked in bold are significant at q<0.05 (the overall effect) or at p<0.05 (pairwise comparisons). Pairwise comparisons shown in this table are APS/BIPS/GRD < HC. CHR subgroup comparisons are reported in part B.*

**eTable 12B.** CHR subgroup (APS vs. BIPS vs. GRD vs. HC) effect size overview (continued)

| | | Overall Test Statistics | | | APS < BIPS | | | APS < GRD | | | GRD < BIPS | | |
|---|---|---|---|---|---|---|---|---|---|---|---|---|---|
| Region of Interest | Hemisphere | F | p | q | Cohen's d | 95% Confidence Interval | p | Cohen's d | 95% Confidence Interval | p | Cohen's d | 95% Confidence Interval | p |
| **Cortical Thickness** | | | | | | | | | | | | | |
| Banks of the Superior Temporal Sulcus | L | 2.41 | 0.07 | 0.19 | -0.14 | (0.15 - -0.43) | 0.33 | -0.16 | (0.05 - -0.37) | 0.14 | 0.02 | (-0.33 - 0.37) | 0.93 |
| | R | 4.18 | 5.84e-03 | 0.06 | -0.19 | (0.09 - -0.48) | 0.18 | -0.04 | (0.17 - -0.25) | 0.73 | -0.16 | (-0.51 - 0.18) | 0.37 |
| Caudal Anterior Cingulate | L | 1.81 | 0.14 | 0.30 | 0.20 | (0.48 - -0.09) | 0.18 | -0.06 | (0.15 - -0.27) | 0.58 | 0.26 | (-0.08 - 0.61) | 0.15 |
| | R | 3.94 | 8.08e-03 | 0.06 | 0.14 | (0.42 - -0.15) | 0.35 | **-0.29** | **(-0.08 - -0.5)** | **6.30e-03** | **0.44** | **(0.09 - 0.78)** | **0.01** |
| Caudal Middle Frontal | L | 2.36 | 0.07 | 0.20 | 0.13 | (0.42 - -0.16) | 0.39 | -0.11 | (0.09 - -0.32) | 0.28 | 0.25 | (-0.1 - 0.6) | 0.17 |
| | R | 3.98 | 7.73e-03 | 0.06 | -0.18 | (0.11 - -0.46) | 0.22 | -0.21 | (0 - -0.41) | 0.05 | 0.03 | (-0.32 - 0.37) | 0.88 |
| Cuneus | L | 1.44 | 0.23 | 0.41 | -0.28 | (0.01 - -0.57) | 0.06 | -0.09 | (0.12 - -0.3) | 0.40 | -0.22 | (-0.56 - 0.13) | 0.28 |
| | R | 2.55 | 0.05 | 0.17 | **-0.38** | **(-0.1 - -0.67)** | **7.95e-03** | -0.02 | (0.19 - -0.23) | 0.84 | **-0.43** | **(-0.78 - -0.09)** | **0.04** |
| Entorhinal | L | 0.29 | 0.83 | 0.91 | 0.10 | (0.39 - -0.18) | 0.48 | 0.07 | (0.27 - -0.14) | 0.52 | 0.04 | (-0.3 - 0.38) | 0.84 |
| | R | 0.75 | 0.52 | 0.70 | 0.12 | (0.41 - -0.16) | 0.40 | 0.09 | (0.3 - -0.12) | 0.40 | 0.03 | (-0.31 - 0.38) | 0.85 |
| Frontal Pole | L | 1.21 | 0.30 | 0.49 | -0.22 | (0.06 - -0.5) | 0.13 | -0.06 | (0.15 - -0.27) | 0.58 | -0.16 | (-0.51 - 0.18) | 0.36 |
| | R | 1.47 | 0.22 | 0.40 | 0.12 | (0.41 - -0.17) | 0.41 | -0.10 | (0.11 - -0.31) | 0.36 | 0.23 | (-0.12 - 0.57) | 0.22 |
| Fusiform | L | **5.81** | **5.92e-04** | **0.02** | -0.08 | (0.2 - -0.36) | 0.58 | -0.17 | (0.04 - -0.37) | 0.12 | 0.09 | (-0.25 - 0.43) | 0.62 |
| | R | 1.86 | 0.13 | 0.29 | -0.10 | (0.19 - -0.38) | 0.51 | -0.02 | (0.19 - -0.22) | 0.87 | -0.08 | (-0.42 - 0.26) | 0.65 |
| Inferior Parietal | L | **6.35** | **2.75e-04** | **0.02** | 0.06 | (0.35 - -0.22) | 0.68 | **-0.21** | **(-0.01 - -0.42)** | **0.04** | 0.29 | (-0.06 - 0.63) | 0.12 |
| | R | 3.93 | 8.24e-03 | 0.06 | -0.05 | (0.23 - -0.34) | 0.73 | **-0.23** | **(-0.03 - -0.44)** | **0.03** | 0.19 | (-0.16 - 0.53) | 0.30 |
| Inferior Temporal | L | 2.94 | 0.03 | 0.12 | 0.00 | (0.28 - -0.29) | 0.97 | 0.08 | (0.29 - -0.13) | 0.45 | -0.09 | (-0.43 - 0.25) | 0.63 |
| | R | 3.83 | 9.51e-03 | 0.06 | 0.03 | (0.32 - -0.25) | 0.82 | 0.00 | (0.21 - -0.21) | 1.00 | 0.03 | (-0.31 - 0.38) | 0.86 |
| Insula | L | **5.56** | **8.41e-04** | **0.02** | -0.09 | (0.19 - -0.38) | 0.52 | 0.06 | (0.27 - -0.14) | 0.56 | -0.16 | (-0.5 - 0.18) | 0.37 |
| | R | **4.80** | **2.46e-03** | **0.04** | 0.11 | (0.4 - -0.17) | 0.43 | -0.03 | (0.17 - -0.24) | 0.75 | 0.15 | (-0.19 - 0.49) | 0.40 |
| Isthmus Cingulate | L | 0.58 | 0.63 | 0.77 | 0.18 | (0.46 - -0.11) | 0.23 | -0.05 | (0.16 - -0.25) | 0.66 | 0.23 | (-0.11 - 0.58) | 0.21 |
| | R | 0.46 | 0.71 | 0.84 | -0.01 | (0.27 - -0.3) | 0.94 | -0.03 | (0.18 - -0.24) | 0.77 | 0.02 | (-0.32 - 0.37) | 0.91 |
| Lateral Occipital | L | 3.72 | 0.01 | 0.06 | 0.01 | (0.29 - -0.28) | 0.97 | -0.20 | (0.01 - -0.4) | 0.07 | 0.21 | (-0.13 - 0.55) | 0.25 |
| | R | **4.58** | **3.31e-03** | **0.05** | 0.06 | (0.35 - -0.22) | 0.66 | -0.07 | (0.14 - -0.28) | 0.52 | 0.14 | (-0.21 - 0.48) | 0.46 |
| | | | | | | (0.26 - | | | (0.05 - | | | (-0.22 - | |

| | | | | | | | | | | | | | |
|---|---|---|---|---|---|---|---|---|---|---|---|---|---|
| Lateral Orbitofrontal | L | 2.03 | 0.11 | 0.27 | -0.03 | -0.31) | 0.84 | -0.16 | -0.36) | 0.14 | 0.13 | 0.47) | 0.47 |
| | R | 1.71 | 0.16 | 0.32 | -0.09 | (0.2 - -0.37) | 0.56 | -0.12 | (0.08 - -0.33) | 0.24 | 0.04 | (-0.31 - 0.39) | 0.83 |
| Lingual | L | 1.87 | 0.13 | 0.29 | **-0.31** | **(-0.03 - -0.6)** | **0.03** | -0.11 | (0.1 - -0.32) | 0.32 | -0.22 | (-0.57 - 0.12) | 0.25 |
| | R | 0.23 | 0.88 | 0.94 | -0.09 | (0.19 - -0.38) | 0.52 | -0.05 | (0.16 - -0.26) | 0.62 | -0.05 | (-0.39 - 0.3) | 0.82 |
| Medial Orbitofrontal | L | **4.57** | **3.38e-03** | **0.05** | -0.12 | (0.17 - -0.4) | 0.43 | 0.02 | (0.22 - -0.19) | 0.88 | -0.14 | (-0.48 - 0.2) | 0.46 |
| | R | 2.61 | 0.05 | 0.16 | -0.08 | (0.21 - -0.36) | 0.59 | -0.12 | (0.08 - -0.33) | 0.24 | 0.05 | (-0.3 - 0.39) | 0.80 |
| Middle Temporal | L | 1.95 | 0.12 | 0.28 | 0.00 | (0.29 - -0.29) | 1.00 | -0.05 | (0.15 - -0.26) | 0.62 | 0.06 | (-0.29 - 0.4) | 0.77 |
| | R | **7.38** | **6.35e-05** | **9.84e-03** | 0.03 | (0.31 - -0.26) | 0.85 | -0.14 | (0.07 - -0.35) | 0.20 | 0.17 | (-0.17 - 0.52) | 0.35 |
| Paracentral | L | **5.17** | **1.45e-03** | **0.03** | -0.23 | (0.06 - -0.51) | 0.12 | -0.15 | (0.05 - -0.36) | 0.14 | -0.08 | (-0.42 - 0.27) | 0.68 |
| | R | 3.07 | 0.03 | 0.12 | **-0.31** | **(-0.02 - -0.59)** | **0.03** | -0.12 | (0.09 - -0.32) | 0.27 | -0.20 | (-0.54 - 0.14) | 0.28 |
| Parahippocampal | L | 3.82 | 9.55e-03 | 0.06 | -0.28 | (0.01 - -0.56) | 0.06 | -0.01 | (0.2 - -0.22) | 0.92 | -0.27 | (-0.61 - 0.07) | 0.13 |
| | R | 1.31 | 0.27 | 0.46 | -0.19 | (0.09 - -0.47) | 0.19 | 0.02 | (0.23 - -0.18) | 0.84 | -0.22 | (-0.56 - 0.13) | 0.23 |
| Pars Opercularis | L | 0.70 | 0.55 | 0.71 | -0.18 | (0.11 - -0.47) | 0.22 | -0.05 | (0.16 - -0.25) | 0.66 | -0.14 | (-0.48 - 0.21) | 0.46 |
| | R | 0.41 | 0.74 | 0.86 | -0.06 | (0.23 - -0.34) | 0.70 | -0.05 | (0.16 - -0.25) | 0.65 | -0.01 | (-0.35 - 0.33) | 0.96 |
| Pars Orbitalis | L | 0.40 | 0.76 | 0.87 | -0.08 | (0.21 - -0.36) | 0.60 | -0.05 | (0.16 - -0.26) | 0.64 | -0.03 | (-0.37 - 0.31) | 0.88 |
| | R | 0.49 | 0.69 | 0.84 | 0.08 | (0.36 - -0.21) | 0.60 | 0.12 | (0.33 - -0.1) | 0.28 | -0.04 | (-0.39 - 0.31) | 0.82 |
| Pars Triangularis | L | 0.60 | 0.61 | 0.77 | -0.06 | (0.22 - -0.34) | 0.68 | 0.08 | (0.29 - -0.12) | 0.42 | -0.15 | (-0.49 - 0.19) | 0.41 |
| | R | 0.70 | 0.55 | 0.71 | -0.15 | (0.13 - -0.44) | 0.30 | 0.00 | (0.21 - -0.21) | 1.00 | -0.16 | (-0.51 - 0.19) | 0.39 |
| Pericalcarine | L | 2.93 | 0.03 | 0.12 | **-0.41** | **(-0.12 - -0.7)** | **5.39e-03** | -0.10 | (0.1 - -0.31) | 0.33 | -0.35 | (-0.69 - 0) | 0.08 |
| | R | 2.77 | 0.04 | 0.14 | -0.27 | (0.02 - -0.56) | 0.07 | -0.12 | (0.09 - -0.33) | 0.25 | -0.17 | (-0.52 - 0.18) | 0.41 |
| Postcentral | L | 1.57 | 0.19 | 0.36 | -0.07 | (0.22 - -0.36) | 0.64 | 0.09 | (0.3 - -0.12) | 0.40 | -0.17 | (-0.52 - 0.17) | 0.37 |
| | R | 3.20 | 0.02 | 0.10 | 0.09 | (0.38 - -0.2) | 0.55 | -0.12 | (0.09 - -0.32) | 0.27 | 0.23 | (-0.12 - 0.58) | 0.25 |
| Posterior Cingulate | L | 2.46 | 0.06 | 0.19 | 0.09 | (0.38 - -0.2) | 0.54 | -0.13 | (0.08 - -0.34) | 0.22 | 0.23 | (-0.12 - 0.57) | 0.21 |
| | R | 3.42 | 0.02 | 0.08 | -0.04 | (0.24 - -0.32) | 0.79 | -0.08 | (0.13 - -0.29) | 0.46 | 0.04 | (-0.3 - 0.39) | 0.81 |
| Precentral | L | 2.06 | 0.10 | 0.26 | -0.03 | (0.25 - -0.31) | 0.83 | -0.05 | (0.16 - -0.25) | 0.66 | 0.02 | (-0.32 - 0.36) | 0.93 |
| | R | 3.47 | 0.02 | 0.08 | -0.11 | (0.17 - -0.39) | 0.44 | -0.18 | (0.03 - -0.38) | 0.10 | 0.07 | (-0.27 - 0.41) | 0.71 |
| Precuneus | L | 3.77 | 0.01 | 0.06 | -0.25 | (0.03 - -0.54) | 0.09 | -0.20 | (0.01 - -0.41) | 0.06 | -0.06 | (-0.4 - 0.29) | 0.76 |
| | R | 2.96 | 0.03 | 0.12 | -0.07 | (0.22 - -0.36) | 0.63 | -0.03 | (0.18 - -0.24) | 0.78 | -0.04 | (-0.39 - 0.3) | 0.82 |
| Rostral Anterior | L | 1.77 | 0.15 | 0.31 | 0.01 | (0.3 - -0.27) | 0.92 | 0.03 | (0.23 - -0.18) | 0.79 | -0.01 | (-0.36 - 0.33) | 0.94 |

| | | | | | | | | | | | | | |
|---|---|---|---|---|---|---|---|---|---|---|---|---|---|
| Cingulate | R | 1.97 | 0.12 | 0.28 | -0.23 | (0.06 - -0.51) | 0.12 | 0.03 | (0.23 - -0.18) | 0.81 | -0.27 | (-0.61 - 0.07) | 0.15 |
| Rostral Middle Frontal | L | 4.11 | 6.45e-03 | 0.06 | 0.12 | (0.41 - -0.16) | 0.40 | -0.14 | (0.06 - -0.35) | 0.17 | 0.27 | (-0.07 - 0.62) | 0.13 |
| | R | 2.69 | 0.04 | 0.15 | 0.02 | (0.31 - -0.27) | 0.89 | -0.04 | (0.17 - -0.25) | 0.70 | 0.06 | (-0.28 - 0.41) | 0.73 |
| Superior Frontal | L | 1.59 | 0.19 | 0.36 | -0.01 | (0.27 - -0.3) | 0.93 | -0.08 | (0.13 - -0.29) | 0.45 | 0.07 | (-0.27 - 0.41) | 0.70 |
| | R | 1.62 | 0.18 | 0.35 | -0.04 | (0.25 - -0.32) | 0.79 | -0.14 | (0.07 - -0.34) | 0.20 | 0.10 | (-0.24 - 0.44) | 0.58 |
| Superior Parietal | L | 2.12 | 0.10 | 0.25 | -0.07 | (0.21 - -0.36) | 0.61 | -0.15 | (0.06 - -0.35) | 0.17 | 0.07 | (-0.27 - 0.42) | 0.69 |
| | R | 4.16 | 5.98e-03 | 0.06 | -0.07 | (0.21 - -0.36) | 0.61 | **-0.25** | **(-0.05 - -0.46)** | **0.02** | 0.19 | (-0.16 - 0.53) | 0.31 |
| Superior Temporal | L | 2.77 | 0.04 | 0.14 | -0.01 | (0.28 - -0.3) | 0.96 | -0.11 | (0.1 - -0.32) | 0.32 | 0.10 | (-0.24 - 0.45) | 0.58 |
| | R | **4.52** | **3.64e-03** | **0.05** | 0.07 | (0.36 - -0.22) | 0.65 | 0.05 | (0.26 - -0.15) | 0.62 | 0.02 | (-0.33 - 0.36) | 0.93 |
| Supramarginal | L | 1.53 | 0.21 | 0.38 | -0.03 | (0.25 - -0.31) | 0.85 | 0.05 | (0.26 - -0.16) | 0.63 | -0.09 | (-0.43 - 0.25) | 0.65 |
| | R | 1.84 | 0.14 | 0.30 | 0.02 | (0.3 - -0.27) | 0.90 | -0.10 | (0.1 - -0.31) | 0.33 | 0.13 | (-0.21 - 0.48) | 0.48 |
| Temporal Pole | L | 1.05 | 0.37 | 0.54 | -0.04 | (0.24 - -0.33) | 0.76 | 0.03 | (0.24 - -0.18) | 0.79 | -0.08 | (-0.42 - 0.26) | 0.68 |
| | R | 1.66 | 0.17 | 0.34 | -0.04 | (0.24 - -0.32) | 0.77 | -0.05 | (0.15 - -0.26) | 0.61 | 0.01 | (-0.33 - 0.36) | 0.94 |
| Transverse Temporal | L | 0.45 | 0.72 | 0.85 | -0.03 | (0.26 - -0.31) | 0.84 | 0.02 | (0.23 - -0.18) | 0.81 | -0.06 | (-0.4 - 0.29) | 0.76 |
| | R | 2.80 | 0.04 | 0.14 | 0.25 | (0.54 - -0.03) | 0.08 | -0.05 | (0.15 - -0.26) | 0.61 | 0.31 | (-0.03 - 0.66) | 0.08 |
| **Surface Area** | | | | | | | | | | | | | |
| Banks of the Superior Temporal Sulcus | L | 1.27 | 0.28 | 0.47 | -0.16 | (0.13 - -0.45) | 0.28 | -0.03 | (0.18 - -0.24) | 0.81 | -0.15 | (-0.5 - 0.2) | 0.45 |
| | R | 2.25 | 0.08 | 0.22 | -0.26 | (0.03 - -0.54) | 0.08 | 0.09 | (0.3 - -0.12) | 0.39 | **-0.36** | **(-0.71 - -0.02)** | **0.05** |
| Caudal Anterior Cingulate | L | 1.17 | 0.32 | 0.49 | -0.26 | (0.03 - -0.54) | 0.08 | 0.04 | (0.25 - -0.16) | 0.68 | -0.31 | (-0.66 - 0.04) | 0.09 |
| | R | 1.91 | 0.13 | 0.29 | -0.13 | (0.16 - -0.41) | 0.38 | -0.07 | (0.13 - -0.28) | 0.50 | -0.06 | (-0.4 - 0.28) | 0.74 |
| Caudal Middle Frontal | L | 0.99 | 0.40 | 0.57 | -0.16 | (0.12 - -0.45) | 0.27 | -0.07 | (0.14 - -0.27) | 0.54 | -0.10 | (-0.44 - 0.25) | 0.59 |
| | R | 1.86 | 0.13 | 0.29 | -0.25 | (0.03 - -0.54) | 0.08 | -0.16 | (0.05 - -0.37) | 0.13 | -0.10 | (-0.44 - 0.24) | 0.59 |
| Cuneus | L | 2.21 | 0.09 | 0.23 | **-0.31** | **(-0.03 - -0.6)** | **0.03** | -0.03 | (0.18 - -0.24) | 0.75 | -0.31 | (-0.66 - 0.04) | 0.12 |
| | R | 1.73 | 0.16 | 0.32 | -0.13 | (0.16 - -0.41) | 0.39 | 0.01 | (0.22 - -0.2) | 0.90 | -0.16 | (-0.51 - 0.18) | 0.43 |
| Entorhinal | L | 2.64 | 0.05 | 0.16 | 0.01 | (0.29 - -0.28) | 0.95 | -0.07 | (0.13 - -0.28) | 0.48 | 0.09 | (-0.26 - 0.43) | 0.64 |
| | R | 1.40 | 0.24 | 0.42 | 0.00 | (0.29 - -0.28) | 0.98 | -0.20 | (0.01 - -0.4) | 0.06 | 0.21 | (-0.13 - 0.55) | 0.25 |
| Frontal Pole | L | 0.48 | 0.70 | 0.84 | 0.00 | (0.28 - -0.28) | 0.99 | -0.06 | (0.15 - -0.26) | 0.59 | 0.06 | (-0.29 - 0.4) | 0.75 |
| | R | 1.94 | 0.12 | 0.28 | -0.27 | (0.01 - -0.56) | 0.06 | 0.08 | (0.28 - -0.13) | 0.48 | **-0.36** | **(-0.7 - -0.02)** | **0.05** |
| Fusiform | L | 1.82 | 0.14 | 0.30 | -0.28 | (0.01 - -0.56) | 0.06 | 0.08 | (0.29 - -0.12) | 0.43 | **-0.37** | **(-0.71 - -0.02)** | **0.04** |

| | | | | | | | | | | | | | |
|---|---|---|---|---|---|---|---|---|---|---|---|---|---|
| | R | 2.68 | 0.05 | 0.15 | **-0.31** | **(-0.03 - -0.59)** | **0.03** | -0.04 | (0.17 - -0.24) | 0.73 | -0.28 | (-0.62 - 0.06) | 0.12 |
| Inferior Parietal | L | 0.95 | 0.41 | 0.58 | -0.07 | (0.21 - -0.35) | 0.63 | 0.04 | (0.24 - -0.17) | 0.72 | -0.11 | (-0.45 - 0.23) | 0.54 |
| | R | 1.96 | 0.12 | 0.28 | -0.11 | (0.17 - -0.4) | 0.43 | 0.06 | (0.27 - -0.14) | 0.56 | -0.18 | (-0.52 - 0.16) | 0.31 |
| Inferior Temporal | L | 0.81 | 0.49 | 0.66 | -0.19 | (0.1 - -0.47) | 0.20 | -0.01 | (0.2 - -0.21) | 0.94 | -0.18 | (-0.52 - 0.16) | 0.31 |
| | R | 2.42 | 0.06 | 0.19 | **-0.36** | **(-0.07 - -0.64)** | **0.01** | 0.10 | (0.3 - -0.11) | 0.37 | **-0.46** | **(-0.8 - -0.11)** | **9.76e-03** |
| Insula | L | 0.98 | 0.40 | 0.57 | 0.04 | (0.33 - -0.24) | 0.78 | -0.07 | (0.14 - -0.28) | 0.51 | 0.12 | (-0.23 - 0.46) | 0.53 |
| | R | 0.37 | 0.78 | 0.88 | -0.01 | (0.28 - -0.3) | 0.95 | 0.01 | (0.21 - -0.2) | 0.95 | -0.02 | (-0.36 - 0.33) | 0.92 |
| Isthmus Cingulate | L | 0.16 | 0.93 | 0.96 | -0.07 | (0.21 - -0.36) | 0.62 | 0.03 | (0.24 - -0.17) | 0.76 | -0.11 | (-0.45 - 0.24) | 0.56 |
| | R | 3.48 | 0.02 | 0.08 | **-0.33** | **(-0.04 - -0.61)** | **0.03** | 0.03 | (0.23 - -0.18) | 0.80 | **-0.36** | **(-0.71 - -0.02)** | **0.04** |
| Lateral Occipital | L | 3.73 | 0.01 | 0.06 | **-0.41** | **(-0.12 - -0.69)** | **5.32e-03** | 0.18 | (0.39 - -0.03) | 0.09 | **-0.62** | **(-0.97 - -0.27)** | **8.48e-04** |
| | R | 0.62 | 0.60 | 0.75 | -0.18 | (0.1 - -0.46) | 0.22 | 0.00 | (0.21 - -0.21) | 0.98 | -0.18 | (-0.53 - 0.16) | 0.31 |
| Lateral Orbitofrontal | L | 0.32 | 0.81 | 0.90 | -0.04 | (0.25 - -0.32) | 0.81 | -0.09 | (0.11 - -0.3) | 0.37 | 0.06 | (-0.28 - 0.4) | 0.74 |
| | R | 0.34 | 0.80 | 0.89 | -0.07 | (0.21 - -0.36) | 0.62 | -0.06 | (0.15 - -0.26) | 0.59 | -0.02 | (-0.36 - 0.33) | 0.93 |
| Lingual | L | 1.12 | 0.34 | 0.51 | 0.07 | (0.36 - -0.21) | 0.62 | 0.08 | (0.28 - -0.13) | 0.47 | 0.00 | (-0.35 - 0.34) | 0.98 |
| | R | 0.53 | 0.66 | 0.81 | -0.18 | (0.11 - -0.47) | 0.22 | 0.01 | (0.21 - -0.2) | 0.95 | -0.21 | (-0.56 - 0.14) | 0.29 |
| Medial Orbitofrontal | L | 0.13 | 0.94 | 0.97 | -0.06 | (0.22 - -0.35) | 0.66 | 0.01 | (0.22 - -0.19) | 0.91 | -0.08 | (-0.42 - 0.26) | 0.66 |
| | R | 1.57 | 0.19 | 0.36 | -0.06 | (0.22 - -0.35) | 0.66 | 0.04 | (0.25 - -0.17) | 0.71 | -0.11 | (-0.45 - 0.24) | 0.56 |
| Middle Temporal | L | 0.73 | 0.54 | 0.70 | -0.21 | (0.08 - -0.49) | 0.16 | -0.04 | (0.17 - -0.24) | 0.72 | -0.18 | (-0.52 - 0.17) | 0.34 |
| | R | 1.17 | 0.32 | 0.49 | -0.09 | (0.2 - -0.37) | 0.56 | 0.04 | (0.25 - -0.16) | 0.69 | -0.13 | (-0.47 - 0.21) | 0.47 |
| Paracentral | L | 0.17 | 0.92 | 0.96 | -0.04 | (0.24 - -0.32) | 0.79 | -0.02 | (0.18 - -0.23) | 0.83 | -0.02 | (-0.36 - 0.33) | 0.93 |
| | R | 0.84 | 0.47 | 0.65 | 0.00 | (0.28 - -0.29) | 0.97 | -0.17 | (0.04 - -0.37) | 0.11 | 0.17 | (-0.17 - 0.51) | 0.36 |
| Parahippocampal | L | 1.52 | 0.21 | 0.38 | 0.24 | (0.52 - -0.04) | 0.10 | 0.06 | (0.27 - -0.14) | 0.56 | 0.18 | (-0.16 - 0.52) | 0.31 |
| | R | 0.75 | 0.52 | 0.70 | 0.01 | (0.29 - -0.27) | 0.94 | -0.02 | (0.19 - -0.23) | 0.85 | 0.03 | (-0.31 - 0.37) | 0.86 |
| Pars Opercularis | L | 2.11 | 0.10 | 0.25 | 0.05 | (0.33 - -0.24) | 0.76 | -0.12 | (0.09 - -0.33) | 0.25 | 0.17 | (-0.17 - 0.52) | 0.34 |
| | R | 0.59 | 0.62 | 0.77 | -0.09 | (0.19 - -0.38) | 0.51 | -0.12 | (0.09 - -0.32) | 0.27 | 0.02 | (-0.32 - 0.37) | 0.90 |
| Pars Orbitalis | L | 1.12 | 0.34 | 0.51 | -0.18 | (0.11 - -0.46) | 0.22 | -0.13 | (0.07 - -0.34) | 0.21 | -0.05 | (-0.39 - 0.29) | 0.79 |
| | R | 0.69 | 0.56 | 0.71 | -0.14 | (0.14 - -0.43) | 0.32 | -0.04 | (0.17 - -0.25) | 0.71 | -0.11 | (-0.45 - 0.23) | 0.55 |
| Pars Triangularis | L | 1.25 | 0.29 | 0.47 | -0.08 | (0.2 - -0.36) | 0.59 | -0.12 | (0.09 - -0.32) | 0.27 | 0.04 | (-0.3 - 0.38) | 0.82 |
| | R | 0.18 | 0.91 | 0.96 | -0.09 | (0.19 - | 0.54 | 0.01 | (0.22 - -0.2) | 0.92 | -0.10 | (-0.44 - | 0.57 |

| | | | | | | | | | | | | | |
|---|---|---|---|---|---|---|---|---|---|---|---|---|---|
| | | | | | | -0.37) | | | | | | 0.24) | |
| Pericalcarine | L | 1.05 | 0.37 | 0.54 | -0.18 | (0.1 - -0.47) | 0.21 | -0.05 | (0.16 - -0.26) | 0.64 | -0.15 | (-0.5 - 0.2) | 0.45 |
| | R | 2.27 | 0.08 | 0.22 | -0.04 | (0.25 - -0.33) | 0.81 | -0.02 | (0.19 - -0.23) | 0.86 | -0.02 | (-0.37 - 0.33) | 0.92 |
| Postcentral | L | 0.10 | 0.96 | 0.97 | 0.03 | (0.31 - -0.25) | 0.84 | 0.05 | (0.26 - -0.15) | 0.62 | -0.03 | (-0.37 - 0.32) | 0.89 |
| | R | 1.24 | 0.30 | 0.48 | 0.20 | (0.49 - -0.08) | 0.17 | -0.07 | (0.14 - -0.27) | 0.53 | 0.29 | (-0.05 - 0.64) | 0.13 |
| Posterior Cingulate | L | 1.39 | 0.24 | 0.42 | -0.19 | (0.09 - -0.47) | 0.18 | 0.09 | (0.3 - -0.12) | 0.39 | -0.30 | (-0.64 - 0.04) | 0.10 |
| | R | 1.76 | 0.15 | 0.31 | -0.05 | (0.23 - -0.33) | 0.72 | 0.13 | (0.34 - -0.07) | 0.21 | -0.19 | (-0.53 - 0.15) | 0.29 |
| Precentral | L | 1.20 | 0.31 | 0.49 | -0.20 | (0.08 - -0.49) | 0.16 | 0.01 | (0.22 - -0.2) | 0.91 | -0.22 | (-0.57 - 0.12) | 0.22 |
| | R | 0.23 | 0.87 | 0.94 | -0.10 | (0.18 - -0.39) | 0.47 | -0.02 | (0.19 - -0.22) | 0.88 | -0.09 | (-0.44 - 0.25) | 0.61 |
| Precuneus | L | 1.05 | 0.37 | 0.54 | 0.08 | (0.37 - -0.2) | 0.57 | 0.12 | (0.33 - -0.08) | 0.25 | -0.04 | (-0.38 - 0.3) | 0.82 |
| | R | 0.44 | 0.73 | 0.85 | 0.07 | (0.35 - -0.22) | 0.64 | 0.00 | (0.21 - -0.2) | 0.99 | 0.07 | (-0.28 - 0.41) | 0.70 |
| Rostral Anterior Cingulate | L | 3.78 | 0.01 | 0.06 | -0.20 | (0.08 - -0.48) | 0.16 | -0.14 | (0.06 - -0.35) | 0.18 | -0.06 | (-0.4 - 0.28) | 0.73 |
| | R | 2.16 | 0.09 | 0.24 | -0.17 | (0.12 - -0.45) | 0.25 | -0.13 | (0.08 - -0.34) | 0.22 | -0.04 | (-0.38 - 0.3) | 0.82 |
| Rostral Middle Frontal | L | 2.43 | 0.06 | 0.19 | **-0.36** | **(-0.07 - -0.64)** | **0.01** | 0.10 | (0.31 - -0.11) | 0.35 | **-0.47** | **(-0.82 - -0.12)** | **9.50e-03** |
| | R | **4.48** | **3.81e-03** | **0.05** | **-0.43** | **(-0.15 - -0.71)** | **2.90e-03** | 0.20 | (0.41 - 0) | 0.06 | **-0.65** | **(-1 - -0.3)** | **2.92e-04** |
| Superior Frontal | L | 3.24 | 0.02 | 0.10 | **-0.34** | **(-0.06 - -0.62)** | **0.02** | -0.21 | (0 - -0.41) | 0.05 | -0.14 | (-0.48 - 0.2) | 0.43 |
| | R | 3.65 | 0.01 | 0.06 | -0.22 | (0.06 - -0.51) | 0.12 | -0.13 | (0.08 - -0.33) | 0.23 | -0.10 | (-0.44 - 0.24) | 0.58 |
| Superior Parietal | L | 0.12 | 0.95 | 0.97 | 0.07 | (0.35 - -0.22) | 0.64 | -0.03 | (0.18 - -0.24) | 0.77 | 0.10 | (-0.24 - 0.44) | 0.58 |
| | R | 0.63 | 0.59 | 0.75 | -0.15 | (0.13 - -0.43) | 0.30 | 0.08 | (0.28 - -0.13) | 0.47 | -0.23 | (-0.58 - 0.11) | 0.19 |
| Superior Temporal | L | 0.15 | 0.93 | 0.96 | -0.01 | (0.28 - -0.3) | 0.96 | -0.01 | (0.2 - -0.22) | 0.92 | 0.00 | (-0.34 - 0.35) | 0.98 |
| | R | 0.91 | 0.44 | 0.60 | -0.08 | (0.2 - -0.36) | 0.58 | 0.04 | (0.25 - -0.16) | 0.69 | -0.13 | (-0.47 - 0.22) | 0.49 |
| Supramarginal | L | 2.82 | 0.04 | 0.14 | 0.11 | (0.4 - -0.18) | 0.47 | 0.04 | (0.25 - -0.17) | 0.70 | 0.07 | (-0.27 - 0.42) | 0.71 |
| | R | 1.47 | 0.22 | 0.40 | 0.04 | (0.32 - -0.24) | 0.78 | 0.08 | (0.29 - -0.12) | 0.43 | -0.05 | (-0.39 - 0.3) | 0.81 |
| Temporal Pole | L | 0.06 | 0.98 | 0.98 | 0.05 | (0.33 - -0.23) | 0.73 | -0.01 | (0.2 - -0.22) | 0.92 | 0.06 | (-0.28 - 0.4) | 0.73 |
| | R | 3.25 | 0.02 | 0.10 | -0.21 | (0.08 - -0.49) | 0.15 | **-0.26** | **(-0.05 - -0.47)** | **0.01** | 0.05 | (-0.29 - 0.4) | 0.77 |
| Transverse Temporal | L | 1.16 | 0.32 | 0.49 | 0.13 | (0.41 - -0.16) | 0.39 | -0.11 | (0.09 - -0.32) | 0.29 | 0.26 | (-0.09 - 0.6) | 0.18 |
| | R | 0.96 | 0.41 | 0.58 | -0.03 | (0.25 - -0.32) | 0.83 | 0.16 | (0.37 - -0.04) | 0.12 | -0.20 | (-0.55 - 0.14) | 0.27 |
| **Subcortical Volume** | | | | | | | | | | | | | |
| Amygdala | L | 0.73 | 0.54 | 0.70 | -0.13 | (0.16 - -0.41) | 0.38 | -0.08 | (0.12 - -0.29) | 0.42 | -0.05 | (-0.39 - 0.3) | 0.80 |

| | | | | | | | | | | | | | |
|---|---|---|---|---|---|---|---|---|---|---|---|---|---|
| | R | 3.05 | 0.03 | 0.12 | **-0.29** | **(-0.01 - -0.57)** | **0.05** | -0.10 | (0.11 - -0.3) | 0.36 | -0.20 | (-0.54 - 0.14) | 0.27 |
| Caudate | L | 0.47 | 0.71 | 0.84 | 0.16 | (0.45 - -0.13) | 0.28 | 0.04 | (0.25 - -0.17) | 0.69 | 0.12 | (-0.22 - 0.47) | 0.51 |
| | R | 0.25 | 0.86 | 0.93 | 0.09 | (0.38 - -0.2) | 0.54 | 0.01 | (0.22 - -0.2) | 0.93 | 0.08 | (-0.26 - 0.43) | 0.65 |
| Hippocampus | L | 1.25 | 0.29 | 0.47 | -0.01 | (0.27 - -0.29) | 0.96 | -0.09 | (0.11 - -0.3) | 0.38 | 0.09 | (-0.25 - 0.43) | 0.63 |
| | R | **5.59** | **8.01e-04** | **0.02** | 0.04 | (0.32 - -0.24) | 0.78 | -0.10 | (0.11 - -0.31) | 0.34 | 0.15 | (-0.2 - 0.49) | 0.42 |
| Lateral Ventricles | L | 4.09 | 6.54e-03 | 0.06 | 0.15 | (0.44 - -0.14) | 0.30 | 0.18 | (0.39 - -0.02) | 0.08 | -0.03 | (-0.38 - 0.31) | 0.86 |
| | R | 3.79 | 9.99e-03 | 0.06 | **0.30** | **(0.59 - 0.01)** | **0.05** | 0.01 | (0.21 - -0.2) | 0.95 | 0.33 | (-0.02 - 0.68) | 0.10 |
| Nucleus Accumbens | L | 0.28 | 0.84 | 0.91 | 0.08 | (0.37 - -0.2) | 0.57 | 0.06 | (0.27 - -0.15) | 0.57 | 0.03 | (-0.32 - 0.37) | 0.89 |
| | R | 0.05 | 0.98 | 0.98 | 0.04 | (0.33 - -0.24) | 0.77 | 0.02 | (0.23 - -0.19) | 0.84 | 0.02 | (-0.32 - 0.37) | 0.90 |
| Pallidum | L | 0.35 | 0.79 | 0.89 | -0.04 | (0.24 - -0.33) | 0.76 | -0.10 | (0.11 - -0.3) | 0.36 | 0.05 | (-0.29 - 0.39) | 0.77 |
| | R | 1.06 | 0.36 | 0.54 | 0.06 | (0.35 - -0.22) | 0.65 | -0.18 | (0.03 - -0.38) | 0.10 | 0.25 | (-0.1 - 0.59) | 0.17 |
| Putamen | L | 0.16 | 0.93 | 0.96 | -0.05 | (0.24 - -0.33) | 0.74 | 0.06 | (0.26 - -0.15) | 0.60 | -0.11 | (-0.45 - 0.24) | 0.55 |
| | R | 0.31 | 0.82 | 0.91 | -0.12 | (0.17 - -0.41) | 0.41 | -0.05 | (0.15 - -0.26) | 0.62 | -0.07 | (-0.42 - 0.28) | 0.69 |
| Thalamus | L | 1.30 | 0.27 | 0.46 | -0.09 | (0.2 - -0.37) | 0.55 | -0.18 | (0.03 - -0.38) | 0.10 | 0.09 | (-0.25 - 0.43) | 0.62 |
| | R | 2.45 | 0.06 | 0.19 | -0.01 | (0.28 - -0.29) | 0.97 | -0.17 | (0.03 - -0.38) | 0.10 | 0.17 | (-0.17 - 0.51) | 0.34 |
| **Global** | | | | | | | | | | | | | |
| Estimated Intracranial Volume | | 4.28 | 5.03e-03 | 0.06 | 0.01 | (0.3 - -0.27) | 0.92 | 0.00 | (0.21 - -0.2) | 0.97 | 0.01 | (-0.33 - 0.35) | 0.95 |
| Mean Cortical Thickness | N/A | **5.87** | **5.42e-04** | **0.02** | -0.05 | (0.24 - -0.34) | 0.74 | -0.12 | (0.09 - -0.32) | 0.28 | 0.07 | (-0.28 - 0.41) | 0.71 |
| Total Surface Area | | **6.29** | **3.01e-04** | **0.02** | -0.19 | (0.09 - -0.48) | 0.18 | 0.02 | (0.23 - -0.18) | 0.84 | -0.22 | (-0.56 - 0.12) | 0.22 |

*Results marked in bold are significant at q<0.05 (the overall effect) or at p<0.05 (pairwise comparisons). Pairwise comparisons shown in this table are APS < BIPS, APS < GRD, and GRD < BIPS*

**eTable 13.** APS subgroup (APS vs. no APS assignment vs. HC) effect size overview

| | | Overall Test Statistics | | | APS < HC | | | APS < no APS | | | no APS < HC | | |
|---|---|---|---|---|---|---|---|---|---|---|---|---|---|
| **Region of Interest** | **Hemisphere** | **F** | **p** | **q** | **Cohen's d** | **95% Confidence Interval** | **p** | **Cohen's d** | **95% Confidence Interval** | **p** | **Cohen's d** | **95% Confidence Interval** | **p** |
| **Cortical Thickness** | | | | | | | | | | | | | |
| Banks of the Superior Temporal Sulcus | L | 4.05 | 0.02 | 0.07 | **-0.10** | **(-0.17 - -0.02)** | **0.01** | -0.16 | (-0.33 - 0.01) | 0.07 | 0.06 | (-0.11 - 0.23) | 0.50 |
| | R | **6.76** | **1.18e-03** | **0.01** | **-0.14** | **(-0.21 - -0.06)** | **2.44e-04** | -0.08 | (-0.25 - 0.09) | 0.37 | -0.06 | (-0.23 - 0.11) | 0.46 |
| Caudal Anterior Cingulate | L | 1.80 | 0.17 | 0.30 | -0.06 | (-0.14 - 0.01) | 0.09 | 0.04 | (-0.13 - 0.21) | 0.61 | -0.11 | (-0.28 - 0.06) | 0.21 |
| | R | 2.78 | 0.06 | 0.15 | **-0.08** | **(-0.15 - 0)** | **0.04** | -0.14 | (-0.31 - 0.03) | 0.11 | 0.06 | (-0.11 - 0.23) | 0.47 |
| Caudal Middle Frontal | L | 3.78 | 0.02 | 0.08 | **-0.10** | **(-0.18 - -0.03)** | **6.00e-03** | -0.05 | (-0.22 - 0.12) | 0.57 | -0.06 | (-0.23 - 0.11) | 0.52 |
| | R | **6.86** | **1.07e-03** | **0.01** | **-0.13** | **(-0.2 - -0.05)** | **7.91e-04** | **-0.19** | **(-0.36 - -0.02)** | **0.03** | 0.06 | (-0.11 - 0.23) | 0.48 |
| Cuneus | L | 1.33 | 0.27 | 0.43 | 0.01 | (-0.06 - 0.09) | 0.70 | -0.13 | (-0.3 - 0.04) | 0.14 | 0.14 | (-0.03 - 0.31) | 0.10 |
| | R | 1.88 | 0.15 | 0.29 | -0.05 | (-0.12 - 0.03) | 0.23 | -0.15 | (-0.32 - 0.02) | 0.08 | 0.10 | (-0.07 - 0.27) | 0.23 |
| Entorhinal | L | 0.37 | 0.69 | 0.79 | -0.01 | (-0.09 - 0.06) | 0.70 | 0.06 | (-0.11 - 0.23) | 0.50 | -0.07 | (-0.24 - 0.1) | 0.40 |
| | R | 1.47 | 0.23 | 0.38 | -0.04 | (-0.12 - 0.03) | 0.24 | 0.08 | (-0.09 - 0.25) | 0.33 | -0.13 | (-0.3 - 0.04) | 0.14 |
| Frontal Pole | L | 1.17 | 0.31 | 0.47 | -0.04 | (-0.12 - 0.03) | 0.25 | -0.10 | (-0.27 - 0.06) | 0.23 | 0.06 | (-0.11 - 0.23) | 0.49 |
| | R | 1.87 | 0.15 | 0.29 | -0.07 | (-0.15 - 0) | 0.05 | -0.03 | (-0.2 - 0.15) | 0.77 | -0.05 | (-0.22 - 0.12) | 0.58 |
| Fusiform | L | **9.93** | **5.05e-05** | **1.75e-03** | **-0.17** | **(-0.24 - -0.09)** | **1.17e-05** | -0.14 | (-0.31 - 0.03) | 0.10 | -0.03 | (-0.2 - 0.14) | 0.76 |
| | R | 3.25 | 0.04 | 0.11 | **-0.10** | **(-0.17 - -0.02)** | **0.01** | -0.05 | (-0.22 - 0.12) | 0.54 | -0.04 | (-0.21 - 0.13) | 0.61 |
| Inferior Parietal | L | **10.21** | **3.82e-05** | **1.75e-03** | **-0.17** | **(-0.24 - -0.09)** | **8.60e-06** | -0.14 | (-0.31 - 0.03) | 0.10 | -0.03 | (-0.2 - 0.14) | 0.74 |
| | R | **6.20** | **2.06e-03** | **0.02** | **-0.12** | **(-0.19 - -0.05)** | **1.38e-03** | **-0.18** | **(-0.35 - -0.01)** | **0.04** | 0.06 | (-0.11 - 0.23) | 0.51 |
| Inferior Temporal | L | 4.24 | 0.01 | 0.06 | **-0.10** | **(-0.18 - -0.03)** | **6.67e-03** | 0.04 | (-0.13 - 0.21) | 0.63 | -0.14 | (-0.31 - 0.02) | 0.09 |
| | R | **6.47** | **1.57e-03** | **0.01** | **-0.13** | **(-0.21 - -0.06)** | **4.79e-04** | 0.01 | (-0.16 - 0.18) | 0.91 | -0.14 | (-0.31 - 0.03) | 0.10 |
| Insula | L | **9.17** | **1.08e-04** | **2.78e-03** | **-0.16** | **(-0.23 - -0.08)** | **2.97e-05** | 0.00 | (-0.16 - 0.17) | 0.97 | -0.16 | (-0.33 - 0.01) | 0.06 |
| | R | **7.19** | **7.68e-04** | **0.01** | **-0.14** | **(-0.21 - -0.07)** | **2.31e-04** | 0.01 | (-0.16 - 0.18) | 0.91 | -0.15 | (-0.32 - 0.02) | 0.08 |
| Isthmus Cingulate | L | 0.14 | 0.87 | 0.92 | -0.01 | (-0.09 - 0.06) | 0.69 | 0.02 | (-0.15 - 0.19) | 0.80 | -0.04 | (-0.21 - 0.13) | 0.67 |
| | R | 0.94 | 0.39 | 0.55 | -0.05 | (-0.13 - 0.02) | 0.17 | -0.02 | (-0.19 - 0.15) | 0.84 | -0.03 | (-0.21 - 0.14) | 0.69 |
| Lateral Occipital | L | **4.94** | **7.23e-03** | **0.04** | **-0.11** | **(-0.19 - -0.04)** | **2.70e-03** | -0.13 | (-0.3 - 0.04) | 0.13 | 0.02 | (-0.15 - 0.19) | 0.85 |
| | R | **7.90** | **3.79e-04** | **7.34e-03** | **-0.15** | **(-0.22 - -0.08)** | **7.86e-05** | -0.04 | (-0.21 - 0.14) | 0.69 | -0.12 | (-0.29 - 0.06) | 0.19 |
| | | | | | | | | | (-0.26 - | | | (-0.15 - | |

| | | | | | | | | | | | | | |
|---|---|---|---|---|---|---|---|---|---|---|---|---|---|
| Lateral Orbitofrontal | L | 2.00 | 0.14 | 0.27 | -0.07 | (-0.14 - 0) | 0.06 | -0.09 | 0.08) | 0.28 | 0.02 | 0.19) | 0.81 |
| | R | 2.74 | 0.06 | 0.16 | **-0.08** | **(-0.15 - -0.01)** | **0.03** | -0.12 | (-0.29 - 0.05) | 0.17 | 0.04 | (-0.13 - 0.21) | 0.67 |
| Lingual | L | 1.85 | 0.16 | 0.29 | -0.03 | (-0.11 - 0.04) | 0.40 | -0.16 | (-0.33 - 0.01) | 0.06 | 0.13 | (-0.04 - 0.3) | 0.14 |
| | R | 0.31 | 0.73 | 0.82 | 0.00 | (-0.07 - 0.08) | 0.92 | -0.06 | (-0.24 - 0.11) | 0.46 | 0.07 | (-0.1 - 0.24) | 0.43 |
| Medial Orbitofrontal | L | **4.81** | **8.21e-03** | **0.04** | **-0.12** | **(-0.19 - -0.04)** | **2.22e-03** | -0.01 | (-0.18 - 0.16) | 0.89 | -0.11 | (-0.27 - 0.06) | 0.23 |
| | R | 3.49 | 0.03 | 0.10 | **-0.10** | **(-0.17 - -0.02)** | **0.01** | -0.10 | (-0.27 - 0.07) | 0.23 | 0.00 | (-0.16 - 0.17) | 0.96 |
| Middle Temporal | L | 2.88 | 0.06 | 0.15 | **-0.09** | **(-0.16 - -0.02)** | **0.02** | -0.04 | (-0.21 - 0.13) | 0.61 | -0.05 | (-0.22 - 0.12) | 0.58 |
| | R | **11.84** | **7.59e-06** | **1.18e-03** | **-0.18** | **(-0.26 - -0.11)** | **1.20e-06** | -0.09 | (-0.26 - 0.08) | 0.30 | -0.10 | (-0.27 - 0.07) | 0.27 |
| Paracentral | L | **8.84** | **1.49e-04** | **3.29e-03** | **-0.15** | **(-0.22 - -0.08)** | **6.45e-05** | **-0.18** | **(-0.35 - -0.01)** | **0.04** | 0.03 | (-0.14 - 0.2) | 0.76 |
| | R | **5.33** | **4.88e-03** | **0.03** | **-0.11** | **(-0.18 - -0.03)** | **5.03e-03** | **-0.19** | **(-0.36 - -0.02)** | **0.03** | 0.08 | (-0.09 - 0.25) | 0.34 |
| Parahippocampal | L | **5.72** | **3.32e-03** | **0.02** | **-0.13** | **(-0.2 - -0.05)** | **8.44e-04** | -0.10 | (-0.27 - 0.07) | 0.23 | -0.03 | (-0.19 - 0.14) | 0.77 |
| | R | 1.48 | 0.23 | 0.38 | -0.06 | (-0.14 - 0.01) | 0.09 | -0.06 | (-0.23 - 0.11) | 0.50 | -0.01 | (-0.18 - 0.16) | 0.93 |
| Pars Opercularis | L | 1.12 | 0.33 | 0.49 | -0.04 | (-0.12 - 0.03) | 0.25 | -0.10 | (-0.27 - 0.07) | 0.24 | 0.06 | (-0.11 - 0.23) | 0.51 |
| | R | 1.68 | 0.19 | 0.33 | -0.07 | (-0.14 - 0.01) | 0.08 | -0.08 | (-0.25 - 0.09) | 0.36 | 0.01 | (-0.16 - 0.18) | 0.89 |
| Pars Orbitalis | L | 1.17 | 0.31 | 0.47 | -0.05 | (-0.12 - 0.02) | 0.19 | -0.09 | (-0.26 - 0.08) | 0.30 | 0.04 | (-0.13 - 0.21) | 0.66 |
| | R | 0.66 | 0.52 | 0.66 | 0.02 | (-0.06 - 0.09) | 0.63 | 0.10 | (-0.07 - 0.27) | 0.26 | -0.08 | (-0.25 - 0.09) | 0.37 |
| Pars Triangularis | L | 0.41 | 0.67 | 0.78 | 0.03 | (-0.04 - 0.11) | 0.39 | 0.04 | (-0.13 - 0.2) | 0.68 | 0.00 | (-0.17 - 0.17) | 0.98 |
| | R | 0.81 | 0.45 | 0.61 | -0.05 | (-0.12 - 0.03) | 0.22 | -0.05 | (-0.22 - 0.12) | 0.55 | 0.01 | (-0.17 - 0.18) | 0.95 |
| Pericalcarine | L | 2.70 | 0.07 | 0.16 | 0.02 | (-0.06 - 0.09) | 0.67 | **-0.19** | **(-0.36 - -0.02)** | **0.03** | **0.20** | **(0.03 - 0.37)** | **0.02** |
| | R | 3.39 | 0.03 | 0.11 | 0.05 | (-0.02 - 0.13) | 0.17 | -0.17 | (-0.34 - 0.01) | 0.06 | **0.22** | **(0.05 - 0.39)** | **0.01** |
| Postcentral | L | 2.59 | 0.08 | 0.17 | **-0.08** | **(-0.16 - -0.01)** | **0.03** | 0.02 | (-0.15 - 0.19) | 0.78 | -0.11 | (-0.28 - 0.06) | 0.22 |
| | R | **4.67** | **9.48e-03** | **0.05** | **-0.12** | **(-0.19 - -0.04)** | **2.28e-03** | -0.05 | (-0.22 - 0.12) | 0.57 | -0.07 | (-0.24 - 0.1) | 0.44 |
| Posterior Cingulate | L | 3.08 | 0.05 | 0.13 | **-0.09** | **(-0.17 - -0.02)** | **0.01** | -0.03 | (-0.2 - 0.14) | 0.75 | -0.07 | (-0.24 - 0.1) | 0.44 |
| | R | **5.41** | **4.50e-03** | **0.03** | **-0.12** | **(-0.2 - -0.05)** | **1.01e-03** | -0.06 | (-0.23 - 0.11) | 0.50 | -0.07 | (-0.24 - 0.1) | 0.44 |
| Precentral | L | 4.42 | 0.01 | 0.05 | **-0.11** | **(-0.19 - -0.04)** | **2.98e-03** | -0.05 | (-0.22 - 0.11) | 0.53 | -0.06 | (-0.23 - 0.11) | 0.49 |
| | R | **6.02** | **2.45e-03** | **0.02** | **-0.12** | **(-0.2 - -0.05)** | **1.05e-03** | -0.15 | (-0.32 - 0.02) | 0.08 | 0.03 | (-0.14 - 0.2) | 0.75 |
| Precuneus | L | **6.51** | **1.50e-03** | **0.01** | **-0.11** | **(-0.19 - -0.04)** | **2.63e-03** | **-0.22** | **(-0.39 - -0.05)** | **0.01** | 0.11 | (-0.06 - 0.28) | 0.22 |
| | R | **6.30** | **1.87e-03** | **0.02** | **-0.13** | **(-0.21 - -0.06)** | **3.93e-04** | -0.07 | (-0.24 - 0.1) | 0.44 | -0.07 | (-0.24 - 0.1) | 0.43 |
| Rostral Anterior | L | 3.33 | 0.04 | 0.11 | **-0.09** | **(-0.16 - -0.02)** | **0.02** | 0.04 | (-0.13 - 0.21) | 0.67 | -0.13 | (-0.3 - 0.04) | 0.14 |

| | | | | | | | | | | | | | |
|---|---|---|---|---|---|---|---|---|---|---|---|---|---|
| Cingulate | R | 1.93 | 0.15 | 0.29 | -0.07 | (-0.15 - 0) | 0.05 | -0.06 | (-0.22 - 0.11) | 0.52 | -0.02 | (-0.19 - 0.15) | 0.83 |
| Rostral Middle Frontal | L | **5.11** | **6.09e-03** | **0.03** | **-0.12** | **(-0.19 - -0.05)** | **1.40e-03** | -0.05 | (-0.22 - 0.12) | 0.54 | -0.07 | (-0.24 - 0.1) | 0.43 |
| | R | 4.12 | 0.02 | 0.06 | **-0.11** | **(-0.18 - -0.03)** | **4.40e-03** | -0.02 | (-0.19 - 0.15) | 0.81 | -0.09 | (-0.26 - 0.08) | 0.32 |
| Superior Frontal | L | 3.43 | 0.03 | 0.11 | **-0.10** | **(-0.17 - -0.02)** | **9.21e-03** | -0.07 | (-0.24 - 0.1) | 0.44 | -0.03 | (-0.2 - 0.14) | 0.71 |
| | R | 2.82 | 0.06 | 0.15 | **-0.08** | **(-0.16 - -0.01)** | **0.03** | -0.11 | (-0.28 - 0.06) | 0.21 | 0.02 | (-0.15 - 0.19) | 0.78 |
| Superior Parietal | L | 4.14 | 0.02 | 0.06 | **-0.10** | **(-0.17 - -0.03)** | **7.41e-03** | -0.14 | (-0.3 - 0.03) | 0.12 | 0.03 | (-0.14 - 0.2) | 0.71 |
| | R | **6.70** | **1.25e-03** | **0.01** | **-0.12** | **(-0.2 - -0.05)** | **1.06e-03** | **-0.20** | **(-0.36 - -0.03)** | **0.02** | 0.07 | (-0.1 - 0.24) | 0.42 |
| Superior Temporal | L | 4.46 | 0.01 | 0.05 | **-0.11** | **(-0.19 - -0.04)** | **2.92e-03** | -0.07 | (-0.24 - 0.1) | 0.41 | -0.04 | (-0.21 - 0.13) | 0.63 |
| | R | **6.96** | **9.69e-04** | **0.01** | **-0.13** | **(-0.2 - -0.05)** | **7.18e-04** | 0.07 | (-0.1 - 0.24) | 0.40 | **-0.20** | **(-0.37 - -0.03)** | **0.02** |
| Supramarginal | L | 2.94 | 0.05 | 0.14 | **-0.09** | **(-0.16 - -0.02)** | **0.02** | 0.00 | (-0.16 - 0.17) | 0.97 | -0.09 | (-0.26 - 0.07) | 0.28 |
| | R | 2.65 | 0.07 | 0.16 | **-0.09** | **(-0.16 - -0.01)** | **0.02** | -0.05 | (-0.22 - 0.12) | 0.56 | -0.04 | (-0.21 - 0.13) | 0.66 |
| Temporal Pole | L | 1.00 | 0.37 | 0.52 | -0.05 | (-0.12 - 0.02) | 0.18 | 0.01 | (-0.16 - 0.18) | 0.90 | -0.06 | (-0.23 - 0.11) | 0.47 |
| | R | 2.28 | 0.10 | 0.23 | **-0.08** | **(-0.15 - -0.01)** | **0.03** | -0.05 | (-0.22 - 0.12) | 0.59 | -0.04 | (-0.2 - 0.13) | 0.69 |
| Transverse Temporal | L | 1.25 | 0.29 | 0.45 | -0.06 | (-0.13 - 0.01) | 0.12 | -0.01 | (-0.18 - 0.16) | 0.88 | -0.05 | (-0.22 - 0.12) | 0.59 |
| | R | 3.31 | 0.04 | 0.11 | **-0.09** | **(-0.16 - -0.01)** | **0.02** | 0.05 | (-0.12 - 0.22) | 0.54 | -0.14 | (-0.31 - 0.03) | 0.10 |
| **Surface Area** | | | | | | | | | | | | | |
| Banks of the Superior Temporal Sulcus | L | 2.18 | 0.11 | 0.24 | **-0.08** | **(-0.15 - 0)** | **0.04** | -0.08 | (-0.25 - 0.09) | 0.38 | 0.00 | (-0.17 - 0.17) | 0.98 |
| | R | 2.23 | 0.11 | 0.23 | **-0.08** | **(-0.15 - -0.01)** | **0.03** | -0.04 | (-0.21 - 0.13) | 0.66 | -0.04 | (-0.21 - 0.13) | 0.63 |
| Caudal Anterior Cingulate | L | 0.29 | 0.75 | 0.83 | -0.02 | (-0.09 - 0.06) | 0.64 | -0.06 | (-0.23 - 0.11) | 0.50 | 0.04 | (-0.13 - 0.21) | 0.64 |
| | R | 2.91 | 0.05 | 0.14 | **-0.09** | **(-0.16 - -0.01)** | **0.02** | -0.09 | (-0.26 - 0.08) | 0.31 | 0.00 | (-0.17 - 0.17) | 0.97 |
| Caudal Middle Frontal | L | 1.76 | 0.17 | 0.31 | -0.06 | (-0.14 - 0.01) | 0.10 | -0.10 | (-0.27 - 0.06) | 0.23 | 0.04 | (-0.13 - 0.21) | 0.63 |
| | R | 2.73 | 0.07 | 0.16 | -0.04 | (-0.11 - 0.03) | 0.28 | **-0.19** | **(-0.36 - -0.03)** | **0.02** | 0.15 | (-0.02 - 0.32) | 0.08 |
| Cuneus | L | 2.17 | 0.11 | 0.24 | -0.07 | (-0.14 - 0.01) | 0.07 | -0.12 | (-0.29 - 0.05) | 0.16 | 0.05 | (-0.12 - 0.23) | 0.54 |
| | R | 2.67 | 0.07 | 0.16 | **-0.09** | **(-0.16 - -0.01)** | **0.02** | -0.02 | (-0.19 - 0.15) | 0.78 | -0.06 | (-0.24 - 0.11) | 0.46 |
| Entorhinal | L | 3.39 | 0.03 | 0.11 | **-0.10** | **(-0.17 - -0.02)** | **9.54e-03** | -0.02 | (-0.19 - 0.14) | 0.77 | -0.07 | (-0.24 - 0.1) | 0.39 |
| | R | 1.27 | 0.28 | 0.45 | -0.04 | (-0.11 - 0.04) | 0.35 | -0.12 | (-0.29 - 0.04) | 0.15 | 0.09 | (-0.08 - 0.26) | 0.31 |
| Frontal Pole | L | 0.56 | 0.57 | 0.69 | 0.03 | (-0.04 - 0.1) | 0.43 | -0.05 | (-0.21 - 0.12) | 0.59 | 0.08 | (-0.09 - 0.25) | 0.38 |
| | R | 0.92 | 0.40 | 0.56 | 0.04 | (-0.04 - 0.11) | 0.31 | -0.06 | (-0.23 - 0.11) | 0.51 | 0.10 | (-0.07 - 0.27) | 0.27 |
| Fusiform | L | 1.02 | 0.36 | 0.52 | -0.05 | (-0.13 - 0.02) | 0.16 | -0.05 | (-0.22 - 0.12) | 0.56 | 0.00 | (-0.17 - 0.17) | 0.97 |

| | | | | | | | | | | | | | |
|---|---|---|---|---|---|---|---|---|---|---|---|---|---|
| | R | 3.24 | 0.04 | 0.11 | **-0.08** | **(-0.16 - -0.01)** | **0.03** | -0.14 | (-0.31 - 0.03) | 0.10 | 0.05 | (-0.11 - 0.22) | 0.53 |
| Inferior Parietal | L | 1.19 | 0.30 | 0.47 | -0.06 | (-0.13 - 0.02) | 0.13 | 0.00 | (-0.17 - 0.17) | 0.98 | -0.06 | (-0.22 - 0.11) | 0.52 |
| | R | 3.67 | 0.03 | 0.09 | **-0.10** | **(-0.17 - -0.03)** | **7.38e-03** | -0.01 | (-0.18 - 0.15) | 0.87 | -0.09 | (-0.26 - 0.08) | 0.30 |
| Inferior Temporal | L | 0.64 | 0.53 | 0.66 | -0.04 | (-0.11 - 0.04) | 0.32 | -0.06 | (-0.23 - 0.1) | 0.46 | 0.03 | (-0.14 - 0.2) | 0.76 |
| | R | 0.47 | 0.63 | 0.74 | -0.03 | (-0.1 - 0.04) | 0.44 | -0.06 | (-0.23 - 0.11) | 0.47 | 0.03 | (-0.14 - 0.2) | 0.70 |
| Insula | L | 1.51 | 0.22 | 0.38 | -0.07 | (-0.14 - 0.01) | 0.08 | -0.02 | (-0.19 - 0.15) | 0.79 | -0.04 | (-0.21 - 0.13) | 0.62 |
| | R | 0.75 | 0.47 | 0.64 | -0.05 | (-0.12 - 0.03) | 0.23 | 0.00 | (-0.17 - 0.17) | 0.97 | -0.04 | (-0.21 - 0.13) | 0.62 |
| Isthmus Cingulate | L | 0.03 | 0.97 | 0.97 | -0.01 | (-0.08 - 0.07) | 0.86 | 0.01 | (-0.16 - 0.18) | 0.90 | -0.02 | (-0.19 - 0.15) | 0.84 |
| | R | 3.47 | 0.03 | 0.10 | **0.08** | **(0.01 - 0.15)** | **0.04** | -0.10 | (-0.27 - 0.07) | 0.26 | **0.18** | **(0.01 - 0.35)** | **0.04** |
| Lateral Occipital | L | 0.05 | 0.95 | 0.97 | -0.01 | (-0.08 - 0.06) | 0.82 | -0.02 | (-0.19 - 0.15) | 0.79 | 0.01 | (-0.16 - 0.18) | 0.87 |
| | R | 0.31 | 0.74 | 0.82 | -0.02 | (-0.09 - 0.05) | 0.59 | -0.06 | (-0.23 - 0.11) | 0.51 | 0.04 | (-0.13 - 0.21) | 0.67 |
| Lateral Orbitofrontal | L | 0.54 | 0.58 | 0.70 | -0.03 | (-0.1 - 0.04) | 0.43 | -0.07 | (-0.24 - 0.1) | 0.40 | 0.04 | (-0.13 - 0.21) | 0.63 |
| | R | 0.50 | 0.61 | 0.72 | -0.03 | (-0.11 - 0.04) | 0.37 | -0.05 | (-0.22 - 0.12) | 0.53 | 0.02 | (-0.15 - 0.19) | 0.82 |
| Lingual | L | 1.87 | 0.15 | 0.29 | -0.05 | (-0.13 - 0.02) | 0.15 | 0.08 | (-0.09 - 0.25) | 0.34 | -0.14 | (-0.31 - 0.03) | 0.11 |
| | R | 0.39 | 0.68 | 0.78 | -0.03 | (-0.1 - 0.05) | 0.50 | -0.06 | (-0.23 - 0.11) | 0.49 | 0.03 | (-0.14 - 0.2) | 0.70 |
| Medial Orbitofrontal | L | 0.17 | 0.84 | 0.90 | -0.02 | (-0.1 - 0.05) | 0.56 | -0.01 | (-0.18 - 0.16) | 0.92 | -0.01 | (-0.18 - 0.16) | 0.87 |
| | R | 2.92 | 0.05 | 0.14 | **-0.09** | **(-0.16 - -0.02)** | **0.02** | 0.00 | (-0.17 - 0.17) | 1.00 | -0.09 | (-0.26 - 0.08) | 0.30 |
| Middle Temporal | L | 0.61 | 0.54 | 0.67 | 0.00 | (-0.07 - 0.08) | 0.93 | -0.09 | (-0.26 - 0.08) | 0.29 | 0.10 | (-0.08 - 0.27) | 0.27 |
| | R | 1.88 | 0.15 | 0.29 | -0.07 | (-0.15 - 0) | 0.06 | 0.00 | (-0.17 - 0.17) | 0.99 | -0.07 | (-0.24 - 0.1) | 0.39 |
| Paracentral | L | 0.04 | 0.96 | 0.97 | -0.01 | (-0.08 - 0.06) | 0.78 | -0.01 | (-0.18 - 0.16) | 0.91 | 0.00 | (-0.17 - 0.17) | 0.99 |
| | R | 0.69 | 0.50 | 0.65 | 0.00 | (-0.07 - 0.08) | 0.94 | -0.10 | (-0.27 - 0.07) | 0.26 | 0.10 | (-0.07 - 0.27) | 0.25 |
| Parahippocampal | L | 1.82 | 0.16 | 0.30 | -0.04 | (-0.11 - 0.03) | 0.29 | 0.12 | (-0.05 - 0.28) | 0.18 | -0.16 | (-0.32 - 0.01) | 0.07 |
| | R | 1.27 | 0.28 | 0.45 | 0.06 | (-0.02 - 0.13) | 0.12 | 0.00 | (-0.17 - 0.17) | 0.98 | 0.06 | (-0.11 - 0.23) | 0.48 |
| Pars Opercularis | L | 2.56 | 0.08 | 0.17 | **-0.08** | **(-0.16 - -0.01)** | **0.02** | -0.06 | (-0.23 - 0.11) | 0.47 | -0.02 | (-0.19 - 0.15) | 0.79 |
| | R | 0.78 | 0.46 | 0.63 | -0.01 | (-0.09 - 0.06) | 0.70 | -0.11 | (-0.28 - 0.06) | 0.22 | 0.09 | (-0.08 - 0.26) | 0.29 |
| Pars Orbitalis | L | 1.26 | 0.28 | 0.45 | 0.01 | (-0.07 - 0.08) | 0.83 | -0.13 | (-0.3 - 0.04) | 0.13 | 0.14 | (-0.03 - 0.31) | 0.11 |
| | R | 0.65 | 0.52 | 0.66 | 0.02 | (-0.05 - 0.09) | 0.58 | -0.07 | (-0.24 - 0.09) | 0.39 | 0.10 | (-0.07 - 0.26) | 0.27 |
| Pars Triangularis | L | 1.51 | 0.22 | 0.38 | -0.06 | (-0.13 - 0.01) | 0.12 | -0.09 | (-0.26 - 0.08) | 0.29 | 0.03 | (-0.14 - 0.2) | 0.71 |
| | R | 0.14 | 0.87 | 0.92 | 0.01 | (-0.06 - | 0.73 | -0.03 | (-0.2 - 0.14) | 0.74 | 0.04 | (-0.13 - | 0.64 |

| | | | | | | | | | | | | | |
|---|---|---|---|---|---|---|---|---|---|---|---|---|---|
| | | | | | | 0.09) | | | | | | 0.21) | |
| Pericalcarine | L | 1.68 | 0.19 | 0.33 | -0.06 | (-0.14 - 0.01) | 0.09 | -0.09 | (-0.26 - 0.08) | 0.29 | 0.03 | (-0.14 - 0.2) | 0.75 |
| | R | 3.94 | 0.02 | 0.07 | **-0.11** | **(-0.18 - -0.03)** | **5.36e-03** | -0.02 | (-0.19 - 0.15) | 0.81 | -0.09 | (-0.26 - 0.09) | 0.33 |
| Postcentral | L | 0.11 | 0.89 | 0.94 | 0.00 | (-0.07 - 0.08) | 0.93 | 0.04 | (-0.13 - 0.21) | 0.64 | -0.04 | (-0.21 - 0.13) | 0.67 |
| | R | 0.67 | 0.51 | 0.66 | -0.04 | (-0.11 - 0.03) | 0.29 | 0.02 | (-0.15 - 0.19) | 0.81 | -0.06 | (-0.23 - 0.11) | 0.48 |
| Posterior Cingulate | L | 0.69 | 0.50 | 0.65 | -0.04 | (-0.12 - 0.03) | 0.25 | 0.00 | (-0.17 - 0.17) | 0.98 | -0.05 | (-0.22 - 0.12) | 0.60 |
| | R | 2.23 | 0.11 | 0.23 | -0.06 | (-0.14 - 0.01) | 0.09 | 0.07 | (-0.09 - 0.24) | 0.39 | -0.14 | (-0.31 - 0.03) | 0.11 |
| Precentral | L | 1.11 | 0.33 | 0.49 | 0.04 | (-0.03 - 0.12) | 0.26 | -0.06 | (-0.23 - 0.11) | 0.47 | 0.11 | (-0.06 - 0.28) | 0.22 |
| | R | 0.18 | 0.83 | 0.90 | 0.01 | (-0.07 - 0.08) | 0.83 | -0.04 | (-0.21 - 0.12) | 0.61 | 0.05 | (-0.12 - 0.22) | 0.55 |
| Precuneus | L | 1.46 | 0.23 | 0.38 | -0.03 | (-0.1 - 0.05) | 0.49 | 0.12 | (-0.05 - 0.29) | 0.16 | -0.15 | (-0.31 - 0.02) | 0.09 |
| | R | 0.71 | 0.49 | 0.65 | -0.04 | (-0.11 - 0.04) | 0.34 | 0.04 | (-0.13 - 0.21) | 0.62 | -0.08 | (-0.25 - 0.09) | 0.36 |
| Rostral Anterior Cingulate | L | **6.11** | **2.25e-03** | **0.02** | **-0.12** | **(-0.2 - -0.05)** | **1.05e-03** | -0.16 | (-0.33 - 0.01) | 0.06 | 0.03 | (-0.13 - 0.2) | 0.69 |
| | R | 3.13 | 0.04 | 0.12 | **-0.08** | **(-0.16 - -0.01)** | **0.03** | -0.14 | (-0.31 - 0.03) | 0.11 | 0.05 | (-0.12 - 0.22) | 0.55 |
| Rostral Middle Frontal | L | 0.23 | 0.79 | 0.86 | -0.02 | (-0.09 - 0.05) | 0.59 | -0.04 | (-0.21 - 0.12) | 0.61 | 0.02 | (-0.15 - 0.19) | 0.78 |
| | R | 0.25 | 0.77 | 0.85 | -0.03 | (-0.1 - 0.05) | 0.48 | -0.01 | (-0.18 - 0.16) | 0.92 | -0.02 | (-0.19 - 0.15) | 0.83 |
| Superior Frontal | L | 4.09 | 0.02 | 0.07 | -0.05 | (-0.12 - 0.02) | 0.19 | **-0.24** | **(-0.41 - -0.07)** | **5.92e-03** | **0.19** | **(0.02 - 0.36)** | **0.03** |
| | R | **5.23** | **5.41e-03** | **0.03** | **-0.11** | **(-0.19 - -0.04)** | **2.57e-03** | -0.15 | (-0.32 - 0.02) | 0.08 | 0.04 | (-0.13 - 0.2) | 0.68 |
| Superior Parietal | L | 0.05 | 0.95 | 0.97 | 0.01 | (-0.06 - 0.09) | 0.75 | 0.01 | (-0.16 - 0.18) | 0.92 | 0.00 | (-0.17 - 0.17) | 0.96 |
| | R | 0.08 | 0.93 | 0.96 | 0.01 | (-0.06 - 0.09) | 0.69 | 0.01 | (-0.16 - 0.18) | 0.93 | 0.01 | (-0.16 - 0.18) | 0.93 |
| Superior Temporal | L | 0.54 | 0.58 | 0.70 | -0.04 | (-0.11 - 0.03) | 0.30 | -0.02 | (-0.19 - 0.15) | 0.79 | -0.02 | (-0.19 - 0.15) | 0.85 |
| | R | 1.24 | 0.29 | 0.45 | -0.06 | (-0.13 - 0.02) | 0.13 | 0.00 | (-0.17 - 0.17) | 0.97 | -0.06 | (-0.23 - 0.11) | 0.48 |
| Supramarginal | L | **5.25** | **5.28e-03** | **0.03** | **-0.11** | **(-0.19 - -0.04)** | **3.40e-03** | 0.07 | (-0.1 - 0.24) | 0.45 | **-0.18** | **(-0.35 - -0.01)** | **0.04** |
| | R | 1.77 | 0.17 | 0.31 | -0.06 | (-0.13 - 0.02) | 0.14 | 0.07 | (-0.1 - 0.24) | 0.40 | -0.13 | (-0.3 - 0.04) | 0.14 |
| Temporal Pole | L | 0.40 | 0.67 | 0.78 | -0.03 | (-0.11 - 0.04) | 0.37 | -0.02 | (-0.18 - 0.15) | 0.86 | -0.02 | (-0.19 - 0.15) | 0.83 |
| | R | **4.89** | **7.61e-03** | **0.04** | 0.02 | (-0.05 - 0.1) | 0.52 | **-0.25** | **(-0.42 - -0.08)** | **4.24e-03** | **0.27** | **(0.1 - 0.44)** | **1.80e-03** |
| Transverse Temporal | L | 1.09 | 0.34 | 0.49 | -0.06 | (-0.13 - 0.02) | 0.14 | -0.02 | (-0.19 - 0.14) | 0.78 | -0.03 | (-0.2 - 0.14) | 0.71 |
| | R | 0.73 | 0.48 | 0.65 | -0.01 | (-0.08 - 0.07) | 0.86 | 0.10 | (-0.07 - 0.27) | 0.26 | -0.10 | (-0.27 - 0.07) | 0.23 |
| **Subcortical Volume** | | | | | | | | | | | | | |
| Amygdala | L | 1.08 | 0.34 | 0.49 | -0.05 | (-0.12 - 0.03) | 0.23 | -0.09 | (-0.26 - 0.08) | 0.28 | 0.05 | (-0.12 - 0.22) | 0.59 |

| | | | | | | | | | | | | | |
|---|---|---|---|---|---|---|---|---|---|---|---|---|---|
| | R | 4.20 | 0.02 | 0.06 | **-0.10** | **(-0.17 - -0.02)** | **8.86e-03** | -0.15 | (-0.32 - 0.02) | 0.08 | 0.05 | (-0.12 - 0.22) | 0.56 |
| Caudate | L | 0.61 | 0.55 | 0.67 | 0.02 | (-0.05 - 0.1) | 0.51 | 0.09 | (-0.08 - 0.26) | 0.32 | -0.06 | (-0.23 - 0.11) | 0.48 |
| | R | 0.23 | 0.80 | 0.86 | -0.01 | (-0.08 - 0.06) | 0.81 | 0.05 | (-0.12 - 0.22) | 0.57 | -0.06 | (-0.23 - 0.11) | 0.50 |
| Hippocampus | L | 2.02 | 0.13 | 0.27 | -0.07 | (-0.15 - 0) | 0.05 | -0.08 | (-0.25 - 0.09) | 0.37 | 0.00 | (-0.17 - 0.17) | 0.98 |
| | R | **7.41** | **6.18e-04** | **9.59e-03** | **-0.14** | **(-0.22 - -0.07)** | **1.23e-04** | -0.05 | (-0.22 - 0.12) | 0.54 | -0.09 | (-0.26 - 0.08) | 0.28 |
| Lateral Ventricles | L | **6.53** | **1.48e-03** | **0.01** | **0.13** | **(0.05 - 0.2)** | **7.29e-04** | 0.17 | (0 - 0.34) | 0.05 | -0.04 | (-0.21 - 0.13) | 0.67 |
| | R | **4.62** | **9.96e-03** | **0.05** | **0.11** | **(0.04 - 0.19)** | **2.88e-03** | 0.10 | (-0.07 - 0.27) | 0.24 | 0.01 | (-0.16 - 0.18) | 0.88 |
| Nucleus Accumbens | L | 0.57 | 0.57 | 0.69 | -0.02 | (-0.1 - 0.05) | 0.55 | 0.06 | (-0.1 - 0.23) | 0.46 | -0.09 | (-0.26 - 0.08) | 0.32 |
| | R | 0.06 | 0.94 | 0.97 | 0.00 | (-0.07 - 0.08) | 0.90 | 0.03 | (-0.14 - 0.2) | 0.73 | -0.02 | (-0.19 - 0.15) | 0.78 |
| Pallidum | L | 0.61 | 0.54 | 0.67 | -0.03 | (-0.1 - 0.05) | 0.45 | -0.08 | (-0.25 - 0.09) | 0.34 | 0.05 | (-0.12 - 0.22) | 0.54 |
| | R | 0.83 | 0.44 | 0.61 | -0.03 | (-0.1 - 0.05) | 0.46 | -0.10 | (-0.27 - 0.07) | 0.24 | 0.07 | (-0.1 - 0.24) | 0.40 |
| Putamen | L | 0.03 | 0.97 | 0.97 | 0.00 | (-0.08 - 0.07) | 0.94 | 0.02 | (-0.15 - 0.19) | 0.83 | -0.02 | (-0.19 - 0.15) | 0.81 |
| | R | 0.32 | 0.73 | 0.82 | -0.01 | (-0.08 - 0.06) | 0.77 | -0.07 | (-0.24 - 0.1) | 0.43 | 0.06 | (-0.11 - 0.23) | 0.52 |
| Thalamus | L | 2.26 | 0.10 | 0.23 | -0.06 | (-0.13 - 0.01) | 0.12 | -0.15 | (-0.32 - 0.02) | 0.09 | 0.09 | (-0.08 - 0.26) | 0.31 |
| | R | 3.73 | 0.02 | 0.09 | **-0.10** | **(-0.17 - -0.02)** | **0.01** | -0.12 | (-0.29 - 0.05) | 0.16 | 0.02 | (-0.15 - 0.19) | 0.79 |
| **Global** | | | | | | | | | | | | | |
| Estimated Intracranial Volume | N/A | **7.74** | **4.46e-04** | **7.68e-03** | **-0.15** | **(-0.22 - -0.07)** | **1.13e-04** | -0.01 | (-0.18 - 0.16) | 0.94 | -0.14 | (-0.31 - 0.03) | 0.11 |
| Mean Cortical Thickness | | **9.82** | **5.63e-05** | **1.75e-03** | **-0.17** | **(-0.24 - -0.09)** | **1.00e-05** | -0.10 | (-0.27 - 0.07) | 0.26 | -0.07 | (-0.24 - 0.1) | 0.42 |
| Total Surface Area | | **9.88** | **5.30e-05** | **1.75e-03** | **-0.17** | **(-0.24 - -0.09)** | **9.54e-06** | -0.05 | (-0.22 - 0.11) | 0.54 | -0.12 | (-0.28 - 0.05) | 0.18 |

*Results marked in bold are significant at q<0.05 (the overall effect) or at p<0.05 (pairwise comparisons).*

**eTable 14.** BIPS subgroup (BIPS vs. no BIPS assignment vs. HC) effect size overview

| | | Overall Test Statistics | | | BIPS < HC | | | BIPS < no BIPS | | | no BIPS < HC | | |
|---|---|---|---|---|---|---|---|---|---|---|---|---|---|
| **Region of Interest** | **Hemisphere** | **F** | **p** | **q** | **Cohen's d** | **95% Confidence Interval** | **p** | **Cohen's d** | **95% Confidence Interval** | **p** | **Cohen's d** | **95% Confidence Interval** | **p** |
| **Cortical Thickness** | | | | | | | | | | | | | |
| Banks of the Superior Temporal Sulcus | L | 2.52 | 0.08 | 0.19 | -0.04 | (-0.26 - 0.17) | 0.69 | 0.04 | (-0.17 - 0.26) | 0.70 | **-0.08** | **(-0.16 - -0.01)** | **0.02** |
| | R | **7.01** | **9.19e-04** | **0.01** | -0.02 | (-0.23 - 0.19) | 0.87 | 0.12 | (-0.09 - 0.33) | 0.26 | **-0.14** | **(-0.21 - -0.07)** | **2.15e-04** |
| Caudal Anterior Cingulate | L | 2.54 | 0.08 | 0.19 | -0.20 | (-0.42 - 0.01) | 0.06 | -0.14 | (-0.36 - 0.07) | 0.19 | -0.06 | (-0.13 - 0.01) | 0.11 |
| | R | 1.82 | 0.16 | 0.29 | -0.15 | (-0.36 - 0.06) | 0.17 | -0.09 | (-0.3 - 0.12) | 0.41 | -0.06 | (-0.13 - 0.01) | 0.12 |
| Caudal Middle Frontal | L | 3.91 | 0.02 | 0.08 | -0.18 | (-0.39 - 0.04) | 0.10 | -0.08 | (-0.3 - 0.13) | 0.45 | **-0.09** | **(-0.17 - -0.02)** | **0.01** |
| | R | 4.79 | 8.35e-03 | 0.05 | -0.02 | (-0.24 - 0.19) | 0.83 | 0.09 | (-0.12 - 0.31) | 0.39 | **-0.11** | **(-0.19 - -0.04)** | **2.13e-03** |
| Cuneus | L | 2.67 | 0.07 | 0.18 | **0.26** | **(0.04 - 0.48)** | **0.02** | **0.24** | **(0.03 - 0.46)** | **0.03** | 0.01 | (-0.06 - 0.09) | 0.74 |
| | R | 1.98 | 0.14 | 0.26 | 0.15 | (-0.06 - 0.36) | 0.17 | 0.20 | (-0.02 - 0.41) | 0.07 | -0.04 | (-0.12 - 0.03) | 0.25 |
| Entorhinal | L | 3.49 | 0.03 | 0.11 | **-0.28** | **(-0.5 - -0.07)** | **9.04e-03** | **-0.28** | **(-0.49 - -0.07)** | **9.76e-03** | 0.00 | (-0.08 - 0.07) | 0.92 |
| | R | 3.03 | 0.05 | 0.14 | **-0.26** | **(-0.47 - -0.05)** | **0.02** | **-0.22** | **(-0.43 - -0.01)** | **0.04** | -0.04 | (-0.11 - 0.03) | 0.29 |
| Frontal Pole | L | 1.31 | 0.27 | 0.41 | 0.10 | (-0.11 - 0.31) | 0.36 | 0.14 | (-0.07 - 0.35) | 0.19 | -0.04 | (-0.12 - 0.03) | 0.26 |
| | R | 2.35 | 0.10 | 0.21 | -0.18 | (-0.39 - 0.04) | 0.11 | -0.11 | (-0.32 - 0.1) | 0.31 | -0.06 | (-0.14 - 0.01) | 0.08 |
| Fusiform | L | **8.60** | **1.89e-04** | **4.88e-03** | -0.17 | (-0.38 - 0.04) | 0.11 | -0.02 | (-0.23 - 0.19) | 0.87 | **-0.15** | **(-0.22 - -0.08)** | **5.03e-05** |
| | R | 3.33 | 0.04 | 0.12 | -0.02 | (-0.23 - 0.2) | 0.88 | 0.08 | (-0.13 - 0.29) | 0.46 | **-0.10** | **(-0.17 - -0.02)** | **0.01** |
| Inferior Parietal | L | **9.26** | **9.78e-05** | **3.03e-03** | **-0.25** | **(-0.46 - -0.04)** | **0.02** | -0.10 | (-0.31 - 0.12) | 0.38 | **-0.15** | **(-0.22 - -0.08)** | **6.27e-05** |
| | R | 4.26 | 0.01 | 0.07 | -0.04 | (-0.25 - 0.18) | 0.74 | 0.07 | (-0.14 - 0.29) | 0.49 | **-0.11** | **(-0.18 - -0.04)** | **3.62e-03** |
| Inferior Temporal | L | 4.14 | 0.02 | 0.07 | -0.09 | (-0.3 - 0.12) | 0.41 | 0.02 | (-0.19 - 0.23) | 0.86 | **-0.11** | **(-0.18 - -0.03)** | **4.23e-03** |
| | R | **6.63** | **1.34e-03** | **0.02** | -0.07 | (-0.29 - 0.14) | 0.50 | 0.06 | (-0.15 - 0.28) | 0.56 | **-0.14** | **(-0.21 - -0.06)** | **2.75e-04** |
| Insula | L | **9.49** | **7.82e-05** | **3.03e-03** | -0.08 | (-0.29 - 0.13) | 0.47 | 0.09 | (-0.13 - 0.3) | 0.42 | **-0.16** | **(-0.24 - -0.09)** | **1.38e-05** |
| | R | **7.78** | **4.25e-04** | **7.57e-03** | **-0.25** | **(-0.47 - -0.04)** | **0.02** | -0.12 | (-0.33 - 0.09) | 0.27 | **-0.13** | **(-0.21 - -0.06)** | **3.77e-04** |
| Isthmus Cingulate | L | 1.52 | 0.22 | 0.36 | -0.19 | (-0.4 - 0.02) | 0.08 | -0.18 | (-0.4 - 0.03) | 0.09 | -0.01 | (-0.08 - 0.07) | 0.86 |
| | R | 0.93 | 0.39 | 0.53 | -0.04 | (-0.25 - 0.18) | 0.74 | 0.02 | (-0.2 - 0.23) | 0.88 | -0.05 | (-0.12 - 0.02) | 0.17 |
| Lateral Occipital | L | 3.78 | 0.02 | 0.09 | -0.11 | (-0.32 - 0.11) | 0.33 | 0.00 | (-0.22 - 0.21) | 0.97 | **-0.10** | **(-0.17 - -0.03)** | **6.85e-03** |
| | R | **7.82** | **4.11e-04** | **7.57e-03** | -0.15 | (-0.36 - 0.07) | 0.19 | 0.00 | (-0.21 - 0.22) | 0.98 | **-0.15** | **(-0.22 - -0.07)** | **9.61e-05** |
| | | | | | | (-0.16 - | | | (-0.09 - | | | | |

| | | | | | | | | | | | | | |
|---|---|---|---|---|---|---|---|---|---|---|---|---|---|
| Lateral Orbitofrontal | L | 2.04 | 0.13 | 0.25 | 0.05 | 0.26) | 0.64 | 0.12 | 0.33) | 0.27 | -0.07 | (-0.14 - 0) | 0.06 |
| | R | 3.01 | 0.05 | 0.14 | 0.09 | (-0.12 - 0.3) | 0.41 | 0.17 | (-0.04 - 0.38) | 0.12 | **-0.08** | **(-0.15 - -0.01)** | **0.03** |
| Lingual | L | 0.87 | 0.42 | 0.55 | 0.11 | (-0.11 - 0.33) | 0.32 | 0.14 | (-0.08 - 0.35) | 0.22 | -0.03 | (-0.1 - 0.05) | 0.50 |
| | R | 0.04 | 0.96 | 0.96 | 0.02 | (-0.19 - 0.24) | 0.84 | 0.01 | (-0.2 - 0.23) | 0.91 | 0.01 | (-0.06 - 0.08) | 0.81 |
| Medial Orbitofrontal | L | **5.84** | **2.95e-03** | **0.03** | 0.03 | (-0.18 - 0.25) | 0.77 | 0.16 | (-0.06 - 0.37) | 0.15 | **-0.12** | **(-0.2 - -0.05)** | **9.98e-04** |
| | R | 3.16 | 0.04 | 0.13 | 0.00 | (-0.21 - 0.22) | 0.99 | 0.10 | (-0.12 - 0.31) | 0.38 | **-0.09** | **(-0.17 - -0.02)** | **0.01** |
| Middle Temporal | L | 2.75 | 0.06 | 0.17 | -0.10 | (-0.31 - 0.12) | 0.38 | -0.01 | (-0.22 - 0.21) | 0.94 | **-0.09** | **(-0.16 - -0.01)** | **0.02** |
| | R | **11.32** | **1.26e-05** | **1.96e-03** | -0.20 | (-0.42 - 0.01) | 0.07 | -0.02 | (-0.24 - 0.19) | 0.82 | **-0.17** | **(-0.25 - -0.1)** | **3.43e-06** |
| Paracentral | L | **7.00** | **9.28e-04** | **0.01** | -0.05 | (-0.27 - 0.16) | 0.63 | 0.09 | (-0.13 - 0.3) | 0.42 | **-0.14** | **(-0.21 - -0.07)** | **1.91e-04** |
| | R | 3.10 | 0.05 | 0.13 | -0.02 | (-0.24 - 0.19) | 0.82 | 0.07 | (-0.14 - 0.28) | 0.52 | **-0.09** | **(-0.17 - -0.02)** | **0.01** |
| Parahippocampal | L | **5.06** | **6.42e-03** | **0.05** | -0.09 | (-0.3 - 0.13) | 0.43 | 0.03 | (-0.18 - 0.25) | 0.75 | **-0.12** | **(-0.19 - -0.05)** | **1.52e-03** |
| | R | 1.39 | 0.25 | 0.39 | -0.01 | (-0.22 - 0.21) | 0.96 | 0.06 | (-0.16 - 0.27) | 0.60 | -0.06 | (-0.13 - 0.01) | 0.10 |
| Pars Opercularis | L | 2.02 | 0.13 | 0.25 | 0.15 | (-0.07 - 0.36) | 0.18 | 0.19 | (-0.02 - 0.41) | 0.08 | -0.05 | (-0.12 - 0.03) | 0.23 |
| | R | 1.30 | 0.27 | 0.41 | -0.09 | (-0.3 - 0.13) | 0.42 | -0.03 | (-0.24 - 0.18) | 0.78 | -0.06 | (-0.13 - 0.02) | 0.13 |
| Pars Orbitalis | L | 0.65 | 0.52 | 0.65 | -0.06 | (-0.27 - 0.16) | 0.61 | -0.01 | (-0.23 - 0.2) | 0.90 | -0.04 | (-0.11 - 0.03) | 0.27 |
| | R | 0.09 | 0.92 | 0.94 | 0.04 | (-0.17 - 0.26) | 0.70 | 0.03 | (-0.18 - 0.25) | 0.75 | 0.01 | (-0.07 - 0.08) | 0.83 |
| Pars Triangularis | L | 0.33 | 0.72 | 0.80 | 0.04 | (-0.17 - 0.26) | 0.69 | 0.01 | (-0.2 - 0.23) | 0.90 | 0.03 | (-0.04 - 0.1) | 0.44 |
| | R | 0.83 | 0.43 | 0.56 | 0.02 | (-0.19 - 0.24) | 0.83 | 0.07 | (-0.14 - 0.28) | 0.52 | -0.05 | (-0.12 - 0.03) | 0.23 |
| Pericalcarine | L | 2.45 | 0.09 | 0.20 | **0.25** | **(0.03 - 0.47)** | **0.03** | **0.23** | **(0.01 - 0.45)** | **0.04** | 0.02 | (-0.05 - 0.09) | 0.58 |
| | R | 1.59 | 0.20 | 0.35 | 0.08 | (-0.14 - 0.3) | 0.46 | 0.01 | (-0.2 - 0.23) | 0.89 | 0.07 | (-0.01 - 0.14) | 0.08 |
| Postcentral | L | 2.74 | 0.07 | 0.17 | -0.02 | (-0.24 - 0.19) | 0.84 | 0.07 | (-0.15 - 0.28) | 0.54 | **-0.09** | **(-0.16 - -0.01)** | **0.02** |
| | R | 4.50 | 0.01 | 0.06 | -0.11 | (-0.33 - 0.11) | 0.32 | 0.00 | (-0.22 - 0.22) | 0.99 | **-0.11** | **(-0.18 - -0.04)** | **3.05e-03** |
| Posterior Cingulate | L | 3.29 | 0.04 | 0.13 | -0.17 | (-0.38 - 0.05) | 0.13 | -0.08 | (-0.29 - 0.13) | 0.47 | **-0.09** | **(-0.16 - -0.01)** | **0.02** |
| | R | **5.38** | **4.67e-03** | **0.04** | -0.06 | (-0.27 - 0.15) | 0.60 | 0.07 | (-0.14 - 0.28) | 0.53 | **-0.12** | **(-0.2 - -0.05)** | **1.06e-03** |
| Precentral | L | 4.22 | 0.01 | 0.07 | -0.11 | (-0.33 - 0.1) | 0.30 | 0.00 | (-0.22 - 0.21) | 0.97 | **-0.11** | **(-0.18 - -0.03)** | **4.25e-03** |
| | R | 4.48 | 0.01 | 0.06 | -0.08 | (-0.29 - 0.13) | 0.47 | 0.03 | (-0.18 - 0.25) | 0.76 | **-0.11** | **(-0.19 - -0.04)** | **2.82e-03** |
| Precuneus | L | 3.52 | 0.03 | 0.11 | -0.02 | (-0.23 - 0.19) | 0.86 | 0.08 | (-0.13 - 0.29) | 0.46 | **-0.10** | **(-0.17 - -0.03)** | **8.51e-03** |
| | R | **6.31** | **1.84e-03** | **0.02** | -0.05 | (-0.26 - 0.16) | 0.65 | 0.09 | (-0.13 - 0.3) | 0.43 | **-0.13** | **(-0.21 - -0.06)** | **3.97e-04** |
| Rostral Anterior | L | 3.27 | 0.04 | 0.13 | -0.12 | (-0.34 - 0.09) | 0.26 | -0.03 | (-0.24 - 0.18) | 0.79 | **-0.09** | **(-0.17 - -0.02)** | **0.01** |

| | | | | | | | | | | | | | |
|---|---|---|---|---|---|---|---|---|---|---|---|---|---|
| Cingulate | R | 2.46 | 0.09 | 0.20 | 0.05 | (-0.16 - 0.27) | 0.62 | 0.13 | (-0.08 - 0.34) | 0.23 | **-0.08** | **(-0.15 - 0)** | **0.04** |
| Rostral Middle Frontal | L | **4.94** | **7.20e-03** | **0.05** | -0.10 | (-0.31 - 0.12) | 0.37 | 0.02 | (-0.19 - 0.23) | 0.85 | **-0.12** | **(-0.19 - -0.04)** | **1.78e-03** |
| | R | 4.11 | 0.02 | 0.07 | -0.08 | (-0.3 - 0.13) | 0.44 | 0.02 | (-0.19 - 0.24) | 0.82 | **-0.11** | **(-0.18 - -0.03)** | **4.29e-03** |
| Superior Frontal | L | 3.13 | 0.04 | 0.13 | -0.09 | (-0.3 - 0.12) | 0.41 | 0.00 | (-0.21 - 0.22) | 0.97 | **-0.09** | **(-0.17 - -0.02)** | **0.01** |
| | R | 2.02 | 0.13 | 0.25 | -0.07 | (-0.28 - 0.14) | 0.52 | 0.01 | (-0.21 - 0.22) | 0.96 | **-0.07** | **(-0.15 - 0)** | **0.05** |
| Superior Parietal | L | 3.33 | 0.04 | 0.12 | 0.00 | (-0.21 - 0.22) | 0.97 | 0.10 | (-0.11 - 0.31) | 0.36 | **-0.09** | **(-0.17 - -0.02)** | **0.01** |
| | R | 4.26 | 0.01 | 0.07 | -0.06 | (-0.27 - 0.16) | 0.60 | 0.05 | (-0.16 - 0.27) | 0.61 | **-0.11** | **(-0.18 - -0.04)** | **3.55e-03** |
| Superior Temporal | L | 4.12 | 0.02 | 0.07 | -0.11 | (-0.33 - 0.1) | 0.31 | 0.00 | (-0.22 - 0.21) | 0.97 | **-0.11** | **(-0.18 - -0.03)** | **4.73e-03** |
| | R | **6.60** | **1.38e-03** | **0.02** | -0.13 | (-0.34 - 0.09) | 0.25 | 0.01 | (-0.21 - 0.23) | 0.93 | **-0.13** | **(-0.21 - -0.06)** | **3.25e-04** |
| Supramarginal | L | 2.95 | 0.05 | 0.15 | -0.10 | (-0.31 - 0.11) | 0.36 | -0.01 | (-0.22 - 0.2) | 0.94 | **-0.09** | **(-0.16 - -0.02)** | **0.02** |
| | R | 2.50 | 0.08 | 0.19 | -0.11 | (-0.32 - 0.11) | 0.34 | -0.02 | (-0.24 - 0.19) | 0.84 | **-0.08** | **(-0.15 - -0.01)** | **0.03** |
| Temporal Pole | L | 1.37 | 0.25 | 0.39 | -0.14 | (-0.35 - 0.07) | 0.19 | -0.09 | (-0.31 - 0.12) | 0.39 | -0.05 | (-0.12 - 0.03) | 0.21 |
| | R | 2.15 | 0.12 | 0.24 | -0.10 | (-0.31 - 0.12) | 0.38 | -0.02 | (-0.23 - 0.19) | 0.86 | **-0.08** | **(-0.15 - 0)** | **0.04** |
| Transverse Temporal | L | 1.38 | 0.25 | 0.39 | -0.11 | (-0.33 - 0.1) | 0.30 | -0.06 | (-0.27 - 0.16) | 0.60 | -0.05 | (-0.13 - 0.02) | 0.14 |
| | R | 4.37 | 0.01 | 0.07 | **-0.26** | **(-0.47 - -0.04)** | **0.02** | -0.17 | (-0.39 - 0.04) | 0.11 | **-0.08** | **(-0.16 - -0.01)** | **0.03** |
| **Surface Area** | | | | | | | | | | | | | |
| Banks of the Superior Temporal Sulcus | L | 1.80 | 0.17 | 0.29 | -0.07 | (-0.28 - 0.15) | 0.54 | 0.00 | (-0.21 - 0.22) | 0.97 | -0.07 | (-0.14 - 0) | 0.06 |
| | R | 2.63 | 0.07 | 0.18 | 0.02 | (-0.19 - 0.24) | 0.82 | 0.11 | (-0.1 - 0.32) | 0.32 | **-0.08** | **(-0.16 - -0.01)** | **0.03** |
| Caudal Anterior Cingulate | L | 0.30 | 0.74 | 0.81 | 0.06 | (-0.16 - 0.27) | 0.59 | 0.08 | (-0.14 - 0.29) | 0.48 | -0.02 | (-0.09 - 0.06) | 0.65 |
| | R | 2.51 | 0.08 | 0.19 | -0.03 | (-0.25 - 0.18) | 0.75 | 0.05 | (-0.16 - 0.26) | 0.64 | **-0.08** | **(-0.16 - -0.01)** | **0.03** |
| Caudal Middle Frontal | L | 1.04 | 0.35 | 0.48 | -0.04 | (-0.25 - 0.18) | 0.74 | 0.02 | (-0.19 - 0.23) | 0.86 | -0.05 | (-0.13 - 0.02) | 0.15 |
| | R | 2.21 | 0.11 | 0.23 | 0.18 | (-0.03 - 0.4) | 0.10 | **0.22** | **(0.01 - 0.43)** | **0.05** | -0.04 | (-0.11 - 0.04) | 0.34 |
| Cuneus | L | 1.18 | 0.31 | 0.43 | -0.06 | (-0.28 - 0.16) | 0.60 | 0.00 | (-0.22 - 0.22) | 0.99 | -0.06 | (-0.13 - 0.02) | 0.13 |
| | R | 2.67 | 0.07 | 0.18 | -0.11 | (-0.33 - 0.1) | 0.31 | -0.03 | (-0.24 - 0.19) | 0.80 | **-0.08** | **(-0.16 - -0.01)** | **0.03** |
| Entorhinal | L | 3.37 | 0.03 | 0.12 | -0.12 | (-0.33 - 0.1) | 0.29 | -0.02 | (-0.23 - 0.19) | 0.85 | **-0.09** | **(-0.17 - -0.02)** | **0.01** |
| | R | 0.42 | 0.65 | 0.74 | 0.04 | (-0.17 - 0.25) | 0.70 | 0.07 | (-0.14 - 0.28) | 0.52 | -0.03 | (-0.1 - 0.04) | 0.45 |
| Frontal Pole | L | 0.44 | 0.64 | 0.74 | 0.05 | (-0.16 - 0.27) | 0.62 | 0.02 | (-0.19 - 0.23) | 0.85 | 0.03 | (-0.04 - 0.11) | 0.38 |
| | R | 0.73 | 0.48 | 0.61 | 0.07 | (-0.14 - 0.28) | 0.52 | 0.03 | (-0.19 - 0.24) | 0.81 | 0.04 | (-0.03 - 0.12) | 0.26 |
| Fusiform | L | 1.42 | 0.24 | 0.39 | 0.06 | (-0.15 - 0.28) | 0.58 | 0.12 | (-0.1 - 0.33) | 0.28 | -0.05 | (-0.13 - 0.02) | 0.14 |

| | | | | | | | | | | | | | |
|---|---|---|---|---|---|---|---|---|---|---|---|---|---|
| | R | 4.55 | 0.01 | 0.06 | 0.16 | (-0.05 - 0.38) | 0.14 | **0.25** | **(0.04 - 0.46)** | **0.02** | **-0.09** | **(-0.16 - -0.01)** | **0.02** |
| Inferior Parietal | L | 1.86 | 0.16 | 0.28 | 0.06 | (-0.15 - 0.27) | 0.58 | 0.13 | (-0.09 - 0.34) | 0.25 | -0.06 | (-0.14 - 0.01) | 0.09 |
| | R | 4.41 | 0.01 | 0.07 | 0.02 | (-0.19 - 0.24) | 0.83 | 0.13 | (-0.08 - 0.34) | 0.22 | **-0.11** | **(-0.18 - -0.03)** | **4.13e-03** |
| Inferior Temporal | L | 2.35 | 0.10 | 0.21 | 0.17 | (-0.04 - 0.39) | 0.11 | **0.22** | **(0 - 0.43)** | **0.05** | -0.04 | (-0.12 - 0.03) | 0.24 |
| | R | 2.05 | 0.13 | 0.25 | 0.17 | (-0.04 - 0.38) | 0.11 | 0.21 | (0 - 0.42) | 0.05 | -0.03 | (-0.11 - 0.04) | 0.35 |
| Insula | L | 2.02 | 0.13 | 0.25 | -0.17 | (-0.39 - 0.04) | 0.12 | -0.11 | (-0.33 - 0.1) | 0.30 | -0.06 | (-0.13 - 0.02) | 0.13 |
| | R | 0.75 | 0.47 | 0.61 | -0.04 | (-0.26 - 0.17) | 0.68 | 0.00 | (-0.21 - 0.21) | 0.99 | -0.04 | (-0.12 - 0.03) | 0.23 |
| Isthmus Cingulate | L | 0.64 | 0.53 | 0.65 | -0.12 | (-0.34 - 0.09) | 0.26 | -0.12 | (-0.34 - 0.09) | 0.26 | 0.00 | (-0.07 - 0.07) | 0.98 |
| | R | 2.96 | 0.05 | 0.15 | 0.14 | (-0.07 - 0.35) | 0.20 | 0.05 | (-0.16 - 0.27) | 0.63 | **0.08** | **(0.01 - 0.16)** | **0.02** |
| Lateral Occipital | L | 1.95 | 0.14 | 0.26 | 0.20 | (-0.02 - 0.41) | 0.07 | **0.21** | **(0 - 0.43)** | **0.05** | -0.02 | (-0.09 - 0.05) | 0.62 |
| | R | 0.69 | 0.50 | 0.63 | 0.10 | (-0.12 - 0.31) | 0.37 | 0.12 | (-0.09 - 0.33) | 0.27 | -0.02 | (-0.09 - 0.05) | 0.56 |
| Lateral Orbitofrontal | L | 0.27 | 0.77 | 0.82 | -0.06 | (-0.28 - 0.15) | 0.57 | -0.04 | (-0.25 - 0.17) | 0.71 | -0.02 | (-0.09 - 0.05) | 0.58 |
| | R | 0.43 | 0.65 | 0.74 | -0.08 | (-0.29 - 0.13) | 0.47 | -0.05 | (-0.27 - 0.16) | 0.62 | -0.03 | (-0.1 - 0.05) | 0.49 |
| Lingual | L | 3.15 | 0.04 | 0.13 | **-0.26** | **(-0.47 - -0.04)** | **0.02** | -0.21 | (-0.42 - 0.01) | 0.06 | -0.05 | (-0.12 - 0.02) | 0.17 |
| | R | 0.29 | 0.75 | 0.81 | -0.08 | (-0.29 - 0.14) | 0.49 | -0.06 | (-0.27 - 0.16) | 0.60 | -0.02 | (-0.09 - 0.06) | 0.65 |
| Medial Orbitofrontal | L | 0.17 | 0.85 | 0.88 | -0.02 | (-0.23 - 0.2) | 0.87 | 0.00 | (-0.21 - 0.22) | 0.97 | -0.02 | (-0.09 - 0.05) | 0.57 |
| | R | 2.92 | 0.05 | 0.15 | -0.08 | (-0.29 - 0.13) | 0.47 | 0.01 | (-0.2 - 0.22) | 0.91 | **-0.09** | **(-0.16 - -0.02)** | **0.02** |
| Middle Temporal | L | 1.17 | 0.31 | 0.43 | 0.17 | (-0.05 - 0.38) | 0.13 | 0.16 | (-0.05 - 0.38) | 0.13 | 0.00 | (-0.07 - 0.08) | 0.95 |
| | R | 2.14 | 0.12 | 0.24 | 0.00 | (-0.21 - 0.21) | 0.99 | 0.08 | (-0.13 - 0.29) | 0.47 | **-0.08** | **(-0.15 - 0)** | **0.04** |
| Paracentral | L | 1.27 | 0.28 | 0.41 | 0.15 | (-0.06 - 0.36) | 0.17 | 0.17 | (-0.04 - 0.38) | 0.12 | -0.02 | (-0.09 - 0.05) | 0.61 |
| | R | 0.05 | 0.95 | 0.95 | 0.02 | (-0.19 - 0.24) | 0.82 | 0.01 | (-0.2 - 0.23) | 0.90 | 0.01 | (-0.06 - 0.08) | 0.78 |
| Parahippocampal | L | 1.29 | 0.28 | 0.41 | -0.14 | (-0.35 - 0.07) | 0.20 | -0.09 | (-0.31 - 0.12) | 0.39 | -0.04 | (-0.12 - 0.03) | 0.23 |
| | R | 1.31 | 0.27 | 0.41 | 0.09 | (-0.13 - 0.3) | 0.42 | 0.03 | (-0.18 - 0.24) | 0.78 | 0.06 | (-0.02 - 0.13) | 0.13 |
| Pars Opercularis | L | 2.36 | 0.10 | 0.21 | -0.12 | (-0.33 - 0.1) | 0.29 | -0.04 | (-0.25 - 0.17) | 0.72 | **-0.08** | **(-0.15 - 0)** | **0.04** |
| | R | 0.50 | 0.60 | 0.72 | 0.10 | (-0.12 - 0.31) | 0.38 | 0.11 | (-0.1 - 0.32) | 0.32 | -0.01 | (-0.08 - 0.06) | 0.76 |
| Pars Orbitalis | L | 0.31 | 0.73 | 0.81 | 0.08 | (-0.13 - 0.29) | 0.46 | 0.06 | (-0.15 - 0.28) | 0.56 | 0.02 | (-0.06 - 0.09) | 0.67 |
| | R | 0.31 | 0.73 | 0.81 | 0.00 | (-0.21 - 0.22) | 0.98 | -0.03 | (-0.24 - 0.19) | 0.81 | 0.03 | (-0.04 - 0.1) | 0.43 |
| Pars Triangularis | L | 1.00 | 0.37 | 0.49 | -0.09 | (-0.3 - 0.12) | 0.42 | -0.04 | (-0.25 - 0.17) | 0.72 | -0.05 | (-0.12 - 0.02) | 0.20 |
| | R | 0.23 | 0.79 | 0.84 | 0.07 | (-0.14 - | 0.52 | 0.06 | (-0.15 - | 0.59 | 0.01 | (-0.06 - | 0.75 |

| | | | | | | | | | | | | | |
|---|---|---|---|---|---|---|---|---|---|---|---|---|---|
| | | | | | | 0.28) | | | 0.27) | | | 0.09) | |
| Pericalcarine | L | 1.40 | 0.25 | 0.39 | -0.14 | (-0.36 - 0.08) | 0.23 | -0.08 | (-0.3 - 0.13) | 0.45 | -0.05 | (-0.12 - 0.02) | 0.18 |
| | R | **4.98** | **6.95e-03** | **0.05** | **-0.26** | **(-0.48 - -0.04)** | **0.02** | -0.16 | (-0.38 - 0.06) | 0.14 | **-0.09** | **(-0.17 - -0.02)** | **0.01** |
| Postcentral | L | 0.07 | 0.93 | 0.95 | 0.04 | (-0.18 - 0.25) | 0.73 | 0.04 | (-0.17 - 0.25) | 0.71 | 0.00 | (-0.08 - 0.07) | 0.94 |
| | R | 1.20 | 0.30 | 0.43 | -0.15 | (-0.37 - 0.06) | 0.17 | -0.12 | (-0.33 - 0.1) | 0.29 | -0.04 | (-0.11 - 0.04) | 0.34 |
| Posterior Cingulate | L | 0.86 | 0.42 | 0.55 | 0.02 | (-0.2 - 0.23) | 0.89 | 0.06 | (-0.15 - 0.27) | 0.56 | -0.05 | (-0.12 - 0.03) | 0.21 |
| | R | 1.86 | 0.16 | 0.28 | -0.08 | (-0.3 - 0.13) | 0.44 | -0.01 | (-0.22 - 0.2) | 0.91 | -0.07 | (-0.14 - 0) | 0.06 |
| Precentral | L | 1.28 | 0.28 | 0.41 | 0.14 | (-0.07 - 0.36) | 0.19 | 0.10 | (-0.11 - 0.31) | 0.35 | 0.04 | (-0.03 - 0.12) | 0.26 |
| | R | 0.05 | 0.95 | 0.95 | 0.01 | (-0.21 - 0.22) | 0.96 | -0.01 | (-0.22 - 0.21) | 0.95 | 0.01 | (-0.06 - 0.09) | 0.74 |
| Precuneus | L | 0.50 | 0.61 | 0.72 | -0.02 | (-0.24 - 0.19) | 0.83 | 0.02 | (-0.2 - 0.23) | 0.89 | -0.04 | (-0.11 - 0.04) | 0.32 |
| | R | 1.10 | 0.33 | 0.46 | -0.15 | (-0.36 - 0.07) | 0.19 | -0.11 | (-0.33 - 0.1) | 0.31 | -0.03 | (-0.11 - 0.04) | 0.36 |
| Rostral Anterior Cingulate | L | **5.43** | **4.40e-03** | **0.04** | 0.04 | (-0.18 - 0.25) | 0.73 | 0.16 | (-0.06 - 0.37) | 0.15 | **-0.12** | **(-0.19 - -0.04)** | **1.60e-03** |
| | R | 2.22 | 0.11 | 0.23 | 0.02 | (-0.2 - 0.23) | 0.89 | 0.09 | (-0.12 - 0.31) | 0.40 | **-0.08** | **(-0.15 - 0)** | **0.04** |
| Rostral Middle Frontal | L | 0.88 | 0.41 | 0.55 | 0.11 | (-0.1 - 0.32) | 0.31 | 0.14 | (-0.08 - 0.35) | 0.21 | -0.02 | (-0.1 - 0.05) | 0.52 |
| | R | 1.74 | 0.18 | 0.31 | 0.15 | (-0.06 - 0.36) | 0.17 | 0.19 | (-0.02 - 0.4) | 0.08 | -0.04 | (-0.11 - 0.04) | 0.33 |
| Superior Frontal | L | 2.23 | 0.11 | 0.23 | 0.17 | (-0.04 - 0.38) | 0.11 | **0.21** | **(0 - 0.42)** | **0.05** | -0.04 | (-0.11 - 0.03) | 0.28 |
| | R | 3.94 | 0.02 | 0.08 | -0.03 | (-0.24 - 0.18) | 0.79 | 0.08 | (-0.13 - 0.29) | 0.47 | **-0.10** | **(-0.18 - -0.03)** | **5.20e-03** |
| Superior Parietal | L | 0.18 | 0.84 | 0.88 | -0.04 | (-0.25 - 0.17) | 0.71 | -0.06 | (-0.27 - 0.16) | 0.61 | 0.01 | (-0.06 - 0.09) | 0.70 |
| | R | 1.50 | 0.22 | 0.37 | 0.19 | (-0.03 - 0.4) | 0.09 | 0.18 | (-0.03 - 0.4) | 0.09 | 0.00 | (-0.07 - 0.08) | 0.92 |
| Superior Temporal | L | 0.61 | 0.54 | 0.66 | -0.09 | (-0.3 - 0.13) | 0.44 | -0.05 | (-0.27 - 0.16) | 0.65 | -0.03 | (-0.11 - 0.04) | 0.36 |
| | R | 1.33 | 0.26 | 0.41 | -0.02 | (-0.23 - 0.2) | 0.89 | 0.05 | (-0.16 - 0.26) | 0.67 | -0.06 | (-0.13 - 0.01) | 0.10 |
| Supramarginal | L | **5.38** | **4.64e-03** | **0.04** | -0.21 | (-0.43 - 0) | 0.05 | -0.10 | (-0.32 - 0.11) | 0.36 | **-0.11** | **(-0.18 - -0.04)** | **3.08e-03** |
| | R | 1.56 | 0.21 | 0.35 | -0.01 | (-0.22 - 0.21) | 0.95 | 0.06 | (-0.15 - 0.27) | 0.59 | -0.07 | (-0.14 - 0.01) | 0.08 |
| Temporal Pole | L | 0.51 | 0.60 | 0.72 | -0.08 | (-0.3 - 0.13) | 0.44 | -0.05 | (-0.27 - 0.16) | 0.62 | -0.03 | (-0.1 - 0.04) | 0.44 |
| | R | 1.26 | 0.28 | 0.41 | 0.15 | (-0.07 - 0.36) | 0.18 | 0.10 | (-0.11 - 0.32) | 0.33 | 0.04 | (-0.03 - 0.11) | 0.28 |
| Transverse Temporal | L | 1.07 | 0.34 | 0.47 | -0.07 | (-0.29 - 0.14) | 0.50 | -0.02 | (-0.23 - 0.19) | 0.85 | -0.05 | (-0.13 - 0.02) | 0.16 |
| | R | 0.21 | 0.81 | 0.85 | 0.04 | (-0.18 - 0.25) | 0.75 | 0.05 | (-0.16 - 0.27) | 0.62 | -0.02 | (-0.09 - 0.05) | 0.62 |
| **Subcortical Volume** | | | | | | | | | | | | | |
| Amygdala | L | 0.51 | 0.60 | 0.72 | -0.06 | (-0.27 - 0.16) | 0.62 | -0.02 | (-0.23 - 0.2) | 0.86 | -0.04 | (-0.11 - 0.04) | 0.34 |
| | R | 3.18 | 0.04 | 0.13 | 0.02 | (-0.2 - 0.23) | 0.87 | 0.11 | (-0.1 - 0.32) | 0.31 | **-0.09** | **(-0.16 -** | **0.01** |

| | | | | | | | | | | | | | |
|---|---|---|---|---|---|---|---|---|---|---|---|---|---|
| | | | | | | | | | | | | **-0.02)** | |
| Caudate | L | 0.48 | 0.62 | 0.72 | -0.07 | (-0.29 - 0.14) | 0.52 | -0.09 | (-0.31 - 0.12) | 0.39 | 0.02 | (-0.05 - 0.1) | 0.55 |
| | R | 0.48 | 0.62 | 0.72 | -0.11 | (-0.32 - 0.11) | 0.33 | -0.10 | (-0.32 - 0.11) | 0.36 | -0.01 | (-0.08 - 0.07) | 0.83 |
| Hippocampus | L | 2.03 | 0.13 | 0.25 | -0.16 | (-0.37 - 0.05) | 0.14 | -0.10 | (-0.31 - 0.11) | 0.36 | -0.06 | (-0.13 - 0.01) | 0.10 |
| | R | **7.27** | **7.06e-04** | **0.01** | -0.18 | (-0.39 - 0.04) | 0.11 | -0.04 | (-0.25 - 0.18) | 0.74 | **-0.14** | **(-0.21 - -0.06)** | **2.25e-04** |
| Lateral Ventricles | L | 4.66 | 9.51e-03 | 0.06 | 0.13 | (-0.08 - 0.34) | 0.24 | 0.02 | (-0.2 - 0.23) | 0.88 | **0.11** | **(0.04 - 0.19)** | **2.86e-03** |
| | R | **5.35** | **4.78e-03** | **0.04** | -0.07 | (-0.29 - 0.15) | 0.52 | -0.19 | (-0.4 - 0.03) | 0.09 | **0.11** | **(0.04 - 0.19)** | **2.40e-03** |
| Nucleus Accumbens | L | 1.62 | 0.20 | 0.34 | -0.20 | (-0.41 - 0.02) | 0.07 | -0.18 | (-0.39 - 0.04) | 0.10 | -0.02 | (-0.09 - 0.05) | 0.63 |
| | R | 0.66 | 0.52 | 0.65 | -0.12 | (-0.33 - 0.1) | 0.29 | -0.12 | (-0.34 - 0.09) | 0.25 | 0.01 | (-0.06 - 0.08) | 0.81 |
| Pallidum | L | 0.27 | 0.76 | 0.82 | -0.07 | (-0.28 - 0.14) | 0.53 | -0.05 | (-0.26 - 0.16) | 0.64 | -0.02 | (-0.09 - 0.06) | 0.63 |
| | R | 0.44 | 0.64 | 0.74 | -0.10 | (-0.31 - 0.11) | 0.36 | -0.09 | (-0.3 - 0.13) | 0.43 | -0.01 | (-0.09 - 0.06) | 0.71 |
| Putamen | L | 0.33 | 0.72 | 0.80 | -0.09 | (-0.3 - 0.13) | 0.43 | -0.09 | (-0.3 - 0.13) | 0.42 | 0.00 | (-0.07 - 0.07) | 0.99 |
| | R | 0.15 | 0.86 | 0.89 | -0.06 | (-0.28 - 0.16) | 0.59 | -0.06 | (-0.27 - 0.16) | 0.60 | 0.00 | (-0.07 - 0.07) | 0.96 |
| Thalamus | L | 1.55 | 0.21 | 0.35 | -0.17 | (-0.39 - 0.04) | 0.11 | -0.13 | (-0.35 - 0.08) | 0.21 | -0.04 | (-0.11 - 0.03) | 0.30 |
| | R | 4.20 | 0.02 | 0.07 | **-0.26** | **(-0.47 - -0.05)** | **0.02** | -0.19 | (-0.4 - 0.03) | 0.09 | **-0.08** | **(-0.15 - 0)** | **0.04** |
| **Global** | | | | | | | | | | | | | |
| Estimated Intracranial Volume | | **7.75** | **4.40e-04** | **7.57e-03** | -0.17 | (-0.38 - 0.05) | 0.13 | -0.02 | (-0.23 - 0.19) | 0.85 | **-0.14** | **(-0.22 - -0.07)** | **1.21e-04** |
| Mean Cortical Thickness | N/A | **9.27** | **9.67e-05** | **3.03e-03** | -0.12 | (-0.33 - 0.1) | 0.29 | 0.05 | (-0.17 - 0.26) | 0.67 | **-0.16** | **(-0.23 - -0.09)** | **1.79e-05** |
| Total Surface Area | | **10.09** | **4.31e-05** | **3.03e-03** | -0.07 | (-0.29 - 0.14) | 0.51 | 0.10 | (-0.12 - 0.31) | 0.37 | **-0.17** | **(-0.24 - -0.09)** | **7.43e-06** |

*Results marked in bold are significant at q<0.05 (the overall effect) or at p<0.05 (pairwise comparisons).*

**eTable 15.** GRD subgroup (GRD vs. no GRD assignment vs. HC) effect size overview

| | | Overall Test Statistics | | | GRD < HC | | | GRD < no GRD | | | no GRD < HC | | |
|---|---|---|---|---|---|---|---|---|---|---|---|---|---|
| Region of Interest | Hemisphere | F | p | q | Cohen's d | 95% Confidence Interval | p | Cohen's d | 95% Confidence Interval | p | Cohen's d | 95% Confidence Interval | p |
| **Cortical Thickness** | | | | | | | | | | | | | |
| Banks of the Superior Temporal Sulcus | L | 2.57 | 0.08 | 0.18 | -0.05 | (-0.19 - 0.08) | 0.44 | 0.03 | (-0.1 - 0.17) | 0.62 | **-0.09** | **(-0.16 - -0.01)** | **0.02** |
| | R | **6.49** | **1.55e-03** | **0.01** | **-0.16** | **(-0.3 - -0.03)** | **0.02** | -0.04 | (-0.17 - 0.1) | 0.61 | **-0.13** | **(-0.2 - -0.05)** | **9.98e-04** |
| Caudal Anterior Cingulate | L | 1.91 | 0.15 | 0.31 | -0.03 | (-0.16 - 0.11) | 0.70 | 0.05 | (-0.09 - 0.19) | 0.49 | -0.07 | (-0.15 - 0) | 0.05 |
| | R | 3.38 | 0.03 | 0.12 | 0.05 | (-0.09 - 0.19) | 0.47 | 0.14 | (0 - 0.27) | 0.05 | **-0.08** | **(-0.16 - -0.01)** | **0.03** |
| Caudal Middle Frontal | L | 3.66 | 0.03 | 0.10 | -0.12 | (-0.25 - 0.02) | 0.10 | -0.02 | (-0.15 - 0.12) | 0.79 | **-0.10** | **(-0.17 - -0.02)** | **0.01** |
| | R | 4.43 | 0.01 | 0.07 | -0.10 | (-0.24 - 0.03) | 0.14 | 0.01 | (-0.12 - 0.15) | 0.87 | **-0.11** | **(-0.19 - -0.04)** | **3.70e-03** |
| Cuneus | L | 0.50 | 0.61 | 0.72 | 0.07 | (-0.07 - 0.2) | 0.33 | 0.05 | (-0.09 - 0.19) | 0.47 | 0.02 | (-0.06 - 0.09) | 0.64 |
| | R | 0.37 | 0.69 | 0.80 | -0.03 | (-0.17 - 0.1) | 0.63 | 0.00 | (-0.14 - 0.13) | 0.98 | -0.03 | (-0.11 - 0.04) | 0.41 |
| Entorhinal | L | 0.56 | 0.57 | 0.68 | -0.07 | (-0.21 - 0.06) | 0.29 | -0.06 | (-0.2 - 0.07) | 0.36 | -0.01 | (-0.08 - 0.06) | 0.79 |
| | R | 1.61 | 0.20 | 0.37 | -0.12 | (-0.25 - 0.02) | 0.09 | -0.08 | (-0.21 - 0.06) | 0.27 | -0.04 | (-0.12 - 0.03) | 0.29 |
| Frontal Pole | L | 0.77 | 0.46 | 0.62 | 0.01 | (-0.12 - 0.15) | 0.84 | 0.06 | (-0.08 - 0.19) | 0.41 | -0.04 | (-0.12 - 0.03) | 0.26 |
| | R | 1.84 | 0.16 | 0.32 | -0.06 | (-0.2 - 0.07) | 0.36 | 0.01 | (-0.13 - 0.15) | 0.90 | -0.07 | (-0.15 - 0) | 0.06 |
| Fusiform | L | **8.80** | **1.55e-04** | **4.01e-03** | -0.12 | (-0.25 - 0.02) | 0.09 | 0.05 | (-0.09 - 0.18) | 0.52 | **-0.16** | **(-0.23 - -0.08)** | **3.23e-05** |
| | R | 3.19 | 0.04 | 0.13 | -0.12 | (-0.26 - 0.01) | 0.08 | -0.04 | (-0.17 - 0.1) | 0.60 | **-0.09** | **(-0.16 - -0.01)** | **0.03** |
| Inferior Parietal | L | **8.95** | **1.33e-04** | **4.01e-03** | -0.13 | (-0.27 - 0) | 0.06 | 0.03 | (-0.11 - 0.16) | 0.68 | **-0.16** | **(-0.23 - -0.08)** | **3.22e-05** |
| | R | 4.21 | 0.01 | 0.07 | -0.07 | (-0.21 - 0.07) | 0.31 | 0.04 | (-0.09 - 0.18) | 0.54 | **-0.11** | **(-0.19 - -0.04)** | **3.80e-03** |
| Inferior Temporal | L | 4.13 | 0.02 | 0.07 | -0.10 | (-0.24 - 0.03) | 0.14 | 0.00 | (-0.13 - 0.14) | 0.95 | **-0.11** | **(-0.18 - -0.03)** | **5.44e-03** |
| | R | **6.94** | **9.86e-04** | **0.01** | **-0.19** | **(-0.33 - -0.06)** | **5.94e-03** | -0.07 | (-0.2 - 0.07) | 0.33 | **-0.12** | **(-0.2 - -0.05)** | **1.43e-03** |
| Insula | L | **9.69** | **6.40e-05** | **3.31e-03** | **-0.22** | **(-0.36 - -0.08)** | **1.54e-03** | -0.07 | (-0.21 - 0.06) | 0.31 | **-0.15** | **(-0.22 - -0.07)** | **1.27e-04** |
| | R | **7.29** | **6.95e-04** | **9.79e-03** | -0.11 | (-0.25 - 0.02) | 0.10 | 0.03 | (-0.1 - 0.17) | 0.64 | **-0.14** | **(-0.22 - -0.07)** | **1.65e-04** |
| Isthmus Cingulate | L | 0.11 | 0.90 | 0.92 | -0.02 | (-0.15 - 0.12) | 0.80 | 0.00 | (-0.14 - 0.13) | 0.99 | -0.02 | (-0.09 - 0.06) | 0.66 |
| | R | 1.21 | 0.30 | 0.47 | -0.10 | (-0.23 - 0.04) | 0.17 | -0.05 | (-0.19 - 0.08) | 0.45 | -0.04 | (-0.12 - 0.03) | 0.27 |
| Lateral Occipital | L | 4.40 | 0.01 | 0.07 | -0.04 | (-0.17 - 0.1) | 0.59 | 0.08 | (-0.06 - 0.21) | 0.27 | **-0.11** | **(-0.19 - -0.04)** | **3.22e-03** |
| | R | **8.04** | **3.30e-04** | **5.84e-03** | **-0.19** | **(-0.32 - -0.05)** | **7.38e-03** | -0.05 | (-0.18 - 0.09) | 0.51 | **-0.14** | **(-0.21 - -0.06)** | **2.96e-04** |
| | | | | | | | | | (-0.02 - | | | | |

| | | | | | | | | | | | | | |
|---|---|---|---|---|---|---|---|---|---|---|---|---|---|
| Lateral Orbitofrontal | L | 2.86 | 0.06 | 0.15 | 0.04 | (-0.1 - 0.17) | 0.60 | 0.12 | 0.25) | 0.09 | **-0.08** | **(-0.15 - 0)** | **0.04** |
| | R | 1.79 | 0.17 | 0.33 | -0.07 | (-0.21 - 0.06) | 0.30 | 0.00 | (-0.14 - 0.13) | 0.98 | -0.07 | (-0.14 - 0.01) | 0.07 |
| Lingual | L | 0.42 | 0.66 | 0.77 | 0.03 | (-0.11 - 0.16) | 0.69 | 0.05 | (-0.08 - 0.19) | 0.44 | -0.03 | (-0.1 - 0.05) | 0.50 |
| | R | 0.04 | 0.96 | 0.96 | 0.02 | (-0.12 - 0.15) | 0.80 | 0.01 | (-0.13 - 0.15) | 0.90 | 0.01 | (-0.07 - 0.08) | 0.83 |
| Medial Orbitofrontal | L | **5.72** | **3.32e-03** | **0.03** | -0.04 | (-0.17 - 0.1) | 0.60 | 0.09 | (-0.04 - 0.23) | 0.18 | **-0.13** | **(-0.2 - -0.05)** | **8.12e-04** |
| | R | 3.54 | 0.03 | 0.11 | -0.02 | (-0.15 - 0.12) | 0.81 | 0.09 | (-0.05 - 0.22) | 0.22 | **-0.10** | **(-0.17 - -0.02)** | **9.23e-03** |
| Middle Temporal | L | 3.18 | 0.04 | 0.13 | -0.03 | (-0.17 - 0.1) | 0.63 | 0.06 | (-0.07 - 0.2) | 0.35 | **-0.10** | **(-0.17 - -0.02)** | **0.01** |
| | R | **11.41** | **1.16e-05** | **1.79e-03** | **-0.15** | **(-0.29 - -0.01)** | **0.03** | 0.03 | (-0.1 - 0.17) | 0.63 | **-0.18** | **(-0.26 - -0.11)** | **2.68e-06** |
| Paracentral | L | **6.68** | **1.27e-03** | **0.01** | -0.13 | (-0.26 - 0.01) | 0.07 | 0.01 | (-0.13 - 0.15) | 0.88 | **-0.14** | **(-0.21 - -0.06)** | **3.85e-04** |
| | R | 2.90 | 0.06 | 0.15 | -0.10 | (-0.24 - 0.04) | 0.16 | -0.01 | (-0.15 - 0.13) | 0.88 | **-0.09** | **(-0.16 - -0.01)** | **0.02** |
| Parahippocampal | L | **5.02** | **6.65e-03** | **0.05** | -0.13 | (-0.26 - 0.01) | 0.06 | -0.01 | (-0.15 - 0.12) | 0.86 | **-0.11** | **(-0.19 - -0.04)** | **2.77e-03** |
| | R | 1.25 | 0.29 | 0.46 | -0.06 | (-0.19 - 0.08) | 0.41 | 0.00 | (-0.13 - 0.14) | 0.97 | -0.06 | (-0.13 - 0.02) | 0.13 |
| Pars Opercularis | L | 0.71 | 0.49 | 0.63 | -0.08 | (-0.21 - 0.06) | 0.26 | -0.05 | (-0.19 - 0.08) | 0.46 | -0.03 | (-0.1 - 0.05) | 0.49 |
| | R | 2.14 | 0.12 | 0.26 | **-0.14** | **(-0.27 - 0)** | **0.05** | -0.09 | (-0.23 - 0.04) | 0.18 | -0.04 | (-0.12 - 0.03) | 0.24 |
| Pars Orbitalis | L | 0.66 | 0.52 | 0.64 | -0.05 | (-0.19 - 0.08) | 0.44 | -0.01 | (-0.15 - 0.12) | 0.85 | -0.04 | (-0.11 - 0.04) | 0.30 |
| | R | 0.73 | 0.48 | 0.63 | -0.06 | (-0.2 - 0.08) | 0.39 | -0.08 | (-0.22 - 0.05) | 0.24 | 0.02 | (-0.05 - 0.1) | 0.56 |
| Pars Triangularis | L | 1.06 | 0.35 | 0.51 | -0.04 | (-0.18 - 0.09) | 0.55 | -0.08 | (-0.22 - 0.05) | 0.22 | 0.04 | (-0.03 - 0.12) | 0.27 |
| | R | 0.86 | 0.43 | 0.59 | -0.08 | (-0.22 - 0.05) | 0.24 | -0.05 | (-0.18 - 0.09) | 0.50 | -0.03 | (-0.11 - 0.04) | 0.37 |
| Pericalcarine | L | 1.55 | 0.21 | 0.38 | 0.12 | (-0.01 - 0.26) | 0.08 | 0.11 | (-0.03 - 0.24) | 0.13 | 0.02 | (-0.06 - 0.09) | 0.67 |
| | R | 1.71 | 0.18 | 0.35 | 0.10 | (-0.04 - 0.23) | 0.17 | 0.04 | (-0.1 - 0.17) | 0.61 | 0.06 | (-0.01 - 0.14) | 0.12 |
| Postcentral | L | 3.58 | 0.03 | 0.11 | **-0.17** | **(-0.31 - -0.03)** | **0.02** | -0.10 | (-0.24 - 0.04) | 0.15 | -0.07 | (-0.14 - 0.01) | 0.08 |
| | R | 4.52 | 0.01 | 0.07 | -0.10 | (-0.24 - 0.03) | 0.14 | 0.01 | (-0.12 - 0.15) | 0.87 | **-0.11** | **(-0.19 - -0.04)** | **3.43e-03** |
| Posterior Cingulate | L | 3.07 | 0.05 | 0.14 | -0.07 | (-0.21 - 0.06) | 0.28 | 0.02 | (-0.12 - 0.16) | 0.77 | **-0.09** | **(-0.17 - -0.02)** | **0.01** |
| | R | **5.20** | **5.59e-03** | **0.04** | -0.13 | (-0.27 - 0.01) | 0.06 | -0.01 | (-0.15 - 0.12) | 0.88 | **-0.12** | **(-0.19 - -0.04)** | **2.28e-03** |
| Precentral | L | 4.56 | 0.01 | 0.07 | **-0.16** | **(-0.29 - -0.02)** | **0.02** | -0.06 | (-0.19 - 0.08) | 0.41 | **-0.10** | **(-0.17 - -0.02)** | **0.01** |
| | R | 4.53 | 0.01 | 0.07 | -0.09 | (-0.22 - 0.05) | 0.22 | 0.03 | (-0.11 - 0.17) | 0.66 | **-0.11** | **(-0.19 - -0.04)** | **2.86e-03** |
| Precuneus | L | 3.26 | 0.04 | 0.13 | -0.09 | (-0.22 - 0.05) | 0.22 | 0.01 | (-0.12 - 0.15) | 0.86 | **-0.10** | **(-0.17 - -0.02)** | **0.01** |
| | R | **7.90** | **3.77e-04** | **5.84e-03** | **-0.24** | **(-0.38 - -0.11)** | **4.55e-04** | -0.14 | (-0.27 - 0) | 0.05 | **-0.11** | **(-0.18 - -0.03)** | **5.23e-03** |
| Rostral Anterior | L | 3.38 | 0.03 | 0.12 | -0.13 | (-0.26 - 0.01) | 0.07 | -0.04 | (-0.17 - 0.1) | 0.59 | **-0.09** | **(-0.16 - -0.01)** | **0.02** |

| | | | | | | | | | | | | | |
|---|---|---|---|---|---|---|---|---|---|---|---|---|---|
| Cingulate | R | 1.81 | 0.16 | 0.33 | -0.09 | (-0.23 - 0.04) | 0.18 | -0.03 | (-0.16 - 0.11) | 0.68 | -0.06 | (-0.14 - 0.01) | 0.10 |
| Rostral Middle Frontal | L | **5.18** | **5.68e-03** | **0.04** | -0.08 | (-0.21 - 0.06) | 0.28 | 0.05 | (-0.09 - 0.19) | 0.47 | **-0.12** | **(-0.2 - -0.05)** | **1.32e-03** |
| | R | 4.09 | 0.02 | 0.07 | -0.11 | (-0.25 - 0.03) | 0.12 | 0.00 | (-0.14 - 0.13) | 0.96 | **-0.10** | **(-0.18 - -0.03)** | **6.22e-03** |
| Superior Frontal | L | 3.55 | 0.03 | 0.11 | **-0.15** | **(-0.28 - -0.01)** | **0.03** | -0.06 | (-0.2 - 0.07) | 0.35 | **-0.08** | **(-0.16 - -0.01)** | **0.03** |
| | R | 2.03 | 0.13 | 0.29 | -0.07 | (-0.2 - 0.07) | 0.34 | 0.01 | (-0.13 - 0.15) | 0.88 | **-0.08** | **(-0.15 - 0)** | **0.05** |
| Superior Parietal | L | 2.98 | 0.05 | 0.14 | -0.11 | (-0.25 - 0.02) | 0.10 | -0.03 | (-0.16 - 0.11) | 0.69 | **-0.08** | **(-0.16 - -0.01)** | **0.03** |
| | R | 4.32 | 0.01 | 0.07 | -0.07 | (-0.21 - 0.07) | 0.31 | 0.04 | (-0.09 - 0.18) | 0.54 | **-0.11** | **(-0.19 - -0.04)** | **3.37e-03** |
| Superior Temporal | L | 4.16 | 0.02 | 0.07 | -0.09 | (-0.23 - 0.05) | 0.19 | 0.02 | (-0.12 - 0.16) | 0.79 | **-0.11** | **(-0.18 - -0.03)** | **4.60e-03** |
| | R | **6.71** | **1.24e-03** | **0.01** | **-0.16** | **(-0.3 - -0.03)** | **0.02** | -0.03 | (-0.17 - 0.1) | 0.63 | **-0.13** | **(-0.2 - -0.05)** | **7.87e-04** |
| Supramarginal | L | 3.82 | 0.02 | 0.09 | **-0.17** | **(-0.3 - -0.03)** | **0.02** | -0.09 | (-0.23 - 0.04) | 0.19 | **-0.08** | **(-0.15 - 0)** | **0.05** |
| | R | 2.68 | 0.07 | 0.17 | -0.05 | (-0.18 - 0.09) | 0.50 | 0.04 | (-0.09 - 0.18) | 0.53 | **-0.09** | **(-0.16 - -0.01)** | **0.02** |
| Temporal Pole | L | 3.21 | 0.04 | 0.13 | 0.07 | (-0.07 - 0.21) | 0.31 | **0.15** | **(0.01 - 0.28)** | **0.04** | -0.07 | (-0.15 - 0) | 0.05 |
| | R | 2.79 | 0.06 | 0.16 | -0.01 | (-0.15 - 0.13) | 0.89 | 0.08 | (-0.06 - 0.22) | 0.25 | **-0.09** | **(-0.16 - -0.01)** | **0.02** |
| Transverse Temporal | L | 1.77 | 0.17 | 0.34 | -0.12 | (-0.26 - 0.02) | 0.09 | -0.07 | (-0.21 - 0.06) | 0.31 | -0.05 | (-0.12 - 0.03) | 0.22 |
| | R | 3.15 | 0.04 | 0.13 | -0.11 | (-0.24 - 0.03) | 0.13 | -0.02 | (-0.15 - 0.12) | 0.82 | **-0.09** | **(-0.16 - -0.01)** | **0.02** |
| **Surface Area** | | | | | | | | | | | | | |
| Banks of the Superior Temporal Sulcus | L | 1.94 | 0.14 | 0.31 | -0.10 | (-0.24 - 0.03) | 0.14 | -0.04 | (-0.17 - 0.1) | 0.59 | -0.06 | (-0.14 - 0.01) | 0.09 |
| | R | 4.25 | 0.01 | 0.07 | **-0.20** | **(-0.33 - -0.06)** | **4.49e-03** | **-0.14** | **(-0.28 - -0.01)** | **0.04** | -0.05 | (-0.13 - 0.02) | 0.15 |
| Caudal Anterior Cingulate | L | 0.16 | 0.85 | 0.91 | 0.01 | (-0.12 - 0.15) | 0.84 | 0.03 | (-0.1 - 0.17) | 0.64 | -0.02 | (-0.09 - 0.06) | 0.65 |
| | R | 2.45 | 0.09 | 0.20 | -0.07 | (-0.2 - 0.07) | 0.35 | 0.02 | (-0.12 - 0.16) | 0.77 | **-0.08** | **(-0.16 - -0.01)** | **0.03** |
| Caudal Middle Frontal | L | 1.04 | 0.35 | 0.51 | -0.04 | (-0.18 - 0.09) | 0.55 | 0.01 | (-0.12 - 0.15) | 0.84 | -0.05 | (-0.13 - 0.02) | 0.15 |
| | R | 0.96 | 0.38 | 0.55 | 0.05 | (-0.09 - 0.19) | 0.48 | 0.09 | (-0.05 - 0.22) | 0.22 | -0.04 | (-0.11 - 0.04) | 0.35 |
| Cuneus | L | 1.53 | 0.22 | 0.38 | -0.01 | (-0.15 - 0.13) | 0.90 | 0.06 | (-0.08 - 0.19) | 0.40 | -0.07 | (-0.14 - 0.01) | 0.09 |
| | R | 2.67 | 0.07 | 0.17 | -0.10 | (-0.24 - 0.03) | 0.14 | -0.02 | (-0.16 - 0.12) | 0.78 | **-0.08** | **(-0.16 - -0.01)** | **0.03** |
| Entorhinal | L | 3.68 | 0.03 | 0.10 | -0.05 | (-0.19 - 0.09) | 0.48 | 0.06 | (-0.08 - 0.19) | 0.42 | **-0.10** | **(-0.18 - -0.03)** | **6.71e-03** |
| | R | 1.38 | 0.25 | 0.42 | 0.07 | (-0.07 - 0.2) | 0.35 | 0.11 | (-0.03 - 0.24) | 0.13 | -0.04 | (-0.12 - 0.03) | 0.29 |
| Frontal Pole | L | 0.46 | 0.63 | 0.74 | 0.02 | (-0.12 - 0.15) | 0.80 | -0.02 | (-0.16 - 0.12) | 0.77 | 0.04 | (-0.04 - 0.11) | 0.34 |
| | R | 0.70 | 0.50 | 0.63 | 0.04 | (-0.1 - 0.18) | 0.56 | 0.00 | (-0.14 - 0.13) | 0.96 | 0.04 | (-0.03 - 0.12) | 0.25 |
| Fusiform | L | 1.61 | 0.20 | 0.37 | -0.12 | (-0.26 - 0.01) | 0.08 | -0.09 | (-0.22 - 0.05) | 0.22 | -0.04 | (-0.11 - 0.04) | 0.36 |

| | | | | | | | | | | | | |
|---|---|---|---|---|---|---|---|---|---|---|---|---|
| | R | 1.94 | 0.14 | 0.31 | -0.09 | (-0.23 - 0.05) | 0.19 | -0.02 | (-0.16 - 0.12) | 0.77 | -0.07 | (-0.14 - 0.01) | 0.07 |
| Inferior Parietal | L | 1.26 | 0.29 | 0.46 | -0.08 | (-0.22 - 0.06) | 0.25 | -0.03 | (-0.16 - 0.11) | 0.71 | -0.05 | (-0.13 - 0.02) | 0.17 |
| | R | **7.12** | **8.23e-04** | **0.01** | **-0.25** | **(-0.39 - -0.12)** | **2.43e-04** | **-0.18** | **(-0.32 - -0.05)** | **8.64e-03** | -0.07 | (-0.15 - 0) | 0.06 |
| Inferior Temporal | L | 0.52 | 0.59 | 0.71 | -0.06 | (-0.2 - 0.07) | 0.35 | -0.04 | (-0.17 - 0.1) | 0.57 | -0.03 | (-0.1 - 0.05) | 0.51 |
| | R | 1.15 | 0.32 | 0.49 | -0.10 | (-0.24 - 0.03) | 0.13 | -0.10 | (-0.23 - 0.04) | 0.17 | -0.01 | (-0.08 - 0.07) | 0.82 |
| Insula | L | 1.57 | 0.21 | 0.38 | -0.04 | (-0.17 - 0.1) | 0.57 | 0.03 | (-0.11 - 0.17) | 0.67 | -0.07 | (-0.14 - 0.01) | 0.08 |
| | R | 0.76 | 0.47 | 0.62 | -0.06 | (-0.19 - 0.08) | 0.42 | -0.01 | (-0.15 - 0.12) | 0.86 | -0.04 | (-0.12 - 0.03) | 0.26 |
| Isthmus Cingulate | L | 0.28 | 0.75 | 0.85 | 0.03 | (-0.1 - 0.17) | 0.62 | 0.05 | (-0.09 - 0.19) | 0.47 | -0.02 | (-0.09 - 0.06) | 0.69 |
| | R | 2.92 | 0.05 | 0.15 | 0.11 | (-0.02 - 0.25) | 0.10 | 0.03 | (-0.11 - 0.16) | 0.68 | **0.08** | **(0.01 - 0.16)** | **0.03** |
| Lateral Occipital | L | 1.53 | 0.22 | 0.38 | -0.11 | (-0.25 - 0.03) | 0.12 | -0.12 | (-0.26 - 0.02) | 0.08 | 0.01 | (-0.06 - 0.09) | 0.76 |
| | R | 0.12 | 0.88 | 0.92 | 0.00 | (-0.14 - 0.14) | 0.99 | 0.02 | (-0.12 - 0.16) | 0.78 | -0.02 | (-0.09 - 0.06) | 0.64 |
| Lateral Orbitofrontal | L | 0.23 | 0.79 | 0.87 | -0.01 | (-0.14 - 0.13) | 0.91 | 0.02 | (-0.12 - 0.15) | 0.79 | -0.03 | (-0.1 - 0.05) | 0.50 |
| | R | 0.77 | 0.46 | 0.62 | 0.03 | (-0.11 - 0.16) | 0.70 | 0.07 | (-0.07 - 0.2) | 0.34 | -0.04 | (-0.11 - 0.04) | 0.31 |
| Lingual | L | 1.41 | 0.24 | 0.41 | -0.06 | (-0.2 - 0.07) | 0.37 | 0.00 | (-0.14 - 0.14) | 0.99 | -0.06 | (-0.14 - 0.01) | 0.11 |
| | R | 0.29 | 0.75 | 0.85 | -0.05 | (-0.19 - 0.08) | 0.46 | -0.04 | (-0.17 - 0.1) | 0.60 | -0.01 | (-0.09 - 0.06) | 0.70 |
| Medial Orbitofrontal | L | 0.24 | 0.79 | 0.87 | -0.04 | (-0.18 - 0.09) | 0.52 | -0.03 | (-0.16 - 0.11) | 0.70 | -0.02 | (-0.09 - 0.06) | 0.65 |
| | R | 3.33 | 0.04 | 0.12 | **-0.14** | **(-0.28 - -0.01)** | **0.04** | -0.06 | (-0.2 - 0.07) | 0.37 | **-0.08** | **(-0.15 - 0)** | **0.04** |
| Middle Temporal | L | 0.13 | 0.88 | 0.92 | -0.01 | (-0.15 - 0.12) | 0.87 | -0.03 | (-0.16 - 0.11) | 0.69 | 0.02 | (-0.06 - 0.09) | 0.69 |
| | R | 2.56 | 0.08 | 0.18 | **-0.14** | **(-0.28 - 0)** | **0.04** | -0.08 | (-0.22 - 0.05) | 0.25 | -0.06 | (-0.13 - 0.02) | 0.12 |
| Paracentral | L | 0.23 | 0.79 | 0.87 | 0.03 | (-0.11 - 0.16) | 0.69 | 0.04 | (-0.09 - 0.18) | 0.53 | -0.02 | (-0.09 - 0.06) | 0.67 |
| | R | 1.07 | 0.34 | 0.51 | 0.10 | (-0.04 - 0.23) | 0.17 | 0.10 | (-0.04 - 0.24) | 0.15 | 0.00 | (-0.08 - 0.07) | 0.92 |
| Parahippocampal | L | 1.20 | 0.30 | 0.47 | -0.09 | (-0.23 - 0.04) | 0.18 | -0.05 | (-0.19 - 0.08) | 0.46 | -0.04 | (-0.12 - 0.03) | 0.27 |
| | R | 1.32 | 0.27 | 0.44 | 0.04 | (-0.09 - 0.18) | 0.55 | -0.02 | (-0.16 - 0.11) | 0.75 | 0.06 | (-0.01 - 0.14) | 0.11 |
| Pars Opercularis | L | 2.88 | 0.06 | 0.15 | -0.02 | (-0.15 - 0.12) | 0.81 | 0.07 | (-0.06 - 0.21) | 0.28 | **-0.09** | **(-0.17 - -0.02)** | **0.02** |
| | R | 1.11 | 0.33 | 0.50 | 0.08 | (-0.05 - 0.22) | 0.24 | 0.10 | (-0.03 - 0.24) | 0.14 | -0.02 | (-0.1 - 0.05) | 0.59 |
| Pars Orbitalis | L | 0.22 | 0.81 | 0.87 | 0.04 | (-0.09 - 0.18) | 0.54 | 0.03 | (-0.11 - 0.16) | 0.69 | 0.02 | (-0.06 - 0.09) | 0.69 |
| | R | 0.29 | 0.75 | 0.85 | 0.02 | (-0.11 - 0.16) | 0.75 | -0.01 | (-0.14 - 0.13) | 0.92 | 0.03 | (-0.05 - 0.1) | 0.45 |
| Pars Triangularis | L | 3.04 | 0.05 | 0.14 | 0.07 | (-0.07 - 0.2) | 0.33 | **0.14** | **(0.01 - 0.28)** | **0.04** | -0.07 | (-0.15 - 0) | 0.06 |
| | R | 0.11 | 0.90 | 0.92 | 0.03 | (-0.11 - 0.16) | 0.69 | 0.01 | (-0.12 - 0.15) | 0.84 | 0.01 | (-0.06 - 0.09) | 0.73 |
| | L | 1.15 | 0.32 | 0.49 | -0.04 | (-0.18 - 0.1) | 0.57 | 0.02 | (-0.12 - | 0.78 | -0.06 | (-0.13 - | 0.13 |

| | | | | | | | | | | | | | |
|---|---|---|---|---|---|---|---|---|---|---|---|---|---|
| Pericalcarine | | | | | | | | | 0.16) | | | 0.02) | |
| | R | 3.94 | 0.02 | 0.08 | -0.09 | (-0.23 - 0.05) | 0.20 | 0.02 | (-0.12 - 0.15) | 0.80 | **-0.11** | **(-0.18 - -0.03)** | **5.86e-03** |
| Postcentral | L | 0.85 | 0.43 | 0.59 | -0.08 | (-0.21 - 0.06) | 0.27 | -0.09 | (-0.23 - 0.05) | 0.19 | 0.01 | (-0.06 - 0.09) | 0.73 |
| | R | 0.73 | 0.48 | 0.63 | -0.02 | (-0.15 - 0.12) | 0.79 | 0.03 | (-0.11 - 0.16) | 0.68 | -0.05 | (-0.12 - 0.03) | 0.23 |
| Posterior Cingulate | L | 0.70 | 0.50 | 0.63 | -0.04 | (-0.17 - 0.1) | 0.59 | 0.01 | (-0.13 - 0.14) | 0.91 | -0.04 | (-0.12 - 0.03) | 0.24 |
| | R | 2.00 | 0.14 | 0.30 | -0.10 | (-0.24 - 0.03) | 0.14 | -0.04 | (-0.17 - 0.1) | 0.60 | -0.07 | (-0.14 - 0.01) | 0.09 |
| Precentral | L | 0.90 | 0.41 | 0.57 | 0.03 | (-0.11 - 0.17) | 0.66 | -0.02 | (-0.16 - 0.11) | 0.76 | 0.05 | (-0.02 - 0.13) | 0.18 |
| | R | 0.07 | 0.93 | 0.93 | 0.00 | (-0.14 - 0.14) | 1.00 | -0.01 | (-0.15 - 0.12) | 0.83 | 0.01 | (-0.06 - 0.09) | 0.71 |
| Precuneus | L | 0.62 | 0.54 | 0.66 | -0.07 | (-0.2 - 0.07) | 0.34 | -0.04 | (-0.17 - 0.1) | 0.61 | -0.03 | (-0.11 - 0.04) | 0.42 |
| | R | 0.59 | 0.55 | 0.67 | -0.05 | (-0.18 - 0.09) | 0.52 | -0.01 | (-0.14 - 0.13) | 0.94 | -0.04 | (-0.11 - 0.04) | 0.31 |
| Rostral Anterior Cingulate | L | 4.46 | 0.01 | 0.07 | -0.09 | (-0.22 - 0.05) | 0.20 | 0.03 | (-0.11 - 0.16) | 0.71 | **-0.11** | **(-0.19 - -0.04)** | **3.19e-03** |
| | R | 2.21 | 0.11 | 0.25 | -0.02 | (-0.16 - 0.11) | 0.74 | 0.06 | (-0.08 - 0.19) | 0.40 | **-0.08** | **(-0.15 - 0)** | **0.04** |
| Rostral Middle Frontal | L | 0.15 | 0.86 | 0.92 | -0.04 | (-0.17 - 0.1) | 0.61 | -0.02 | (-0.16 - 0.11) | 0.74 | -0.01 | (-0.09 - 0.06) | 0.74 |
| | R | 1.72 | 0.18 | 0.35 | -0.13 | (-0.26 - 0.01) | 0.07 | -0.12 | (-0.25 - 0.02) | 0.09 | -0.01 | (-0.08 - 0.07) | 0.83 |
| Superior Frontal | L | 1.57 | 0.21 | 0.38 | 0.07 | (-0.07 - 0.2) | 0.35 | 0.11 | (-0.03 - 0.25) | 0.11 | -0.05 | (-0.12 - 0.03) | 0.24 |
| | R | 4.48 | 0.01 | 0.07 | -0.03 | (-0.16 - 0.11) | 0.69 | 0.09 | (-0.05 - 0.22) | 0.21 | **-0.11** | **(-0.19 - -0.04)** | **3.13e-03** |
| Superior Parietal | L | 0.11 | 0.90 | 0.92 | 0.03 | (-0.1 - 0.17) | 0.65 | 0.02 | (-0.11 - 0.16) | 0.73 | 0.01 | (-0.07 - 0.08) | 0.84 |
| | R | 1.49 | 0.23 | 0.39 | -0.08 | (-0.22 - 0.05) | 0.23 | -0.12 | (-0.25 - 0.02) | 0.09 | 0.03 | (-0.04 - 0.11) | 0.41 |
| Superior Temporal | L | 0.66 | 0.51 | 0.64 | -0.07 | (-0.21 - 0.07) | 0.31 | -0.04 | (-0.18 - 0.1) | 0.57 | -0.03 | (-0.11 - 0.04) | 0.42 |
| | R | 1.27 | 0.28 | 0.46 | -0.07 | (-0.21 - 0.06) | 0.28 | -0.02 | (-0.15 - 0.12) | 0.79 | -0.06 | (-0.13 - 0.02) | 0.15 |
| Supramarginal | L | **5.56** | **3.88e-03** | **0.03** | **-0.18** | **(-0.32 - -0.05)** | **8.78e-03** | -0.08 | (-0.21 - 0.06) | 0.28 | **-0.11** | **(-0.18 - -0.03)** | **6.39e-03** |
| | R | 1.41 | 0.25 | 0.41 | -0.06 | (-0.2 - 0.07) | 0.38 | 0.00 | (-0.13 - 0.14) | 0.97 | -0.06 | (-0.14 - 0.01) | 0.10 |
| Temporal Pole | L | 2.46 | 0.09 | 0.20 | **-0.15** | **(-0.29 - -0.02)** | **0.03** | **-0.14** | **(-0.28 - -0.01)** | **0.04** | -0.01 | (-0.09 - 0.06) | 0.78 |
| | R | 1.40 | 0.25 | 0.41 | 0.11 | (-0.02 - 0.25) | 0.11 | 0.08 | (-0.06 - 0.21) | 0.27 | 0.03 | (-0.04 - 0.11) | 0.37 |
| Transverse Temporal | L | 1.05 | 0.35 | 0.51 | -0.05 | (-0.19 - 0.08) | 0.44 | 0.00 | (-0.14 - 0.14) | 0.99 | -0.05 | (-0.13 - 0.02) | 0.16 |
| | R | 0.09 | 0.91 | 0.92 | -0.02 | (-0.16 - 0.12) | 0.77 | -0.01 | (-0.14 - 0.13) | 0.94 | -0.01 | (-0.09 - 0.06) | 0.70 |
| **Subcortical Volume** | | | | | | | | | | | | | |
| Amygdala | L | 0.72 | 0.49 | 0.63 | 0.00 | (-0.13 - 0.14) | 0.98 | 0.05 | (-0.09 - 0.18) | 0.51 | -0.04 | (-0.12 - 0.03) | 0.25 |
| | R | 2.67 | 0.07 | 0.17 | -0.08 | (-0.22 - 0.05) | 0.22 | 0.00 | (-0.13 - 0.14) | 0.98 | **-0.09** | **(-0.16 - -0.01)** | **0.03** |

| | | | | | | | | | | | | | |
|---|---|---|---|---|---|---|---|---|---|---|---|---|---|
| Caudate | L | 0.14 | 0.87 | 0.92 | 0.03 | (-0.1 - 0.17) | 0.65 | 0.02 | (-0.12 - 0.15) | 0.80 | 0.01 | (-0.06 - 0.09) | 0.70 |
| | R | 0.59 | 0.55 | 0.67 | 0.05 | (-0.09 - 0.18) | 0.51 | 0.07 | (-0.06 - 0.21) | 0.31 | -0.02 | (-0.1 - 0.05) | 0.53 |
| Hippocampus | L | 1.91 | 0.15 | 0.31 | -0.02 | (-0.16 - 0.11) | 0.75 | 0.05 | (-0.08 - 0.19) | 0.45 | -0.07 | (-0.15 - 0) | 0.05 |
| | R | **8.61** | **1.87e-04** | **4.14e-03** | -0.04 | (-0.18 - 0.09) | 0.52 | 0.12 | (-0.02 - 0.25) | 0.10 | **-0.16** | **(-0.23 - -0.08)** | **4.04e-05** |
| Lateral Ventricles | L | **5.74** | **3.25e-03** | **0.03** | 0.03 | (-0.11 - 0.16) | 0.68 | -0.10 | (-0.24 - 0.03) | 0.14 | **0.13** | **(0.05 - 0.2)** | **8.52e-04** |
| | R | 3.98 | 0.02 | 0.08 | 0.09 | (-0.05 - 0.22) | 0.21 | -0.02 | (-0.16 - 0.12) | 0.76 | **0.11** | **(0.03 - 0.18)** | **5.47e-03** |
| Nucleus Accumbens | L | 0.93 | 0.40 | 0.56 | -0.09 | (-0.23 - 0.04) | 0.17 | -0.08 | (-0.21 - 0.06) | 0.26 | -0.02 | (-0.09 - 0.06) | 0.67 |
| | R | 0.12 | 0.89 | 0.92 | -0.03 | (-0.16 - 0.11) | 0.70 | -0.03 | (-0.17 - 0.1) | 0.62 | 0.01 | (-0.07 - 0.08) | 0.85 |
| Pallidum | L | 0.32 | 0.72 | 0.83 | 0.01 | (-0.12 - 0.15) | 0.86 | 0.04 | (-0.1 - 0.18) | 0.57 | -0.03 | (-0.1 - 0.05) | 0.48 |
| | R | 0.68 | 0.51 | 0.64 | 0.04 | (-0.09 - 0.18) | 0.54 | 0.07 | (-0.06 - 0.21) | 0.29 | -0.03 | (-0.1 - 0.05) | 0.44 |
| Putamen | L | 0.23 | 0.79 | 0.87 | 0.03 | (-0.1 - 0.17) | 0.62 | 0.05 | (-0.09 - 0.18) | 0.50 | -0.01 | (-0.09 - 0.06) | 0.76 |
| | R | 0.81 | 0.44 | 0.61 | 0.07 | (-0.07 - 0.2) | 0.32 | 0.09 | (-0.05 - 0.22) | 0.21 | -0.02 | (-0.09 - 0.06) | 0.63 |
| Thalamus | L | 1.11 | 0.33 | 0.50 | 0.00 | (-0.14 - 0.14) | 1.00 | 0.06 | (-0.08 - 0.19) | 0.42 | -0.05 | (-0.13 - 0.02) | 0.16 |
| | R | 3.29 | 0.04 | 0.12 | -0.03 | (-0.16 - 0.11) | 0.71 | 0.07 | (-0.06 - 0.21) | 0.29 | **-0.10** | **(-0.17 - -0.02)** | **0.01** |
| **Global** | | | | | | | | | | | | | |
| Estimated Intracranial Volume | N/A | **7.93** | **3.68e-04** | **5.84e-03** | **-0.18** | **(-0.32 - -0.05)** | **8.36e-03** | -0.04 | (-0.18 - 0.09) | 0.53 | **-0.14** | **(-0.21 - -0.06)** | **3.11e-04** |
| Mean Cortical Thickness | | **9.19** | **1.05e-04** | **4.01e-03** | **-0.15** | **(-0.29 - -0.02)** | **0.03** | 0.01 | (-0.13 - 0.14) | 0.92 | **-0.16** | **(-0.23 - -0.08)** | **3.44e-05** |
| Total Surface Area | | **9.86** | **5.39e-05** | **3.31e-03** | **-0.20** | **(-0.33 - -0.06)** | **4.16e-03** | -0.04 | (-0.18 - 0.09) | 0.56 | **-0.16** | **(-0.23 - -0.08)** | **4.91e-05** |

*Results marked in bold are significant at q<0.05 (the overall effect) or at p<0.05 (pairwise comparisons).*

**eTable 16.** CHR-PS+ vs. CHR-PS- vs. HC effect size overview (post-ComBat mega analysis)

| | | Overall Test Statistics | | | CHR-PS+ < HC | | | CHR-PS+ < CHR-PS- | | | CHR-PS- < HC | | |
|---|---|---|---|---|---|---|---|---|---|---|---|---|---|
| Region of Interest | Hemisphere | F | p | q | Cohen's d | 95% Confidence Interval | p | Cohen's d | 95% Confidence Interval | p | Cohen's d | 95% Confidence Interval | p |
| **Cortical Thickness** | | | | | | | | | | | | | |
| Banks of the Superior Temporal Sulcus | L | 3.91 | 0.02 | 0.06 | -0.10 | (-0.24 - 0.03) | 0.13 | 0.00 | (-0.17 - 0.18) | 0.97 | **-0.11** | **(-0.19 - -0.03)** | **7.27e-03** |
| | R | **9.34** | **9.08e-05** | **1.17e-03** | **-0.23** | **(-0.37 - -0.09)** | **9.04e-04** | -0.09 | (-0.26 - 0.09) | 0.21 | **-0.14** | **(-0.22 - -0.07)** | **3.21e-04** |
| Caudal Anterior Cingulate | L | 2.53 | 0.08 | 0.19 | -0.08 | (-0.21 - 0.06) | 0.26 | 0.01 | (-0.17 - 0.18) | 0.90 | **-0.09** | **(-0.16 - -0.01)** | **0.03** |
| | R | 1.41 | 0.24 | 0.39 | -0.07 | (-0.21 - 0.06) | 0.29 | -0.01 | (-0.19 - 0.17) | 0.88 | -0.06 | (-0.14 - 0.02) | 0.12 |
| Caudal Middle Frontal | L | **7.05** | **8.86e-04** | **7.16e-03** | **-0.21** | **(-0.34 - -0.07)** | **2.47e-03** | -0.09 | (-0.27 - 0.08) | 0.19 | **-0.12** | **(-0.2 - -0.04)** | **2.77e-03** |
| | R | **7.21** | **7.51e-04** | **6.75e-03** | **-0.16** | **(-0.3 - -0.03)** | **0.02** | -0.02 | (-0.19 - 0.16) | 0.79 | **-0.14** | **(-0.22 - -0.06)** | **3.91e-04** |
| Cuneus | L | 0.61 | 0.54 | 0.67 | -0.05 | (-0.19 - 0.08) | 0.43 | -0.08 | (-0.25 - 0.1) | 0.28 | 0.02 | (-0.06 - 0.1) | 0.61 |
| | R | 1.28 | 0.28 | 0.43 | -0.09 | (-0.23 - 0.04) | 0.19 | -0.04 | (-0.22 - 0.14) | 0.55 | -0.05 | (-0.13 - 0.03) | 0.21 |
| Entorhinal | L | 0.89 | 0.41 | 0.54 | -0.09 | (-0.22 - 0.05) | 0.20 | -0.09 | (-0.26 - 0.09) | 0.20 | 0.00 | (-0.08 - 0.08) | 0.98 |
| | R | 1.99 | 0.14 | 0.26 | **-0.14** | **(-0.27 - 0)** | **0.05** | -0.10 | (-0.28 - 0.07) | 0.15 | -0.04 | (-0.11 - 0.04) | 0.37 |
| Frontal Pole | L | 1.45 | 0.23 | 0.38 | -0.11 | (-0.25 - 0.02) | 0.11 | -0.07 | (-0.24 - 0.11) | 0.32 | -0.04 | (-0.12 - 0.04) | 0.30 |
| | R | 2.26 | 0.10 | 0.22 | -0.13 | (-0.26 - 0.01) | 0.07 | -0.07 | (-0.24 - 0.11) | 0.35 | -0.06 | (-0.14 - 0.02) | 0.12 |
| Fusiform | L | **12.41** | **4.33e-06** | **1.68e-04** | **-0.29** | **(-0.42 - -0.15)** | **2.96e-05** | **-0.14** | **(-0.31 - 0.04)** | **0.05** | **-0.15** | **(-0.23 - -0.07)** | **1.52e-04** |
| | R | **6.42** | **1.66e-03** | **9.19e-03** | **-0.23** | **(-0.36 - -0.09)** | **1.12e-03** | -0.13 | (-0.31 - 0.04) | 0.06 | **-0.09** | **(-0.17 - -0.02)** | **0.02** |
| Inferior Parietal | L | **10.83** | **2.06e-05** | **4.56e-04** | **-0.22** | **(-0.35 - -0.08)** | **1.45e-03** | -0.05 | (-0.23 - 0.12) | 0.44 | **-0.16** | **(-0.24 - -0.09)** | **3.20e-05** |
| | R | **5.82** | **3.01e-03** | **0.01** | -0.10 | (-0.24 - 0.03) | 0.13 | 0.03 | (-0.14 - 0.21) | 0.66 | **-0.13** | **(-0.21 - -0.06)** | **7.76e-04** |
| Inferior Temporal | L | **6.03** | **2.43e-03** | **0.01** | **-0.19** | **(-0.32 - -0.05)** | **6.73e-03** | -0.07 | (-0.25 - 0.1) | 0.29 | **-0.11** | **(-0.19 - -0.04)** | **4.28e-03** |
| | R | **9.37** | **8.76e-05** | **1.17e-03** | **-0.17** | **(-0.3 - -0.03)** | **0.02** | 0.00 | (-0.18 - 0.17) | 0.96 | **-0.16** | **(-0.24 - -0.09)** | **3.73e-05** |
| Insula | L | **11.21** | **1.42e-05** | **3.66e-04** | **-0.19** | **(-0.33 - -0.06)** | **5.44e-03** | -0.01 | (-0.19 - 0.16) | 0.85 | **-0.18** | **(-0.25 - -0.1)** | **8.07e-06** |
| | R | **9.59** | **7.06e-05** | **1.17e-03** | **-0.19** | **(-0.32 - -0.05)** | **6.83e-03** | -0.02 | (-0.2 - 0.15) | 0.72 | **-0.16** | **(-0.24 - -0.08)** | **4.67e-05** |
| Isthmus Cingulate | L | 0.30 | 0.74 | 0.86 | -0.05 | (-0.18 - 0.09) | 0.50 | -0.02 | (-0.2 - 0.15) | 0.72 | -0.02 | (-0.1 - 0.06) | 0.58 |
| | R | 1.90 | 0.15 | 0.27 | -0.05 | (-0.18 - 0.09) | 0.51 | 0.03 | (-0.14 - 0.21) | 0.64 | -0.08 | (-0.15 - 0) | 0.05 |
| Lateral Occipital | L | **6.56** | **1.44e-03** | **8.59e-03** | **-0.20** | **(-0.34 - -0.07)** | **3.04e-03** | -0.09 | (-0.27 - 0.08) | 0.19 | **-0.11** | **(-0.19 - -0.03)** | **4.50e-03** |
| | R | **9.95** | **4.96e-05** | **9.62e-04** | **-0.22** | **(-0.35 - -0.08)** | **1.63e-03** | -0.06 | (-0.24 - 0.11) | 0.38 | **-0.16** | **(-0.23 - -0.08)** | **8.69e-05** |
| | | | | | | **(-0.29 -** | | | (-0.26 - | | | | |

| | | | | | | | | | | | | | |
|---|---|---|---|---|---|---|---|---|---|---|---|---|---|
| Lateral Orbitofrontal | L | 3.35 | 0.04 | 0.09 | **-0.15** | **-0.02)** | **0.02** | -0.08 | 0.09) | 0.25 | -0.07 | (-0.15 - 0) | 0.06 |
| | R | 2.53 | 0.08 | 0.19 | -0.08 | (-0.22 - 0.05) | 0.23 | 0.00 | (-0.17 - 0.18) | 0.95 | **-0.09** | **(-0.16 - -0.01)** | **0.03** |
| Lingual | L | 0.55 | 0.58 | 0.71 | -0.04 | (-0.18 - 0.09) | 0.53 | 0.00 | (-0.18 - 0.17) | 0.95 | -0.04 | (-0.12 - 0.04) | 0.33 |
| | R | 0.04 | 0.96 | 0.99 | -0.02 | (-0.16 - 0.12) | 0.78 | -0.01 | (-0.19 - 0.16) | 0.85 | -0.01 | (-0.08 - 0.07) | 0.88 |
| Medial Orbitofrontal | L | **7.00** | **9.23e-04** | **7.16e-03** | -0.08 | (-0.21 - 0.06) | 0.27 | 0.07 | (-0.1 - 0.25) | 0.29 | **-0.15** | **(-0.23 - -0.07)** | **1.86e-04** |
| | R | **4.53** | **0.01** | **0.04** | -0.10 | (-0.24 - 0.03) | 0.13 | 0.01 | (-0.16 - 0.19) | 0.86 | **-0.12** | **(-0.19 - -0.04)** | **3.43e-03** |
| Middle Temporal | L | 3.74 | 0.02 | 0.07 | **-0.17** | **(-0.31 - -0.04)** | **0.01** | -0.10 | (-0.27 - 0.08) | 0.15 | -0.07 | (-0.15 - 0.01) | 0.07 |
| | R | **13.73** | **1.16e-06** | **1.19e-04** | **-0.27** | **(-0.41 - -0.14)** | **8.29e-05** | -0.09 | (-0.27 - 0.08) | 0.17 | **-0.18** | **(-0.25 - -0.1)** | **9.15e-06** |
| Paracentral | L | **12.18** | **5.42e-06** | **1.68e-04** | **-0.32** | **(-0.45 - -0.18)** | **4.60e-06** | **-0.20** | **(-0.37 - -0.02)** | **4.76e-03** | **-0.12** | **(-0.2 - -0.04)** | **2.25e-03** |
| | R | **8.88** | **1.43e-04** | **1.71e-03** | **-0.28** | **(-0.42 - -0.15)** | **4.51e-05** | **-0.20** | **(-0.37 - -0.02)** | **4.59e-03** | **-0.09** | **(-0.16 - -0.01)** | **0.03** |
| Parahippocampal | L | **6.79** | **1.14e-03** | **7.59e-03** | -0.13 | (-0.26 - 0.01) | 0.06 | 0.01 | (-0.16 - 0.19) | 0.83 | **-0.14** | **(-0.22 - -0.06)** | **3.45e-04** |
| | R | 1.66 | 0.19 | 0.33 | -0.08 | (-0.21 - 0.06) | 0.26 | -0.01 | (-0.18 - 0.16) | 0.88 | -0.07 | (-0.14 - 0.01) | 0.09 |
| Pars Opercularis | L | 0.96 | 0.38 | 0.52 | -0.03 | (-0.16 - 0.11) | 0.68 | 0.03 | (-0.15 - 0.2) | 0.69 | -0.05 | (-0.13 - 0.02) | 0.17 |
| | R | 2.42 | 0.09 | 0.19 | -0.12 | (-0.25 - 0.02) | 0.09 | -0.04 | (-0.22 - 0.13) | 0.53 | -0.07 | (-0.15 - 0) | 0.07 |
| Pars Orbitalis | L | 1.39 | 0.25 | 0.40 | -0.10 | (-0.23 - 0.03) | 0.15 | -0.05 | (-0.23 - 0.12) | 0.45 | -0.05 | (-0.12 - 0.03) | 0.23 |
| | R | 0.30 | 0.74 | 0.86 | -0.05 | (-0.19 - 0.08) | 0.46 | -0.04 | (-0.21 - 0.14) | 0.61 | -0.02 | (-0.09 - 0.06) | 0.68 |
| Pars Triangularis | L | 0.13 | 0.87 | 0.93 | -0.02 | (-0.15 - 0.12) | 0.81 | -0.03 | (-0.21 - 0.14) | 0.65 | 0.01 | (-0.06 - 0.09) | 0.71 |
| | R | 2.05 | 0.13 | 0.25 | -0.12 | (-0.25 - 0.02) | 0.09 | -0.06 | (-0.23 - 0.12) | 0.42 | -0.06 | (-0.14 - 0.02) | 0.12 |
| Pericalcarine | L | 0.14 | 0.87 | 0.93 | 0.02 | (-0.12 - 0.15) | 0.79 | 0.00 | (-0.18 - 0.17) | 0.98 | 0.02 | (-0.06 - 0.1) | 0.60 |
| | R | 0.70 | 0.50 | 0.63 | 0.04 | (-0.09 - 0.18) | 0.54 | 0.00 | (-0.18 - 0.17) | 0.96 | 0.05 | (-0.03 - 0.12) | 0.25 |
| Postcentral | L | 3.83 | 0.02 | 0.07 | -0.13 | (-0.27 - 0) | 0.06 | -0.03 | (-0.21 - 0.14) | 0.65 | **-0.10** | **(-0.18 - -0.02)** | **0.01** |
| | R | **6.38** | **1.72e-03** | **9.21e-03** | **-0.20** | **(-0.34 - -0.07)** | **3.86e-03** | -0.09 | (-0.27 - 0.09) | 0.21 | **-0.11** | **(-0.19 - -0.04)** | **4.57e-03** |
| Posterior Cingulate | L | 3.57 | 0.03 | 0.08 | -0.07 | (-0.21 - 0.06) | 0.28 | 0.03 | (-0.14 - 0.21) | 0.65 | **-0.10** | **(-0.18 - -0.03)** | **8.04e-03** |
| | R | **6.76** | **1.17e-03** | **7.59e-03** | **-0.19** | **(-0.33 - -0.06)** | **5.73e-03** | -0.07 | (-0.24 - 0.11) | 0.33 | **-0.12** | **(-0.2 - -0.05)** | **1.83e-03** |
| Precentral | L | **6.63** | **1.34e-03** | **8.34e-03** | -0.13 | (-0.27 - 0) | 0.06 | 0.01 | (-0.17 - 0.19) | 0.91 | **-0.14** | **(-0.22 - -0.06)** | **4.37e-04** |
| | R | **7.67** | **4.76e-04** | **4.92e-03** | **-0.22** | **(-0.36 - -0.09)** | **1.37e-03** | -0.10 | (-0.28 - 0.08) | 0.16 | **-0.12** | **(-0.2 - -0.04)** | **2.06e-03** |
| Precuneus | L | **4.84** | **7.99e-03** | **0.03** | -0.11 | (-0.25 - 0.02) | 0.11 | 0.01 | (-0.17 - 0.18) | 0.90 | **-0.12** | **(-0.2 - -0.04)** | **2.63e-03** |
| | R | **9.40** | **8.55e-05** | **1.17e-03** | **-0.17** | **(-0.3 - -0.03)** | **0.02** | 0.00 | (-0.18 - 0.18) | 1.00 | **-0.16** | **(-0.24 - -0.09)** | **3.43e-05** |
| Rostral Anterior | L | **5.13** | **5.96e-03** | **0.03** | **-0.17** | **(-0.31 - -0.04)** | **0.01** | -0.07 | (-0.24 - 0.11) | 0.34 | **-0.11** | **(-0.18 - -0.03)** | **8.06e-03** |

| | | | | | | | | | | | | | |
|---|---|---|---|---|---|---|---|---|---|---|---|---|---|
| Cingulate | R | 3.68 | 0.03 | 0.07 | -0.14 | (-0.27 - 0) | 0.05 | -0.04 | (-0.22 - 0.14) | 0.56 | **-0.09** | **(-0.17 - -0.02)** | **0.02** |
| Rostral Middle Frontal | L | **7.18** | **7.77e-04** | **6.75e-03** | **-0.19** | **(-0.32 - -0.05)** | **7.37e-03** | -0.05 | (-0.23 - 0.12) | 0.45 | **-0.13** | **(-0.21 - -0.05)** | **8.63e-04** |
| | R | **6.48** | **1.55e-03** | **8.90e-03** | **-0.19** | **(-0.33 - -0.06)** | **5.78e-03** | -0.07 | (-0.25 - 0.1) | 0.31 | **-0.12** | **(-0.2 - -0.04)** | **2.63e-03** |
| Superior Frontal | L | **5.43** | **4.45e-03** | **0.02** | -0.13 | (-0.27 - 0) | 0.05 | -0.01 | (-0.19 - 0.16) | 0.87 | **-0.12** | **(-0.2 - -0.05)** | **1.95e-03** |
| | R | **4.51** | **0.01** | **0.04** | **-0.15** | **(-0.29 - -0.02)** | **0.03** | -0.05 | (-0.23 - 0.12) | 0.47 | **-0.10** | **(-0.18 - -0.02)** | **0.01** |
| Superior Parietal | L | **4.92** | **7.37e-03** | **0.03** | **-0.16** | **(-0.29 - -0.02)** | **0.02** | -0.05 | (-0.22 - 0.13) | 0.48 | **-0.11** | **(-0.19 - -0.03)** | **6.53e-03** |
| | R | **5.87** | **2.87e-03** | **0.01** | -0.13 | (-0.26 - 0.01) | 0.07 | 0.01 | (-0.17 - 0.18) | 0.94 | **-0.13** | **(-0.21 - -0.05)** | **9.76e-04** |
| Superior Temporal | L | **6.88** | **1.05e-03** | **7.37e-03** | **-0.23** | **(-0.36 - -0.09)** | **9.44e-04** | -0.13 | (-0.3 - 0.05) | 0.07 | **-0.10** | **(-0.18 - -0.02)** | **0.01** |
| | R | **12.28** | **4.88e-06** | **1.68e-04** | **-0.31** | **(-0.44 - -0.17)** | **9.93e-06** | **-0.17** | **(-0.35 - 0)** | **0.01** | **-0.14** | **(-0.21 - -0.06)** | **6.49e-04** |
| Supramarginal | L | **4.28** | **0.01** | **0.05** | -0.11 | (-0.25 - 0.02) | 0.10 | 0.00 | (-0.18 - 0.18) | 0.98 | **-0.11** | **(-0.19 - -0.03)** | **5.22e-03** |
| | R | 4.08 | 0.02 | 0.06 | -0.11 | (-0.25 - 0.02) | 0.10 | -0.01 | (-0.18 - 0.17) | 0.94 | **-0.11** | **(-0.19 - -0.03)** | **6.68e-03** |
| Temporal Pole | L | 0.94 | 0.39 | 0.53 | -0.09 | (-0.22 - 0.05) | 0.21 | -0.05 | (-0.22 - 0.13) | 0.47 | -0.04 | (-0.11 - 0.04) | 0.36 |
| | R | 2.05 | 0.13 | 0.25 | -0.10 | (-0.23 - 0.04) | 0.16 | -0.03 | (-0.2 - 0.15) | 0.70 | -0.07 | (-0.15 - 0.01) | 0.07 |
| Transverse Temporal | L | 3.29 | 0.04 | 0.10 | **-0.15** | **(-0.28 - -0.01)** | **0.03** | -0.07 | (-0.25 - 0.1) | 0.30 | -0.08 | (-0.15 - 0) | 0.05 |
| | R | **6.90** | **1.02e-03** | **7.37e-03** | **-0.22** | **(-0.35 - -0.08)** | **1.58e-03** | -0.11 | (-0.28 - 0.07) | 0.12 | **-0.11** | **(-0.19 - -0.03)** | **5.45e-03** |
| **Surface Area** | | | | | | | | | | | | | |
| Banks of the Superior Temporal Sulcus | L | 2.21 | 0.11 | 0.22 | -0.02 | (-0.15 - 0.12) | 0.81 | 0.07 | (-0.11 - 0.24) | 0.34 | **-0.08** | **(-0.16 - 0)** | **0.04** |
| | R | 4.08 | 0.02 | 0.06 | **-0.19** | **(-0.33 - -0.06)** | **5.88e-03** | -0.13 | (-0.31 - 0.04) | 0.06 | -0.06 | (-0.14 - 0.02) | 0.13 |
| Caudal Anterior Cingulate | L | 0.32 | 0.73 | 0.86 | 0.04 | (-0.09 - 0.18) | 0.53 | 0.06 | (-0.12 - 0.23) | 0.43 | -0.01 | (-0.09 - 0.07) | 0.77 |
| | R | 3.32 | 0.04 | 0.09 | -0.02 | (-0.15 - 0.12) | 0.79 | 0.08 | (-0.09 - 0.26) | 0.23 | **-0.10** | **(-0.18 - -0.02)** | **0.01** |
| Caudal Middle Frontal | L | 1.61 | 0.20 | 0.34 | 0.07 | (-0.06 - 0.21) | 0.30 | 0.12 | (-0.06 - 0.29) | 0.10 | -0.04 | (-0.12 - 0.03) | 0.26 |
| | R | 1.25 | 0.29 | 0.43 | 0.09 | (-0.05 - 0.22) | 0.21 | 0.11 | (-0.07 - 0.29) | 0.11 | -0.02 | (-0.1 - 0.06) | 0.58 |
| Cuneus | L | 0.30 | 0.74 | 0.86 | -0.05 | (-0.19 - 0.09) | 0.48 | -0.03 | (-0.21 - 0.15) | 0.67 | -0.02 | (-0.1 - 0.06) | 0.61 |
| | R | 1.54 | 0.22 | 0.35 | -0.08 | (-0.22 - 0.06) | 0.25 | -0.02 | (-0.19 - 0.16) | 0.82 | -0.06 | (-0.14 - 0.01) | 0.11 |
| Entorhinal | L | 3.79 | 0.02 | 0.07 | -0.10 | (-0.24 - 0.03) | 0.14 | 0.00 | (-0.17 - 0.18) | 0.95 | **-0.11** | **(-0.18 - -0.03)** | **8.06e-03** |
| | R | 1.25 | 0.29 | 0.43 | 0.05 | (-0.08 - 0.19) | 0.44 | 0.10 | (-0.08 - 0.27) | 0.16 | -0.04 | (-0.12 - 0.03) | 0.27 |
| Frontal Pole | L | 1.93 | 0.14 | 0.27 | 0.13 | (0 - 0.27) | 0.05 | 0.12 | (-0.05 - 0.3) | 0.08 | 0.01 | (-0.06 - 0.09) | 0.71 |
| | R | 1.20 | 0.30 | 0.44 | 0.07 | (-0.07 - 0.2) | 0.31 | 0.01 | (-0.16 - 0.19) | 0.85 | 0.06 | (-0.02 - 0.13) | 0.16 |
| Fusiform | L | 0.80 | 0.45 | 0.58 | 0.01 | (-0.12 - 0.14) | 0.88 | 0.06 | (-0.12 - 0.23) | 0.41 | -0.05 | (-0.12 - 0.03) | 0.25 |

| | | | | | | | | | | | | | |
|---|---|---|---|---|---|---|---|---|---|---|---|---|---|
| | R | 1.57 | 0.21 | 0.35 | -0.05 | (-0.18 - 0.09) | 0.50 | 0.02 | (-0.15 - 0.2) | 0.73 | -0.07 | (-0.15 - 0.01) | 0.08 |
| Inferior Parietal | L | 3.62 | 0.03 | 0.07 | 0.08 | (-0.06 - 0.21) | 0.26 | **0.16** | **(-0.02 - 0.33)** | **0.02** | **-0.08** | **(-0.16 - 0)** | **0.04** |
| | R | 3.95 | 0.02 | 0.06 | -0.05 | (-0.19 - 0.08) | 0.44 | 0.06 | (-0.11 - 0.23) | 0.39 | **-0.11** | **(-0.19 - -0.03)** | **4.99e-03** |
| Inferior Temporal | L | 1.18 | 0.31 | 0.44 | 0.04 | (-0.09 - 0.18) | 0.55 | 0.09 | (-0.09 - 0.26) | 0.20 | -0.05 | (-0.12 - 0.03) | 0.24 |
| | R | 2.26 | 0.10 | 0.22 | 0.09 | (-0.04 - 0.23) | 0.17 | **0.14** | **(-0.03 - 0.32)** | **0.04** | -0.05 | (-0.12 - 0.03) | 0.24 |
| Insula | L | 1.97 | 0.14 | 0.26 | -0.13 | (-0.26 - 0.01) | 0.06 | -0.08 | (-0.26 - 0.09) | 0.24 | -0.05 | (-0.13 - 0.03) | 0.23 |
| | R | 0.83 | 0.43 | 0.57 | -0.09 | (-0.22 - 0.05) | 0.21 | -0.06 | (-0.24 - 0.11) | 0.36 | -0.02 | (-0.1 - 0.05) | 0.54 |
| Isthmus Cingulate | L | 0.27 | 0.77 | 0.87 | -0.04 | (-0.17 - 0.1) | 0.61 | -0.05 | (-0.22 - 0.13) | 0.48 | 0.01 | (-0.06 - 0.09) | 0.73 |
| | R | **4.44** | **0.01** | **0.05** | 0.12 | (-0.02 - 0.25) | 0.08 | 0.01 | (-0.17 - 0.18) | 0.91 | **0.11** | **(0.03 - 0.19)** | **4.85e-03** |
| Lateral Occipital | L | 0.03 | 0.97 | 0.99 | 0.01 | (-0.12 - 0.15) | 0.86 | 0.00 | (-0.17 - 0.18) | 0.96 | 0.01 | (-0.07 - 0.09) | 0.83 |
| | R | 0.02 | 0.98 | 1.00 | 0.00 | (-0.14 - 0.13) | 0.96 | -0.01 | (-0.18 - 0.16) | 0.88 | 0.01 | (-0.07 - 0.08) | 0.87 |
| Lateral Orbitofrontal | L | 0.21 | 0.81 | 0.90 | 0.01 | (-0.12 - 0.15) | 0.87 | 0.03 | (-0.14 - 0.21) | 0.63 | -0.02 | (-0.1 - 0.06) | 0.58 |
| | R | 1.86 | 0.16 | 0.28 | 0.09 | (-0.05 - 0.22) | 0.20 | 0.13 | (-0.05 - 0.31) | 0.06 | -0.04 | (-0.12 - 0.04) | 0.31 |
| Lingual | L | 2.08 | 0.12 | 0.25 | **-0.14** | **(-0.27 - 0)** | **0.05** | -0.10 | (-0.28 - 0.07) | 0.15 | -0.04 | (-0.12 - 0.04) | 0.34 |
| | R | 2.20 | 0.11 | 0.22 | -0.13 | (-0.27 - 0.01) | 0.06 | **-0.15** | **(-0.32 - 0.03)** | **0.04** | 0.02 | (-0.06 - 0.09) | 0.70 |
| Medial Orbitofrontal | L | 1.93 | 0.15 | 0.27 | 0.10 | (-0.03 - 0.23) | 0.15 | 0.13 | (-0.04 - 0.31) | 0.05 | -0.03 | (-0.11 - 0.04) | 0.40 |
| | R | 2.39 | 0.09 | 0.20 | -0.05 | (-0.19 - 0.08) | 0.45 | 0.04 | (-0.14 - 0.21) | 0.61 | **-0.09** | **(-0.16 - -0.01)** | **0.03** |
| Middle Temporal | L | 0.25 | 0.78 | 0.88 | 0.05 | (-0.09 - 0.18) | 0.49 | 0.05 | (-0.13 - 0.22) | 0.52 | 0.00 | (-0.07 - 0.08) | 0.94 |
| | R | 2.59 | 0.08 | 0.18 | **-0.14** | **(-0.27 - 0)** | **0.05** | -0.07 | (-0.24 - 0.11) | 0.32 | -0.07 | (-0.14 - 0.01) | 0.10 |
| Paracentral | L | 1.16 | 0.31 | 0.45 | 0.10 | (-0.04 - 0.23) | 0.16 | 0.10 | (-0.07 - 0.28) | 0.13 | -0.01 | (-0.08 - 0.07) | 0.85 |
| | R | 0.41 | 0.66 | 0.79 | 0.04 | (-0.1 - 0.17) | 0.61 | 0.00 | (-0.17 - 0.18) | 0.99 | 0.03 | (-0.04 - 0.11) | 0.39 |
| Parahippocampal | L | 0.80 | 0.45 | 0.58 | -0.04 | (-0.18 - 0.09) | 0.53 | 0.01 | (-0.17 - 0.18) | 0.93 | -0.05 | (-0.13 - 0.03) | 0.22 |
| | R | 1.37 | 0.25 | 0.40 | 0.01 | (-0.13 - 0.14) | 0.89 | -0.06 | (-0.23 - 0.12) | 0.43 | 0.06 | (-0.01 - 0.14) | 0.11 |
| Pars Opercularis | L | 2.42 | 0.09 | 0.19 | -0.06 | (-0.19 - 0.07) | 0.38 | 0.03 | (-0.15 - 0.2) | 0.69 | **-0.09** | **(-0.16 - -0.01)** | **0.03** |
| | R | 0.01 | 0.99 | 1.00 | 0.00 | (-0.13 - 0.14) | 0.96 | 0.01 | (-0.17 - 0.18) | 0.92 | 0.00 | (-0.08 - 0.07) | 0.92 |
| Pars Orbitalis | L | 1.08 | 0.34 | 0.48 | 0.10 | (-0.04 - 0.23) | 0.16 | 0.10 | (-0.08 - 0.27) | 0.16 | 0.00 | (-0.08 - 0.08) | 1.00 |
| | R | 0.96 | 0.38 | 0.52 | 0.10 | (-0.04 - 0.23) | 0.17 | 0.08 | (-0.09 - 0.26) | 0.23 | 0.01 | (-0.06 - 0.09) | 0.74 |
| Pars Triangularis | L | 1.26 | 0.28 | 0.43 | -0.09 | (-0.22 - 0.05) | 0.20 | -0.04 | (-0.21 - 0.14) | 0.58 | -0.05 | (-0.13 - 0.03) | 0.20 |
| | R | 1.07 | 0.34 | 0.48 | 0.10 | (-0.03 - | 0.15 | 0.09 | (-0.08 - | 0.19 | 0.01 | (-0.07 - | 0.82 |

| | | | | | | | | | | | | | |
|---|---|---|---|---|---|---|---|---|---|---|---|---|---|
| | | | | | | 0.24) | | | 0.27) | | | 0.09) | |
| Pericalcarine | L | 1.25 | 0.29 | 0.43 | -0.11 | (-0.24 - 0.03) | 0.12 | -0.08 | (-0.25 - 0.1) | 0.28 | -0.03 | (-0.11 - 0.05) | 0.43 |
| | R | 4.20 | 0.02 | 0.05 | **-0.18** | **(-0.32 - -0.05)** | **9.21e-03** | -0.10 | (-0.28 - 0.07) | 0.14 | -0.08 | (-0.16 - 0) | 0.05 |
| Postcentral | L | 0.50 | 0.60 | 0.73 | 0.06 | (-0.07 - 0.2) | 0.36 | 0.07 | (-0.11 - 0.25) | 0.32 | 0.00 | (-0.08 - 0.07) | 0.91 |
| | R | 0.69 | 0.50 | 0.63 | 0.02 | (-0.12 - 0.16) | 0.79 | 0.06 | (-0.12 - 0.24) | 0.39 | -0.04 | (-0.12 - 0.04) | 0.31 |
| Posterior Cingulate | L | 2.43 | 0.09 | 0.19 | 0.08 | (-0.06 - 0.21) | 0.27 | **0.14** | **(-0.04 - 0.31)** | **0.05** | -0.06 | (-0.14 - 0.02) | 0.13 |
| | R | 3.05 | 0.05 | 0.12 | 0.05 | (-0.08 - 0.19) | 0.46 | 0.13 | (-0.04 - 0.31) | 0.05 | **-0.08** | **(-0.16 - 0)** | **0.04** |
| Precentral | L | 1.41 | 0.24 | 0.39 | 0.09 | (-0.04 - 0.23) | 0.18 | 0.04 | (-0.14 - 0.21) | 0.58 | 0.05 | (-0.02 - 0.13) | 0.18 |
| | R | 2.27 | 0.10 | 0.22 | **0.14** | **(0.01 - 0.28)** | **0.04** | **0.14** | **(-0.04 - 0.32)** | **0.04** | 0.00 | (-0.07 - 0.08) | 0.93 |
| Precuneus | L | 0.47 | 0.62 | 0.75 | -0.01 | (-0.14 - 0.13) | 0.93 | 0.03 | (-0.14 - 0.21) | 0.64 | -0.04 | (-0.12 - 0.04) | 0.34 |
| | R | 0.19 | 0.82 | 0.90 | -0.02 | (-0.16 - 0.11) | 0.72 | 0.00 | (-0.18 - 0.17) | 0.99 | -0.02 | (-0.1 - 0.05) | 0.55 |
| Rostral Anterior Cingulate | L | 4.08 | 0.02 | 0.06 | -0.06 | (-0.2 - 0.07) | 0.36 | 0.05 | (-0.12 - 0.23) | 0.46 | **-0.11** | **(-0.19 - -0.04)** | **4.35e-03** |
| | R | 2.97 | 0.05 | 0.13 | 0.05 | (-0.09 - 0.18) | 0.48 | 0.13 | (-0.05 - 0.31) | 0.06 | **-0.08** | **(-0.16 - 0)** | **0.04** |
| Rostral Middle Frontal | L | 2.44 | 0.09 | 0.19 | 0.12 | (-0.02 - 0.25) | 0.09 | **0.15** | **(-0.02 - 0.33)** | **0.03** | -0.03 | (-0.11 - 0.04) | 0.40 |
| | R | 0.93 | 0.39 | 0.53 | 0.06 | (-0.07 - 0.2) | 0.36 | 0.09 | (-0.08 - 0.27) | 0.19 | -0.03 | (-0.11 - 0.05) | 0.48 |
| Superior Frontal | L | 0.77 | 0.46 | 0.59 | 0.06 | (-0.08 - 0.19) | 0.42 | 0.08 | (-0.09 - 0.26) | 0.23 | -0.03 | (-0.1 - 0.05) | 0.51 |
| | R | 2.70 | 0.07 | 0.17 | 0.00 | (-0.14 - 0.13) | 0.98 | 0.09 | (-0.09 - 0.26) | 0.20 | **-0.09** | **(-0.17 - -0.01)** | **0.03** |
| Superior Parietal | L | 0.20 | 0.82 | 0.90 | 0.04 | (-0.09 - 0.18) | 0.53 | 0.04 | (-0.14 - 0.21) | 0.57 | 0.00 | (-0.07 - 0.08) | 0.91 |
| | R | 1.21 | 0.30 | 0.44 | 0.10 | (-0.04 - 0.23) | 0.15 | 0.11 | (-0.07 - 0.28) | 0.13 | 0.00 | (-0.08 - 0.07) | 0.90 |
| Superior Temporal | L | 0.22 | 0.81 | 0.90 | -0.05 | (-0.18 - 0.09) | 0.51 | -0.04 | (-0.22 - 0.13) | 0.56 | 0.00 | (-0.08 - 0.07) | 0.90 |
| | R | 1.64 | 0.20 | 0.33 | -0.12 | (-0.26 - 0.01) | 0.07 | -0.11 | (-0.28 - 0.07) | 0.12 | -0.02 | (-0.09 - 0.06) | 0.68 |
| Supramarginal | L | **4.30** | **0.01** | **0.05** | **-0.18** | **(-0.31 - -0.04)** | **0.01** | -0.09 | (-0.27 - 0.09) | 0.20 | **-0.09** | **(-0.16 - -0.01)** | **0.03** |
| | R | 0.90 | 0.41 | 0.54 | -0.04 | (-0.18 - 0.09) | 0.52 | 0.01 | (-0.17 - 0.18) | 0.91 | -0.05 | (-0.13 - 0.03) | 0.19 |
| Temporal Pole | L | 0.69 | 0.50 | 0.63 | -0.08 | (-0.21 - 0.05) | 0.24 | -0.07 | (-0.24 - 0.1) | 0.31 | -0.01 | (-0.09 - 0.07) | 0.79 |
| | R | 1.77 | 0.17 | 0.30 | 0.13 | (-0.01 - 0.26) | 0.06 | 0.10 | (-0.07 - 0.28) | 0.14 | 0.03 | (-0.05 - 0.1) | 0.50 |
| Transverse Temporal | L | 0.09 | 0.91 | 0.95 | -0.02 | (-0.16 - 0.11) | 0.72 | -0.01 | (-0.19 - 0.16) | 0.87 | -0.01 | (-0.09 - 0.06) | 0.74 |
| | R | 0.00 | 1.00 | 1.00 | 0.00 | (-0.13 - 0.14) | 0.95 | 0.00 | (-0.17 - 0.18) | 0.96 | 0.00 | (-0.08 - 0.08) | 0.98 |
| **Subcortical Volume** | | | | | | | | | | | | | |
| Amygdala | L | 0.15 | 0.86 | 0.93 | -0.02 | (-0.16 - 0.11) | 0.73 | 0.00 | (-0.18 - 0.17) | 0.96 | -0.02 | (-0.1 - 0.06) | 0.61 |

| | | | | | | | | | | | | | |
|---|---|---|---|---|---|---|---|---|---|---|---|---|---|
| | R | 1.64 | 0.19 | 0.33 | -0.07 | (-0.21 - 0.06) | 0.29 | 0.00 | (-0.18 - 0.17) | 0.95 | -0.07 | (-0.15 - 0.01) | 0.09 |
| Caudate | L | 0.28 | 0.76 | 0.87 | 0.04 | (-0.09 - 0.18) | 0.54 | 0.02 | (-0.16 - 0.19) | 0.78 | 0.02 | (-0.05 - 0.1) | 0.56 |
| | R | 0.04 | 0.96 | 0.99 | 0.00 | (-0.14 - 0.14) | 1.00 | 0.01 | (-0.17 - 0.19) | 0.88 | -0.01 | (-0.09 - 0.07) | 0.78 |
| Hippocampus | L | 2.61 | 0.07 | 0.18 | -0.03 | (-0.17 - 0.1) | 0.61 | 0.06 | (-0.12 - 0.23) | 0.42 | **-0.09** | **(-0.17 - -0.01)** | **0.02** |
| | R | **8.38** | **2.35e-04** | **2.60e-03** | **-0.16** | **(-0.29 - -0.02)** | **0.02** | 0.00 | (-0.18 - 0.17) | 0.96 | **-0.15** | **(-0.23 - -0.08)** | **9.76e-05** |
| Lateral Ventricles | L | **5.11** | **6.06e-03** | **0.03** | 0.03 | (-0.1 - 0.17) | 0.62 | -0.09 | (-0.27 - 0.08) | 0.18 | **0.13** | **(0.05 - 0.2)** | **1.55e-03** |
| | R | 3.82 | 0.02 | 0.07 | 0.06 | (-0.08 - 0.19) | 0.42 | -0.06 | (-0.23 - 0.12) | 0.43 | **0.11** | **(0.03 - 0.19)** | **5.73e-03** |
| Nucleus Accumbens | L | 0.59 | 0.55 | 0.68 | -0.07 | (-0.21 - 0.06) | 0.28 | -0.06 | (-0.24 - 0.11) | 0.37 | -0.01 | (-0.09 - 0.06) | 0.73 |
| | R | 0.02 | 0.98 | 1.00 | 0.01 | (-0.13 - 0.14) | 0.92 | 0.01 | (-0.16 - 0.19) | 0.87 | 0.00 | (-0.08 - 0.07) | 0.90 |
| Pallidum | L | 0.20 | 0.82 | 0.90 | -0.02 | (-0.15 - 0.12) | 0.82 | 0.01 | (-0.16 - 0.18) | 0.89 | -0.02 | (-0.1 - 0.05) | 0.53 |
| | R | 1.03 | 0.36 | 0.50 | -0.10 | (-0.23 - 0.04) | 0.15 | -0.08 | (-0.25 - 0.1) | 0.26 | -0.02 | (-0.1 - 0.06) | 0.59 |
| Putamen | L | 0.11 | 0.89 | 0.94 | -0.03 | (-0.16 - 0.11) | 0.67 | -0.03 | (-0.21 - 0.14) | 0.64 | 0.00 | (-0.07 - 0.08) | 0.93 |
| | R | 0.13 | 0.88 | 0.93 | 0.01 | (-0.13 - 0.14) | 0.92 | 0.03 | (-0.15 - 0.2) | 0.72 | -0.02 | (-0.09 - 0.06) | 0.66 |
| Thalamus | L | 1.93 | 0.15 | 0.27 | -0.08 | (-0.21 - 0.06) | 0.26 | 0.00 | (-0.18 - 0.17) | 0.95 | -0.07 | (-0.15 - 0) | 0.06 |
| | R | **4.28** | **0.01** | **0.05** | **-0.14** | **(-0.27 - 0)** | **0.05** | -0.03 | (-0.21 - 0.14) | 0.64 | **-0.10** | **(-0.18 - -0.03)** | **8.62e-03** |
| **Global** | | | | | | | | | | | | | |
| Estimated Intracranial Volume | | **6.18** | **2.10e-03** | **0.01** | **-0.17** | **(-0.3 - -0.03)** | **0.02** | -0.04 | (-0.22 - 0.13) | 0.55 | **-0.12** | **(-0.2 - -0.05)** | **1.69e-03** |
| Mean Cortical Thickness | N/A | **13.45** | **1.54e-06** | **1.19e-04** | **-0.26** | **(-0.4 - -0.13)** | **1.50e-04** | -0.08 | (-0.26 - 0.09) | 0.22 | **-0.18** | **(-0.25 - -0.1)** | **8.07e-06** |
| Total Surface Area | | **7.17** | **7.83e-04** | **6.75e-03** | -0.09 | (-0.23 - 0.04) | 0.17 | 0.06 | (-0.12 - 0.23) | 0.43 | **-0.15** | **(-0.23 - -0.07)** | **1.64e-04** |

*For “Overall Test Statistics”: results marked in bold are significant at q<0.05 For pairwise comparisons: results marked in bold are significant at p<0.05*

**eTable 17.** CHR-PS+ vs. CHR-PS- vs. HC antipsychotic medication effects

| | | Overall stats: Original Results | | Cohens's d Group Difference: Original Results | | | Overall stats: AP-corrected | | Cohens's d Group Difference: AP-corrected | | |
|---|---|---|---|---|---|---|---|---|---|---|---|
| Region of Interest | Hemisphere | F | q | CHR-PS+ < HC | CHR-PS+ < CHR-PS- | CHR-PS- < HC | F | q | CHR-PS+ < HC | CHR-PS+ < CHR-PS- | CHR-PS- < HC |
| **Cortical Thickness** | | | | | | | | | | | |
| Banks of the Superior Temporal Sulcus | R | 9.34 | 1.17e-03 | -0.23 | -0.09 | -0.14 | 8.25 | 1.18e-03 | -0.22 | -0.09 | -0.13 |
| Caudal Middle Frontal | L | 7.05 | 7.16e-03 | -0.21 | -0.09 | -0.12 | 5.75 | 5.44e-03 | -0.20 | -0.09 | -0.11 |
| | R | 7.21 | 6.75e-03 | -0.16 | -0.02 | -0.14 | 6.15 | 4.51e-03 | -0.15 | -0.02 | -0.13 |
| **Fusiform** | L | 12.41 | 1.68e-04 | -0.29 | -0.14 | -0.15 | 10.88 | 1.44e-04 | -0.28 | -0.14 | -0.14 |
| | R | 6.42 | 9.19e-03 | -0.23 | -0.13 | -0.09 | 5.38 | 6.41e-03 | -0.21 | -0.13 | -0.08 |
| Inferior Parietal | L | 10.83 | 4.56e-04 | -0.22 | -0.05 | -0.16 | 9.96 | 2.69e-04 | -0.22 | -0.05 | -0.16 |
| | R | 5.82 | 0.01 | -0.10 | 0.03 | -0.13 | 4.50 | 0.01 | -0.09 | 0.03 | -0.12 |
| Inferior Temporal | L | 6.03 | 0.01 | -0.19 | -0.07 | -0.11 | 5.45 | 6.19e-03 | -0.18 | -0.07 | -0.11 |
| | R | 9.37 | 1.17e-03 | -0.17 | 0.00 | -0.16 | 6.80 | 3.12e-03 | -0.15 | 0.00 | -0.14 |
| Insula | L | 11.21 | 3.66e-04 | -0.19 | -0.01 | -0.18 | 11.69 | 1.21e-04 | -0.20 | -0.01 | -0.18 |
| | R | 9.59 | 1.17e-03 | -0.19 | -0.02 | -0.16 | 8.76 | 7.84e-04 | -0.18 | -0.02 | -0.15 |
| Lateral Occipital | L | 6.56 | 8.59e-03 | -0.20 | -0.09 | -0.11 | 7.27 | 2.60e-03 | -0.22 | -0.09 | -0.12 |
| | R | 9.95 | 9.62e-04 | -0.22 | -0.06 | -0.16 | 10.26 | 2.27e-04 | -0.22 | -0.06 | -0.16 |
| Medial Orbitofrontal | L | 7.00 | 7.16e-03 | -0.08 | 0.07 | -0.15 | 7.04 | 2.81e-03 | -0.08 | 0.07 | -0.15 |
| | R | 4.53 | 0.04 | -0.10 | 0.01 | -0.12 | 4.54 | 0.01 | -0.11 | 0.01 | -0.12 |
| Middle Temporal | R | 13.73 | 1.19e-04 | -0.27 | -0.09 | -0.18 | 11.45 | 1.21e-04 | -0.26 | -0.09 | -0.16 |
| **Paracentral** | L | 12.18 | 1.68e-04 | -0.32 | -0.20 | -0.12 | 11.57 | 1.21e-04 | -0.31 | -0.20 | -0.12 |
| | R | 8.88 | 1.71e-03 | -0.28 | -0.20 | -0.09 | 8.01 | 1.36e-03 | -0.27 | -0.20 | -0.08 |
| Parahippocampal | L | 6.79 | 7.59e-03 | -0.13 | 0.01 | -0.14 | 6.47 | 4.05e-03 | -0.13 | 0.01 | -0.14 |
| Postcentral | R | 6.38 | 9.21e-03 | -0.20 | -0.09 | -0.11 | 6.39 | 4.11e-03 | -0.20 | -0.09 | -0.11 |
| Posterior Cingulate | R | 6.76 | 7.59e-03 | -0.19 | -0.07 | -0.12 | 5.78 | 5.44e-03 | -0.18 | -0.07 | -0.11 |
| Precentral | L | 6.63 | 8.34e-03 | -0.13 | 0.01 | -0.14 | 5.45 | 6.19e-03 | -0.12 | 0.01 | -0.13 |
| | R | 7.67 | 4.92e-03 | -0.22 | -0.10 | -0.12 | 6.89 | 3.05e-03 | -0.22 | -0.10 | -0.12 |
| Precuneus | L | 4.84 | 0.03 | -0.11 | 0.01 | -0.12 | 3.78 | 0.03 | -0.10 | 0.01 | -0.11 |
| | R | 9.40 | 1.17e-03 | -0.17 | 0.00 | -0.16 | 7.06 | 2.81e-03 | -0.15 | 0.00 | -0.14 |
| Rostral Anterior Cingulate | L | 5.13 | 0.03 | -0.17 | -0.07 | -0.11 | 5.48 | 6.19e-03 | -0.18 | -0.07 | -0.11 |
| Rostral Middle Frontal | L | 7.18 | 6.75e-03 | -0.19 | -0.05 | -0.13 | 5.96 | 5.05e-03 | -0.17 | -0.05 | -0.12 |
| | R | 6.48 | 8.90e-03 | -0.19 | -0.07 | -0.12 | 5.87 | 5.22e-03 | -0.19 | -0.07 | -0.11 |
| Superior Frontal | L | 5.43 | 0.02 | -0.13 | -0.01 | -0.12 | 4.16 | 0.02 | -0.12 | -0.01 | -0.11 |
| | R | 4.51 | 0.04 | -0.15 | -0.05 | -0.10 | 4.58 | 0.01 | -0.16 | -0.05 | -0.10 |
| Superior Parietal | L | 4.92 | 0.03 | -0.16 | -0.05 | -0.11 | 4.90 | 9.73e-03 | -0.16 | -0.05 | -0.11 |
| | R | 5.87 | 0.01 | -0.13 | 0.01 | -0.13 | 5.58 | 6.04e-03 | -0.13 | 0.01 | -0.13 |
| **Superior Temporal** | L | 6.88 | 7.37e-03 | -0.23 | -0.13 | -0.10 | 5.57 | 6.04e-03 | -0.21 | -0.13 | -0.09 |
| | R | 12.28 | 1.68e-04 | -0.31 | -0.17 | -0.14 | 11.24 | 1.21e-04 | -0.30 | -0.17 | -0.13 |
| Supramarginal | L | 4.28 | 0.05 | -0.11 | 0.00 | -0.11 | 3.26 | 0.04 | -0.10 | 0.00 | -0.10 |
| Transverse Temporal | R | 6.90 | 7.37e-03 | -0.22 | -0.11 | -0.11 | 6.25 | 4.29e-03 | -0.21 | -0.11 | -0.10 |
| **Surface Area** | | | | | | | | | | | |
| Isthmus Cingulate | R | 4.44 | 0.05 | 0.12 | 0.01 | 0.11 | 2.66 | 0.07 | 0.10 | 0.01 | 0.09 |
| Supramarginal | L | 4.30 | 0.05 | -0.18 | -0.09 | -0.09 | 3.71 | 0.03 | -0.17 | -0.09 | -0.08 |
| **Subcortical Volume** | | | | | | | | | | | |
| Hippocampus | R | 8.38 | 2.60e-03 | -0.16 | 0.00 | -0.15 | 5.17 | 7.63e-03 | -0.13 | 0.00 | -0.12 |
| Lateral Ventricles | L | 5.11 | 0.03 | 0.03 | -0.09 | 0.13 | 4.25 | 0.02 | 0.03 | -0.09 | 0.11 |

| | | | | | | | | | | | |
|---|---|---|---|---|---|---|---|---|---|---|---|
| Thalamus | R | 4.28 | 0.05 | -0.14 | -0.03 | -0.10 | 2.96 | 0.05 | -0.12 | -0.03 | -0.09 |
| **Global** | | | | | | | | | | | |
| Estimated Intracranial Volume | N/A | 6.18 | 0.01 | -0.17 | -0.04 | -0.12 | 6.35 | 4.11e-03 | -0.17 | -0.04 | -0.13 |
| Mean Cortical Thickness | N/A | 13.45 | 1.19e-04 | -0.26 | -0.08 | -0.18 | 12.02 | 1.21e-04 | -0.25 | -0.08 | -0.17 |
| Total Surface Area | N/A | 7.17 | 6.75e-03 | -0.09 | 0.06 | -0.15 | 5.95 | 5.05e-03 | -0.08 | 0.06 | -0.14 |

*Results reported under “Original Results” are from the CHR-PS+ vs. CHR-PS- vs. HC post-ComBat mega analysis (see also eTable 11). All reported regions showed an overall effect significant at $q<0.05$. “AP-corrected” refers to post-ComBat mega analysis including antipsychotic medication as an additional covariate.*

**eTable 18.** CHR-PS+ vs. CHR-PS- vs. HC equivalence testing results

| | | | Equivalence Testing Results | | | | | Equivalence Testing Results | | | | | Equivalence Testing Results | | | |
|---|---|---|---|---|---|---|---|---|---|---|---|---|---|---|---|---|
| | | CHR-PS+ < HC | | | | | CHR-PS+ < CHR-PS- | | | | | CHR-PS- < HC | | | | |
| Region of Interest | Hemisphere | Cohen's d | 90% CI | p ΔL | p ΔU | equivalent to SESOI | Cohen's d | 90% CI | p ΔL | p ΔU | equivalent to SESOI | Cohen's d | 90% CI | p ΔL | p ΔU | equivalent to SESOI |
| **Cortical Thickness** | | | | | | | | | | | | | | | | |
| Banks of the Superior Temporal Sulcus | R | -0.23 | (-0.35 - -0.12) | 0.88 | 2.08e-08 | no | -0.09 | • | • | • | • | -0.14 | (-0.21 - -0.08) | 0.43 | 7.42e-14 | no |
| Caudal Middle Frontal | L | -0.21 | (-0.32 - -0.1) | 0.80 | 9.51e-08 | no | -0.09 | • | • | • | • | -0.12 | (-0.18 - -0.05) | 0.21 | 5.66e-12 | no |
| | R | -0.16 | (-0.27 - -0.05) | 0.56 | 3.38e-06 | no | -0.02 | • | • | • | • | -0.14 | (-0.21 - -0.08) | 0.41 | 1.09e-13 | no |
| **Fusiform** | L | -0.29 | (-0.4 - -0.17) | 0.98 | 1.01e-10 | no | -0.14 | (-0.29 - 0.01) | 0.45 | 6.30e-04 | no | -0.15 | (-0.22 - -0.09) | 0.50 | 1.70e-14 | no |
| | R | -0.23 | (-0.34 - -0.11) | 0.86 | 2.72e-08 | no | -0.13 | • | • | • | • | -0.09 | (-0.16 - -0.03) | 0.07 | 3.78e-10 | no |
| Inferior Parietal | L | -0.22 | (-0.33 - -0.11) | 0.84 | 3.94e-08 | no | -0.05 | • | • | • | • | -0.16 | (-0.23 - -0.1) | 0.65 | 9.10e-16 | no |
| | R | -0.10 | • | • | • | • | 0.03 | • | • | • | • | -0.13 | (-0.2 - -0.07) | 0.34 | 4.32e-13 | no |
| Inferior Temporal | L | -0.19 | (-0.3 - -0.07) | 0.70 | 5.00e-07 | no | -0.07 | • | • | • | • | -0.11 | (-0.18 - -0.05) | 0.18 | 1.43e-11 | no |
| | R | -0.17 | (-0.28 - -0.05) | 0.60 | 2.08e-06 | no | 0.00 | • | • | • | • | -0.16 | (-0.23 - -0.1) | 0.63 | 1.24e-15 | no |
| Insula | L | -0.19 | (-0.3 - -0.08) | 0.73 | 3.37e-07 | no | -0.01 | • | • | • | • | -0.18 | (-0.24 - -0.11) | 0.75 | 7.58e-17 | no |
| | R | -0.19 | (-0.3 - -0.07) | 0.70 | 5.13e-07 | no | -0.02 | • | • | • | • | -0.16 | (-0.23 - -0.1) | 0.61 | 1.82e-15 | no |
| Lateral Occipital | L | -0.20 | (-0.32 - -0.09) | 0.79 | 1.34e-07 | no | -0.09 | • | • | • | • | -0.11 | (-0.18 - -0.05) | 0.17 | 1.59e-11 | no |
| | R | -0.22 | (-0.33 - -0.1) | 0.83 | 4.66e-08 | no | -0.06 | • | • | • | • | -0.16 | (-0.22 - -0.09) | 0.56 | 5.97e-15 | no |
| Medial Orbitofrontal | L | -0.08 | • | • | • | • | 0.07 | • | • | • | • | -0.15 | (-0.21 - -0.08) | 0.48 | 2.56e-14 | no |
| | R | -0.10 | • | • | • | • | 0.01 | • | • | • | • | -0.12 | (-0.18 - -0.05) | 0.19 | 9.06e-12 | no |
| Middle Temporal | R | -0.27 | (-0.38 - -0.16) | 0.96 | 4.62e-10 | no | -0.09 | • | • | • | • | -0.18 | (-0.24 - -0.11) | 0.75 | 1.01e-16 | no |

| | | | | | | | | | | | | | | | | |
|---|---|---|---|---|---|---|---|---|---|---|---|---|---|---|---|---|
| **Paracentral** | L | -0.32 | (-0.43 - -0.2) | 0.99 | 6.98e-12 | no | -0.20 | (-0.34 - -0.05) | 0.70 | 5.50e-05 | no | -0.12 | (-0.19 - -0.06) | 0.23 | 3.72e-12 | no |
| | R | -0.28 | (-0.4 - -0.17) | 0.97 | 1.94e-10 | no | -0.20 | (-0.35 - -0.05) | 0.70 | 5.31e-05 | no | -0.09 | (-0.15 - -0.02) | 0.05 | 1.24e-09 | no |
| Parahippocampal | L | -0.13 | • | • | • | • | 0.01 | • | • | • | • | -0.14 | (-0.21 - -0.08) | 0.42 | 7.97e-14 | no |
| Postcentral | R | -0.20 | (-0.32 - -0.09) | 0.77 | 2.25e-07 | no | -0.09 | • | • | • | • | -0.11 | (-0.18 - -0.05) | 0.18 | 2.02e-11 | no |
| Posterior Cingulate | R | -0.19 | (-0.31 - -0.08) | 0.73 | 4.06e-07 | no | -0.07 | • | • | • | • | -0.12 | (-0.19 - -0.06) | 0.25 | 2.40e-12 | no |
| Precentral | L | -0.13 | • | • | • | • | 0.01 | • | • | • | • | -0.14 | (-0.21 - -0.07) | 0.40 | 1.54e-13 | no |
| | R | -0.22 | (-0.34 - -0.11) | 0.85 | 3.90e-08 | no | -0.10 | • | • | • | • | -0.12 | (-0.19 - -0.06) | 0.25 | 3.59e-12 | no |
| Precuneus | L | -0.11 | • | • | • | • | 0.01 | • | • | • | • | -0.12 | (-0.18 - -0.05) | 0.22 | 5.23e-12 | no |
| | R | -0.17 | (-0.28 - -0.05) | 0.59 | 2.42e-06 | no | 0.00 | • | • | • | • | -0.16 | (-0.23 - -0.1) | 0.64 | 1.07e-15 | no |
| Rostral Anterior Cingulate | L | -0.17 | (-0.28 - -0.06) | 0.62 | 1.51e-06 | no | -0.07 | • | • | • | • | -0.11 | (-0.17 - -0.04) | 0.13 | 5.95e-11 | no |
| Rostral Middle Frontal | L | -0.19 | (-0.3 - -0.07) | 0.70 | 5.93e-07 | no | -0.05 | • | • | • | • | -0.13 | (-0.2 - -0.07) | 0.32 | 4.97e-13 | no |
| | R | -0.19 | (-0.3 - -0.08) | 0.72 | 3.80e-07 | no | -0.07 | • | • | • | • | -0.12 | (-0.18 - -0.05) | 0.22 | 5.01e-12 | no |
| Superior Frontal | L | -0.13 | • | • | • | • | -0.01 | • | • | • | • | -0.12 | (-0.19 - -0.06) | 0.25 | 2.75e-12 | no |
| | R | -0.15 | (-0.27 - -0.04) | 0.52 | 5.28e-06 | no | -0.05 | • | • | • | • | -0.10 | (-0.17 - -0.04) | 0.11 | 9.39e-11 | no |
| Superior Parietal | L | -0.16 | (-0.27 - -0.04) | 0.54 | 4.14e-06 | no | -0.05 | • | • | • | • | -0.11 | (-0.17 - -0.04) | 0.14 | 3.64e-11 | no |
| | R | -0.13 | • | • | • | • | 0.01 | • | • | • | • | -0.13 | (-0.2 - -0.07) | 0.31 | 6.77e-13 | no |
| **Superior Temporal** | L | -0.23 | (-0.34 - -0.11) | 0.87 | 1.96e-08 | no | -0.13 | • | • | • | • | -0.10 | (-0.17 - -0.04) | 0.11 | 1.19e-10 | no |
| | R | -0.31 | (-0.42 - -0.19) | 0.99 | 2.18e-11 | no | -0.17 | (-0.32 - -0.02) | 0.59 | 1.71e-04 | no | -0.14 | (-0.2 - -0.07) | 0.36 | 3.02e-13 | no |
| Supramarginal | L | -0.11 | • | • | • | • | 0.00 | • | • | • | • | -0.11 | (-0.18 - | 0.16 | 2.48e- | no |

| | | | | | | | | | | | | | | | | |
|---|---|---|---|---|---|---|---|---|---|---|---|---|---|---|---|---|
| | | | | | | | | | | | | | -0.05) | | 11 | |
| Transverse Temporal | R | -0.22 | (-0.33 - -0.1) | 0.84 | 4.54e-08 | no | -0.11 | • | • | • | • | -0.11 | (-0.18 - -0.05) | 0.16 | 2.44e-11 | no |
| **Surface Area** | | | | | | | | | | | | | | | | |
| Isthmus Cingulate | R | 0.12 | • | • | • | • | 0.01 | • | • | • | • | 0.11 | (0.05 - 0.18) | 1.92e-11 | 0.17 | no |
| Supramarginal | L | -0.18 | (-0.29 - -0.06) | 0.65 | 1.34e-06 | no | -0.09 | • | • | • | • | -0.09 | (-0.15 - -0.02) | 0.05 | 1.60e-09 | no |
| **Subcortical Volume** | | | | | | | | | | | | | | | | |
| Hippocampus | R | -0.16 | (-0.27 - -0.05) | 0.55 | 3.36e-06 | no | 0.00 | • | • | • | • | -0.15 | (-0.22 - -0.09) | 0.54 | 6.98e-15 | no |
| Lateral Ventricles | L | 0.03 | • | • | • | • | -0.09 | • | • | • | • | 0.13 | (0.06 - 0.19) | 1.98e-12 | 0.27 | no |
| Thalamus | R | -0.14 | (-0.25 - -0.02) | 0.42 | 1.46e-05 | no | -0.03 | • | • | • | • | -0.10 | (-0.17 - -0.04) | 0.12 | 6.64e-11 | no |
| **Global** | | | | | | | | | | | | | | | | |
| Estimated Intracranial Volume | N/A | -0.17 | (-0.28 - -0.05) | 0.59 | 2.10e-06 | no | -0.04 | • | • | • | • | -0.12 | (-0.19 - -0.06) | 0.25 | 1.81e-12 | no |
| Mean Cortical Thickness | N/A | -0.26 | (-0.38 - -0.15) | 0.95 | 1.18e-09 | no | -0.08 | • | • | • | • | -0.18 | (-0.24 - -0.11) | 0.75 | 7.26e-17 | no |
| Total Surface Area | N/A | -0.09 | • | • | • | • | 0.06 | • | • | • | • | -0.15 | (-0.21 - -0.08) | 0.49 | 1.83e-14 | no |

*ΔU/L refer to upper (+0.15) and lower (-0.15) equivalence bounds. SESOI = smallest effect size of interest. All regions of interest showed an overall effect of conversion status at q<0.05 in the CHR-PS+ vs. CHR-PS- vs. HC post-ComBat mega analysis (see also eTable 11). Regions that failed to show a group difference at p<0.05 in pairwise comparisons are marked as "•".*

**eTable 19.** CHR subgroup-by-psychosis conversion interaction analyses effect size overview

| | | all-by-conversion | | | APS-by-conversion | | | BIPS-by-conversion | | | GRD-by-conversion | | |
|---|---|---|---|---|---|---|---|---|---|---|---|---|---|
| **Region of Interest** | **Hemisphere** | **F** | **p** | **q** | **F** | **p** | **q** | **F** | **p** | **q** | **F** | **p** | **q** |
| **Cortical Thickness** | | | | | | | | | | | | | |
| Banks of the Superior Temporal Sulcus | L | 0.96 | 0.38 | 0.86 | 0.43 | 0.51 | 0.94 | 1.33 | 0.25 | 0.98 | 6.64 | 0.01 | 0.35 |
| | R | 0.47 | 0.62 | 0.91 | 0.41 | 0.52 | 0.94 | 0.01 | 0.91 | 0.98 | 1.68 | 0.20 | 0.64 |
| Caudal Anterior Cingulate | L | 3.74 | 0.02 | 0.75 | 2.51 | 0.11 | 0.84 | 0.35 | 0.56 | 0.98 | 0.10 | 0.75 | 0.90 |
| | R | 0.12 | 0.88 | 0.96 | 0.02 | 0.89 | 0.98 | 1.83 | 0.18 | 0.98 | 0.76 | 0.38 | 0.70 |
| Caudal Middle Frontal | L | 1.61 | 0.20 | 0.75 | 3.66 | 0.06 | 0.74 | 0.18 | 0.67 | 0.98 | 1.70 | 0.19 | 0.64 |
| | R | 3.54 | 0.03 | 0.75 | 6.80 | 9.16e-03 | 0.63 | 0.77 | 0.38 | 0.98 | 9.66 | 1.90e-03 | 0.21 |
| Cuneus | L | 2.11 | 0.12 | 0.75 | 2.96 | 0.09 | 0.82 | 0.01 | 0.91 | 0.98 | 4.22 | 0.04 | 0.48 |
| | R | 2.17 | 0.11 | 0.75 | 1.91 | 0.17 | 0.87 | 0.03 | 0.87 | 0.98 | 2.06 | 0.15 | 0.61 |
| Entorhinal | L | 1.86 | 0.16 | 0.75 | 0.37 | 0.55 | 0.94 | 1.10 | 0.30 | 0.98 | 0.07 | 0.79 | 0.92 |
| | R | 2.25 | 0.11 | 0.75 | 1.95 | 0.16 | 0.87 | 1.76 | 0.18 | 0.98 | 4.65 | 0.03 | 0.48 |
| Frontal Pole | L | 0.53 | 0.59 | 0.91 | 0.04 | 0.84 | 0.98 | 0.23 | 0.63 | 0.98 | 1.56 | 0.21 | 0.64 |
| | R | 0.63 | 0.53 | 0.91 | 0.66 | 0.42 | 0.93 | 2.81 | 0.09 | 0.98 | 1.62 | 0.20 | 0.64 |
| Fusiform | L | 1.22 | 0.29 | 0.86 | 1.06 | 0.30 | 0.91 | 1.45 | 0.23 | 0.98 | 0.56 | 0.45 | 0.78 |
| | R | 0.54 | 0.58 | 0.91 | 0.71 | 0.40 | 0.92 | 0.38 | 0.54 | 0.98 | 3.97 | 0.05 | 0.48 |
| Inferior Parietal | L | 1.18 | 0.31 | 0.86 | 2.26 | 0.13 | 0.86 | 0.00 | 0.97 | 0.98 | 1.01 | 0.31 | 0.70 |
| | R | 1.79 | 0.17 | 0.75 | 3.39 | 0.07 | 0.74 | 0.32 | 0.57 | 0.98 | 1.05 | 0.31 | 0.70 |
| Inferior Temporal | L | 0.51 | 0.60 | 0.91 | 0.67 | 0.41 | 0.93 | 0.71 | 0.40 | 0.98 | 0.16 | 0.68 | 0.89 |
| | R | 0.51 | 0.60 | 0.91 | 0.54 | 0.46 | 0.93 | 0.35 | 0.55 | 0.98 | 0.00 | 0.98 | 0.98 |
| Insula | L | 0.69 | 0.50 | 0.91 | 0.35 | 0.56 | 0.95 | 0.20 | 0.66 | 0.98 | 4.97 | 0.03 | 0.48 |
| | R | 1.05 | 0.35 | 0.86 | 1.47 | 0.23 | 0.87 | 0.23 | 0.63 | 0.98 | 2.40 | 0.12 | 0.59 |
| Isthmus Cingulate | L | 0.34 | 0.71 | 0.94 | 0.79 | 0.37 | 0.91 | 0.21 | 0.65 | 0.98 | 1.99 | 0.16 | 0.61 |
| | R | 0.05 | 0.95 | 0.97 | 0.13 | 0.71 | 0.95 | 0.04 | 0.83 | 0.98 | 3.55 | 0.06 | 0.48 |
| Lateral Occipital | L | 0.70 | 0.50 | 0.91 | 1.52 | 0.22 | 0.87 | 0.00 | 0.96 | 0.98 | 0.23 | 0.63 | 0.85 |
| | R | 3.09 | 0.05 | 0.75 | 6.28 | 0.01 | 0.63 | 0.00 | 0.97 | 0.98 | 0.88 | 0.35 | 0.70 |
| Lateral Orbitofrontal | L | 0.13 | 0.88 | 0.96 | 0.25 | 0.62 | 0.95 | 0.00 | 0.97 | 0.98 | 0.89 | 0.35 | 0.70 |
| | R | 1.29 | 0.28 | 0.86 | 0.04 | 0.85 | 0.98 | 0.03 | 0.86 | 0.98 | 0.25 | 0.62 | 0.85 |
| Lingual | L | 0.38 | 0.68 | 0.94 | 0.10 | 0.76 | 0.95 | 0.32 | 0.57 | 0.98 | 4.62 | 0.03 | 0.48 |
| | R | 0.32 | 0.72 | 0.94 | 0.27 | 0.60 | 0.95 | 0.80 | 0.37 | 0.98 | 2.90 | 0.09 | 0.53 |
| Medial Orbitofrontal | L | 1.56 | 0.21 | 0.75 | 2.46 | 0.12 | 0.84 | 0.01 | 0.93 | 0.98 | 1.59 | 0.21 | 0.64 |
| | R | 2.21 | 0.11 | 0.75 | 2.16 | 0.14 | 0.87 | 0.62 | 0.43 | 0.98 | 2.38 | 0.12 | 0.59 |
| Middle Temporal | L | 1.67 | 0.19 | 0.75 | 0.07 | 0.80 | 0.96 | 1.68 | 0.20 | 0.98 | 5.05 | 0.02 | 0.48 |
| | R | 0.49 | 0.62 | 0.91 | 1.21 | 0.27 | 0.91 | 0.14 | 0.71 | 0.98 | 0.03 | 0.86 | 0.94 |
| Paracentral | L | 0.16 | 0.85 | 0.96 | 0.03 | 0.86 | 0.98 | 2.22 | 0.14 | 0.98 | 0.59 | 0.44 | 0.78 |
| | R | 0.45 | 0.64 | 0.91 | 0.00 | 0.96 | 0.99 | 0.03 | 0.86 | 0.98 | 0.32 | 0.57 | 0.84 |
| Parahippocampal | L | 1.73 | 0.18 | 0.75 | 0.90 | 0.34 | 0.91 | 1.71 | 0.19 | 0.98 | 0.51 | 0.48 | 0.78 |
| | R | 3.46 | 0.03 | 0.75 | 0.17 | 0.68 | 0.95 | 0.77 | 0.38 | 0.98 | 0.15 | 0.70 | 0.89 |
| Pars Opercularis | L | 0.78 | 0.46 | 0.91 | 0.25 | 0.61 | 0.95 | 0.53 | 0.47 | 0.98 | 1.37 | 0.24 | 0.68 |
| | R | 2.46 | 0.09 | 0.75 | 3.92 | 0.05 | 0.74 | 1.41 | 0.24 | 0.98 | 6.42 | 0.01 | 0.35 |
| Pars Orbitalis | L | 0.98 | 0.38 | 0.86 | 0.57 | 0.45 | 0.93 | 0.00 | 0.96 | 0.98 | 1.12 | 0.29 | 0.70 |
| | R | 2.59 | 0.08 | 0.75 | 0.00 | 0.96 | 0.99 | 2.27 | 0.13 | 0.98 | 0.45 | 0.50 | 0.80 |
| Pars Triangularis | L | 1.89 | 0.15 | 0.75 | 0.78 | 0.38 | 0.91 | 0.79 | 0.38 | 0.98 | 1.05 | 0.31 | 0.70 |
| | R | 1.79 | 0.17 | 0.75 | 0.24 | 0.62 | 0.95 | 1.05 | 0.31 | 0.98 | 0.01 | 0.91 | 0.96 |
| Pericalcarine | L | 1.12 | 0.33 | 0.86 | 0.44 | 0.51 | 0.94 | 0.00 | 1.00 | 1.00 | 3.54 | 0.06 | 0.48 |
| | R | 0.12 | 0.88 | 0.96 | 0.00 | 1.00 | 1.00 | 4.07 | 0.04 | 0.98 | 0.11 | 0.74 | 0.90 |
| | L | 0.05 | 0.96 | 0.97 | 0.08 | 0.77 | 0.95 | 0.64 | 0.42 | 0.98 | 0.00 | 0.97 | 0.98 |

| | | | | | | | | | | | | | |
|---|---|---|---|---|---|---|---|---|---|---|---|---|---|
| Postcentral | R | 0.37 | 0.69 | 0.94 | 1.04 | 0.31 | 0.91 | 0.01 | 0.91 | 0.98 | 0.04 | 0.85 | 0.94 |
| Posterior Cingulate | L | 0.00 | 1.00 | 1.00 | 0.57 | 0.45 | 0.93 | 0.03 | 0.86 | 0.98 | 0.45 | 0.50 | 0.80 |
| | R | 0.19 | 0.83 | 0.96 | 0.21 | 0.64 | 0.95 | 0.16 | 0.69 | 0.98 | 0.78 | 0.38 | 0.70 |
| Precentral | L | 0.13 | 0.88 | 0.96 | 0.00 | 0.96 | 0.99 | 0.17 | 0.68 | 0.98 | 1.62 | 0.20 | 0.64 |
| | R | 0.22 | 0.81 | 0.96 | 0.23 | 0.63 | 0.95 | 0.93 | 0.34 | 0.98 | 0.01 | 0.91 | 0.96 |
| Precuneus | L | 1.84 | 0.16 | 0.75 | 3.35 | 0.07 | 0.74 | 0.00 | 0.97 | 0.98 | 3.58 | 0.06 | 0.48 |
| | R | 1.96 | 0.14 | 0.75 | 3.40 | 0.07 | 0.74 | 0.51 | 0.48 | 0.98 | 1.11 | 0.29 | 0.70 |
| Rostral Anterior Cingulate | L | 0.51 | 0.60 | 0.91 | 0.86 | 0.35 | 0.91 | 1.67 | 0.20 | 0.98 | 0.01 | 0.93 | 0.97 |
| | R | 1.52 | 0.22 | 0.75 | 0.34 | 0.56 | 0.95 | 0.23 | 0.63 | 0.98 | 4.04 | 0.04 | 0.48 |
| Rostral Middle Frontal | L | 0.58 | 0.56 | 0.91 | 1.02 | 0.31 | 0.91 | 0.18 | 0.67 | 0.98 | 2.69 | 0.10 | 0.53 |
| | R | 1.69 | 0.18 | 0.75 | 1.77 | 0.18 | 0.87 | 1.02 | 0.31 | 0.98 | 2.14 | 0.14 | 0.61 |
| Superior Frontal | L | 0.56 | 0.57 | 0.91 | 0.40 | 0.53 | 0.94 | 0.14 | 0.71 | 0.98 | 5.13 | 0.02 | 0.48 |
| | R | 0.50 | 0.61 | 0.91 | 0.98 | 0.32 | 0.91 | 0.02 | 0.88 | 0.98 | 1.40 | 0.24 | 0.68 |
| Superior Parietal | L | 0.96 | 0.38 | 0.86 | 2.43 | 0.12 | 0.84 | 0.02 | 0.88 | 0.98 | 1.27 | 0.26 | 0.70 |
| | R | 1.53 | 0.22 | 0.75 | 4.28 | 0.04 | 0.74 | 0.04 | 0.83 | 0.98 | 1.82 | 0.18 | 0.64 |
| Superior Temporal | L | 0.32 | 0.73 | 0.94 | 0.01 | 0.91 | 0.99 | 2.68 | 0.10 | 0.98 | 3.37 | 0.07 | 0.48 |
| | R | 0.69 | 0.50 | 0.91 | 0.06 | 0.80 | 0.96 | 0.33 | 0.57 | 0.98 | 0.22 | 0.64 | 0.85 |
| Supramarginal | L | 0.56 | 0.57 | 0.91 | 0.05 | 0.82 | 0.97 | 0.02 | 0.87 | 0.98 | 2.68 | 0.10 | 0.53 |
| | R | 1.75 | 0.17 | 0.75 | 2.00 | 0.16 | 0.87 | 0.16 | 0.69 | 0.98 | 9.03 | 2.68e-03 | 0.21 |
| Temporal Pole | L | 0.53 | 0.59 | 0.91 | 0.81 | 0.37 | 0.91 | 0.13 | 0.72 | 0.98 | 2.07 | 0.15 | 0.61 |
| | R | 1.13 | 0.32 | 0.86 | 0.08 | 0.78 | 0.95 | 0.33 | 0.57 | 0.98 | 0.50 | 0.48 | 0.78 |
| Transverse Temporal | L | 4.40 | 0.01 | 0.75 | 3.44 | 0.06 | 0.74 | 0.04 | 0.84 | 0.98 | 7.21 | 7.30e-03 | 0.35 |
| | R | 2.40 | 0.09 | 0.75 | 6.58 | 0.01 | 0.63 | 0.01 | 0.91 | 0.98 | 1.68 | 0.20 | 0.64 |
| **Surface Area** | | | | | | | | | | | | | |
| Banks of the Superior Temporal Sulcus | L | 1.07 | 0.34 | 0.86 | 1.10 | 0.30 | 0.91 | 0.01 | 0.92 | 0.98 | 0.22 | 0.64 | 0.85 |
| | R | 1.00 | 0.37 | 0.86 | 0.39 | 0.53 | 0.94 | 1.06 | 0.30 | 0.98 | 0.40 | 0.53 | 0.81 |
| Caudal Anterior Cingulate | L | 0.16 | 0.86 | 0.96 | 0.31 | 0.58 | 0.95 | 0.14 | 0.70 | 0.98 | 0.06 | 0.81 | 0.93 |
| | R | 0.21 | 0.81 | 0.96 | 0.19 | 0.66 | 0.95 | 0.01 | 0.94 | 0.98 | 0.53 | 0.47 | 0.78 |
| Caudal Middle Frontal | L | 0.79 | 0.46 | 0.91 | 2.49 | 0.11 | 0.84 | 1.57 | 0.21 | 0.98 | 0.83 | 0.36 | 0.70 |
| | R | 0.39 | 0.67 | 0.93 | 0.06 | 0.81 | 0.96 | 0.06 | 0.80 | 0.98 | 1.11 | 0.29 | 0.70 |
| Cuneus | L | 0.20 | 0.82 | 0.96 | 0.02 | 0.89 | 0.98 | 0.04 | 0.84 | 0.98 | 3.57 | 0.06 | 0.48 |
| | R | 0.05 | 0.96 | 0.97 | 0.00 | 0.97 | 0.99 | 0.02 | 0.88 | 0.98 | 0.79 | 0.38 | 0.70 |
| Entorhinal | L | 3.36 | 0.04 | 0.75 | 1.63 | 0.20 | 0.87 | 9.12 | 2.56e-03 | 0.39 | 0.03 | 0.87 | 0.94 |
| | R | 2.26 | 0.10 | 0.75 | 0.96 | 0.33 | 0.91 | 4.46 | 0.03 | 0.98 | 0.86 | 0.35 | 0.70 |
| Frontal Pole | L | 0.45 | 0.64 | 0.91 | 0.12 | 0.73 | 0.95 | 0.00 | 0.95 | 0.98 | 0.54 | 0.46 | 0.78 |
| | R | 0.16 | 0.86 | 0.96 | 0.48 | 0.49 | 0.94 | 1.11 | 0.29 | 0.98 | 1.01 | 0.31 | 0.70 |
| Fusiform | L | 2.13 | 0.12 | 0.75 | 0.01 | 0.91 | 0.99 | 3.84 | 0.05 | 0.98 | 0.80 | 0.37 | 0.70 |
| | R | 2.36 | 0.09 | 0.75 | 0.00 | 0.96 | 0.99 | 0.97 | 0.32 | 0.98 | 3.08 | 0.08 | 0.51 |
| Inferior Parietal | L | 0.05 | 0.95 | 0.97 | 0.00 | 0.97 | 0.99 | 1.08 | 0.30 | 0.98 | 2.12 | 0.15 | 0.61 |
| | R | 1.21 | 0.30 | 0.86 | 0.54 | 0.46 | 0.93 | 0.56 | 0.45 | 0.98 | 0.92 | 0.34 | 0.70 |
| Inferior Temporal | L | 1.55 | 0.21 | 0.75 | 1.44 | 0.23 | 0.87 | 0.10 | 0.75 | 0.98 | 0.03 | 0.87 | 0.94 |
| | R | 0.36 | 0.70 | 0.94 | 0.00 | 0.94 | 0.99 | 1.14 | 0.29 | 0.98 | 0.53 | 0.47 | 0.78 |
| Insula | L | 2.23 | 0.11 | 0.75 | 1.46 | 0.23 | 0.87 | 1.62 | 0.20 | 0.98 | 0.31 | 0.58 | 0.84 |
| | R | 2.00 | 0.14 | 0.75 | 1.34 | 0.25 | 0.87 | 2.47 | 0.12 | 0.98 | 1.90 | 0.17 | 0.64 |
| Isthmus Cingulate | L | 0.28 | 0.76 | 0.95 | 0.10 | 0.75 | 0.95 | 0.19 | 0.66 | 0.98 | 2.99 | 0.08 | 0.52 |
| | R | 1.76 | 0.17 | 0.75 | 4.58 | 0.03 | 0.74 | 0.55 | 0.46 | 0.98 | 3.08 | 0.08 | 0.51 |
| Lateral Occipital | L | 1.60 | 0.20 | 0.75 | 5.01 | 0.03 | 0.74 | 0.05 | 0.82 | 0.98 | 0.41 | 0.52 | 0.81 |

| | | | | | | | | | | | | | |
|---|---|---|---|---|---|---|---|---|---|---|---|---|---|
| | R | 0.82 | 0.44 | 0.91 | 0.85 | 0.36 | 0.91 | 1.85 | 0.17 | 0.98 | 4.12 | 0.04 | 0.48 |
| Lateral Orbitofrontal | L | 0.28 | 0.75 | 0.95 | 1.02 | 0.31 | 0.91 | 1.29 | 0.26 | 0.98 | 0.17 | 0.68 | 0.89 |
| | R | 0.11 | 0.90 | 0.96 | 0.00 | 0.95 | 0.99 | 0.01 | 0.91 | 0.98 | 0.04 | 0.84 | 0.94 |
| Lingual | L | 0.07 | 0.93 | 0.97 | 0.02 | 0.89 | 0.98 | 0.00 | 0.96 | 0.98 | 1.16 | 0.28 | 0.70 |
| | R | 0.14 | 0.87 | 0.96 | 0.22 | 0.64 | 0.95 | 0.23 | 0.63 | 0.98 | 0.02 | 0.90 | 0.96 |
| Medial Orbitofrontal | L | 0.05 | 0.95 | 0.97 | 0.01 | 0.93 | 0.99 | 0.71 | 0.40 | 0.98 | 0.47 | 0.50 | 0.80 |
| | R | 1.62 | 0.20 | 0.75 | 0.16 | 0.69 | 0.95 | 2.82 | 0.09 | 0.98 | 2.73 | 0.10 | 0.53 |
| Middle Temporal | L | 0.60 | 0.55 | 0.91 | 1.01 | 0.31 | 0.91 | 0.79 | 0.37 | 0.98 | 0.03 | 0.86 | 0.94 |
| | R | 0.39 | 0.67 | 0.93 | 0.24 | 0.62 | 0.95 | 0.03 | 0.85 | 0.98 | 1.04 | 0.31 | 0.70 |
| Paracentral | L | 0.94 | 0.39 | 0.86 | 0.11 | 0.74 | 0.95 | 0.41 | 0.52 | 0.98 | 0.07 | 0.79 | 0.92 |
| | R | 0.73 | 0.48 | 0.91 | 2.06 | 0.15 | 0.87 | 0.18 | 0.67 | 0.98 | 0.00 | 0.99 | 0.99 |
| Parahippocampal | L | 2.01 | 0.13 | 0.75 | 2.95 | 0.09 | 0.82 | 4.49 | 0.03 | 0.98 | 0.84 | 0.36 | 0.70 |
| | R | 1.04 | 0.35 | 0.86 | 1.51 | 0.22 | 0.87 | 0.01 | 0.91 | 0.98 | 3.43 | 0.06 | 0.48 |
| Pars Opercularis | L | 2.79 | 0.06 | 0.75 | 4.90 | 0.03 | 0.74 | 3.31 | 0.07 | 0.98 | 0.98 | 0.32 | 0.70 |
| | R | 0.09 | 0.91 | 0.97 | 0.23 | 0.63 | 0.95 | 0.06 | 0.81 | 0.98 | 0.13 | 0.72 | 0.89 |
| Pars Orbitalis | L | 0.07 | 0.93 | 0.97 | 0.03 | 0.86 | 0.98 | 3.18 | 0.07 | 0.98 | 0.22 | 0.64 | 0.85 |
| | R | 0.42 | 0.66 | 0.93 | 0.60 | 0.44 | 0.93 | 0.24 | 0.62 | 0.98 | 0.38 | 0.54 | 0.82 |
| Pars Triangularis | L | 0.27 | 0.76 | 0.95 | 0.64 | 0.42 | 0.93 | 1.95 | 0.16 | 0.98 | 0.30 | 0.59 | 0.84 |
| | R | 0.57 | 0.57 | 0.91 | 0.42 | 0.52 | 0.94 | 0.39 | 0.53 | 0.98 | 0.83 | 0.36 | 0.70 |
| Pericalcarine | L | 0.01 | 0.99 | 1.00 | 0.09 | 0.76 | 0.95 | 0.03 | 0.86 | 0.98 | 2.06 | 0.15 | 0.61 |
| | R | 0.17 | 0.84 | 0.96 | 0.10 | 0.75 | 0.95 | 0.55 | 0.46 | 0.98 | 0.14 | 0.71 | 0.89 |
| Postcentral | L | 0.47 | 0.62 | 0.91 | 0.38 | 0.54 | 0.94 | 1.09 | 0.30 | 0.98 | 0.03 | 0.86 | 0.94 |
| | R | 1.86 | 0.16 | 0.75 | 0.00 | 0.97 | 0.99 | 0.02 | 0.88 | 0.98 | 3.67 | 0.06 | 0.48 |
| Posterior Cingulate | L | 0.33 | 0.72 | 0.94 | 0.76 | 0.38 | 0.92 | 0.47 | 0.49 | 0.98 | 0.08 | 0.78 | 0.92 |
| | R | 1.73 | 0.18 | 0.75 | 1.34 | 0.25 | 0.87 | 0.58 | 0.45 | 0.98 | 1.04 | 0.31 | 0.70 |
| Precentral | L | 1.30 | 0.27 | 0.86 | 0.41 | 0.52 | 0.94 | 0.06 | 0.81 | 0.98 | 0.32 | 0.57 | 0.84 |
| | R | 3.46 | 0.03 | 0.75 | 0.62 | 0.43 | 0.93 | 1.95 | 0.16 | 0.98 | 1.50 | 0.22 | 0.66 |
| Precuneus | L | 0.94 | 0.39 | 0.86 | 1.36 | 0.24 | 0.87 | 2.32 | 0.13 | 0.98 | 0.28 | 0.59 | 0.84 |
| | R | 0.57 | 0.56 | 0.91 | 0.20 | 0.65 | 0.95 | 0.91 | 0.34 | 0.98 | 0.22 | 0.64 | 0.85 |
| Rostral Anterior Cingulate | L | 0.23 | 0.80 | 0.96 | 0.08 | 0.78 | 0.95 | 1.32 | 0.25 | 0.98 | 0.12 | 0.73 | 0.90 |
| | R | 0.17 | 0.84 | 0.96 | 0.08 | 0.77 | 0.95 | 0.05 | 0.83 | 0.98 | 0.14 | 0.71 | 0.89 |
| Rostral Middle Frontal | L | 0.23 | 0.79 | 0.96 | 0.72 | 0.40 | 0.92 | 0.07 | 0.79 | 0.98 | 1.31 | 0.25 | 0.70 |
| | R | 1.36 | 0.26 | 0.85 | 0.20 | 0.65 | 0.95 | 0.02 | 0.89 | 0.98 | 0.00 | 0.95 | 0.98 |
| Superior Frontal | L | 3.69 | 0.02 | 0.75 | 0.53 | 0.47 | 0.93 | 2.10 | 0.15 | 0.98 | 2.05 | 0.15 | 0.61 |
| | R | 0.96 | 0.38 | 0.86 | 0.11 | 0.74 | 0.95 | 3.23 | 0.07 | 0.98 | 0.86 | 0.35 | 0.70 |
| Superior Parietal | L | 0.21 | 0.81 | 0.96 | 0.09 | 0.76 | 0.95 | 0.20 | 0.65 | 0.98 | 0.06 | 0.81 | 0.93 |
| | R | 0.35 | 0.71 | 0.94 | 0.19 | 0.66 | 0.95 | 0.54 | 0.46 | 0.98 | 0.44 | 0.51 | 0.80 |
| Superior Temporal | L | 1.22 | 0.29 | 0.86 | 0.14 | 0.71 | 0.95 | 0.50 | 0.48 | 0.98 | 1.78 | 0.18 | 0.64 |
| | R | 0.55 | 0.58 | 0.91 | 0.02 | 0.89 | 0.98 | 0.04 | 0.85 | 0.98 | 0.14 | 0.71 | 0.89 |
| Supramarginal | L | 0.11 | 0.90 | 0.96 | 0.02 | 0.89 | 0.98 | 1.04 | 0.31 | 0.98 | 0.11 | 0.74 | 0.90 |
| | R | 0.26 | 0.77 | 0.95 | 0.00 | 0.99 | 0.99 | 0.00 | 0.95 | 0.98 | 0.01 | 0.91 | 0.96 |
| Temporal Pole | L | 1.56 | 0.21 | 0.75 | 1.65 | 0.20 | 0.87 | 2.29 | 0.13 | 0.98 | 2.31 | 0.13 | 0.60 |
| | R | 0.76 | 0.47 | 0.91 | 0.25 | 0.62 | 0.95 | 0.92 | 0.34 | 0.98 | 2.84 | 0.09 | 0.53 |
| Transverse Temporal | L | 0.50 | 0.61 | 0.91 | 0.04 | 0.85 | 0.98 | 1.48 | 0.22 | 0.98 | 0.79 | 0.37 | 0.70 |
| | R | 0.55 | 0.58 | 0.91 | 0.10 | 0.75 | 0.95 | 0.26 | 0.61 | 0.98 | 0.65 | 0.42 | 0.76 |
| **Subcortical Volume** | | | | | | | | | | | | | |
| Amygdala | L | 0.39 | 0.68 | 0.93 | 0.26 | 0.61 | 0.95 | 1.35 | 0.25 | 0.98 | 0.93 | 0.34 | 0.70 |
| | R | 1.40 | 0.25 | 0.83 | 0.50 | 0.48 | 0.94 | 0.55 | 0.46 | 0.98 | 0.92 | 0.34 | 0.70 |
| Caudate | L | 1.18 | 0.31 | 0.86 | 2.82 | 0.09 | 0.82 | 0.10 | 0.75 | 0.98 | 0.28 | 0.60 | 0.84 |
| | R | 1.03 | 0.36 | 0.86 | 2.26 | 0.13 | 0.86 | 0.09 | 0.77 | 0.98 | 0.00 | 0.97 | 0.98 |

| | | | | | | | | | | | | | |
|---|---|---|---|---|---|---|---|---|---|---|---|---|---|
| Hippocampus | L | 1.00 | 0.37 | 0.86 | 1.70 | 0.19 | 0.87 | 0.17 | 0.68 | 0.98 | 0.30 | 0.58 | 0.84 |
| | R | 1.28 | 0.28 | 0.86 | 1.67 | 0.20 | 0.87 | 0.23 | 0.63 | 0.98 | 0.76 | 0.38 | 0.70 |
| Lateral Ventricles | L | 0.71 | 0.49 | 0.91 | 1.00 | 0.32 | 0.91 | 0.23 | 0.63 | 0.98 | 0.08 | 0.78 | 0.92 |
| | R | 1.54 | 0.22 | 0.75 | 3.48 | 0.06 | 0.74 | 0.01 | 0.91 | 0.98 | 0.52 | 0.47 | 0.78 |
| Nucleus Accumbens | L | 0.46 | 0.63 | 0.91 | 0.81 | 0.37 | 0.91 | 0.00 | 0.95 | 0.98 | 0.13 | 0.72 | 0.89 |
| | R | 0.28 | 0.75 | 0.95 | 0.19 | 0.67 | 0.95 | 0.61 | 0.43 | 0.98 | 1.18 | 0.28 | 0.70 |
| Pallidum | L | 0.73 | 0.48 | 0.91 | 0.37 | 0.54 | 0.94 | 0.00 | 0.99 | 1.00 | 1.56 | 0.21 | 0.64 |
| | R | 0.12 | 0.89 | 0.96 | 0.09 | 0.76 | 0.95 | 0.36 | 0.55 | 0.98 | 0.63 | 0.43 | 0.76 |
| Putamen | L | 1.73 | 0.18 | 0.75 | 2.78 | 0.10 | 0.82 | 0.87 | 0.35 | 0.98 | 0.00 | 0.95 | 0.98 |
| | R | 0.99 | 0.37 | 0.86 | 1.83 | 0.18 | 0.87 | 0.07 | 0.79 | 0.98 | 0.04 | 0.84 | 0.94 |
| Thalamus | L | 0.55 | 0.58 | 0.91 | 0.87 | 0.35 | 0.91 | 0.45 | 0.50 | 0.98 | 0.26 | 0.61 | 0.85 |
| | R | 0.87 | 0.42 | 0.91 | 0.82 | 0.37 | 0.91 | 0.10 | 0.75 | 0.98 | 0.02 | 0.88 | 0.94 |
| **Global** | | | | | | | | | | | | | |
| Estimated Intracranial Volume | N/A | 0.83 | 0.44 | 0.91 | 1.70 | 0.19 | 0.87 | NA | 999.00 | 999.00 | 0.34 | 0.56 | 0.84 |
| Mean Cortical Thickness | | 0.71 | 0.49 | 0.91 | 1.17 | 0.28 | 0.91 | NA | 999.00 | 999.00 | 3.32 | 0.07 | 0.48 |
| Total Surface Area | | 0.53 | 0.59 | 0.91 | 0.57 | 0.45 | 0.93 | NA | 999.00 | 999.00 | 1.39 | 0.24 | 0.68 |

*For all interaction analyses, psychosis conversion consisted of three levels (CHR-PS+, CHR-PS-, HC). For "all-by-conversion", "all" consisted of four levels (APS, BIPS, GRD, HC). For all other interaction analyses (APS/BIPS/GRD-by-psychosis conversion), APS/BIPS/GRD consisted of three levels (APS/BIPS/GRD vs. no APS/BIPS/GRD vs. HC).*

**eTable 20.** Interaction between group (HC vs. CHR) and the smoothed effect of age on structural neuroimaging metrics

| | | Age-interaction statistics | | |
|---|---|---|---|---|
| **Region of Interest** | **Hemisphere** | **F** | **p** | **q** |
| **Cortical Thickness** | | | | |
| Banks of the Superior Temporal Sulcus | L | 0.36 | 0.55 | 1.00 |
| | R | 0.21 | 0.65 | 1.00 |
| Caudal Middle Frontal | L | 0.02 | 0.88 | 1.00 |
| | R | 0.57 | 0.45 | 1.00 |
| Fusiform | L | 4.93 | 0.01 | 0.67 |
| | R | 2.36 | 0.12 | 1.00 |
| Inferior Parietal | L | 0.69 | 0.41 | 1.00 |
| | R | 0.63 | 0.43 | 1.00 |
| Inferior Temporal | L | 5.11 | 0.02 | 1.00 |
| | R | 0.07 | 0.79 | 1.00 |
| Insula | L | 3.02 | 0.09 | 1.00 |
| | R | 3.67 | 0.06 | 1.00 |
| Lateral Occipital | L | 0.25 | 0.62 | 1.00 |
| | R | 1.92 | 0.17 | 1.00 |
| Lateral Orbitofrontal | R | 0.31 | 0.58 | 1.00 |
| Medial Orbitofrontal | L | 0.95 | 0.33 | 1.00 |
| | R | 0.35 | 0.55 | 1.00 |
| Middle Temporal | L | 0.20 | 0.66 | 1.00 |
| | R | 0.52 | 0.47 | 1.00 |
| Paracentral | L | 0.39 | 0.74 | 1.00 |
| | R | 0.07 | 0.79 | 1.00 |
| Parahippocampal | L | 0.21 | 0.65 | 1.00 |
| Postcentral | L | 0.28 | 0.60 | 1.00 |
| | R | 3.02 | 0.08 | 1.00 |
| Posterior Cingulate | L | 1.15 | 0.28 | 1.00 |
| | R | 0.09 | 0.77 | 1.00 |
| Precentral | L | 0.00 | 0.96 | 1.00 |
| | R | 0.66 | 0.61 | 1.00 |
| Precuneus | L | 0.98 | 0.32 | 1.00 |
| | R | 0.04 | 0.85 | 1.00 |
| Rostral Anterior Cingulate | L | 0.53 | 0.47 | 1.00 |
| Rostral Middle Frontal | L | 0.00 | 0.99 | 1.00 |
| | R | 0.03 | 0.87 | 1.00 |
| Superior Frontal | L | 0.03 | 0.86 | 1.00 |
| | R | 0.22 | 0.64 | 1.00 |
| Superior Parietal | L | 0.03 | 0.86 | 1.00 |
| | R | 0.88 | 0.35 | 1.00 |
| Superior Temporal | L | 0.07 | 0.79 | 1.00 |
| | R | 0.20 | 0.66 | 1.00 |
| Supramarginal | L | 0.10 | 0.75 | 1.00 |
| | R | 1.41 | 0.27 | 1.00 |
| Transverse Temporal | R | 4.50 | 0.03 | 1.00 |
| **Surface Area** | | | | |
| Entorhinal | L | 0.09 | 0.77 | 1.00 |
| Inferior Parietal | R | 2.63 | 0.09 | 1.00 |

| | | | | |
|---|---|---|---|---|
| Isthmus Cingulate | R | 0.21 | 0.65 | 1.00 |
| Pars Opercularis | L | 0.00 | 1.00 | 1.00 |
| Pericalcarine | R | 0.83 | 0.37 | 1.00 |
| Rostral Anterior Cingulate | L | 0.07 | 0.79 | 1.00 |
| Superior Frontal | R | 5.19 | 0.02 | 1.00 |
| Supramarginal | L | 1.27 | 0.26 | 1.00 |
| **Subcortical Volume** | | | | |
| Hippocampus | R | 1.15 | 0.25 | 1.00 |
| Lateral Ventricles | L | 0.46 | 0.69 | 1.00 |
| | R | 1.00 | 0.32 | 1.00 |
| **Global** | | | | |
| Estimated Intracranial Volume | N/A | 3.40 | 0.06 | 1.00 |
| Mean Cortical Thickness | N/A | 1.69 | 0.28 | 1.00 |
| Total Surface Area | N/A | 7.60 | 5.69e-03 | 0.32 |

**eTable 21.** Interaction between conversion status (CHR-PS+ vs. HC, CHR-PS+ vs. CHR-PS-, CHR-PS- vs. HC) and the smoothed effect of age on structural neuroimaging metrics.

| | | Age-interaction statistics CHR-PS+ vs. HC | | | Age-interaction statistics CHR-PS+ vs. CHR-PS- | | | Age-interaction statistics CHR-PS- vs. HC | | |
|---|---|---|---|---|---|---|---|---|---|---|
| **Region of Interest** | **Hemisphere** | **F** | **p** | **q** | **F** | **p** | **q** | **F** | **p** | **q** |
| **Cortical Thickness** | | | | | | | | | | |
| Fusiform | L | **9.76** | **4.88e-05** | **5.85e-04** | 1.34 | 0.31 | 0.45 | **8.7** | **1.51e-04** | **9.05e-04** |
| Paracentral | L | **5.29** | **4.68e-03** | **0.02** | 1.89 | 0.18 | 0.45 | 0.23 | 0.68 | 0.74 |
| | R | 1.5 | 0.20 | 0.45 | 1.35 | 0.30 | 0.45 | 0.01 | 0.92 | 0.92 |
| Superior Temporal | R | 1.17 | 0.34 | 0.45 | 0.41 | 0.51 | 0.61 | 1.06 | 0.28 | 0.45 |

*Results marked in bold are significant at q<0.05.*

**eFigure 1.** CHR vs. HC effect size overview (post-ComBat mega analysis)

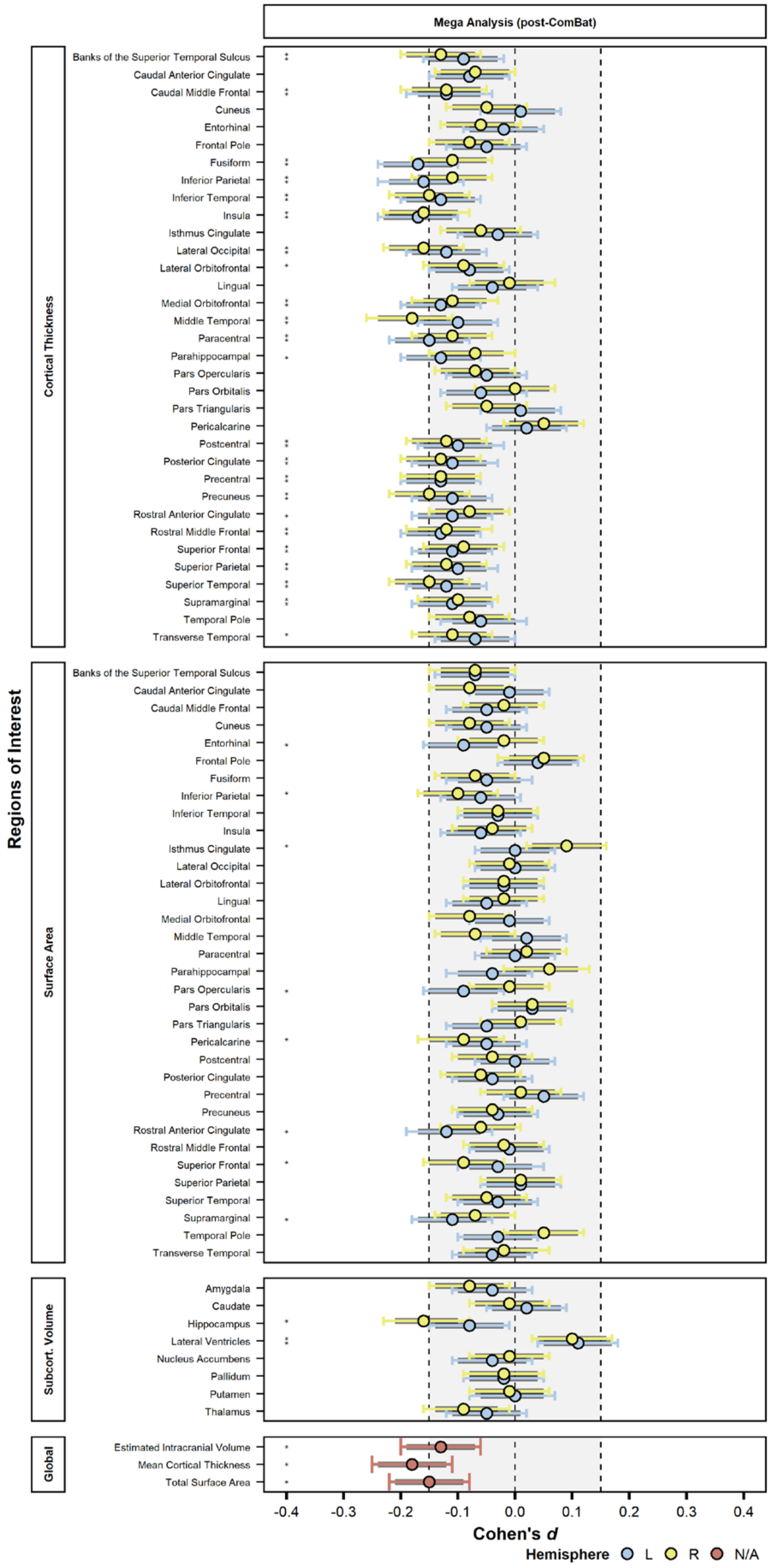

*q<0.05. Colored bars represent 95% confidence intervals. Black lines represent 90% confidence intervals used for equivalence testing. Shaded area represents the interval between upper (left vertical dashed line, -0.15) and lower (right vertical dashed line, +0.15) SESOI bounds.

**eFigure 2.** Jackknife resampling analysis for structural neuroimaging measures meeting statistical significance (q<0.05) in Aim 1

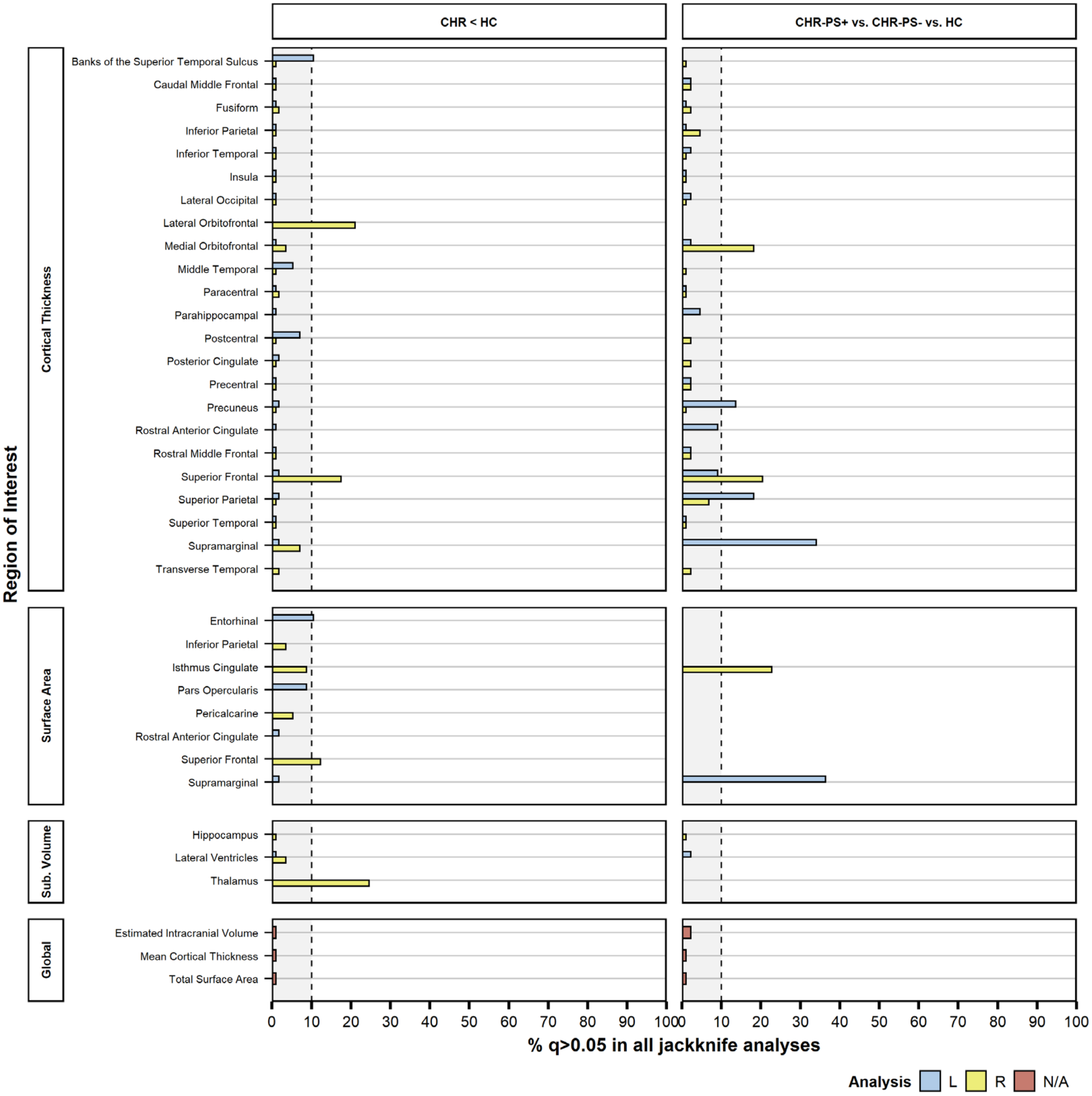

**Left panel:** regions of interest significant at q<0.05 in CHR vs HC post-ComBat mega analysis. **Right panel:** regions of interest significant at q<0.05 in CHR-PS+ vs. CHR-PS- vs. HC post-ComBat mega analysis (overall effect). *Regions that failed to show a psychosis risk/conversion (overall) effect at q<0.05 in more than 10% of all iterations (four study sites) were considered untrustworthy. Vertical dashed line represents 10% cut-off. Absence of colored bar indicates q>0.05 for a particular region in of the two post-ComBat mega analyses; these regions were therefore not included in the jackknife analysis. For visualization purposes, regions of interest with q<0.05 for every iteration are set to 1%.*

**eFigure 3.** CHR-PS+ vs. CHR-PS- vs. HC effect size overview (post-ComBat mega analysis)

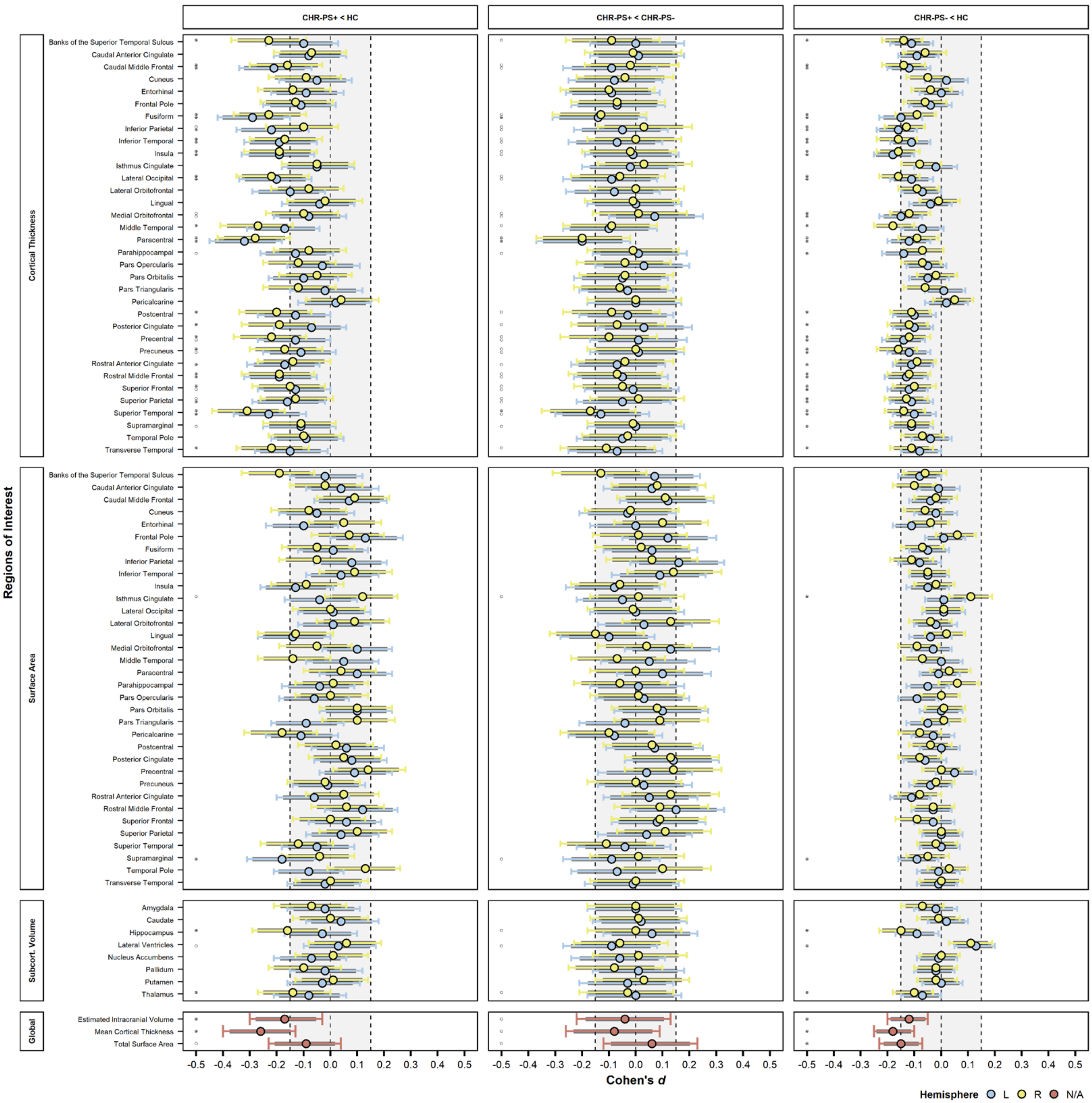

◦overall effect of psychosis conversion q<0.05; * pairwise comparison p<0.05 (note: only displayed for regions for which overall effect was q<0.05). *Colored bars represent 95% confidence intervals. Black lines represent 90% confidence intervals used for equivalence testing. Shaded area represents the interval between upper (left vertical dashed line, -0.15) and lower (right vertical dashed line, +0.15) SESOI bounds.*

**eFigure 4.** CHR-PS+ vs. SZ vs. 22q.11DS effect size overview

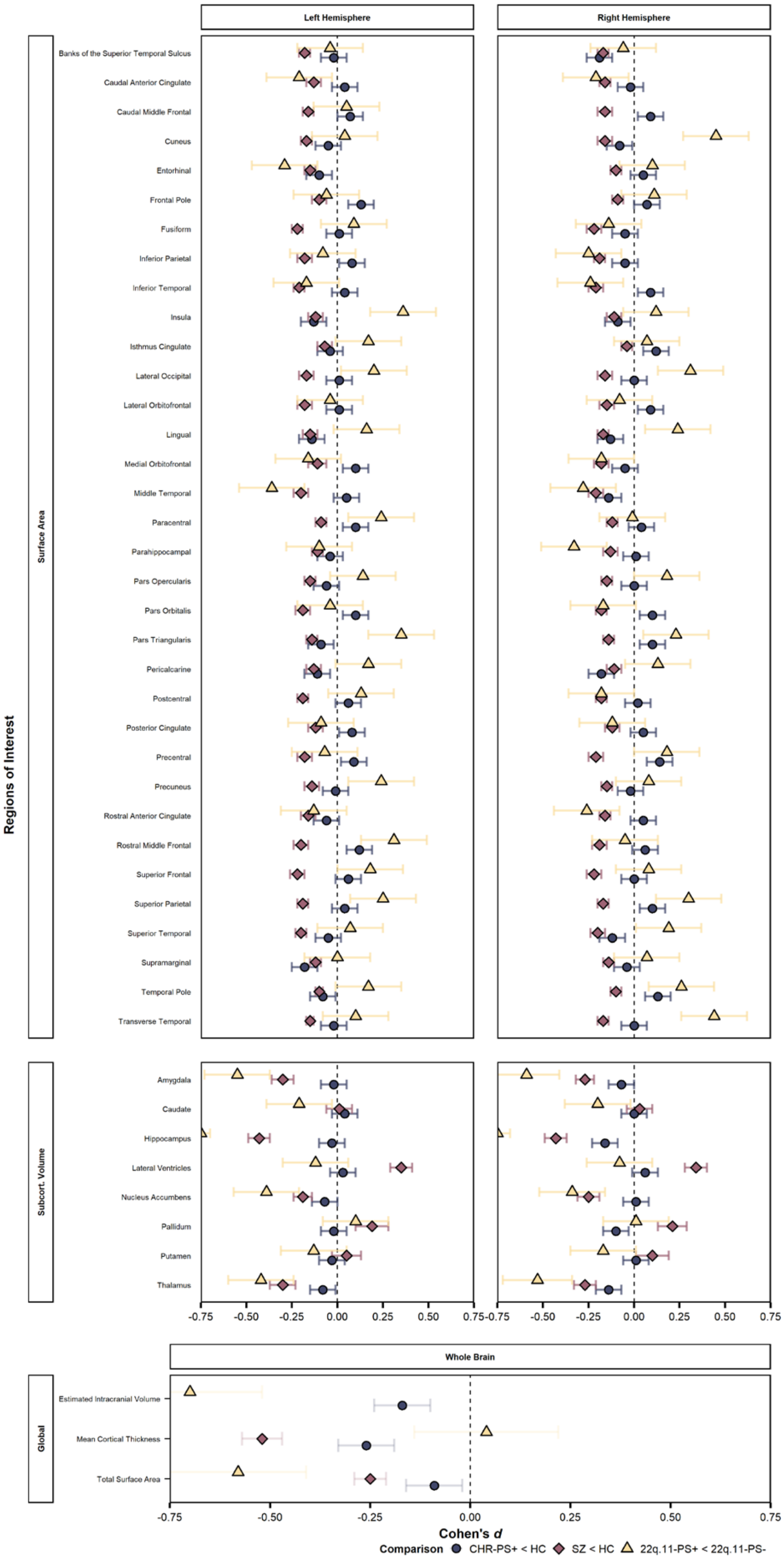

Estimated intracranial volume was not available for individuals with schizophrenia. Colored bars represent standard error.

**eFigure 5.** Surface Area, Subcortical Volume and global measure correlations between CHR-PS+, SZ, and 22q.11DS

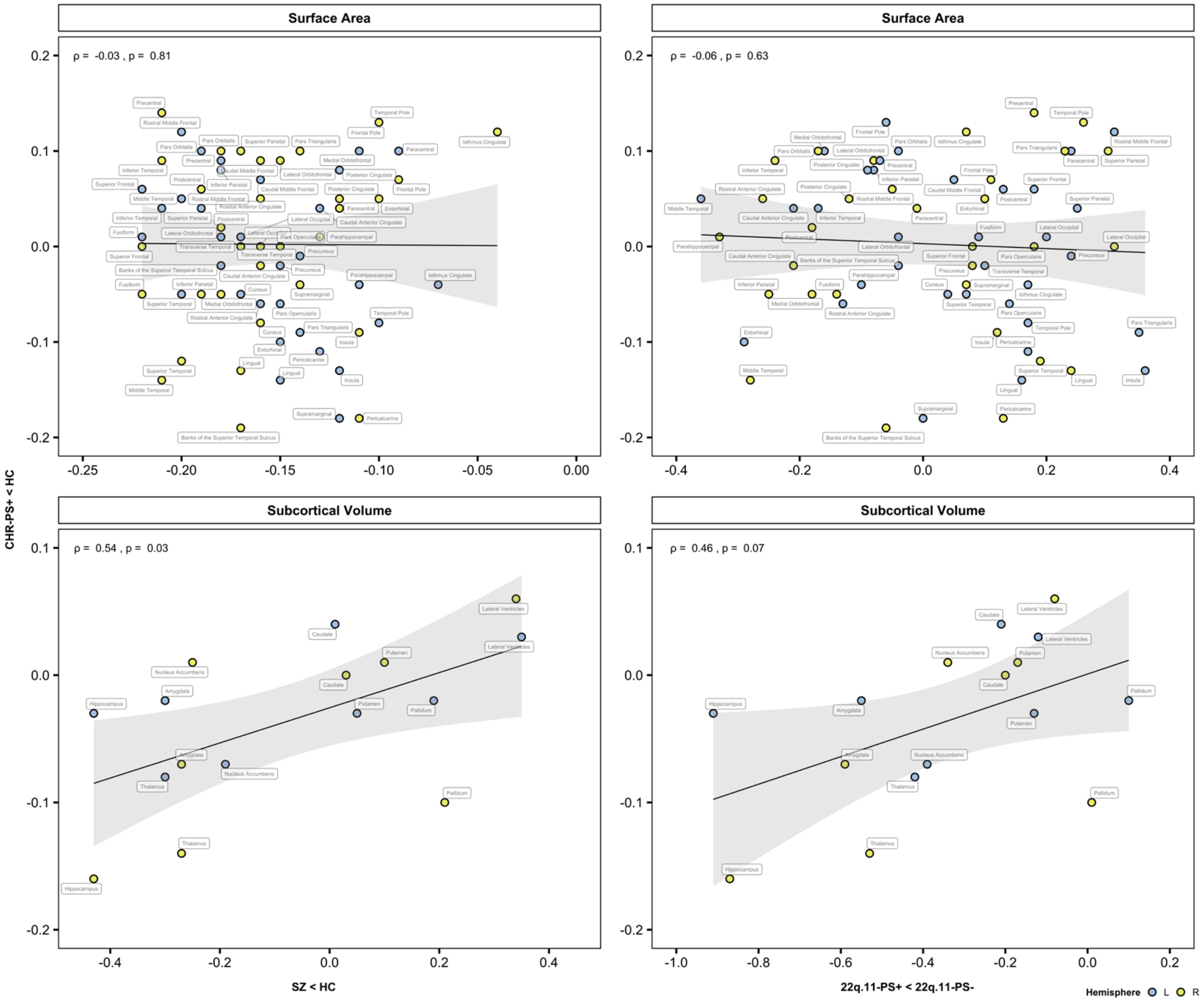

ρ = Spearman’s rank correlation.

12. Lakens D, Scheel AM, Isager PM. Equivalence Testing for Psychological Research: A Tutorial: *Advances in Methods and Practices in Psychological Science*. Published online June 1,

13. doi:10.1177/2515245918770963

13. Calabro FJ, Murty VP, Jalbrzikowski M, Tervo-Clemmens B, Luna B. Development of Hippocampal–Prefrontal Cortex Interactions through Adolescence. *Cereb Cortex*. 2020;30(3):1548-1558. doi:10.1093/cercor/bhz186

14. Wood SN. *Generalized Additive Models: An Introduction with R*. Second edition. CRC Press/Taylor & Francis Group; 2017.

15. Steiger JH. Tests for comparing elements of a correlation matrix. *Psychological Bulletin*. 1980;87(2):245-251. doi:10.1037/0033-2909.87.2.245

16. Diedenhofen B, Musch J. cocor: A Comprehensive Solution for the Statistical Comparison of Correlations. *PLOS ONE*. 2015;10(4):e0121945. doi:10.1371/journal.pone.0121945

17. Lemmers-Jansen IL, Fett A-KJ, van Os J, Veltman DJ, Krabbendam L. Trust and the city: Linking urban upbringing to neural mechanisms of trust in psychosis. *Aust N Z J Psychiatry*. 2020;54(2):138-149. doi:10.1177/0004867419865939

18. Lemmers-Jansen ILJ, Fett A-KJ, Van Doesum NJ, Van Lange PAM, Veltman DJ, Krabbendam L. Social Mindfulness

and Psychosis: Neural Response to Socially Mindful Behavior in First-Episode Psychosis and Patients at Clinical High-Risk. *Front Hum Neurosci*. 2019;13:47. doi:10.3389/fnhum.2019.00047

51. Kwak YB, Kim M, Cho KIK, Lee J, Lee TY, Kwon JS. Reduced cortical thickness in subjects at clinical high risk for

psychosis and clinical attributes. *Aust N Z J Psychiatry*. 2019;53(3):219-227. doi:10.1177/0004867418807299

81. Schultze-Lutter F, Addington J, Ruhrmann S, Klosterkötter J. Schizophrenia proneness instrument, adult version (SPI-A).

82. Klosterkötter J, Ruhrmann S, Schultze-Lutter F, et al. The European Prediction of Psychosis Study (EPOS): integrating early recognition and intervention in Europe. *World Psychiatry*. Published online 2005:7.

83. Lencz T. Nonspecific and attenuated negative symptoms in patients at clinical high-risk for schizophrenia. *Schizophrenia Research*. 2004;68(1):37-48. doi:10.1016/S0920-9964(03)00214-7
